# Global Trends and Risk Factors of Aortic Aneurysm Mortality from 1990 to 2021: An Analysis of the Global Burden of Disease Study 2021

**DOI:** 10.1101/2025.03.26.25324707

**Authors:** Raluca-Maria Câsu, Loïc Metz, David Freiholtz, Xiaofeng Zheng, Hanna M Björck, Xiaowei Zheng

## Abstract

**Background:** Aortic aneurysm (AA) is a life-threatening disease with significant global burden.

**Objectives:** This study aims to evaluate epidemiological trends and risk factors for AA-related mortality from 1990 to 2021 across regions, accounting for age, sex, and socio-economic factors.

**Methods:** Using Global Burden of Disease (GBD) Study 2021, we analyzed AA-related death, death rates, and the age-standardized AA-related death rate (ASDR) per 100,000, along with risk factors. Trends from 1990 to 2021 were compared across global regions and countries by socio-demographic index, health systems, and income. We also examined the impact and trend changes of age, sex, and risk factors on AA.

**Results:** In 2021, global AA-related deaths reached 153,927 (95% uncertainty intervals (UI): 138,413-165,738), a 74.2% increase from 1990. However, accounting for changes in population size and age, ASDR declined from 2.54 (95%UI: 2.35-2.69) to 1.86 (95%UI: 1.67-2.00) deaths per 100,000 people.

Europe and America experienced ASDR reductions of 24.8% and 47.4%, while Asia saw a 38.6% increase. AA mortality remained high in regions with high income, advanced health system, and high socio-demographic index, especially in aged population. In 2021, Japan reported the most AA-related deaths (23,815, 95% UI: 19,180-26,463) and Armenia had the highest ASDR (9.16 per 100,000, 95% UI: 7.61-10.81).

Our results highlight significant sex differences in AA-related mortality. Men had nearly twice the ASDR of women, though the gap narrowed over time. The impact varied by age and region. ASDR declined more in men in Europe and America, especially in Sweden, Norway and Denmark. However, in Russia, Japan and Nauru, women saw greater increase, influencing overall AA-caused mortality.

AA-related risk factors differ by sex: smoking is the primary risk factors for men, while high systolic blood pressure is more significant for women. Other risk factors include high body-mass index, diets low in fruits and vegetables, increased sodium intake and lead exposure. Importantly, the relative contribution of these risk factors has shifted over time, reflecting changes in lifestyle, public health policies, and healthcare access.

**Conclusion:** AA-related mortality remains a global burden with regional and sex disparities. Declines of AA-related ASDR in Western Europe and the America suggest effective interventions, while increases in Eastern Europe, Central and South Asia, and Japan, especially among women, highlight emerging challenges. Smoking, hypertension, and obesity are key contributors, emphasizing the need for targeted prevention, screening and healthcare access.

## Introduction

An aortic aneurysm (AA) is a life-threatening vascular disease defined as the focal and permanent enlargement of the aorta, exceeding 50% of its normal diameter^1,2^. AA generally progresses asymptomatically and are often discovered incidentally during routine physical examinations or on imaging^1^, except for three countries (UK, US and Sweden) where screening programs for men above 65 have been implemented^3–5^. AA most commonly occur between the renal arteries and the aortic bifurcation^6^. Based on the location of the aneurysm, they are classified either as thoracic aortic aneurysm (TAA) or abdominal aortic aneurysm (AAA), with the latter being the most prevalent. The prevalence of AAA ranges from 0.4 to 7.6% depending on the population^7^, while the global prevalence of TAA is 0.16% (95% CI: 0.12-0.20)^8^.

The main risk factors for AAA include advanced age, smoking, male sex, family history of AAA and hypertension^9^. TAA has less well characterized risk factors and is in ∼20% of cases associated with genetic syndromes affecting connective tissue, such as Marfan syndrome and Ehlers-Danlos syndrome, or clear family history^1,10^. TAA also frequently occur in association with a bicuspid aortic valve, denoted BAV-aortopathy^11^. Rupture of an AA is associated with high mortality rates^9^. The primary approach for AA prevention has focused on smoking cessation and hypertension control. Surgical intervention is indicated for large aneurysms or those that are at high risk of rupture. As for now, no pharmacological therapies have been approved for the treatment of aortic aneurysm^1^.

The Global Burden of Diseases, Injuries, and Risk Factors Study (GBD) is a comprehensive research initiative that quantifies the impact of diseases, injuries, and risk factors on global health in more than 200 countries and at the subnational level in more than 20 countries^12,13^. It was initiated more than 30 years ago and has been conducted by the Institute for Health Metrics and Evaluation (IHME). It provides dynamic data since 1990 for various diseases, injuries, and risk factors, broken down by region, age, and sex, and tracks changes over time. Its standardized metrics, such as age-standardized death rates (ASDR), allow for accurate comparisons across populations and time periods while minimizing biases from demographic changes. The goal is to help inform public health policies and priorities by highlighting major health challenges and guiding resource allocation for interventions ^12,13^.

Recent studies have utilized the GBD 2019 data to examine the global burden of AA and its attributable factors or to focus on the disease burden on specific countries, such as Iran and China, to provide a more localized perspective ^14–17^. However, more recent epidemiological data needs to be evaluated to understand the gaps between different regions in the world, and between men and women given emerging evidence of unfavorable outcomes for women in AA-treatment^18^. This study aims to explore the updated data from the GBD 2021 study, in order to assess the global and regional epidemiological trends in AA-related mortality from 1990 to 2021, identify disparities in disease burden, and evaluate the impact of key risk factors. By providing a comprehensive evaluation of past and current AA mortality epidemiology, the findings of this study will help guide targeted screening and preventative efforts to reduced AA-related mortality worldwide.

## Methods

### Data source

The data for this analysis were sourced from The Global Burden of Diseases, Injuries, and Risk Factors Study (GBD) 2021 through the online query tool https://vizhub.healthdata.org/gbd-results/. The GBD study was conducted by the Institute for Health Metrics and Evaluation (IHME) at the University of Washington. The GBD 2021 provides a robust and comprehensive framework for understanding the burden of 288 causes of death and the contribution of 88 risk factors for both men and women worldwide, covering 204 countries and 811 subnational locations, for each year from 1990 until 2021^12,13^.

### Data extracted from GBD 2021

The GBD 2021 provides data on burden of aortic aneurysm combining both abdominal and thoracic aneurysms at global, regional, national and subnational levels. AA definition followed the GBD grouping, with a diagnosis corresponding to one of ICD-10 I71 to 171.9. To evaluate the trend of AA mortality changes, we have extracted data of absolute death number, mortality rate per 100,000 persons, and age-standardized death rate (ASDR). ASDR is a measure that adjusts for differences in age distribution within a population. Older populations naturally have higher death rates due to aging-related health issues. By standardizing death rates across age groups, ASDR allows for fair comparisons between different countries or time periods, ensuring that differences in mortality are not simply due to variations in age structure.

The formula for ASDR per 100,000 is 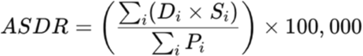

Where: D_i_ = Number of deaths in age group i; S_i_ = Standard population for age group i (from a reference population); P_i_ = Total population in age group i; 100,000 = Scaling factor to express the rate per 100,000 people.

We did not evaluate data of percentage of total death, trying to avoid the influence of deaths from COVID-19 pandemic since late 2019. Disability-adjusted Life Years (DALYs) combine the impact of both premature death and disability caused by the disease^12^. However, since a number of AA cases are undiagnosed until aneurysm rupture or dissections, DALY may be influenced a lot by screening policy and health care access. Therefore, we did not evaluate DALY data, but focused on ASDR for the comparison of different population and over the time period.

In order to evaluate the contribution changes in risk factors for AA-related death, we extracted the percent contribution of risk factors for AA-related mortality, and also the ASDR attributable to individual risk factors. GBD 2021 has used comprehensive methods to estimate the association between each risk factor and disease burden^19^. Each association is considered separately; therefore, the combined height of multiple bar segments of this graph may not be 100%.

### Studied regions, countries and populations

Following analysis of global changes in AA-related mortality and its risk factors. We have evaluated data from four regions that GBD Study 2021 provides (Europe, America, Asia and Africa). We also assessed data from regions defined according to the health system levels: Advanced, Basic, Limited and Minimal Health System regions; regions defined by income including Commonwealth high-, middle- and low-Income regions; and regions with different Socio-Demographic index (SDI). The SDI categorizes populations based on income, education and fertility rates^20^.

We found that Europe and Asia had opposite trends of AA mortality changes over the years, so we have then evaluated the ASDR for AA in different regions and countries in Europe and Asia. The inclusion of countries in different parts of Europe and Asia was based on the classification in the GBD study. The data was further stratified by sex and age groups.

### Data analysis and visualization

The data extracted from GBD was imported into R (version 4.4.2) for visualization through graphs and descriptive analysis, providing a clear depiction of patterns and trends in the population of interest. Initial graphs were generated using ggplot2 package^21^ or GraphPad Prism version 10. All panels were assembled together using Adobe Illustrator 2025. The raw data for each figure are presented in Supplemental Digital Content.

To evaluate trends over time, percentage changes from 1990 to 2021 in the chosen metrics were calculated. Data are presented as mean with 95% uncertainty intervals (UIs), which represent the range between the upper and lower bounds. GBD data provides only summary statistics without raw individual-level data or sample size, which restricts deeper statistical inferences. We therefore only did Descriptive Statistics.

The difference between men and women in ASDR or global death rate by AA in Fig. 5A and 5B was computed as mean of men – mean of women. The lower and upper bounds of the 95%UI of each sex were back transformed to their corresponding SE to estimate mathematically correct UIs as follow: 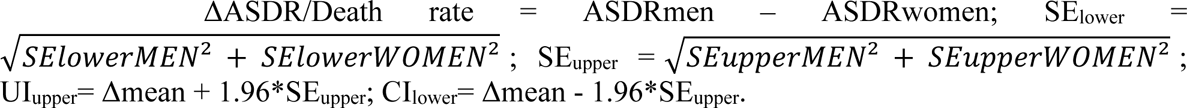

## Results

### Global mortality and age-standardized death rate of aortic aneurysm

In 2021, the global number of AA-related deaths was 153,927 (95% UI: 138,413-165,738), with an average increase of 74.2% from 88,353 (95% UI: 83090 – 93492) in 1990 (Fig. 1A; Table 1). Japan, India, and the United States reported the highest number of AA-related deaths in 2021 (Fig. 1B, Table 2). AA-related death number increased with age, peaking after age of 70 years old (Fig. 1C). However, these data are affected by population number in each country and in each age group.

**Figure 1.**
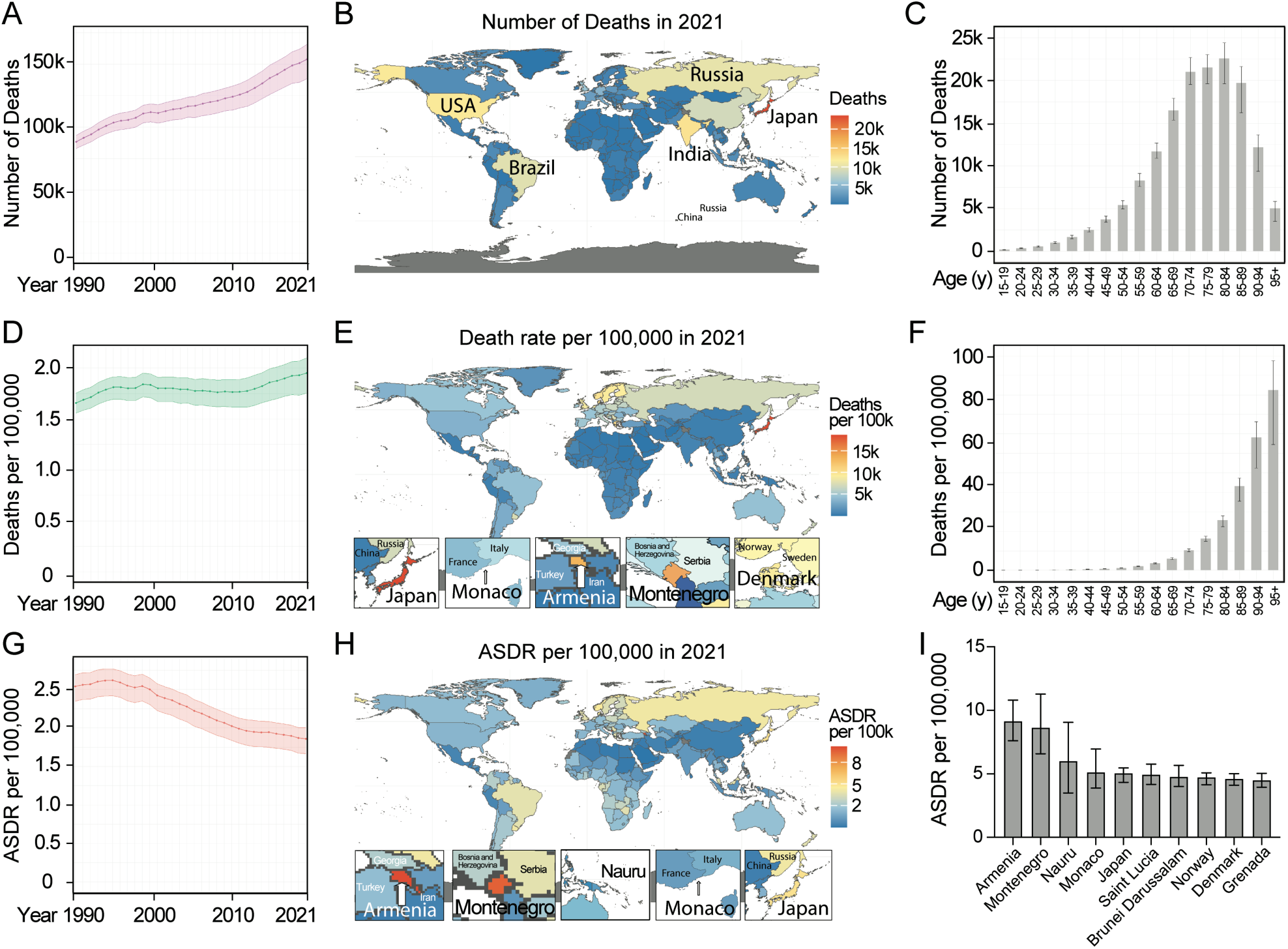
Global trends in AA-caused deaths and death rates. (A) Global number of AA deaths in all ages from 1990 to 2021. (B) Map of AA deaths in all ages in 2021. (C) AA deaths per age group in 2021. (D) Global AA death rate per 100,000 in all ages from 1990 to 2021. (E) Map of AA death rates per 100,000 in all ages in 2021. (F) Global AA death rates per 100,000 per age group in 2021. (G) Global AA-caused ASDR per 100,000 from 1990 to 2021. (H) Map of AA-caused ASDR in 2021. (I) AA-caused ASDR per 100,000 in top 10 countries. Data are shown as mean with 95% Uncertainty Intervals (UI). The top 5 countries with the highest AA-caused absolute death number (B), and mortality rate (E) and ASDR (H) per 100,000 persons in 2021 are marked in the map.

**Table 1.** Death cases and death rates in world regions in 1990 and 2021 along with the percent changes.

| Location | Death cases 1990 | Death cases 2021 | % change in death cases 1990 - 2021 | Death rate per 100.000 1990 | Death rate per 100.000 2021 | % change in death rate per 100.000 1990 - 2021 |
| --- | --- | --- | --- | --- | --- | --- |
| Global | 88353<br>(83090, 93492) | 153927<br>(138413, 165739) | 74.2<br>(63.0, 83.6) | 1.66<br>(1.56, 1.75) | 1.95<br>(1.75, 2.10) | 17.8<br>(10.2, 24.1) |
| **Four World Regions** |  |  |  |  |  |  |
| Africa | 4214<br>(2648, 6608) | 9125<br>(5666, 13820) | 116.6<br>(78.1, 158.1) | 0.67<br>(0.42, 1.05) | 0.66<br>(0.41, 1.00) | -1.2<br>(-18.8, 17.7) |
| America | 26779<br>(25150, 27685) | 31596<br>(28699, 33296) | 18.0<br>(13.2, 22.3) | 3.74<br>(3.51, 3.87) | 3.08<br>(2.79, 3.24) | -17.7<br>(-21.1, -14.7) |
| Asia | 16240<br>(13949, 19497) | 62840<br>(54678, 71295) | 287.0<br>(228.1, 349.3) | 0.51<br>(0.44, 0.61) | 1.36<br>(1.18, 1.54) | 165.8<br>(125.3, 208.6) |
| Europe | 40891<br>(38817, 41959) | 50082<br>(45990, 53067) | 22.5<br>(16.9, 28.3) | 5.10<br>(4.84, 5.23) | 5.90<br>(5.42, 6.25) | 15.8<br>(10.5, 21.3) |

Data are presented in Tables as mean with their 95% UIs with upper and lower bounds.

**Table 2.** Death cases and death rates in 1990 and 2021 in countries with highest burden of AA along with the percent changes.

| Location | Death cases 1990 | Death cases. 2021 | % change in death cases. 1990, 2021 | Location | Death rate per 100.000 1990 | Death rate per 100.000. 2021 | % change in death rate per 100.000. 1990, 2021 |
| --- | --- | --- | --- | --- | --- | --- | --- |
| Japan | 4797<br>(4448, 4982) | 23815<br>(19180, 26463) | 396.4<br>(327.8, 435.2) | Japan | 3.81<br>(3.53, 3.96) | 18.65<br>(15.02, 20.72) | 389.2<br>(321.6, 427.4) |
| India | 2652<br>(1578, 4344) | 12805<br>(9107, 18763) | 382.9<br>(258.3, 576.7) | Monaco | 14.76<br>(11.12, 18.41) | 15.46<br>(11.68, 20.70) | 4.7<br>(-26.0, 54.8) |
| United States | 17399<br>(16107, 18106) | 12195<br>(10940, 12944) | -29.9<br>(-32.6, -27.6) | Armenia | 3.43<br>(2.81, 4.16) | 13.24<br>(11.02, 15.60) | 286.4<br>(191.4, 415.7) |
| Russia | 4699<br>(4559, 4800) | 10445<br>(9555, 11307) | 122.3<br>(104.4, 139.8) | Montenegro | 6.45<br>(5.16, 8.31) | 13.09<br>(9.91, 17.03) | 102.8<br>(46.3, 193.2) |
| Brazil | 2857<br>(2739, 2952) | 10010<br>(9186, 10566) | 250.4<br>(229.5, 269.1) | Denmark | 11.18<br>(10.30, 11.98) | 10.54<br>(9.35, 11.49) | -5.7<br>(-15.5, 4.7) |

Data are presented in Tables as mean with their 95% UIs with upper and lower bounds.

We therefore explored the global mortality rate of AA per 100,000 population. It increased from an average of 1.66 (95% UI: 1.56 – 1.75) deaths in 1990 to 1.95 (95% UI: 1.75 – 2.10) deaths in 2021 in all ages per 100,000 population (17.47% increase) (Fig. 1D, Table 1). Japan, Monaco, Armenia, Montenegro and Denmark ranked high in the mortality rate of AA in 2021 (Fig. 1E, Table 2). The global mortality rate also increased with age, peaking at the highest age (Fig. 1F), indicating that the mortality rate can be affected by the age of the population.

To remove the confounding effects caused by age, we explored data of age-standardized death rate (ASDR). As shown in Figure 1G and Table 3, after age standardization, the global ASDR decreased from an average of 2.54 (95% UI: 2.35 – 2.69) deaths in 1990 to an average of 1.87 (95% UI: 1.67 – 2.00) deaths in 2021 per 100,000 population (26.69% decrease). Armenia, Montenegro, Nauru, Monaco, Japan, Saint Lucia, Brunei Darussalam, Norway, Denmark and Grenada are the top ten countries with highest burden of AA-caused death after age standardization in 2021 (Fig. 1H-1I, Table 4).

**Table 3.** Age-standardized death rate (ASDR) in world regions in 1990 and 2021 along with the percent changes.

| Location | ASDR per 100.000 1990 | ASDR per 100.000 2021 | % change in ASDR 1990, 2021 |
| --- | --- | --- | --- |
| Global | 2.54 (2.35, 2.69) | 1.87 (1.67, 2.00) | -26.7 (-31.1, -23.1) |
| Female | 1.58 (1.41, 1.76) | 1.28 (1.10, 1.42) | -18.8 (-26.0, -12.8) |
| Male | 3.87 (3.61, 4.18) | 2.57 (2.36, 2.79) | -33.6 (-37.1, -29.8) |
| **Four World Regions** |  |  |  |
| Europe | 3.95 (3.74, 4.06) | 2.97 (2.76, 3.15) | -24.8 (-27.9, -21.2) |
| America | 4.43 (4.14, 4.59) | 2.33 (2.12, 2.45) | -47.4 (-49.3, -45.6) |
| Africa | 1.82 (1.14, 2.82) | 1.65 (1.04, 2.47) | -9.1 (-24.2, 6.3) |
| Asia | 1.01 (0.87, 1.18) | 1.40 (1.20, 1.59) | 39.5 (20.4, 59.6) |
| **Health System Grouping** |  |  |  |
| Advanced | 4.15 (3.90, 4.28) | 2.98 (2.68, 3.16) | -28.1 (-31.3, -25.1) |
| Minimal | 1.71 (0.93, 3.14) | 1.70 (0.84, 3.09) | -0.9 (-21.0, 20.2) |
| Limited | 0.99 (0.73, 1.45) | 1.32 (1.06, 1.83) | 33.6 (6.4, 66.0) |
| Basic | 1.02 (0.94, 1.11) | 1.05 (0.95, 1.15) | 2.7 (-10.3, 15.1) |
| **Commonwealth** |  |  |  |
| High income | 8.66 (8.18, 8.94) | 3.22 (2.86, 3.40) | -62.8 (-64.8, -61.6) |
| Low income | 1.27 (0.80, 2.11) | 1.53 (0.98, 2.38) | 19.9 (-10.8, 63.5) |
| Middle income | 1.06 (0.83, 1.45) | 1.39 (1.13, 1.85) | 30.6 (8.1, 56.9) |
| **SDI** |  |  |  |
| High | 4.76 (4.46, 4.91) | 2.87 (2.51, 3.06) | -39.8 (-43.5, -37.2) |
| High-middle | 1.99 (1.88, 2.08) | 1.79 (1.66, 1.92) | -10.1 (-17.3, -3.6) |
| Low | 1.37 (0.83, 2.37) | 1.48 (0.91, 2.44) | 8.4 (-12.0, 31.9) |
| Low-middle | 0.89 (0.71, 1.20) | 1.31 (1.09, 1.76) | 47.9 (22.9, 73.0) |
| Middle | 1.03 (0.94, 1.14) | 1.15 (1.04, 1.25) | 12.5 (-1.4, 25.3) |

Data are presented in Tables as mean with their 95% UIs with upper and lower bounds.

**Table 4.** Age-standardized death rate (ASDR) and its percent changes in 1990 and 2021 in top countries.

| Location | ASDR per 100.000<br>1990 | ASDR per 100.000<br>2021 | % change in ASDR<br>1990, 2021 |
| --- | --- | --- | --- |
| Armenia | 4.55 (3.71, 5.57) | 9.16 (7.61, 10.81) | 101.4 (51.4, 170.2) |
| Montenegro | 6.70 (5.35, 8.64) | 8.65 (6.59, 11.28) | 29.2 (-7.8, 89.2) |
| Nauru | 4.92 (3.95, 6.24) | 6.01 (3.50, 9.06) | 22.3 (-27.0 – 89.0) |
| Monaco | 5.66 (4.29, 7.02) | 5.15 (3.88, 6.95) | -9.1 (-34.8, 33.6) |
| Japan | 2.92 (2.68, 3.04) | 5.07 (4.33, 5.47) | 73.6 (60.4, 81.6) |
| Saint Lucia | 10.31 (9.41, 11.56) | 4.95 (4.17, 5.77) | -52.0 (-60.5, -42.7) |
| Brunei Darussalam | 5.61 (4.47, 6.89) | 4.79(4.00, 5.67) | -14.7 (-35.6, 15.0) |
| Norway | 7.58 (7.16, 7.90) | 4.75 (4.15, 5.08) | -37.4 (-41.8, -34.0) |
| Denmark | 6.71 (6.21, 7.17) | 4.61 (4.10, 5.02) | -31.2 (-38.2, -23.9) |
| Grenada | 4.77 (4.15, 5.70) | 4.51 (3.94, 5.04) | -5.4 (-24.0, 13.4) |

Data are presented in Tables as mean with their 95% UIs with upper and lower bounds.

These data indicate that the global burden of AA-related mortality has been increasing over the time from 1990 to 2021. However, the increase in global population and life span have contributed to the increased mortality of AA. After age standardization, the global ASDR of AA decreased. We therefore decided to use ASDR to evaluate the difference in AA-related mortality among different populations in the world across time and sex.

### Age-standardized death rate of aortic aneurysm in world regions

We next analyzed ASDR of AA among different regions in the world. From 1990 to 2021, AA-caused ASDR decreased 24.8% in Europe and 47.4% in America, but increased 38.6% in Asia. There was big variation among African countries, and the average ASDR did not change much (Fig. 2A, Table 3). Although it decreased, Europe still had the highest ASDR caused by AA (2.97 deaths per 100,000 (95% UI: 2.76 - 3.15)) in 2021, which was followed by America (2.33 deaths per 100,000 (95% UI: 2.12 – 2.45)). Despite a continuous increase, the ASDR of AA remained at lower levels in Asia (1.4 deaths per 100,000 (95% UI: 1.2 – 1.59)), similar as in Africa (1.65 deaths per 100,000 (95% UI: 1.04 – 2.47)) (Fig. 2A and 2E, Table 3).

**Figure 2.**
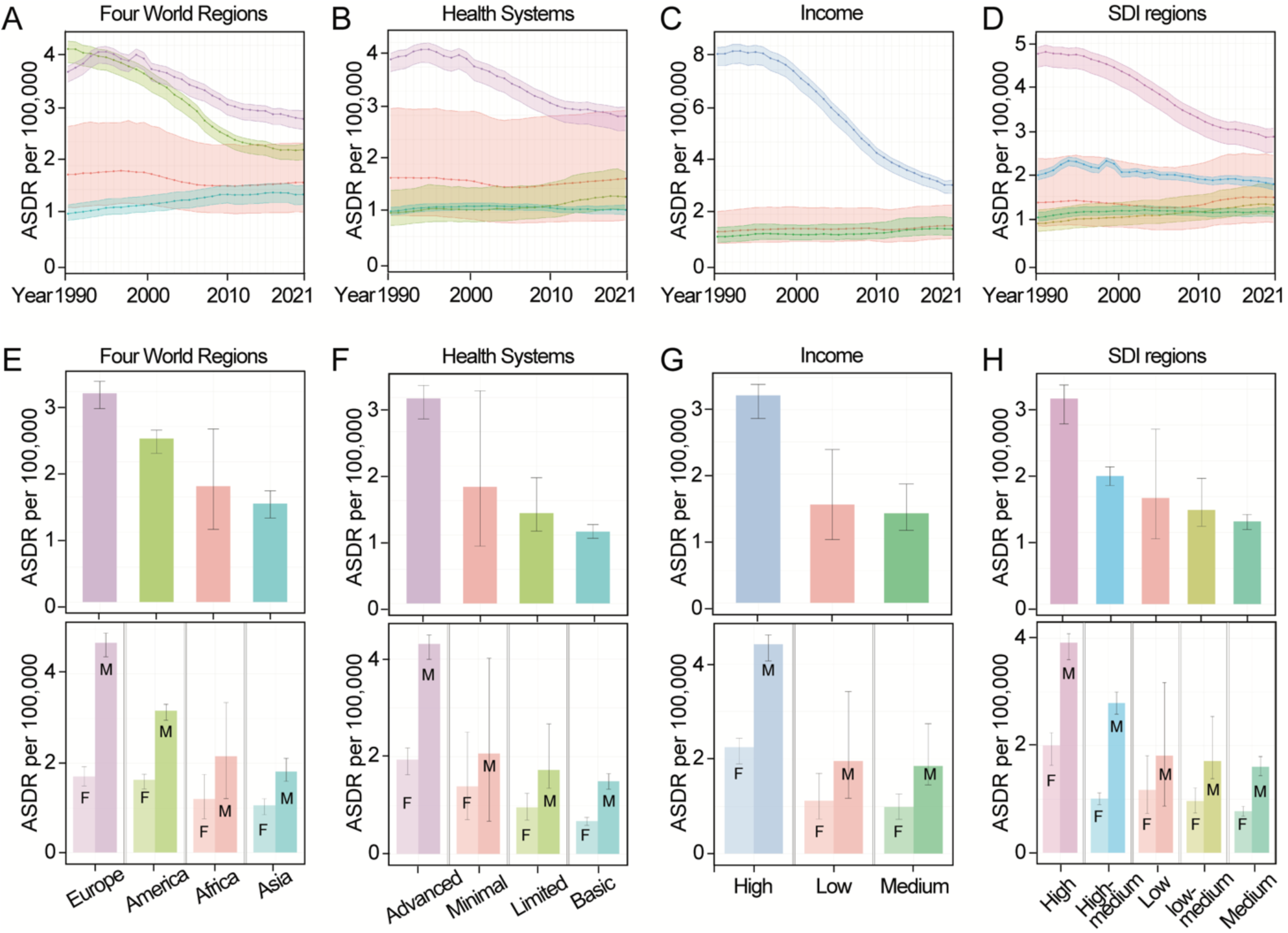
AA-caused ASDR among world regions. (A-D) ASDR of AA per 100,000 analyzed by Four World Regions, Health System Grouping, Commonwealth (Income) and SDI regions from 1990 to 2021. (E-H) ASDR of AA per 100,000 analyzed by Four World Regions, Health System Grouping, Commonwealth (Income) and SDI and their subgroups, stratified by sex in 2021. All regions are denoted by color codes. Data are shown as mean with 95% UI.

Interestingly, ASDR of AA declined from 1990 to 2021 in world regions which had advanced health system (Fig. 2B), high income (Fig. 2C) and high Socio-demographic Index (SDI) (Fig. 2D). However, these regions still had the highest AA-caused ASDR (Fig. 2B-2D). The other regions, with basic to limited health system, middle-low income and SDI, had constantly lower AA-caused ASDR (Fig. 2, Table 3).

Since ASDR of AA decreased in Europe but increased in Asia, we will explore European and Asian countries in detail to understand the burden of AA and its underlying reasons.

### Age-standardized death rate of aortic aneurysm in Europe

In order to understand further ASDR of AA in Europe, we analyzed three GBD European regions, Western, Central and Eastern Europe. The countries that belong to each of the European regions are listed in Table 5. As shown in Fig. 3A, ASDR of AA decreased significantly in Western Europe from 1990 to 2021. It also decreased in Central Europe from 2008 to 2021, but increased in Eastern Europe from 2004 to 2021. In 2021, Eastern Europe had higher ASDR of AA than Western Europe and Central Europe (Fig. 3A and 3E, Table 5).

**Figure 3.**
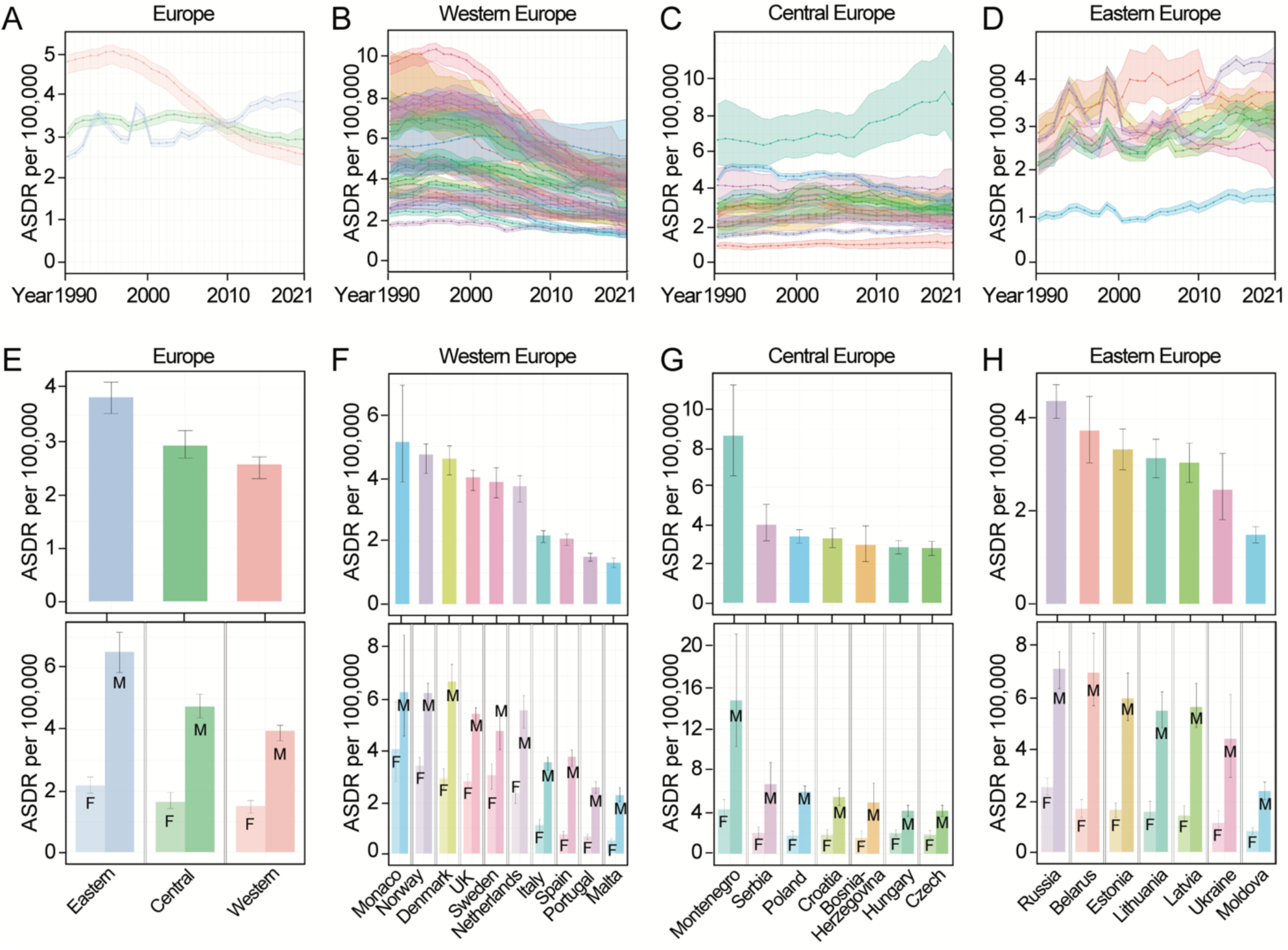
AA-caused ASDR in Europe. (A-D) AA-caused ASDR per 100,000 in Europe, Western Europe, Central Europe and Eastern Europe from 1990 to 2021. (E-H) AA-caused ASDR per 100,000 in European regions and representative countries in 2021. Data for both sexes are shown in upper panel, and stratified data by sex are shown in lower panel. Various regions and countries are denoted by color codes. F: women; M: Men. Data are shown as mean with 95% UI.

**Table 5.** Age-standardized death rate (ASDR) in Europe in 1990 and 2021 along with the percent changes.

| Location | ASDR per 100.000<br>1990 | ASDR per 100.000<br>2021 | % change in ASDR<br>1990, 2021 |
| --- | --- | --- | --- |
| **Western Europe** | 4.78 (4.50, 4.92) | 2.57 (2.30, 2.71) | -46.2 (-48.9, -44.2) |
| Monaco | 5.66 (4.29, 7.02) | 5.15 (3.88, 6.95) | -9.1 (-34.8, 33.6) |
| Norway | 7.58 (7.16, 7.90) | 4.75 (4.15, 5.08) | -37.4 (-41.8, -34.0) |
| Denmark | 6.71 (6.21, 7.17) | 4.61 (4.10, 5.02) | -31.2 (-38.2, -23.9) |
| United Kingdom | 9.72 (9.16, 10.00) | 4.03 (3.61, 4.25) | -58.6 (-60.9, -57.3) |
| Greece | 3.81 (3.55, 4.03) | 3.99 (3.65, 4.26) | 4.7 (-3.7, 13.4) |
| Cyprus | 7.82 (6.01, 10.06) | 3.96 (3.04, 4.88) | -49.3 (-64.9, -25.7) |
| Andorra | 6.87 (4.36, 10.33) | 3.96 (2.50, 5.97) | -42.4 (-62.7, -11.6) |
| Sweden | 7.41 (6.91, 7.79) | 3.88 (3.38, 4.33) | -47.6 (-52.8, -42.2) |
| Netherlands | 7.44 (6.89, 7.96) | 3.74 (3.24, 4.08) | -49.7 (-54.1, -45.2) |
| Finland | 7.88 (7.35, 8.36) | 3.63 (3.20, 3.93) | -53.9 (-58.3, -49.3) |
| Ireland | 6.36 (5.94, 6.78) | 2.93 (2.47, 3.25) | -53.9 (-59.8, -48.7) |
| Iceland | 4.34 (3.91, 4.65) | 2.61 (2.23, 2.92) | -39.8 (-46.5, -32.2) |
| Switzerland | 4.87 (4.43, 5.26) | 2.28 (1.96, 2.51) | -53.2 (-58.3, -47.6) |
| Belgium | 5.11 (4.73, 5.45) | 2.25 (1.96, 2.43) | -55.9 (-59.7, -52.2) |
| Germany | 3.32 (3.07, 3.56) | 2.25 (2.02, 2.41) | -32.4 (-38.3, -25.5) |
| Luxembourg | 4.33 (4.06, 4.64) | 2.23 (1.97, 2.46) | -48.7 (-54.0, -42.6) |
| Italy | 3.10 (2.91, 3.21) | 2.18 (1.95, 2.34) | -29.6 (-33.6, -25.7) |
| Spain | 2.54 (2.37, 2.68) | 2.08 (1.87, 2.23) | -18.3 (-24.2, -12.4) |
| France | 3.65 (3.41, 3.85) | 1.89 (1.67, 2.04) | -48.1 (-53.0, -44.0) |
| Austria | 3.20 (3.03, 3.37) | 1.80 (1.61, 1.94) | -43.7 (-47.9, -39.4) |
| San Marino | 3.09 (2.49, 3.76) | 1.67 (1.06, 2.59) | -46.0 (-67.1, -13.3) |
| Portugal | 1.75 (1.64, 1.86) | 1.51 (1.37, 1.62) | -14.1 (-21.1, -7.0) |
| Malta | 2.60 (2.41, 2.80) | 1.32 (1.17, 1.47) | -49.4 (-55.2, -43.0) |
| Israel | 2.34 (2.15, 2.50) | 1.28 (1.13, 1.40) | -45.1 (-50.9, -38.7) |
| **Central Europe** | 3.07 (2.95, 3.18) | 2.93 (2.69, 3.21) | -4.8 (-12.2, 4.6) |
| Montenegro | 6.70 (5.35, 8.64) | 8.65 (6.59, 11.28) | 29.2 (-7.8, 89.2) |
| Serbia | 4.22 (3.50, 5.14) | 4.05 (3.21, 5.10) | -4.2 (-28.5, 24.7) |
| Poland | 4.54 (4.36, 4.66) | 3.44 (3.09, 3.79) | -24.2 (-31.1, -16.4) |
| Croatia | 2.79 (2.45, 3.19) | 3.34 (2.87, 3.86) | 19.7 (-3.4, 46.5) |
| Bosnia and Herzegovina | 2.12 (1.55, 2.89) | 3.00 (2.15, 3.99) | 41.6 (-12.1, 127.1) |
| Hungary | 3.23 (3.00, 3.48) | 2.87 (2.54, 3.23) | -11.2 (-22.0, -0.1) |
| Czech Republic | 2.96 (2.76, 3.18) | 2.83 (2.46, 3.19) | -4.2 (-17.6, 10.1) |
| North Macedonia | 1.94 (1.51, 2.42) | 2.63 (1.80, 3.81) | 36.0 (-10.4, 97.7) |
| Bulgaria | 2.03 (1.82, 2.23) | 2.62 (2.12, 3.23) | 28.8 (-0.1, 66.1) |
| Slovakia | 2.29(1.97, 2.68) | 2.29 (1.79, 2.90) | -0.01 (-28.2, 38.1) |
| Slovenia | 2.66 (2.47, 2.84) | 2.16 (1.82, 2.58) | -19.0 (-31.3, -2.8) |
| Romania | 1.40 (1.30, 1.52) | 1.90 (1.65, 2.18) | 35.9 (15.1, 58.9) |
| Albania | 0.91 (0.76, 1.08) | 1.09 (0.76, 1.50) | 19.2 (-20.2, 76.7) |
| **Eastern Europe** | 2.52(2.43, 2.62) | 3.82 (3.52, 4.12) | 51.8 (38.9, 64.0) |
| Russian Federation | 2.72 (2.62, 2.78) | 4.38 (4.01, 4.74) | 61.3 (48.2, 74.0) |
| Belarus | 2.67 (2.31, 3.23) | 3.75 (3.05, 4.49) | 40.5 (10.1, 81.4) |
| Estonia | 2.86 (2.60, 3.13) | 3.34 (2.90, 3.78) | 16.9 (0.01, 37.9) |
| Lithuania | 2.16 (1.97, 2.36) | 3.15 (2.73, 3.56) | 46.0 (26.2, 69.3) |
| Latvia | 2.21 (2.03, 2.40) | 3.05 (2.63, 3.47) | 38.2 (18.9, 60.5) |
| Ukraine | 2.13 (1.92, 2.37) | 2.47 (1.82, 3.25) | 15.9 (-18.5, 56.0) |
| Republic of Moldova | 0.96 (0.89, 1.06) | 1.49 (1.32, 1.67) | 54.9 (33.6, 81.1) |

Data are presented in Tables as mean with their 95% UIs with upper and lower bounds.

Almost all the Western European countries had decreased ASDR of AA from 1990 to 2021 (Fig. 3B). Significant decrease was specially observed in UK, Norway, Sweden, Netherlands, Denmark, Finland, France, and Iceland. However, in 2021, the ASDR of AA was still higher in these countries compared with other Western European countries, such as Portugal, Italy, Spain, and Malta. Monaco had marginal change since 1990, but it had the highest ASDR of AA among the Western European countries in 2021 (Fig. 3B and 3F, Table 5).

Most Central European countries had relatively stable ASDR of AA from 1990 to 2021, with the exception of Montenegro, where the ASDR of AA increased from 2008 to 2021, making it the highest ASDR in Europe (Fig. 3C and 3G, Table 5).

The ASDR of AA increased constantly in most Eastern European countries, with the exception of Ukraine where had almost no change (Fig. 3D and 3H, Table 5).

### Age-standardized death rate of aortic aneurysm in Asia

We also explored data among Asian countries using existing GBD regions including Central Asia, South Asia, East Asia, and Southeast Asia, together with data from Japan, Republic of Korea, and Singapore. The ASDR of AA increased constantly in Japan from 1990 to 2021, making it the country with the highest mortality in all ages in the world and the highest ASDR (5.07 deaths per 100,000 (95% UI: 4.33 – 5.47)) in Asia in 2021, almost twice as much as the other Asian countries. Singapore had similar ASDR of AA as in Japan in 1990, but it decreased during the years, ending in 2.15 (95% UI: 1.91 – 2.32) deaths per 100,000 in 2021 (Fig. 4A and 4F, Table 6).

**Figure 4.**
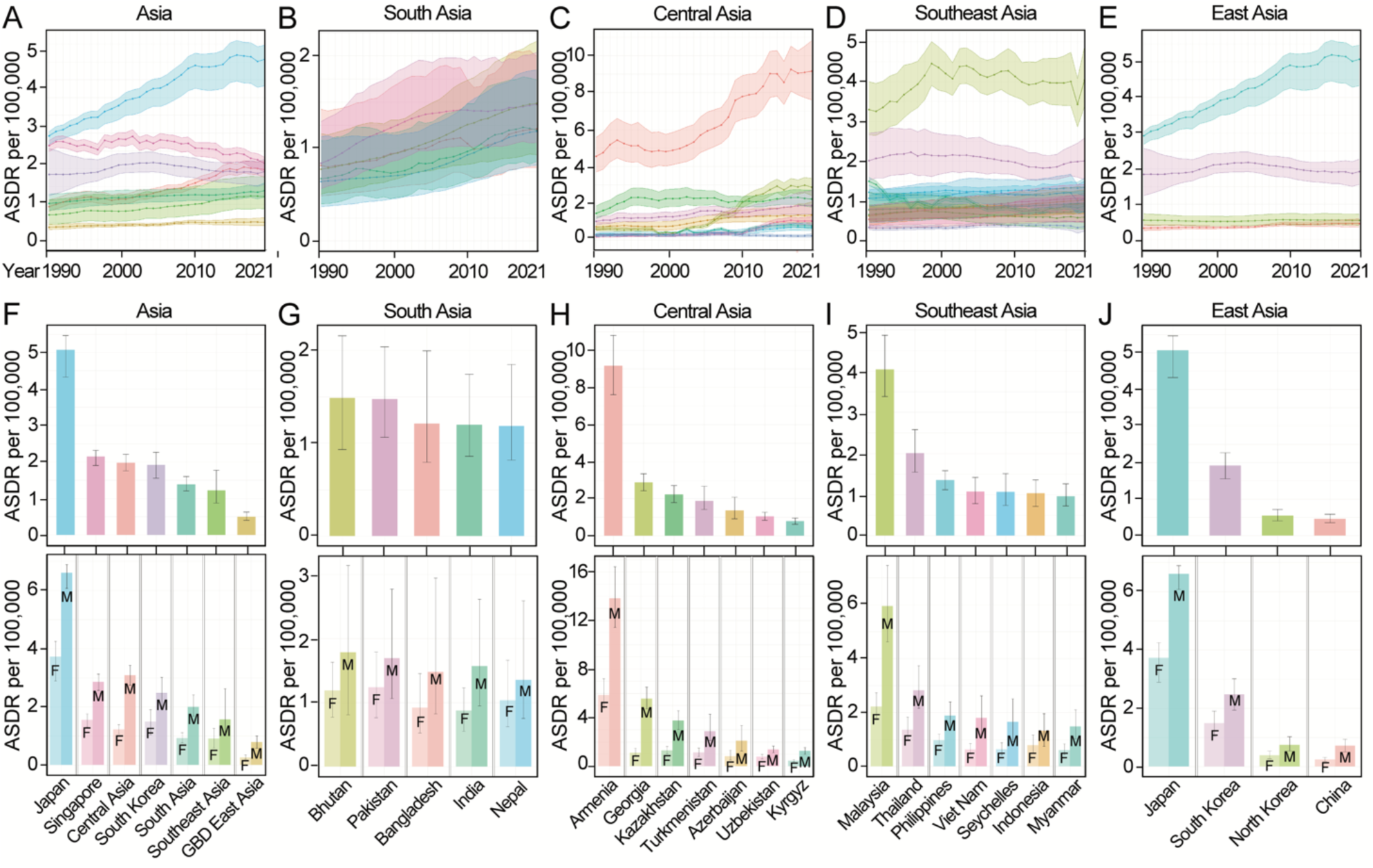
AA-caused ASDR in Asia. (A-E) AA-caused ASDR per 100,000 in Asia, South Asia, Central Asia, Southeast Asia and East Asia from 1990 to 2021. (F-J) AA-caused ASDR per 100,000 in Asian regions and representative countries in 2021. Data for both sexes are shown in upper panel, and stratified data by sex are shown in lower panel. Various regions and countries are denoted by color codes. East Asia in GBD 2021 study (GBD East Asia) contains China and North Korea. F: women; M: Men. Data are shown as mean with 95% UI.

**Table 6.** Age-standardized death rate (ASDR) in Asia in 1990 and 2021 along with the percent changes.

| Location | ASDR per 100.000<br>1990 | ASDR per 100.000<br>2021 | % change in ASDR<br>1990, 2021 |
| --- | --- | --- | --- |
| **South Asia** | 0.71 (0.45, 1.13) | 1.22 (0.88, 1.78) | 71.6 (27.9, 136.1) |
| **Bhutan** | 0.78 (0.51, 1.16) | 1.49 (0.93, 2.16) | 91.6 (5.3, 201.2) |
| **Pakistan** | 0.84 (0.57, 1.29) | 1.48 (1.06, 2.04) | 75.8 (21.9, 148.4) |
| **Bangladesh** | 0.82 (0.49, 1.42) | 1.21 (0.80, 1.99) | 48.6 (2.2, 131.6) |
| **India** | 0.68 (0.40, 1.08) | 1.20 (0.86, 1.74) | 77.5 (30.0, 149.6) |
| **Nepal** | 0.64 (0.37, 1.13) | 1.19 (0.82, 1.85) | 84.6 (24.4, 177.4) |
| **Central Asia** | 0.94 (0.81, 1.13) | 1.98 (1.77, 2.21) | 110.1 (71.5, 150.9) |
| **Armenia** | 4.55 (3.71, 5.57) | 9.16 (7.61, 10.81) | 101.4 (51.4, 170.2) |
| **Georgia** | 0.61 (0.51, 0.72) | 2.88 (2.43, 3.35) | 374.2 (260.2, 510) |
| **Kazakhstan** | 1.46 (1.19, 1.87) | 2.23 (1.80, 2.71) | 52.8 (8.1, 112.2) |
| **Turkmenistan** | 1.03 (0.84, 1.26) | 1.88 (1.42, 2.67) | 83.0 (21.2, 165.9) |
| **Azerbaijan** | 0.73 (0.55, 0.96) | 1.37 (0.93, 2.08) | 87.7 (15.1, 189.9) |
| **Uzbekistan** | 0.21 (0.16, 0.30) | 1.04 (0.85, 1.28) | 386.0 (225.1, 595) |
| **Kyrgyzstan** | 0.28 (0.24, 0.33) | 0.79 (0.63, 0.96) | 185.5 (114, 272.3) |
| **Mongolia** | 0.31 (0.22, 0.42) | 0.70 (0.53, 0.89) | 124.9 (50.1, 251.8) |
| **Tajikistan** | 0.28 (0.19, 0.36) | 0.25 (0.18, 0.34) | -10.0 (-39.0, 38.3) |
| **Southeast Asia** | 1.03 (0.83, 1.29) | 1.39 (1.21, 1.60) | 34.6 (5.7, 70.2) |
| **Malaysia** | 3.30 (2.66, 4.00) | 4.04 (3.38, 4.88) | 22.4 (-9.2, 65.7) |
| **Thailand** | 2.02 (1.53, 2.71) | 2.01 (1.55, 2.58) | -0.4 (-32.8, 44.4) |
| **Philippines** | 1.22 (1.07, 1.39) | 1.35 (1.12, 1.58) | 10.6 (-11.1, 36.1) |
| **Viet Nam** | 0.67 (0.48, 0.90) | 1.07 (0.78, 1.42) | 60.4 (12.1, 146.4) |
| **Seychelles** | 1.08 (0.88, 1.29) | 1.07 (0.74, 1.51) | -0.8 (-28.8, 37.9) |
| **Indonesia** | 0.63 (0.41, 0.87) | 1.03 (0.71, 1.36) | 62.2 (4.4, 130.4) |
| **Myanmar** | 0.80 (0.47, 1.16) | 0.96 (0.73, 1.26) | 20.8 (-18.5, 84.7) |
| **Lao People's Democratic Republic** | 0.78 (0.51, 1.22) | 0.93 (0.68, 1.28) | 18.2 (-18.3, 79.2) |
| **Mauritius** | 1.50 (1.40, 1.60) | 0.88 (0.81, 0.94) | -41.3 (-46.2, -36.7) |
| **Cambodia** | 0.55 (0.32, 0.87) | 0.75 (0.47, 1.21) | 34.8 (-7.5, 107.2) |
| **Timor-Leste** | 0.54 (0.38, 0.82) | 0.74 (0.49, 1.15) | 37.0 (-6.3, 100.3) |
| **Maldives** | 0.66 (0.47, 0.91) | 0.60 (0.31, 0.97) | -9.5 (-47.9, 53.2) |
| **Sri Lanka** | 0.35 (0.29, 0.42) | 0.37 (0.25, 0.50) | 5.8 (-32.9, 66.8) |
| **GBD East Asia** | 0.36 (0.29, 0.44) | 0.50 (0.41, 0.63) | 40.0 (-1.6 – 92.0) |
| **North Korea** | 0.55 (0.41, 0.73) | 0.55 (0.41, 0.72) | -1.3 (-29.0, 42.0) |
| **China** | 0.33 (0.27, 0.42) | 0.46 (0.36, 0.59) | 37.2 (-7.5, 95.7) |
| **Japan** | 2.92 (2.68, 3.04) | 5.07 (4.33, 5.47) | 73.6 (60.4, 81.6) |
| **Singapore** | 2.65 (2.49, 2.79) | 2.15 (1.91, 2.32) | -18.9 (-26.7, -11.6) |
| **Republic of Korea** | 1.84 (1.25, 2.57) | 1.92 (1.56, 2.27) | 4.1 (-34.2, 68.9) |

Data are presented in Tables as mean with their 95% UIs with upper and lower bounds.

ASDR of AA also increased in South Asian countries including Bhutan, Pakistan, Bangladesh, India, and Nepal from 1990 to 2021, although the value had been kept at low levels (Fig. 4B and 4G, Table 6).

Most countries in Central Asia had no significant changes from 1990 to 2021, except Armenia and Georgia. Armenia had constantly much higher ASDR of AA throughout the years. In 2021, Armenia had the highest ASDR of AA in the world, with 9.16 deaths per 100,000 (Fig. 4C and 4H, Table 6).

China and Democratic People’s Republic of Korea (North Korea) from GBD East Asia and most countries in Southeast had constantly lower levels of ASDR of AA. However, Republic of Korea (South Korea) in East Asia and Malaysia and Thailand in Southeast Asia had much higher ASDR (Fig. 4D-4E, 4I-4J, Table 6).

### Sex differences in global mortality rate of aortic aneurysm

We next explored the sex differences in the disease burden of AA. The global ASDR of AA in men have always been higher than in women during 1990 through 2021. It decreased 33.6% in men from 3.87 (95% UI: 3.61 - 4.18) deaths in 1990 to 2.57 (95% UI: 2.36 - 2.79) deaths in 2021, and in women decreased 19% from 1.58 (95% UI: 1.41 - 1.76) to 1.28 (95% UI: 1.10 - 1.42) deaths in 100,000 population (Fig. 5A, Table 3). The difference between men and women decreased over the time from 1990 to 2021 (Fig. 5A).

**Figure 5.**
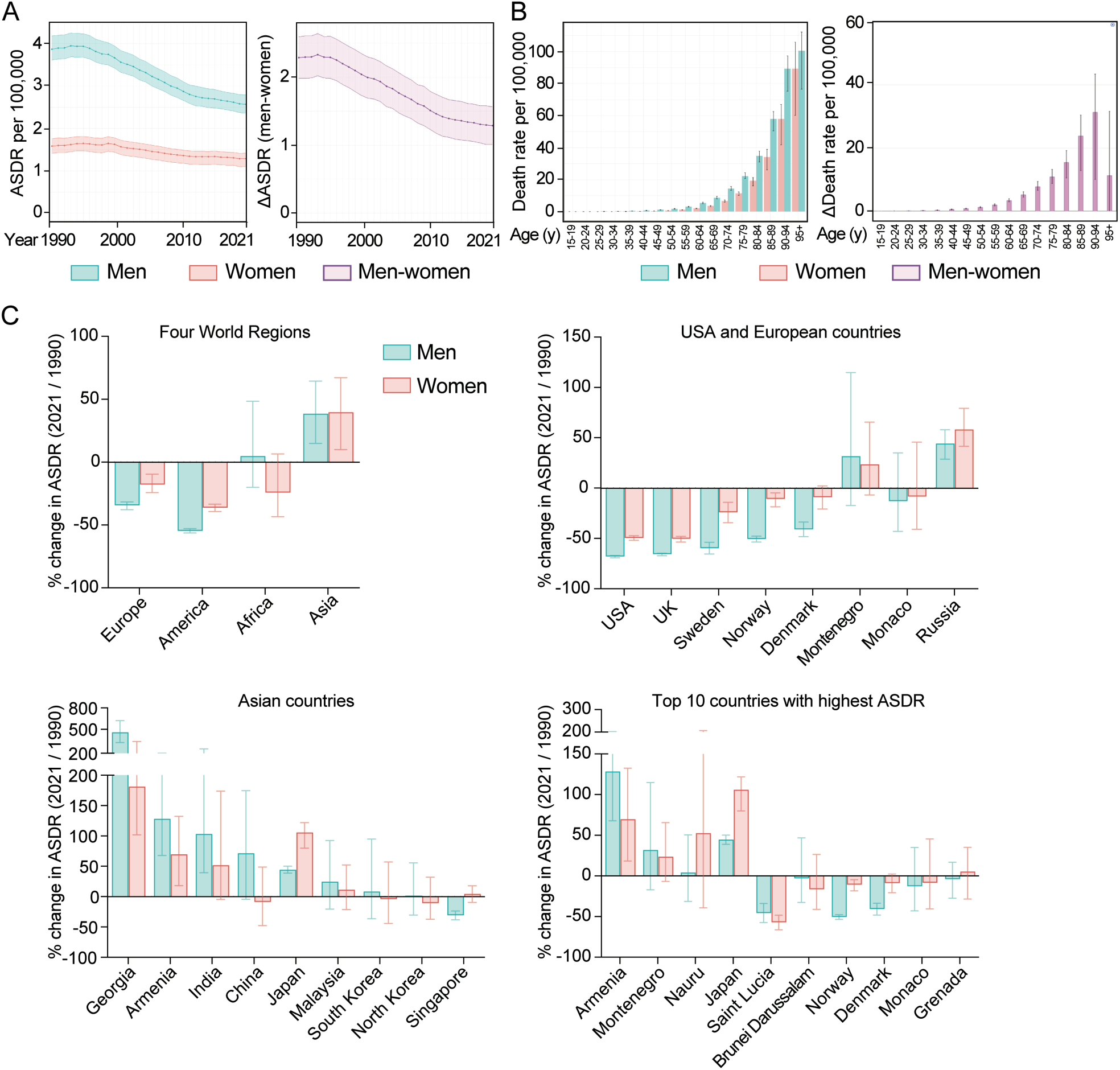
Sex differences in global AA death rates. (A) Global AA-caused ASDR per 100,000 in men (blue) and women (red), as well as the difference between men and women (pink, right panel) from 1990 to 2021. (B) Global death rates per 100,000 in men (blue) and women (red), as well as the difference between men and women (pink, right panel), stratified by age groups in 2021. (C) Percentage changes in AA-caused ASDR of men and women in 2021 compared to 1990, in the indicated world regions and countries. Data are shown as mean with 95% UI.

Mortality rate of AA increased with age in both men and women. Interestingly, the difference between men and women also increased with age (Fig. 5B). However, in the oldest age group of 95+ years, the sex difference diminished, likely due to decreased mortality among elderly men following interventions aligned with treatment guidelines of aortic disease^22^.

In 2021, ASDR of AA in men were about twice that of the women globally (Fig. 5A, Table 3), as well as in the four major regions of the world, Europe, America, Africa and Asia (Fig. 2E). However, the trends in AA-caused ASDR from 1990 to 2021 varied significantly between men and women across different countries (Fig. 5C). In Europe and America, ASDR generally declined more in men than in women. Notably, the differences were particularly pronounced in Sweden, Norway and Denmark, where men experienced percentage changes of −60.0%, −50.6% and −40.9%, respectively, compared to smaller declines in women (−24.1%, −10.8% and −9.0%). However, in Russa, Japan and Nauru, women had greater increase than men, with 58.5% vs 44.5% increase in Russa, 106.1% vs 44.7% increase in Japan, and 52.7% vs 4.5% increase in Nauru. In China, ASDR in men increased 71.6%, but it decreased 8.8% in women. Conversely, in Singapore, men had a 31.0% decrease while women experienced a slight increase of 4.7%. Among the countries with highest ASDR, Armenia increased 128.6% in men and 69.9% in women, however, whereas Monaco and Grenada maintained consistently high levels throughout the study period (Fig. 5C, Table 7).

**Table 7.** Percentage changes in AA-caused ASDR of men and women in 2021 compared to 1990.

| Four World Regions |  |  |
| --- | --- | --- |
| Location | Men | Women |
| Europe | -34.5 (-40.3, -28.2) | -17.9 (-30.9, 0.7) |
| America | -54.6 (-58.8, -50.0) | -36.2 (-47.0, -23.1) |
| Africa | -12.0 (-52.6, 140.7) | -33.3 (-65.7, 19.0) |
| Asia | 65.3 (-30.5, 203.1) | 58.9 (-22.9, 188.5) |
| USA and selected European countries |  |  |
| USA | -68.0 (-71.1, -64.4) | -49.5 (-58.7, -38.0) |
| UK | -65.6 (-69.1, -62.7) | -50.4 (-59.9, -40.0) |
| Sweden | -59.7 (-67.7, -51.1) | -24.2 (-41.7, -1.8) |
| Montenegro | 32.0 (-34.0, 155.7) | 23.8 (-23.2, 97.7) |
| Monaco | -13.0 (-51.1, 53.9) | -8.4 (-52.0, 94.2) |
| Norway | -50.6 (-57.5, -44.9) | -10.8 (-28.6, 8.5) |
| Denmark | -40.9 (-51.4, -29.0) | -9.0 (-28.5, 15.0) |
| Russia | 44.5 (26.1, 63.4) | 58.5 (36.0, 90.1) |
| Selected Asian Countries |  |  |
| Georgia | 478.1 (322.5, 672.4) | 181.5 (83.9, 380.0) |
| Armenia | 128.6 (51.1, 234.0) | 69.9 (5.7, 169.6) |
| India | 103.6 (-45.9, 511.5) | 51.9 (-53.1, 343.0) |
| China | 71.6 (-10.5, 241.5) | -8.8 (-52.6, 67.5) |
| Japan | 44.7 (28.3, 59.4) | 106.1 (49.6, 167.9) |
| Malaysia | 24.8 (-26.4, 113.0) | 11.5 (-27.8, 68.4) |
| South Korea | 8.6 (-43.4, 116.6) | -4.3 (-52.3, 83.5) |
| North Korea | 2.0 (-48.6, 106.8) | -10.8 (-53.9, 74.6) |
| Singapore | -31.0 (-42.0, -19.2) | 4.7 (-18.8, 29.9) |
| Top 10 countries with highest ASDR |  |  |
| Armenia | 128.6 (51.1, 234.0) | 69.9 (5.7, 169.6) |
| Montenegro | 32.0 (-34.0, 155.7) | 23.8 (-23.2, 97.7) |
| Nauru | 4.5 (-50.0, 139.3) | 52.7 (-45.3, 270.9) |
| Japan | 44.7 (28.3, 59.4) | 106.1 (49.6, 167.9) |
| Saint Lucia | -45.9 (-61.2, -29.6) | -57.2 (-68.9, -43.6) |
| Brunei Darussalam | -3.1 (-47.0, 90.7) | -16.6 (-47.8, 46.4) |
| Norway | -50.6 (-57.5, -44.9) | -10.8 (-28.6, 8.5) |
| Denmark | -40.9 (-51.4, -29.0) | -9.0 (-28.5, 15.0) |
| Monaco | -13.0 (-51.1, 53.9) | -8.4 (-52.0, 94.2) |
| Grenada | -4.2 (-36.2, 24.2) | 5.3 (-32.7, 44.7) |

Data are presented in Tables as mean of percentage changes with their 95% UIs with upper and lower bounds.

These data underscore the significant role of sex differences in AA-caused mortality. The impact of sex difference varies with age, being lower in younger and oldest age groups. It fluctuates across different countries over time, influencing the overall AA-caused mortality.

### Risk factors contributing to aortic aneurysm-related deaths

The risk factors that contributed to the ASDR of AA in 2021 were smoking (30%, 95% UI: 25.5 – 34.9%), high systolic blood pressure (17.3%, 95% UI: 13 – 21.9%), high body-mass index (7.4%, 95% UI: 4 – 12.7%), low consumption of fruits (3.6%, 95% UI: 2.5 – 4.8%) and vegetables (2.9%, 95% UI: 1.9 – 4%), high sodium intake (0.9%, 95% UI: 0.1 – 2.7%) and lead exposure (0.7%, 95% UI: −0.1 – 1.7%) (Fig. 6A). We analyzed the percentage contribution of these risk factors across different regions of the world and the top ten countries with the highest ASDR of AA for both men and women (Fig. 6B-6D). The percentage distribution of these risk factors to ASDR of AA in the world is shown in Fig. 6E.

**Figure 6.**
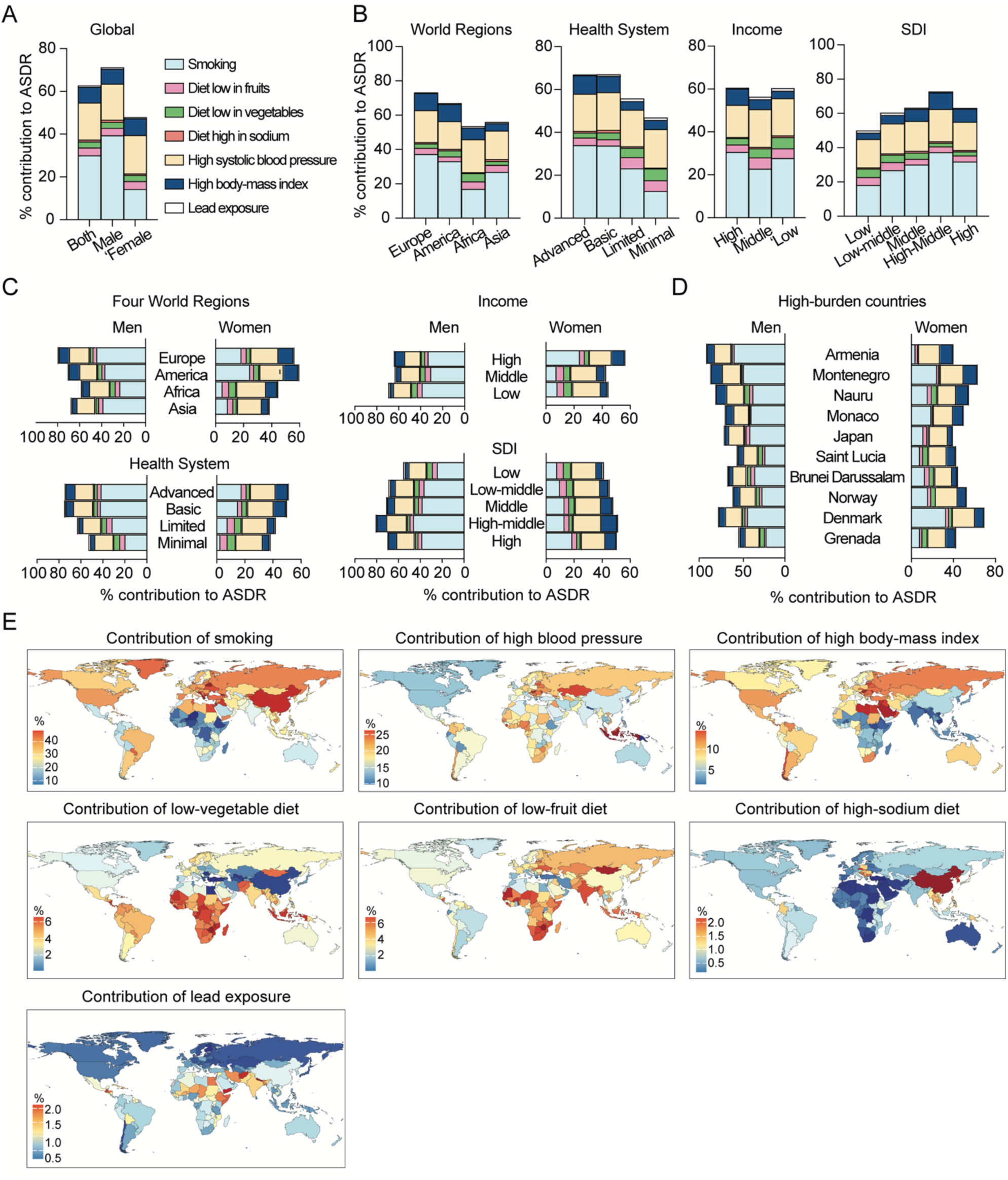
Percentage contribution of risk factors to AA-caused ASDR at global level in 2021. (A) Percentage contribution of various risk factors to global AA-ASDR, stratified by sex. (B) Percentage contribution of risk factors to AA-related ASDR among world regions, including Four World Regions, Health System Grouping, Commonwealth (Income) and SDI. (C) Percentage contribution of risk factors to AA-related ASDR analyzed by Four World Regions, Health System Grouping, Commonwealth (Income) and SDI, stratified by sex. (D) Percentage contribution of risk factors to AA-caused ASDR in top ten countries with the highest ASDR in the world. (E) Maps depicting the global percentage of AA deaths attributable to smoking, high systolic blood pressure, high body-mass index, diet low in vegetables, diet low in fruits, high consumption of sodium and lead exposure. Darker color in red indicates more contribution of the risk factor in that country. To the opposite, darker color in blue suggests less contribution of the risk factor in that country.

### Smoking

The biggest risk factor for AA-caused death was smoking. The percentage contribution of smoking to ASDR of AA was higher in regions with high disease burden of AA (Fig. 6B and 6E). For example, smoking contributed 37.2% (95% UI: 31.8 – 42.9%) and 33.1% (95% UI: 27.5 – 39%) of ASDR in Europe and America while as 16.9% (95% UI: 13.7 – 20.4%) in Africa, and it contributed 33.9% (95% UI: 28.6 – 39.3%) of ASDR in regions having Advanced Health System, but only 12.5% (95% UI: 7.2 – 17.5%) in regions with Minimal Health System (Fig. 6B). The contribution of smoking to AA-caused death was particularly high in the following countries such as Lebanon (48.5%), Georgia (48.1%), Belarus (47.8%), Greece (46.8%), Albania (46.3%), Jordan (46.3%), Bosnia and Herzegovina (45.5%), and China (44.7%) (Fig. 6E).

The percentage contribution of smoking to AA-caused ASDR was much higher in men (39.3%, 95% UI: 33.6 – 45.5%) than in women (14.2%, 95% UI: 11.2 – 17.4%) globally (Fig. 6A), a trend which was maintained among all the world regions and countries with high AA burden (Fig. 6C-6D). Smoking was the biggest risk factor for women in many countries with high burden of AA-caused ASDR, such as in Denmark (33.35%), Montenegro (24.78%), and Monaco (19.31%) (Fig. 6D).

### High systolic blood pressure

The second risk factor for AA-caused death was high systolic blood pressure, accounting for 17.3% (95% UI: 13 – 21.9%) of AA-caused ASDR globally (Fig. 6A). It contributed the most in Indonesia (26.3%), Hungary (25.9%), Republic of Moldova (24.7%), Sierra Leone (24.4%), Kazakhstan (23.9%), Lithuania (23.4%), Malaysia (23.4%), and Georgia (22.8%) (Fig. 6E).

High systolic blood pressure was the biggest risk factor for AA-caused death in women globally and in most of the regions that we have explored, except America and Commonwealth High region where smoking had the highest contribution (Fig. 6C). It was also the biggest risk factor for women in many countries with high burden of AA-caused ASDR, such as in Armenia, Nauru, Japan, Saint Lucia, Brunei Darussalam, Norway, and Grenada (Fig. 6D).

### High body-mass index

High body-mass index contributed to 7.4% (95% UI: 4 – 12.7%) of AA-caused ASDR globally. Its contribution to AA-caused ASDR was higher in regions with high disease burden of AA (Fig. 6B and 6E). For example, it contributed 10% (95% UI: 5.3 – 17.2%) and 9.9% (95% UI: 5.3 – 16.9%) of the AA-caused ASDR in Europe and America while as 4.4% (95% UI: 2.4 – 7%) in Asia, and it contributed 8.6% (95% UI: 4.6 – 14.7%) of the AA-caused ASDR in regions having Advanced Health System, but only 4.3% (95% UI: 2.4 – 7.1%) in regions with Minimal Health System (Fig. 6B). The contribution of high body-mass index to AA-caused death was particularly high in Middle East, Central and Eastern Europe and North Africa, such as in Qatar, Kuwait, Hungary, Saudi Arabia, Republic of Moldova, United Arab Emirates, Libya, and Syrian Arab Republic (Fig. 6E). In general, it played more important roles in women than in men for AA-caused ASDR.

### Diet with low fruits and vegetables but high salt

Diet with low consumption of fruits (3.6%, 95% UI: 2.5 – 4.8%) and vegetables (2.9%, 95% UI: 1.9 – 4%), but high sodium intake (0.9%, 95% UI: 0.1 – 2.7%) were also important risk factors for AA-caused ASDR globally (Fig. 6A and 6E). Their contributions were much lower than smoking, high blood pressure and high body-mass index. It was similar between men and women, and it did not vary a lot among the world regions with different burden of AA (Fig. 6B). However, they are important risk factors for particular countries.

Diet with low fruits or vegetables contributed the most to African countries, such as Zimbabwe, Togo, and Sierra Leone; Asian countries such as Mongolia and countries in Oceania such as Vanuatu (Fig. 6E). Diet with high salt played important roles in the burden of AA-caused death in China, Republic of Korea (South Korea), Democratic People’s Republic of Korea (North Korea), and Singapore in Asia, and in Central and Eastern (Balkans) European countries such as Czechia, Bosnia and Herzegovina, Bulgaria, North Macedonia, Slovakia, Montenegro (Fig. 6E).

### Lead exposure

Lead exposure contributed to 0.7% (95% UI: −0.1 – 1.7%) of AA-caused ASDR globally. Its contribution was particularly high in Southern and eastern Asian countries including Nepal, Yemen, Afghanistan, Bhutan, Bangladesh, Iran; in African countries such as Somalia, Egypt, Ethiopia; and in Central American countries such as Guatemala, Haiti, and Honduras (Fig. 6E).

### Risk factors contributing to aortic aneurysm-related deaths in Europe

In Europe in 2021, smoking was the biggest risk factor for AA-caused death in all the regions for both men and women, except for women in Eastern Europe where high systolic blood pressure was the biggest risk factor (Fig. 7A-7B). High body-mass index contributed more to AA-caused death in women than in men in all the European regions, and high salt diet played particular role in Central Europe, especially in men (Fig. 7B).

**Figure 7.**
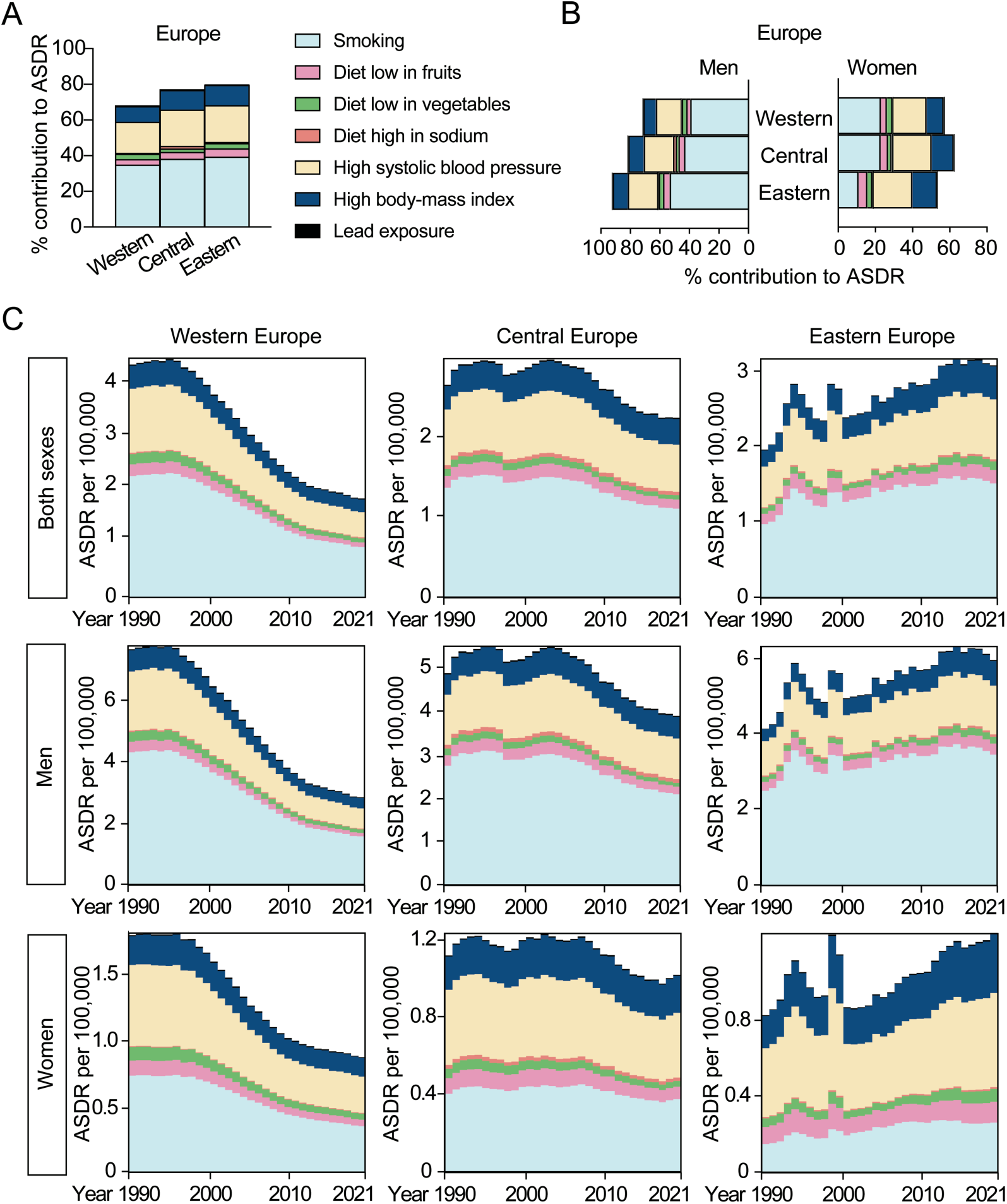
Percentage of AA-caused ASDR attributable to risk factors in Europe. (A) Percentage contribution of risk factors to AA-caused ASDR in Western Europe, Central Europe and Eastern Europe in 2021. (B) Percent contribution of risk factors to AA-related ASDR in Western Europe, Central Europe and Eastern Europe, stratified by sex in 2021. (C) AA-caused ASDR attributable to risk factors in Western Europe, Central Europe and Eastern Europe, stratified by sex from 1990 to 2021.

In order to understand the reasons for the differential changes in ASDR of AA in Western Europe (46.2% decrease) and in Eastern Europe (51.6% increase) from 1990 to 2021, we explored the ASDR attributable to different risk factors. All the known risk factors decreased in Western Europe in both men and women, and increased in Eastern Europe over the time. Smoking contributed the most to the change of AA-caused ASDR in men population, also in women from Western Europe. High systolic blood pressure played the second important role. In women from Eastern Europe, the increases in AA-caused ASDR were particularly attributable to high systolic blood pressure and followed by smoking and high body-mass index (Fig. 7C).

### Risk factors contributing to aortic aneurysm-related deaths in Asia

Smoking was the biggest risk factor for AA-caused death in all Asian regions in men, but high systolic blood pressure was the biggest risk factor for AA-caused death in Asian women. This was observed in both Japan, the country with the continuous increasing AA-related death and the highest ASDR in Asia, and in Singapore, where the AA-caused ASDR was decreasing (Fig. 4A and Fig. 8A-8B).

**Figure 8.**
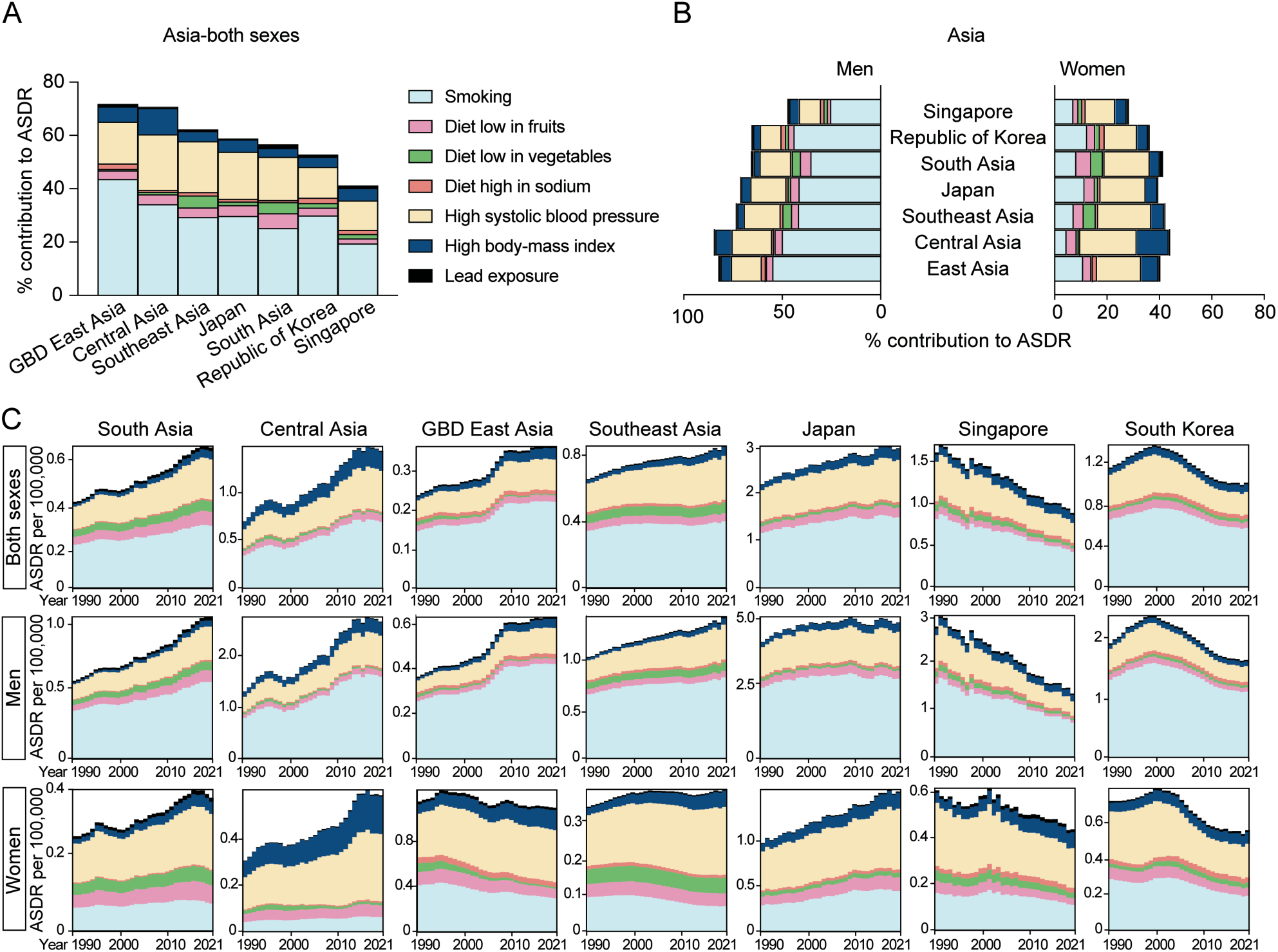
Percentage of AA-caused ASDR attributable to risk factors in Asia. (A) Percentage contribution of risk factors to AA-caused ASDR in South Asia, Central Asia, Southeast Asia, GBD East Asia including China and North Korea, Japan, Republic of Korea and Singapore in 2021. (B) Percent contribution of risk factors to AA-related deaths in the above regions, stratified by sex, in 2021. (C) AA-caused ASDR attributable to risk factors in above regions, stratified by sex, from 1990 to 2021.

The trend in AA-related ASDR attributable to all the risk factors across different regions of Asia from 1990 to 2021 are shown in Fig. 8C. Over the three decades, the contribution of smoking steadily increased among men in most Asian regions, with the exception of Singapore. In women, the contribution of smoking decreased in many regions, although it saw a dramatic increase in Japan. In Japan, AA-related ASDR attributable to smoking rose significantly among women, increasing by 53.6%, from 0.28 (95% UI: 0.22 - 0.36) per 100,000 in 1990 to 0.43 (95% UI: 0.31 – 0.59) in 2021.

In contrast, the increase in men was only 12.6%. The contribution of high blood pressure grew in both men and women across most of Asia, with exceptions in Singapore and South Asia. Additionally, the contribution of high body-mass index increased in both men and women in most regions in Asia, including Singapore, but this trend was particularly prominent in women, especially in Central Asia.

Interestingly, in contrast to the trends observed in most other Asian countries, the ASDR attributable to all the risk factors decreased in Singapore, with a more significant reduction in men than in women. In Japan, which reported the highest AA-related deaths and highest mortality rate per 100,000 globally, the ASDR attributable to all the risk factors increased, with a more pronounced rise observed in women (Fig. 8C).

## Discussion

AA remains a significant global health burden. Despite the trend of decrease in ASDR, AA-related mortality rates have risen over the past three decades worldwide, driven by population growth and increased life expectancy. A comprehensive analysis of these trends is essential for identifying high-risk populations, guiding healthcare policies, and improving prevention strategies. In this study, we explored AA-related mortality data from the Global Burden of Disease Study 2021, assessing epidemiological trends and risk factors for AA-related mortality from 1990 to 2021 across regions, with a focus on age, sex, and socio-economic factors. The findings aim to guide strategies for reducing AA mortality, even amidst a growing and aging population.

### Global Trends in Aortic Aneurysm-related Mortality

From 1990 to 2021, the total AA-related deaths increased by 74.2%, reaching 153,927 deaths in 2021. However, this rise is influenced by population growth and aging demographics. Our findings indicate that AA-related mortality rates increase significantly with age, highlighting the vulnerability of older populations. This is in agreement with previous studies^23^. Consequently, countries with large population and longer life expectancies bear a greater burden of AA-related deaths. Japan, India, and the United States reported the highest absolute numbers of AA-related deaths, while Japan, Monaco, Armenia, Montenegro, and Denmark ranked highest in mortality rates per 100,000 people.

The age-standardized death rate (ASDR), which accounts for population aging, shows a global decline, from 2.54 deaths per 100,000 in 1990 to 1.86 in 2021. This trend suggests that advancements in healthcare and preventive measures can help mitigate AA-related mortality, despite the growing absolute number of cases. After adjusting for age, Armenia, Montenegro, Nauru, Monaco, Japan, Saint Lucia, Brunei Darussalam, Norway, Denmark and Grenada emerged as the top ten countries with highest AA mortality rates.

To better understand and address the high AA mortality in these nations, it is crucial to analyze the underlying risk factors. Additionally, using ASDR allows for more accurate comparisons of mortality rates across different populations and time periods by eliminating the confounding effects of population growth and aging.

### Regional Variations in Aortic Aneurysm-related Mortality

AA-related mortality rates vary significantly across world regions. While Europe and America have experienced a decline in ASDR, Asia has seen an increase. Africa shows high variability among countries, with no clear overall trend.

Interestingly, regions with advanced healthcare systems, high incomes, and high SDI, such as Western Europe, showed a decline in ASDR. However, they still bear a significant burden of AA-related deaths. Our findings suggest that while healthcare advancements have reduced death rates, these regions continue to face a higher prevalence of lifestyle-related risk factors - such as smoking, hypertension, and obesity - that significantly contribute to AA-related deaths. Moreover, many AA cases remain fatal due to their sudden onset and the challenges of timely intervention, particularly in cases of ruptured aneurysms or in clinically frail patients. This underscores the reality that even in well-developed healthcare systems, AA can still remain a serious cause of mortality. Additionally, countries with advanced healthcare systems may have better diagnostic capabilities, leading to better detection and reporting of AA-related deaths.

In contrast, ASDR of AA has increased in some world regions, including Eastern Europe, South Asia, and Central Asia. Our analysis indicates that many risk factors have contributed to the increase in AA-related deaths in both sexes. Among men, smoking is the most significant contributor, whereas hypertension and obesity (high body-mass index) play a larger role in women. Furthermore, limited access to screening and delays in emergency surgical intervention may have led to higher fatality rates in these regions. The rise in AA-related deaths highlights the urgent need for stronger public health measures, including smoking cessation programs, better hypertension management, and improved screening and surgical interventions. Without these measures, AA mortality may continue to increase, particularly as life expectancy rises in many middle-income countries.

Underreporting and misclassification of deaths caused by AA may contribute to the low ASDR of AA in regions with weaker healthcare infrastructure, lower income levels, or lower-middle SDI. Further investigations are needed to address these gaps in data accuracy and reporting.

### Sex Differences in Aortic Aneurysm-related Mortality

Sex significantly influences the burden, progression, and risk factors associated with AA-related mortality ^23^. Our findings show that the ASDR for AA were consistently higher in men than in women. This disparity is likely due to the higher prevalence of AA in men, with subclinical AA being four times more common in men than in women^23^.

However, our study also indicates that the ASDR for AA in men was nearly twice that in women across all major global regions. This may reflect a higher mortality risk of AA for women, as AA in women tends to rupture at smaller sizes and is more likely to experience rapid expansion and rupture, leading to higher mortality rates^24,25^. As a result, women have lower incidence rates but worse outcomes for AA compared to men.

From 1990 to 2021, our results show a global decrease of 33.6% in the ASDR for AA in men, from 3.87 to 2.57 deaths per 100,000. In contrast, the ASDR for women dropped by only 19%, from 1.58 to 1.28 deaths per 100,000. The increased AA mortality in women due to rising rates of smoking, high systolic blood pressure, and high body mass index contributed to the increased ASDR in regions such as South Asia, Central Asia, Japan, and Eastern Europe.

Screening programs for men aged 65 or above have led to a decline in the mortality of abdominal AA in several countries ^26,27^. However, such screening is generally not recommended for women^28^, although AA tends to progress more aggressively in them with more adverse outcomes. Our findings suggest that it is crucial to implement preventive measures and screening for undiagnosed AA in women with risk factors to reduce mortality in this group as well, especially with global trends of equal AA-mortality between sexes at high ages.

### Risk Factors Contributing to Aortic Aneurysm-Related Deaths

Consistent with earlier findings from the GBD 2019 study^17^, our study confirms that smoking continues to be a leading attributable risk factor for AA in men, while hypertension poses a greater risk for women across the majority of the studied countries. Smoking increases aortic inflammation, proteolysis, medial vascular smooth muscle cell apoptosis and angiogenesis within the aortic wall^29–31^. Elevated blood pressure directly increases the mechanical stress on the aortic wall, influencing the smooth muscle cell’s mechano-sensing ability contributing to maladaptive remodeling and aortic dissection ^32,33^. Moreover, both smoking and hypertension are well known to induce endothelial dysfunction, an emerging pathogenesis of TAA^34,35^.

Smoking is the most significant risk factor for AA-related deaths, responsible for nearly 30% of cases globally. Previous studies have reported that smoking increases the risk of AAA by about four times^30^ and nearly doubles the risk of TAA^36^. Effective tobacco control in countries such as Sweden, Norway, and Denmark^37^ may have contributed to a decline in AA-related mortality. In contrast, Japan, which has the highest AA-related mortality in Asia, have weaker tobacco control policies^38^. The impact of smoking is particularly evident in regions with a high burden of aortic aneurysms, such as Europe and America. Countries like Lebanon, Georgia, and Belarus show alarmingly high smoking-related AA mortality rates, suggesting the urgent need for targeted smoking cessation programs.

Patients with hypertension have a 66% higher risk of developing AAA^39^ and more than double the risk of developing TAA^32,36^. We found that high systolic blood pressure accounts for 17.3% of AA-related deaths, with a stronger impact on women, particularly in Eastern Europe and Asia. Countries like Indonesia and Hungary underscore the need for better hypertension management.

As a growing pandemic^40^, obesity can induce chronic inflammation in aorta and contribute to AA development^41^. Positive association between body mass index and AA mortality has been found among Japanese men and smokers^42^. Our research show that obesity is a smaller but significant risk factor for AA mortality particularly in regions with rising obesity rates, such as the Middle East and Eastern Europe, especially among women.

Our findings indicate that poor diet, including low fruit/vegetable intake and high sodium consumption, and lead exposure, also contribute to AA-related mortality. The underlying mechanism may be increased oxidative stress in aorta^43,44^. Their impact is more pronounced in specific countries, such as Zimbabwe and Mongolia for low fruit/vegetable intake, and China and South Korea for high salt consumption. Lead exposure, while minor globally (0.65%), is a significant concern in specific regions, including Southern and Eastern Asia, Africa, and Central America.

### Limitations

The current study has several limitations. Firstly, the GBD 2021 study does not include data on the prevalence and incidence of AA, which may impact the interpretation of the disease burden in this analysis. Additionally, the data retrieved from the GBD 2021 study covers AA without stratifying into TAA or AAA, making it difficult to identify which regions are most affected by specific types of AA. Considering the different effects of risk factors on TAA vs. AAA, attributable effects of modifiable risk factors may be over- or underestimated for each type of AA. Previous reports, however, still highlight smoking and hypertension as significant risk factors of both AAA and TAA^30,32,36^. Underreporting and limited information from some regions and nations may impact the results. The limitation for the estimation of risk-outcome association in GBD study has been summarized recently^19^. GBD data provides only summary statistics without raw individual-level data or sample size, which restricts deeper statistical inferences. We therefore only did Descriptive Statistics.

### Conclusion

Despite global efforts and decreasing trend of AA-related ASDR, the rising total number of AA-related deaths remains a significant burden, with notable regional variations and sex disparities. The decline of AA-related ASDR in Western Europe and the America suggests effective prevention and treatment strategies, while the increase in Eastern Europe, Central and South Asia and Japan, particularly in women highlights emerging public health challenges. Smoking, hypertension, and obesity remain the top contributors to AA mortality, underscoring the need for targeted interventions, particularly in high-risk populations. Addressing these modifiable risk factors through public health campaigns, smoking cessation programs, and improved healthcare access could help reduce the global burden of AA-related deaths in the coming years.

## Data availability statement

The data used in this study were sourced from the Global Burden of Disease (GBD) Study 2021, conducted by the Institute for Health Metrics and Evaluation (IHME). The data were accessed via the GBD Results tool, available at: https://vizhub.healthdata.org/gbd-results/, accessed in March 2025. All raw data used to make the figures are presented in Supplemental Digital Content (SDC).

## Acknowledgements

This research has been conducted as part of the Global Burden of Diseases, Injuries, and Risk Factors Study (GBD), coordinated by the Institute for Health Metrics and Evaluation. The GBD was partially funded by the Bill & Melinda Gates Foundation; the funders had no role in the study design, data analysis, data interpretation, or writing of the report.

## Author Contributions

Conceptualization: X-F.Z. and X-W.Z; Methodology and investigation: R-M.C., L.M. D.F. and X-W.Z.; Validation: R-M.C., L.M., D.F., X-F.Z., H.B., and X-W.Z.; Resources: X-F.Z., H.B., and X-W.Z.; Writing, review & editing: R-M.C., L.M., D.F., X-F.Z., H.B., and X-W.Z.; Visualization: R-M C., L.M. and X-W.Z.; All authors intellectually commented on and edited the manuscript and approved the final version.

