## Supplemental tables for "Global Trends and Risk Factors of Aortic Aneurysm Mortality from 1990 to 2021: An Analysis of the Global Burden of Disease Study 2021"

Raw data are presented in Tables as mean and their 95% uncertainty intervals (UIs) with upper and lower bounds.

Data for Figure 1	4

Fig 1A. Global death cases from 1990 to 2021	4

Fig 1B. World map of death cases in 2021	5

Fig 1C. Global number of deaths per age group in 2021	10

Fig 1D. Global death rates per 100.000 from 1990 to 2021	11

Fig 1E. World map of death rates per 100,000 in 2021	12

Fig 1F. Global death rate per 100,000 per age group in 2021	17

Fig 1G. Global age-standardized death rate (ASDR) from 1990 to 2021	18

Fig 1H and 1I. Global age-standardized death rate (ASDR) in 2021	19

Data for Figure 2	24

Fig 2A. ASDR across Four World Regions from 1990 to 2021	24

Fig 2B. ASDR across Health System Groupings from 1990 to 2021	28

Fig 2C. ASDR across different Commonwealth Income regions from 1990 to 2021	32

Fig 2D. ASDR across SDI regions from 1990 to 2021	35

Fig 2E. ASDR in males and females across the Four World Regions in 2021	39

Fig 2F. ASDR in males and females across Health System Groupings in 2021	39

Fig 2G. ASDR in males and females across different Commonwealth Income regions in 2021	39

Fig 2H. ASDR in males and females across SDI regions in 2021	39

Data for Figure 3	41

Fig 3A. ASDR in Europe from 1990 to 2021	41

Fig 3B. ASDR in Western Europe from 1990 to 2021	44

Fig 3C. ASDR in Central Europe from 1990 to 2021	63

Fig 3D. ASDR in Eastern Europe from 1990 to 2021	73

Fig 3E. ASDR in males and females from Europe in 2021	78

Fig 3F. ASDR in males and females from Western Europe in 2021	78

Fig 3G. ASDR in males and females from Central Europe	79

Fig 3H. ASDR in males and females from Eastern Europe in 2021	80

Data for Figure 4	81

Fig 4A. ASDR in Asia from 1990 to 2021	81

Fig 4B. ASDR in South Asia from 1990 to 2021	87

Fig 4C. ASDR in Central Asia from 1990 to 2021	91

Fig 4D. ASDR in Southeast Asia from 1990 to 2021	98

Fig 4E. ASDR in East Asia from 1990 to 2021	108

Fig 4F. ASDR in males and females from Asia in 2021	110

Fig 4G. ASDR in males and females from South Asia in 2021	110

Fig 4H. ASDR in males and females from Central Asia in 2021	110

Fig 4I. ASDR in males and females from Southeast Asia in 2021	111

Fig 4J. ASDR in males and females from GBD East Asia in 2021	112

Data for Figure 5	113

Fig 5A. Global ASDR in males and females from 1990 to 2021	113

Fig 5B. Global death rate per 100,000 in males and females per age groups in 2021	115

Fig 5C. Percentage changes in AA-caused ASDR of men and women in 2021 compared to 1990	116

Data for Figure 6	118

Fig 6A. Percentage contribution of risk factors to global AA-ASDR grouped by sex in 2021	118

Fig 6B. Percentage contribution of risk factors to AA-ASDR in world regions	119

Fig 6C. Percentage contribution of risk factors to AA-ASDR in world regions grouped by sex in 2021	122

Fig 6D. Percentage contribution of risk factors to AA-ASDR in high-burden countries in 2021	128

Fig 6E. World maps of percentage contribution of risk factors to AA-caused ASDR in 2021	131

Data for Figure 7	165

Fig 7A. Percentage contribution of risk factors to AA- ASDR in Europe in 2021	165

Fig 7B. Percentage contribution of risk factors to AA-ASDR in Europe grouped by sex in 2021	166

Fig 7C. AA-ASDR attributable to risk factors in Europe grouped by sex from 1990 to 2021	167

Data for Figure 8	171

Fig 8A. Percentage contribution of risk factors to AA-ASDR in Asia in 2021	171

Fig 8B. Percentage contribution of risk factors to AA-ASDR in Asia grouped by sex in 2021	173

Fig 8C. AA-ASDR attributable to risk factors in Asia grouped by sex from 1990 to 2021	176

### Data for Figure 1

#### Fig 1A. Global death cases from 1990 to 2021

| **Year** | **Death cases** |
| --- | --- |
| 1990 | 88352.86 (83090.19 to 93491.94) |
| 1991 | 91522.72 (85859.47 to 96845.37) |
| 1992 | 94248.00 (88305.42 to 99617.79) |
| 1993 | 98058.00 (91826.12 to 103627.38) |
| 1994 | 101050.53 (94666.70 to 106801.81) |
| 1995 | 103641.19 (96905.05 to 109549.50) |
| 1996 | 105056.73 (98157.74 to 111170.91) |
| 1997 | 105764.31 (98749.31 to 112013.17) |
| 1998 | 107337.56 (100033.84 to 113578.21) |
| 1999 | 110871.56 (103396.86 to 117443.96) |
| 2000 | 111891.54 (104222.15 to 118437.53) |
| 2001 | 111196.78 (103141.02 to 117903.18) |
| 2002 | 112630.89 (104423.55 to 119485.70) |
| 2003 | 114167.51 (105754.73 to 121059.80) |
| 2004 | 114917.81 (106062.37 to 122119.45) |
| 2005 | 116486.99 (107763.83 to 123827.27) |
| 2006 | 117016.96 (107646.94 to 124342.87) |
| 2007 | 118750.48 (109187.78 to 126633.60) |
| 2008 | 119917.96 (109966.00 to 127769.06) |
| 2009 | 120956.04 (111232.56 to 129205.74) |
| 2010 | 122965.64 (112821.62 to 131022.72) |
| 2011 | 124205.27 (113610.57 to 132604.13) |
| 2012 | 125898.18 (115172.16 to 134509.96) |
| 2013 | 128059.28 (116587.81 to 137271.82) |
| 2014 | 131234.19 (119315.88 to 141021.07) |
| 2015 | 134722.78 (122689.22 to 144516.99) |
| 2016 | 139107.71 (125857.15 to 149208.95) |
| 2017 | 141983.11 (128792.10 to 152396.51) |
| 2018 | 145512.68 (131906.71 to 155795.32) |
| 2019 | 149012.25 (134741.06 to 159764.58) |
| 2020 | 150848.23 (135574.08 to 161878.18) |
| 2021 | 153927.20 (138413.36 to 165738.65) |

#### Fig 1B. World map of death cases in 2021

| **Location** | **Death cases** |
| --- | --- |
| American Samoa | 1.08 (0.90 to 1.31) |
| Antigua and Barbuda | 1.85 (1.69 to 2.09) |
| Arab Republic of Egypt | 250.50 (200.05 to 307.61) |
| Argentine Republic | 1533.23 (1398.22 to 1665.05) |
| Australia | 1186.44 (1032.66 to 1292.39) |
| Barbados | 11.50 (9.33 to 14.14) |
| Belize | 2.30 (2.00 to 2.60) |
| Bermuda | 6.29 (5.36 to 7.54) |
| Bolivarian Republic of Venezuela | 536.31 (419.48 to 670.89) |
| Bosnia and Herzegovina | 189.32 (135.02 to 252.42) |
| Brunei Darussalam | 12.88 (10.87 to 15.31) |
| Burkina Faso | 155.28 (74.97 to 299.16) |
| Canada | 1773.60 (1541.12 to 1928.66) |
| Central African Republic | 37.08 (19.11 to 68.17) |
| Commonwealth of Dominica | 3.08 (2.48 to 3.82) |
| Commonwealth of the Bahamas | 10.31 (8.51 to 12.39) |
| Cook Islands | 0.73 (0.42 to 1.24) |
| Czech Republic | 637.49 (554.46 to 721.95) |
| Democratic People's Republic of Korea | 174.70 (131.40 to 232.47) |
| Democratic Republic of Sao Tome and Principe | 2.16 (1.07 to 3.79) |
| Democratic Republic of the Congo | 626.42 (309.20 to 1114.51) |
| Democratic Republic of Timor-Leste | 5.39 (3.56 to 8.27) |
| Democratic Socialist Republic of Sri Lanka | 87.53 (59.23 to 122.43) |
| Dominican Republic | 157.96 (115.35 to 208.30) |
| Eastern Republic of Uruguay | 255.95 (233.21 to 275.61) |
| Federal Democratic Republic of Ethiopia | 424.03 (218.94 to 719.77) |
| Federal Democratic Republic of Nepal | 235.84 (162.42 to 367.86) |
| Federal Republic of Germany | 4734.70 (4138.78 to 5130.86) |
| Federal Republic of Nigeria | 1691.03 (768.67 to 3087.52) |
| Federal Republic of Somalia | 43.90 (19.89 to 106.22) |
| Federated States of Micronesia | 1.95 (1.40 to 2.61) |
| Federative Republic of Brazil | 10010.25 (9185.86 to 10566.45) |
| French Republic | 3144.53 (2713.06 to 3400.73) |
| Gabonese Republic | 33.10 (20.12 to 47.97) |
| Georgia | 173.00 (146.35 to 200.39) |
| Grand Duchy of Luxembourg | 25.53 (22.49 to 28.28) |
| Greenland | 1.07 (0.87 to 1.35) |
| Grenada | 4.71 (4.11 to 5.29) |
| Guam | 3.65 (3.08 to 4.27) |
| Hashemite Kingdom of Jordan | 68.64 (52.50 to 88.67) |
| Hellenic Republic | 1065.64 (955.05 to 1148.99) |
| Hungary | 574.37 (507.39 to 647.34) |
| Independent State of Papua New Guinea | 65.80 (43.83 to 96.42) |
| Independent State of Samoa | 3.52 (2.66 to 4.55) |
| Ireland | 247.58 (207.38 to 274.90) |
| Islamic Republic of Afghanistan | 29.10 (18.46 to 43.66) |
| Islamic Republic of Iran | 383.79 (340.23 to 432.35) |
| Islamic Republic of Mauritania | 44.45 (16.20 to 78.07) |
| Islamic Republic of Pakistan | 1472.16 (1080.82 to 2066.44) |
| Jamaica | 55.71 (43.49 to 70.23) |
| Japan | 23815.49 (19179.92 to 26463.45) |
| Kingdom of Bahrain | 5.18 (3.91 to 7.01) |
| Kingdom of Belgium | 606.22 (516.67 to 658.71) |
| Kingdom of Bhutan | 8.29 (5.16 to 12.17) |
| Kingdom of Cambodia | 73.62 (46.97 to 118.67) |
| Kingdom of Denmark | 617.04 (547.43 to 672.54) |
| Kingdom of Eswatini | 10.46 (6.84 to 14.86) |
| Kingdom of Lesotho | 17.39 (9.21 to 27.46) |
| Kingdom of Morocco | 139.56 (97.97 to 181.93) |
| Kingdom of Norway | 540.60 (466.25 to 580.59) |
| Kingdom of Saudi Arabia | 40.89 (28.58 to 54.80) |
| Kingdom of Spain | 2255.90 (1978.97 to 2434.46) |
| Kingdom of Sweden | 984.67 (845.59 to 1097.85) |
| Kingdom of Thailand | 2133.88 (1640.10 to 2754.61) |
| Kingdom of the Netherlands | 1465.59 (1262.87 to 1605.02) |
| Kingdom of Tonga | 2.33 (1.66 to 3.05) |
| Kyrgyz Republic | 36.04 (28.70 to 44.02) |
| Lao People's Democratic Republic | 34.13 (24.87 to 46.86) |
| Lebanese Republic | 139.61 (113.94 to 174.64) |
| Malaysia | 971.33 (811.87 to 1158.65) |
| Mongolia | 14.32 (10.89 to 18.32) |
| Montenegro | 80.89 (61.25 to 105.23) |
| New Zealand | 362.23 (317.81 to 392.13) |
| North Macedonia | 79.51 (51.28 to 119.78) |
| Northern Mariana Islands | 1.36 (1.07 to 1.70) |
| Palestine | 17.39 (13.55 to 21.81) |
| People's Democratic Republic of Algeria | 127.69 (95.98 to 169.99) |
| People's Republic of Bangladesh | 1457.82 (952.13 to 2421.76) |
| People's Republic of China | 9033.46 (7052.89 to 11643.53) |
| Plurinational State of Bolivia | 111.14 (83.88 to 149.99) |
| Portuguese Republic | 404.98 (361.46 to 436.78) |
| Principality of Andorra | 6.44 (4.09 to 9.69) |
| Principality of Monaco | 5.85 (4.42 to 7.84) |
| Puerto Rico | 77.01 (63.32 to 90.36) |
| Republic of Albania | 46.83 (32.41 to 65.26) |
| Republic of Angola | 272.60 (162.44 to 408.84) |
| Republic of Armenia | 396.67 (330.04 to 467.31) |
| Republic of Austria | 364.71 (321.35 to 394.95) |
| Republic of Azerbaijan | 130.31 (81.38 to 211.56) |
| Republic of Belarus | 595.76 (485.73 to 714.72) |
| Republic of Benin | 70.18 (27.97 to 125.91) |
| Republic of Botswana | 25.48 (15.55 to 35.03) |
| Republic of Bulgaria | 362.88 (294.50 to 448.66) |
| Republic of Burundi | 57.45 (25.50 to 105.88) |
| Republic of Cabo Verde | 10.21 (4.96 to 17.83) |
| Republic of Cameroon | 246.56 (146.45 to 429.80) |
| Republic of Chad | 72.50 (32.53 to 142.95) |
| Republic of Chile | 563.14 (514.26 to 603.97) |
| Republic of Colombia | 1635.74 (1357.03 to 1937.00) |
| Republic of Costa Rica | 131.80 (114.59 to 148.00) |
| Republic of Côte d'Ivoire | 225.61 (100.21 to 392.95) |
| Republic of Croatia | 316.78 (271.04 to 364.68) |
| Republic of Cuba | 770.49 (655.31 to 868.02) |
| Republic of Cyprus | 80.41 (60.20 to 101.42) |
| Republic of Djibouti | 9.74 (4.15 to 15.81) |
| Republic of Ecuador | 207.72 (164.69 to 262.06) |
| Republic of El Salvador | 33.78 (26.66 to 43.15) |
| Republic of Equatorial Guinea | 14.57 (6.84 to 25.64) |
| Republic of Estonia | 97.61 (85.07 to 110.32) |
| Republic of Fiji | 21.40 (16.02 to 27.44) |
| Republic of Finland | 519.49 (449.99 to 566.43) |
| Republic of Ghana | 403.84 (184.09 to 682.71) |
| Republic of Guatemala | 44.55 (38.06 to 52.15) |
| Republic of Guinea | 93.21 (34.23 to 180.58) |
| Republic of Guinea-Bissau | 13.36 (7.22 to 23.88) |
| Republic of Guyana | 19.82 (15.30 to 25.13) |
| Republic of Haiti | 131.10 (81.08 to 204.92) |
| Republic of Honduras | 66.40 (48.19 to 89.77) |
| Republic of Iceland | 16.74 (14.12 to 18.73) |
| Republic of India | 12805.03 (9106.77 to 18763.13) |
| Republic of Indonesia | 1819.52 (1246.16 to 2445.73) |
| Republic of Iraq | 88.75 (64.87 to 117.52) |
| Republic of Italy | 3654.92 (3186.88 to 3938.73) |
| Republic of Kazakhstan | 376.69 (302.21 to 461.90) |
| Republic of Kenya | 336.58 (198.11 to 468.32) |
| Republic of Kiribati | 0.28 (0.20 to 0.36) |
| Republic of Korea | 1767.00 (1440.81 to 2092.51) |
| Republic of Latvia | 124.94 (107.23 to 141.58) |
| Republic of Liberia | 33.77 (14.92 to 66.01) |
| Republic of Lithuania | 186.51 (162.26 to 210.89) |
| Republic of Madagascar | 243.96 (118.60 to 402.66) |
| Republic of Malawi | 122.48 (59.16 to 208.90) |
| Republic of Maldives | 1.79 (0.81 to 3.03) |
| Republic of Mali | 87.73 (38.44 to 168.99) |
| Republic of Malta | 14.03 (12.32 to 15.72) |
| Republic of Mauritius | 14.88 (13.60 to 15.78) |
| Republic of Moldova | 88.90 (78.62 to 99.68) |
| Republic of Mozambique | 232.76 (94.07 to 453.84) |
| Republic of Namibia | 27.48 (19.85 to 37.90) |
| Republic of Nauru | 0.24 (0.17 to 0.32) |
| Republic of Nicaragua | 15.82 (12.73 to 19.42) |
| Republic of Niue | 0.07 (0.05 to 0.08) |
| Republic of Palau | 0.49 (0.38 to 0.65) |
| Republic of Panama | 78.60 (60.13 to 95.56) |
| Republic of Paraguay | 162.90 (125.81 to 206.75) |
| Republic of Peru | 219.23 (163.28 to 289.57) |
| Republic of Poland | 2575.97 (2306.76 to 2830.65) |
| Republic of Rwanda | 95.95 (56.15 to 156.76) |
| Republic of San Marino | 1.46 (0.95 to 2.27) |
| Republic of Senegal | 142.59 (60.36 to 263.44) |
| Republic of Serbia | 691.68 (548.90 to 871.74) |
| Republic of Seychelles | 1.08 (0.74 to 1.52) |
| Republic of Sierra Leone | 62.72 (27.61 to 121.49) |
| Republic of Singapore | 177.36 (157.51 to 191.33) |
| Republic of Slovenia | 105.28 (88.59 to 126.29) |
| Republic of South Africa | 935.35 (819.86 to 1052.21) |
| Republic of South Sudan | 44.88 (19.54 to 89.43) |
| Republic of Sudan | 66.77 (41.21 to 97.68) |
| Republic of Suriname | 12.11 (8.48 to 16.72) |
| Republic of Tajikistan | 12.75 (9.10 to 17.32) |
| Republic of the Congo | 73.53 (43.54 to 110.81) |
| Republic of the Gambia | 21.22 (9.90 to 36.82) |
| Republic of the Marshall Islands | 0.77 (0.50 to 1.11) |
| Republic of the Niger | 75.55 (28.59 to 175.18) |
| Republic of the Philippines | 950.08 (779.01 to 1119.23) |
| Republic of the Union of Myanmar | 392.21 (296.46 to 513.63) |
| Republic of Trinidad and Tobago | 74.21 (57.40 to 93.69) |
| Republic of Tunisia | 55.57 (35.84 to 80.82) |
| Republic of Turkey | 2075.39 (1640.49 to 2605.72) |
| Republic of Uganda | 204.85 (99.15 to 337.63) |
| Republic of Uzbekistan | 231.37 (186.39 to 284.82) |
| Republic of Vanuatu | 3.08 (2.28 to 4.16) |
| Republic of Yemen | 45.53 (26.24 to 71.28) |
| Republic of Zambia | 204.57 (87.46 to 371.74) |
| Republic of Zimbabwe | 220.59 (164.47 to 297.48) |
| Romania | 705.79 (613.50 to 810.36) |
| Russian Federation | 10444.90 (9555.03 to 11307.25) |
| Saint Kitts and Nevis | 1.37 (1.12 to 1.67) |
| Saint Lucia | 11.40 (9.59 to 13.30) |
| Saint Vincent and the Grenadines | 2.78 (2.46 to 3.15) |
| Slovak Republic | 217.54 (170.03 to 275.07) |
| Socialist Republic of Viet Nam | 894.89 (656.44 to 1192.65) |
| Solomon Islands | 4.97 (3.37 to 7.18) |
| State of Eritrea | 42.47 (19.38 to 81.65) |
| State of Israel | 171.17 (150.01 to 186.67) |
| State of Kuwait | 24.22 (20.03 to 29.77) |
| State of Libya | 18.10 (10.87 to 28.95) |
| State of Qatar | 8.11 (4.86 to 12.96) |
| Sultanate of Oman | 11.30 (6.51 to 19.20) |
| Swiss Confederation | 487.50 (411.15 to 539.94) |
| Syrian Arab Republic | 50.96 (36.72 to 68.44) |
| Taiwan (Province of China) | 990.85 (888.32 to 1071.28) |
| Togolese Republic | 70.95 (29.93 to 126.48) |
| Tokelau | 0.05 (0.03 to 0.07) |
| Turkmenistan | 71.57 (53.10 to 102.83) |
| Tuvalu | 0.26 (0.21 to 0.33) |
| Ukraine | 1867.34 (1384.72 to 2442.11) |
| Union of the Comoros | 8.42 (3.15 to 15.34) |
| United Arab Emirates | 43.16 (34.04 to 54.00) |
| United Kingdom of Great Britain and Northern Ireland | 6071.17 (5374.04 to 6434.62) |
| United Mexican States | 749.75 (662.46 to 845.06) |
| United Republic of Tanzania | 561.82 (271.21 to 957.26) |
| United States of America | 12194.98 (10939.91 to 12943.79) |
| United States Virgin Islands | 3.62 (2.75 to 4.58) |

#### Fig 1C. Global number of deaths per age group in 2021

| **Age** | **Death cases** |
| --- | --- |
| 15-19 years | 204.04 (172.69 to 238.52) |
| 20-24 years | 372.37 (324.39 to 427.38) |
| 25-29 years | 589.06 (520.56 to 666.70) |
| 30-34 years | 1015.38 (911.52 to 1150.28) |
| 35-39 years | 1663.93 (1484.60 to 1893.23) |
| 40-44 years | 2499.70 (2253.07 to 2793.16) |
| 45-49 years | 3727.72 (3421.01 to 4117.56) |
| 50-54 years | 5404.79 (4983.78 to 5935.50) |
| 55-59 years | 8290.38 (7597.86 to 9113.56) |
| 60-64 years | 11669.14 (10885.96 to 12673.19) |
| 65-69 years | 16484.09 (15355.40 to 17909.55) |
| 70-74 years | 21017.46 (19538.19 to 22682.30) |
| 75-79 years | 21508.22 (19620.13 to 23035.32) |
| 80-84 years | 22556.80 (19647.35 to 24401.09) |
| 85-89 years | 19701.47 (16222.10 to 21637.05) |
| 90-94 years | 12188.40 (9386.06 to 13647.46) |
| 95+ years | 5034.26 (3520.79 to 5857.42) |

#### Fig 1D. Global death rates per 100.000 from 1990 to 2021

| **Year** | **Death rate per 100,000** |
| --- | --- |
| 1990 | 1.66 (1.56 to 1.75) |
| 1991 | 1.69 (1.59 to 1.79) |
| 1992 | 1.71 (1.61 to 1.81) |
| 1993 | 1.76 (1.65 to 1.86) |
| 1994 | 1.79 (1.68 to 1.89) |
| 1995 | 1.81 (1.69 to 1.91) |
| 1996 | 1.81 (1.69 to 1.92) |
| 1997 | 1.80 (1.68 to 1.91) |
| 1998 | 1.80 (1.68 to 1.91) |
| 1999 | 1.84 (1.72 to 1.95) |
| 2000 | 1.83 (1.71 to 1.94) |
| 2001 | 1.80 (1.67 to 1.91) |
| 2002 | 1.80 (1.67 to 1.91) |
| 2003 | 1.80 (1.67 to 1.91) |
| 2004 | 1.79 (1.65 to 1.90) |
| 2005 | 1.79 (1.66 to 1.90) |
| 2006 | 1.78 (1.63 to 1.89) |
| 2007 | 1.78 (1.64 to 1.90) |
| 2008 | 1.77 (1.62 to 1.89) |
| 2009 | 1.76 (1.62 to 1.88) |
| 2010 | 1.77 (1.62 to 1.89) |
| 2011 | 1.76 (1.61 to 1.88) |
| 2012 | 1.77 (1.62 to 1.89) |
| 2013 | 1.77 (1.62 to 1.90) |
| 2014 | 1.80 (1.63 to 1.93) |
| 2015 | 1.82 (1.66 to 1.95) |
| 2016 | 1.86 (1.68 to 1.99) |
| 2017 | 1.87 (1.70 to 2.01) |
| 2018 | 1.90 (1.72 to 2.03) |
| 2019 | 1.92 (1.74 to 2.06) |
| 2020 | 1.93 (1.73 to 2.07) |
| 2021 | 1.95 (1.75 to 2.10) |

#### Fig 1E. World map of death rates per 100,000 in 2021

| **Location** | **Death rate per 100,000** |
| --- | --- |
| China | 0.63 (0.50 to 0.82) |
| Tajikistan | 0.13 (0.09 to 0.17) |
| Cabo Verde | 1.83 (0.89 to 3.19) |
| Monaco | 15.46 (11.68 to 20.70) |
| Fiji | 2.32 (1.73 to 2.97) |
| Kiribati | 0.23 (0.16 to 0.30) |
| New Zealand | 7.01 (6.15 to 7.59) |
| Democratic People's Republic of Korea | 0.66 (0.50 to 0.88) |
| Syrian Arab Republic | 0.36 (0.26 to 0.49) |
| Seychelles | 1.03 (0.70 to 1.44) |
| Malta | 3.17 (2.78 to 3.55) |
| Bolivia (Plurinational State of) | 0.94 (0.71 to 1.27) |
| Slovenia | 5.09 (4.28 to 6.10) |
| Antigua and Barbuda | 2.07 (1.89 to 2.34) |
| Congo | 1.36 (0.81 to 2.06) |
| Chad | 0.41 (0.18 to 0.81) |
| Turkmenistan | 1.39 (1.03 to 1.99) |
| Nauru | 2.21 (1.58 to 2.94) |
| Democratic Republic of the Congo | 0.70 (0.34 to 1.24) |
| Algeria | 0.29 (0.22 to 0.38) |
| Bahamas | 2.66 (2.19 to 3.19) |
| Marshall Islands | 1.37 (0.90 to 1.97) |
| Uzbekistan | 0.68 (0.54 to 0.83) |
| Netherlands | 8.52 (7.34 to 9.33) |
| Belarus | 6.39 (5.21 to 7.66) |
| United Republic of Tanzania | 0.96 (0.46 to 1.64) |
| Norway | 9.98 (8.61 to 10.72) |
| Equatorial Guinea | 0.96 (0.45 to 1.70) |
| Tunisia | 0.47 (0.30 to 0.68) |
| Gambia | 0.89 (0.41 to 1.54) |
| Taiwan | 4.19 (3.76 to 4.53) |
| Peru | 0.60 (0.45 to 0.80) |
| Somalia | 0.20 (0.09 to 0.49) |
| Côte d'Ivoire | 0.81 (0.36 to 1.41) |
| Micronesia (Federated States of) | 1.90 (1.37 to 2.54) |
| Niue | 3.95 (3.10 to 4.81) |
| Estonia | 7.45 (6.49 to 8.42) |
| Uganda | 0.47 (0.23 to 0.78) |
| United Arab Emirates | 0.45 (0.35 to 0.56) |
| Portugal | 3.82 (3.41 to 4.12) |
| Albania | 1.76 (1.21 to 2.45) |
| Ecuador | 1.15 (0.91 to 1.45) |
| Türkiye | 2.48 (1.96 to 3.12) |
| Papua New Guinea | 0.63 (0.42 to 0.92) |
| Cambodia | 0.43 (0.28 to 0.70) |
| Andorra | 7.53 (4.78 to 11.32) |
| Belgium | 5.29 (4.50 to 5.74) |
| Yemen | 0.14 (0.08 to 0.21) |
| Guinea | 0.69 (0.25 to 1.34) |
| Northern Mariana Islands | 2.80 (2.20 to 3.50) |
| Spain | 4.95 (4.34 to 5.34) |
| Gabon | 1.82 (1.11 to 2.64) |
| Bahrain | 0.34 (0.26 to 0.46) |
| Samoa | 1.65 (1.25 to 2.13) |
| Barbados | 3.85 (3.12 to 4.73) |
| Ghana | 1.18 (0.54 to 1.99) |
| Colombia | 3.33 (2.77 to 3.95) |
| Indonesia | 0.65 (0.45 to 0.88) |
| Burundi | 0.43 (0.19 to 0.80) |
| Sweden | 9.49 (8.15 to 10.58) |
| Cyprus | 5.92 (4.43 to 7.47) |
| Solomon Islands | 0.73 (0.49 to 1.05) |
| Bosnia and Herzegovina | 5.73 (4.09 to 7.64) |
| Dominica | 4.59 (3.70 to 5.69) |
| Palau | 2.72 (2.09 to 3.60) |
| Belize | 0.54 (0.47 to 0.61) |
| Switzerland | 5.46 (4.61 to 6.05) |
| Austria | 4.06 (3.58 to 4.40) |
| Zambia | 1.05 (0.45 to 1.90) |
| Guinea-Bissau | 0.65 (0.35 to 1.16) |
| Puerto Rico | 2.34 (1.92 to 2.74) |
| Saint Kitts and Nevis | 2.33 (1.91 to 2.85) |
| Bulgaria | 5.35 (4.34 to 6.61) |
| Cuba | 6.84 (5.81 to 7.70) |
| Egypt | 0.24 (0.19 to 0.29) |
| Comoros | 1.13 (0.42 to 2.06) |
| Latvia | 6.68 (5.73 to 7.57) |
| Lao People's Democratic Republic | 0.46 (0.34 to 0.64) |
| Croatia | 7.53 (6.44 to 8.67) |
| Botswana | 1.06 (0.65 to 1.46) |
| Jordan | 0.56 (0.43 to 0.72) |
| Republic of Moldova | 2.47 (2.19 to 2.77) |
| Ukraine | 4.33 (3.21 to 5.67) |
| Tonga | 2.19 (1.56 to 2.87) |
| Iran (Islamic Republic of) | 0.45 (0.40 to 0.51) |
| Lithuania | 6.84 (5.95 to 7.73) |
| Dominican Republic | 1.43 (1.05 to 1.89) |
| Russian Federation | 7.21 (6.60 to 7.81) |
| Liberia | 0.62 (0.27 to 1.21) |
| Lesotho | 0.93 (0.49 to 1.47) |
| United Kingdom | 8.95 (7.92 to 9.48) |
| Iraq | 0.22 (0.16 to 0.29) |
| Costa Rica | 2.78 (2.41 to 3.12) |
| Sudan | 0.15 (0.09 to 0.22) |
| Czechia | 6.00 (5.21 to 6.79) |
| Bangladesh | 0.89 (0.58 to 1.47) |
| Djibouti | 0.77 (0.33 to 1.26) |
| Malaysia | 3.05 (2.55 to 3.64) |
| Honduras | 0.66 (0.48 to 0.89) |
| Vanuatu | 0.99 (0.73 to 1.33) |
| El Salvador | 0.52 (0.41 to 0.67) |
| Maldives | 0.35 (0.16 to 0.59) |
| Eritrea | 0.64 (0.29 to 1.24) |
| Tokelau | 3.71 (2.40 to 5.21) |
| Kuwait | 0.52 (0.43 to 0.64) |
| San Marino | 4.47 (2.91 to 6.94) |
| Bhutan | 1.09 (0.68 to 1.61) |
| Haiti | 1.02 (0.63 to 1.59) |
| Mauritania | 1.01 (0.37 to 1.78) |
| Chile | 3.00 (2.74 to 3.21) |
| South Africa | 1.65 (1.44 to 1.85) |
| Argentina | 3.37 (3.07 to 3.66) |
| Armenia | 13.24 (11.02 to 15.60) |
| Greece | 10.47 (9.39 to 11.29) |
| Kenya | 0.67 (0.40 to 0.94) |
| Hungary | 5.98 (5.29 to 6.74) |
| Afghanistan | 0.09 (0.06 to 0.14) |
| Mexico | 0.58 (0.51 to 0.65) |
| Guatemala | 0.28 (0.24 to 0.33) |
| Mali | 0.36 (0.16 to 0.70) |
| Finland | 9.38 (8.13 to 10.23) |
| Jamaica | 1.99 (1.55 to 2.51) |
| Niger | 0.30 (0.11 to 0.70) |
| Lebanon | 2.52 (2.06 to 3.15) |
| France | 4.74 (4.09 to 5.12) |
| Myanmar | 0.70 (0.53 to 0.91) |
| United States Virgin Islands | 4.21 (3.20 to 5.33) |
| Guyana | 2.59 (2.00 to 3.29) |
| Ethiopia | 0.39 (0.20 to 0.66) |
| Brunei Darussalam | 2.86 (2.41 to 3.39) |
| Nigeria | 0.73 (0.33 to 1.34) |
| Bermuda | 9.90 (8.44 to 11.87) |
| Azerbaijan | 1.24 (0.77 to 2.01) |
| Germany | 5.55 (4.85 to 6.01) |
| Morocco | 0.38 (0.26 to 0.49) |
| Japan | 18.65 (15.02 to 20.72) |
| Namibia | 1.13 (0.82 to 1.56) |
| Greenland | 1.91 (1.54 to 2.41) |
| Philippines | 0.84 (0.69 to 0.99) |
| Eswatini | 0.91 (0.59 to 1.29) |
| Denmark | 10.54 (9.35 to 11.49) |
| American Samoa | 2.18 (1.81 to 2.64) |
| Uruguay | 7.52 (6.85 to 8.09) |
| Republic of Korea | 3.43 (2.79 to 4.06) |
| India | 0.91 (0.64 to 1.33) |
| Grenada | 4.59 (4.01 to 5.16) |
| Cook Islands | 4.12 (2.37 to 6.99) |
| Tuvalu | 2.10 (1.67 to 2.64) |
| Saint Lucia | 6.42 (5.40 to 7.49) |
| Palestine | 0.34 (0.26 to 0.42) |
| Nicaragua | 0.24 (0.19 to 0.29) |
| Madagascar | 0.85 (0.42 to 1.41) |
| Panama | 1.83 (1.40 to 2.23) |
| Thailand | 3.20 (2.46 to 4.13) |
| Montenegro | 13.09 (9.91 to 17.03) |
| Kyrgyzstan | 0.53 (0.42 to 0.64) |
| Timor-Leste | 0.39 (0.25 to 0.59) |
| Sri Lanka | 0.39 (0.27 to 0.55) |
| Pakistan | 0.62 (0.46 to 0.88) |
| Romania | 3.73 (3.24 to 4.28) |
| Venezuela (Bolivarian Republic of) | 2.01 (1.58 to 2.52) |
| Kazakhstan | 1.99 (1.59 to 2.44) |
| Mauritius | 1.17 (1.07 to 1.24) |
| Libya | 0.26 (0.16 to 0.42) |
| Canada | 4.73 (4.11 to 5.15) |
| Malawi | 0.63 (0.30 to 1.07) |
| Oman | 0.24 (0.14 to 0.41) |
| Senegal | 0.90 (0.38 to 1.66) |
| Guam | 2.29 (1.93 to 2.68) |
| Italy | 6.11 (5.33 to 6.59) |
| Trinidad and Tobago | 5.33 (4.12 to 6.73) |
| North Macedonia | 3.65 (2.36 to 5.50) |
| Mozambique | 0.75 (0.30 to 1.46) |
| Togo | 0.85 (0.36 to 1.51) |
| Mongolia | 0.43 (0.33 to 0.55) |
| Sao Tome and Principe | 1.00 (0.50 to 1.75) |
| Australia | 4.60 (4.00 to 5.01) |
| Suriname | 2.09 (1.46 to 2.89) |
| Viet Nam | 0.89 (0.65 to 1.19) |
| Israel | 1.78 (1.56 to 1.95) |
| Ireland | 5.01 (4.20 to 5.56) |
| Poland | 6.74 (6.03 to 7.40) |
| Zimbabwe | 1.41 (1.05 to 1.91) |
| Sierra Leone | 0.71 (0.31 to 1.37) |
| Nepal | 0.76 (0.52 to 1.18) |
| United States of America | 3.67 (3.29 to 3.89) |
| Qatar | 0.27 (0.16 to 0.44) |
| Paraguay | 2.27 (1.75 to 2.88) |
| Rwanda | 0.72 (0.42 to 1.18) |
| Luxembourg | 3.96 (3.49 to 4.39) |
| Saint Vincent and the Grenadines | 2.44 (2.16 to 2.76) |
| Georgia | 4.80 (4.06 to 5.56) |
| Saudi Arabia | 0.11 (0.08 to 0.15) |
| Benin | 0.52 (0.21 to 0.93) |
| Burkina Faso | 0.68 (0.33 to 1.31) |
| South Sudan | 0.46 (0.20 to 0.92) |
| Singapore | 3.10 (2.75 to 3.34) |
| Brazil | 4.54 (4.17 to 4.80) |
| Cameroon | 0.78 (0.46 to 1.35) |
| Iceland | 4.78 (4.03 to 5.35) |
| Serbia | 7.75 (6.15 to 9.77) |
| Angola | 0.83 (0.50 to 1.25) |
| Slovakia | 4.01 (3.13 to 5.07) |
| Central African Republic | 0.68 (0.35 to 1.24) |

#### Fig 1F. Global death rate per 100,000 per age group in 2021

| **Age** | **Death rate per 100,000** |
| --- | --- |
| 15-19 years | 0.03 (0.03 to 0.04) |
| 20-24 years | 0.06 (0.05 to 0.07) |
| 25-29 years | 0.10 (0.09 to 0.11) |
| 30-34 years | 0.17 (0.15 to 0.19) |
| 35-39 years | 0.30 (0.26 to 0.34) |
| 40-44 years | 0.50 (0.45 to 0.56) |
| 45-49 years | 0.79 (0.72 to 0.87) |
| 50-54 years | 1.21 (1.12 to 1.33) |
| 55-59 years | 2.09 (1.92 to 2.30) |
| 60-64 years | 3.65 (3.40 to 3.96) |
| 65-69 years | 5.98 (5.57 to 6.49) |
| 70-74 years | 10.21 (9.49 to 11.02) |
| 75-79 years | 16.31 (14.88 to 17.47) |
| 80-84 years | 25.75 (22.43 to 27.86) |
| 85-89 years | 43.09 (35.48 to 47.32) |
| 90-94 years | 68.13 (52.47 to 76.29) |
| 95+ years | 92.37 (64.60 to 107.47) |

#### Fig 1G. Global age-standardized death rate (ASDR) from 1990 to 2021

| **Year** | **ASDR per 100,000** |
| --- | --- |
| 1990 | 2.54 (2.35 to 2.69) |
| 1992 | 2.57 (2.37 to 2.72) |
| 1991 | 2.56 (2.37 to 2.72) |
| 1994 | 2.61 (2.42 to 2.77) |
| 1997 | 2.55 (2.35 to 2.70) |
| 1995 | 2.62 (2.42 to 2.77) |
| 1998 | 2.53 (2.33 to 2.68) |
| 2000 | 2.50 (2.31 to 2.65) |
| 2003 | 2.36 (2.16 to 2.51) |
| 1999 | 2.54 (2.35 to 2.70) |
| 2001 | 2.42 (2.22 to 2.57) |
| 2004 | 2.31 (2.11 to 2.46) |
| 2006 | 2.22 (2.02 to 2.36) |
| 2008 | 2.14 (1.94 to 2.28) |
| 2005 | 2.28 (2.08 to 2.43) |
| 2010 | 2.07 (1.88 to 2.20) |
| 2007 | 2.19 (1.99 to 2.33) |
| 2009 | 2.10 (1.91 to 2.24) |
| 2011 | 2.02 (1.84 to 2.16) |
| 2012 | 1.99 (1.80 to 2.13) |
| 2013 | 1.96 (1.78 to 2.11) |
| 2015 | 1.94 (1.76 to 2.09) |
| 2014 | 1.95 (1.76 to 2.10) |
| 2016 | 1.95 (1.75 to 2.08) |
| 2017 | 1.92 (1.74 to 2.07) |
| 2018 | 1.91 (1.72 to 2.05) |
| 2019 | 1.90 (1.71 to 2.04) |
| 1996 | 2.59 (2.40 to 2.74) |
| 2021 | 1.86 (1.67 to 2.00) |
| 2002 | 2.39 (2.19 to 2.54) |
| 1993 | 2.60 (2.40 to 2.75) |
| 2020 | 1.87 (1.67 to 2.00) |

#### Fig 1H and 1I. Global age-standardized death rate (ASDR) in 2021

| **Location** | **ASDR per 100,000** |
| --- | --- |
| Republic of Armenia | 9.16 (7.61 to 10.81) |
| Montenegro | 8.65 (6.59 to 11.28) |
| Republic of Nauru | 6.01 (3.50 to 9.06) |
| Principality of Monaco | 5.15 (3.88 to 6.95) |
| Japan | 5.07 (4.33 to 5.47) |
| Saint Lucia | 4.95 (4.17 to 5.77) |
| Brunei Darussalam | 4.79 (4.00 to 5.67) |
| Kingdom of Norway | 4.75 (4.15 to 5.08) |
| Kingdom of Denmark | 4.61 (4.10 to 5.02) |
| Grenada | 4.51 (3.94 to 5.04) |
| Russian Federation | 4.38 (4.01 to 4.74) |
| Eastern Republic of Uruguay | 4.35 (3.98 to 4.67) |
| Bermuda | 4.15 (3.55 to 4.99) |
| Federative Republic of Brazil | 4.06 (3.72 to 4.29) |
| Republic of Serbia | 4.05 (3.21 to 5.10) |
| Malaysia | 4.04 (3.38 to 4.88) |
| United Kingdom of Great Britain and Northern Ireland | 4.03 (3.61 to 4.25) |
| Hellenic Republic | 3.99 (3.65 to 4.26) |
| New Zealand | 3.99 (3.52 to 4.30) |
| Republic of Cyprus | 3.96 (3.04 to 4.88) |
| Principality of Andorra | 3.96 (2.50 to 5.97) |
| Republic of Zimbabwe | 3.96 (3.06 to 5.20) |
| Commonwealth of Dominica | 3.94 (3.17 to 4.87) |
| Republic of Trinidad and Tobago | 3.94 (3.05 to 4.96) |
| Kingdom of Sweden | 3.88 (3.38 to 4.33) |
| Gabonese Republic | 3.79 (2.35 to 5.45) |
| Republic of Belarus | 3.75 (3.05 to 4.49) |
| Republic of Cuba | 3.74 (3.18 to 4.22) |
| Kingdom of the Netherlands | 3.74 (3.24 to 4.08) |
| Republic of Fiji | 3.65 (2.86 to 4.58) |
| Republic of Finland | 3.63 (3.20 to 3.93) |
| Federated States of Micronesia | 3.51 (2.73 to 4.51) |
| Republic of Guyana | 3.45 (2.70 to 4.33) |
| Republic of Poland | 3.44 (3.09 to 3.79) |
| Tokelau | 3.41 (2.24 to 4.78) |
| Republic of Equatorial Guinea | 3.39 (1.69 to 5.75) |
| Republic of the Congo | 3.38 (2.01 to 5.05) |
| Northern Mariana Islands | 3.38 (2.76 to 4.11) |
| Republic of Zambia | 3.36 (1.53 to 5.94) |
| Republic of Croatia | 3.34 (2.87 to 3.86) |
| Republic of Estonia | 3.34 (2.90 to 3.78) |
| Republic of Niue | 3.27 (2.58 to 3.98) |
| Republic of Lithuania | 3.15 (2.73 to 3.56) |
| Republic of Latvia | 3.05 (2.63 to 3.47) |
| Kingdom of Tonga | 3.04 (2.18 to 3.99) |
| Tuvalu | 3.02 (2.44 to 3.75) |
| Bosnia and Herzegovina | 3.00 (2.15 to 3.99) |
| Republic of Colombia | 2.97 (2.47 to 3.53) |
| Cook Islands | 2.94 (1.70 to 4.97) |
| Republic of Palau | 2.93 (2.24 to 3.79) |
| Ireland | 2.93 (2.47 to 3.25) |
| Republic of the Marshall Islands | 2.92 (2.03 to 3.99) |
| Republic of Paraguay | 2.90 (2.23 to 3.67) |
| Republic of Angola | 2.89 (1.72 to 4.26) |
| Georgia | 2.88 (2.43 to 3.35) |
| Hungary | 2.87 (2.54 to 3.23) |
| Republic of Ghana | 2.87 (1.33 to 4.74) |
| Commonwealth of the Bahamas | 2.85 (2.37 to 3.40) |
| Czech Republic | 2.83 (2.46 to 3.19) |
| Independent State of Samoa | 2.80 (2.12 to 3.57) |
| American Samoa | 2.72 (2.24 to 3.30) |
| Argentine Republic | 2.67 (2.44 to 2.90) |
| North Macedonia | 2.63 (1.80 to 3.81) |
| Republic of Bulgaria | 2.62 (2.12 to 3.23) |
| Republic of Iceland | 2.61 (2.23 to 2.92) |
| Republic of Madagascar | 2.53 (1.19 to 4.29) |
| United Republic of Tanzania | 2.50 (1.21 to 4.29) |
| Republic of Namibia | 2.48 (1.85 to 3.39) |
| Ukraine | 2.47 (1.82 to 3.25) |
| Republic of Cabo Verde | 2.46 (1.19 to 4.30) |
| Republic of the Gambia | 2.44 (1.12 to 4.26) |
| Republic of Côte d'Ivoire | 2.44 (1.10 to 4.31) |
| Saint Kitts and Nevis | 2.44 (2.04 to 2.90) |
| Republic of Costa Rica | 2.41 (2.10 to 2.70) |
| Republic of Mozambique | 2.40 (1.03 to 4.55) |
| Taiwan (Province of China) | 2.37 (2.14 to 2.56) |
| Islamic Republic of Mauritania | 2.37 (0.85 to 4.15) |
| Australia | 2.35 (2.06 to 2.55) |
| Republic of Cameroon | 2.33 (1.42 to 3.97) |
| Federal Republic of Nigeria | 2.30 (1.09 to 4.12) |
| Democratic Republic of Sao Tome and Principe | 2.29 (1.14 to 4.00) |
| Republic of Haiti | 2.29 (1.42 to 3.57) |
| Togolese Republic | 2.29 (0.96 to 4.18) |
| Slovak Republic | 2.29 (1.79 to 2.90) |
| Republic of South Africa | 2.29 (2.02 to 2.54) |
| Swiss Confederation | 2.28 (1.96 to 2.51) |
| Republic of Turkey | 2.27 (1.80 to 2.83) |
| Canada | 2.26 (1.99 to 2.45) |
| Kingdom of Belgium | 2.25 (1.96 to 2.43) |
| Federal Republic of Germany | 2.25 (2.02 to 2.41) |
| Republic of Guinea-Bissau | 2.24 (1.18 to 3.99) |
| Republic of Vanuatu | 2.23 (1.65 to 2.95) |
| Republic of Kazakhstan | 2.23 (1.80 to 2.71) |
| Grand Duchy of Luxembourg | 2.23 (1.97 to 2.46) |
| Republic of Botswana | 2.22 (1.36 to 3.04) |
| Kingdom of Eswatini | 2.21 (1.47 to 3.03) |
| Lebanese Republic | 2.21 (1.80 to 2.76) |
| Barbados | 2.18 (1.77 to 2.68) |
| Republic of Italy | 2.18 (1.95 to 2.34) |
| Republic of Chile | 2.17 (1.98 to 2.32) |
| Republic of Slovenia | 2.16 (1.82 to 2.58) |
| Republic of Singapore | 2.15 (1.91 to 2.32) |
| Central African Republic | 2.14 (1.08 to 3.73) |
| Democratic Republic of the Congo | 2.14 (1.08 to 3.82) |
| Republic of Senegal | 2.12 (0.90 to 3.88) |
| Saint Vincent and the Grenadines | 2.09 (1.85 to 2.35) |
| Kingdom of Spain | 2.08 (1.87 to 2.23) |
| United States of America | 2.06 (1.87 to 2.18) |
| United States Virgin Islands | 2.02 (1.56 to 2.57) |
| Burkina Faso | 2.02 (0.98 to 3.85) |
| Republic of Suriname | 2.02 (1.41 to 2.79) |
| Kingdom of Thailand | 2.01 (1.55 to 2.58) |
| Union of the Comoros | 1.97 (0.74 to 3.60) |
| Solomon Islands | 1.97 (1.35 to 2.84) |
| Kingdom of Lesotho | 1.96 (1.14 to 2.97) |
| Republic of Liberia | 1.95 (0.86 to 3.70) |
| Republic of Djibouti | 1.94 (0.86 to 3.09) |
| Antigua and Barbuda | 1.92 (1.75 to 2.15) |
| Republic of Korea | 1.92 (1.56 to 2.27) |
| Bolivarian Republic of Venezuela | 1.91 (1.50 to 2.37) |
| State of Eritrea | 1.90 (0.84 to 3.58) |
| Romania | 1.90 (1.65 to 2.18) |
| French Republic | 1.89 (1.67 to 2.04) |
| Republic of Sierra Leone | 1.89 (0.82 to 3.62) |
| Republic of Guinea | 1.89 (0.70 to 3.66) |
| Turkmenistan | 1.88 (1.42 to 2.67) |
| Republic of Malawi | 1.85 (0.93 to 3.18) |
| Republic of Rwanda | 1.84 (1.12 to 3.04) |
| Greenland | 1.84 (1.49 to 2.30) |
| Republic of Austria | 1.80 (1.61 to 1.94) |
| Republic of Kenya | 1.78 (1.05 to 2.49) |
| Guam | 1.78 (1.51 to 2.07) |
| Republic of Panama | 1.77 (1.35 to 2.15) |
| Jamaica | 1.75 (1.36 to 2.21) |
| Republic of San Marino | 1.67 (1.06 to 2.59) |
| Dominican Republic | 1.63 (1.19 to 2.15) |
| Republic of Uganda | 1.60 (0.80 to 2.64) |
| Republic of Benin | 1.60 (0.65 to 2.86) |
| Independent State of Papua New Guinea | 1.58 (1.07 to 2.25) |
| Portuguese Republic | 1.51 (1.37 to 1.62) |
| Republic of Chad | 1.50 (0.66 to 2.99) |
| Republic of Moldova | 1.49 (1.32 to 1.67) |
| Kingdom of Bhutan | 1.49 (0.93 to 2.16) |
| Islamic Republic of Pakistan | 1.48 (1.06 to 2.04) |
| United Arab Emirates | 1.45 (1.17 to 1.84) |
| Republic of Burundi | 1.44 (0.62 to 2.67) |
| Republic of South Sudan | 1.39 (0.62 to 2.68) |
| Republic of Azerbaijan | 1.37 (0.93 to 2.08) |
| Plurinational State of Bolivia | 1.36 (1.03 to 1.84) |
| Republic of the Philippines | 1.35 (1.12 to 1.58) |
| Republic of Ecuador | 1.32 (1.06 to 1.66) |
| Republic of Malta | 1.32 (1.17 to 1.47) |
| State of Israel | 1.28 (1.13 to 1.40) |
| People's Republic of Bangladesh | 1.21 (0.80 to 1.99) |
| Republic of India | 1.20 (0.86 to 1.74) |
| Republic of Honduras | 1.19 (0.87 to 1.60) |
| Federal Democratic Republic of Nepal | 1.19 (0.82 to 1.85) |
| Republic of Mali | 1.18 (0.51 to 2.27) |
| Republic of the Niger | 1.17 (0.45 to 2.69) |
| Federal Democratic Republic of Ethiopia | 1.10 (0.57 to 1.87) |
| Republic of Albania | 1.09 (0.76 to 1.50) |
| Socialist Republic of Viet Nam | 1.07 (0.78 to 1.42) |
| Republic of Seychelles | 1.07 (0.74 to 1.51) |
| Republic of Uzbekistan | 1.04 (0.85 to 1.28) |
| Republic of Indonesia | 1.03 (0.71 to 1.36) |
| Hashemite Kingdom of Jordan | 0.97 (0.76 to 1.24) |
| Republic of the Union of Myanmar | 0.96 (0.73 to 1.26) |
| State of Qatar | 0.94 (0.59 to 1.46) |
| Lao People's Democratic Republic | 0.93 (0.68 to 1.28) |
| Puerto Rico | 0.93 (0.77 to 1.08) |
| Republic of Mauritius | 0.88 (0.81 to 0.94) |
| Belize | 0.88 (0.76 to 0.99) |
| Federal Republic of Somalia | 0.86 (0.41 to 2.05) |
| State of Kuwait | 0.86 (0.71 to 1.06) |
| Kyrgyz Republic | 0.79 (0.63 to 0.96) |
| Palestine | 0.77 (0.58 to 0.98) |
| Kingdom of Cambodia | 0.75 (0.47 to 1.21) |
| Democratic Republic of Timor-Leste | 0.74 (0.49 to 1.15) |
| Kingdom of Bahrain | 0.71 (0.56 to 0.90) |
| Mongolia | 0.70 (0.53 to 0.89) |
| Republic of Peru | 0.66 (0.49 to 0.87) |
| United Mexican States | 0.63 (0.56 to 0.71) |
| Republic of Maldives | 0.60 (0.31 to 0.97) |
| Sultanate of Oman | 0.56 (0.35 to 0.88) |
| Democratic People's Republic of Korea | 0.55 (0.41 to 0.72) |
| Islamic Republic of Iran | 0.53 (0.47 to 0.60) |
| Republic of El Salvador | 0.53 (0.42 to 0.68) |
| People's Republic of China | 0.46 (0.36 to 0.59) |
| Republic of Kiribati | 0.45 (0.34 to 0.58) |
| Republic of Tunisia | 0.45 (0.29 to 0.66) |
| Arab Republic of Egypt | 0.45 (0.36 to 0.56) |
| Kingdom of Morocco | 0.44 (0.31 to 0.57) |
| Republic of Guatemala | 0.43 (0.37 to 0.50) |
| Syrian Arab Republic | 0.42 (0.31 to 0.56) |
| People's Democratic Republic of Algeria | 0.42 (0.32 to 0.55) |
| Republic of Iraq | 0.38 (0.28 to 0.48) |
| Democratic Socialist Republic of Sri Lanka | 0.37 (0.25 to 0.50) |
| State of Libya | 0.35 (0.21 to 0.54) |
| Republic of Nicaragua | 0.34 (0.28 to 0.42) |
| Republic of Sudan | 0.34 (0.22 to 0.49) |
| Republic of Yemen | 0.33 (0.20 to 0.51) |
| Islamic Republic of Afghanistan | 0.29 (0.19 to 0.42) |
| Republic of Tajikistan | 0.25 (0.18 to 0.34) |
| Kingdom of Saudi Arabia | 0.21 (0.16 to 0.28) |

### Data for Figure 2

#### Fig 2A. ASDR across Four World Regions from 1990 to 2021

| **Year** | **Location** | **ASDR per 100,000** |
| --- | --- | --- |
| 1990 | Africa | 1.82 (1.14 to 2.82) |
| 1991 | Africa | 1.82 (1.13 to 2.84) |
| 1992 | Africa | 1.84 (1.16 to 2.88) |
| 1993 | Africa | 1.85 (1.16 to 2.91) |
| 1994 | Africa | 1.86 (1.18 to 2.88) |
| 1995 | Africa | 1.87 (1.18 to 2.90) |
| 1996 | Africa | 1.89 (1.20 to 2.89) |
| 1997 | Africa | 1.90 (1.22 to 2.90) |
| 1998 | Africa | 1.89 (1.22 to 2.94) |
| 1999 | Africa | 1.87 (1.21 to 2.88) |
| 2000 | Africa | 1.86 (1.20 to 2.91) |
| 2001 | Africa | 1.83 (1.19 to 2.83) |
| 2002 | Africa | 1.79 (1.16 to 2.79) |
| 2003 | Africa | 1.74 (1.14 to 2.70) |
| 2004 | Africa | 1.70 (1.11 to 2.64) |
| 2005 | Africa | 1.66 (1.08 to 2.59) |
| 2006 | Africa | 1.64 (1.07 to 2.53) |
| 2007 | Africa | 1.61 (1.04 to 2.48) |
| 2008 | Africa | 1.60 (1.03 to 2.46) |
| 2009 | Africa | 1.59 (1.02 to 2.44) |
| 2010 | Africa | 1.58 (1.02 to 2.43) |
| 2011 | Africa | 1.58 (1.02 to 2.44) |
| 2012 | Africa | 1.58 (1.01 to 2.44) |
| 2013 | Africa | 1.59 (1.01 to 2.41) |
| 2014 | Africa | 1.59 (1.01 to 2.42) |
| 2015 | Africa | 1.60 (1.01 to 2.45) |
| 2016 | Africa | 1.61 (1.02 to 2.44) |
| 2017 | Africa | 1.62 (1.02 to 2.45) |
| 2018 | Africa | 1.63 (1.03 to 2.46) |
| 2019 | Africa | 1.64 (1.03 to 2.46) |
| 2020 | Africa | 1.65 (1.05 to 2.47) |
| 2021 | Africa | 1.65 (1.04 to 2.47) |
| 1990 | America | 4.43 (4.14 to 4.59) |
| 1991 | America | 4.42 (4.13 to 4.58) |
| 1992 | America | 4.35 (4.07 to 4.50) |
| 1993 | America | 4.33 (4.05 to 4.48) |
| 1994 | America | 4.28 (4.00 to 4.42) |
| 1995 | America | 4.26 (3.98 to 4.40) |
| 1996 | America | 4.20 (3.92 to 4.35) |
| 1997 | America | 4.14 (3.87 to 4.29) |
| 1998 | America | 4.06 (3.79 to 4.21) |
| 1999 | America | 4.01 (3.73 to 4.14) |
| 2000 | America | 3.92 (3.65 to 4.05) |
| 2001 | America | 3.81 (3.54 to 3.94) |
| 2002 | America | 3.71 (3.45 to 3.84) |
| 2003 | America | 3.63 (3.37 to 3.75) |
| 2004 | America | 3.48 (3.22 to 3.60) |
| 2005 | America | 3.38 (3.13 to 3.51) |
| 2006 | America | 3.26 (3.01 to 3.38) |
| 2007 | America | 3.13 (2.89 to 3.26) |
| 2008 | America | 2.94 (2.71 to 3.05) |
| 2009 | America | 2.81 (2.59 to 2.92) |
| 2010 | America | 2.70 (2.49 to 2.81) |
| 2011 | America | 2.62 (2.41 to 2.72) |
| 2012 | America | 2.54 (2.33 to 2.64) |
| 2013 | America | 2.49 (2.28 to 2.58) |
| 2014 | America | 2.45 (2.25 to 2.55) |
| 2015 | America | 2.43 (2.23 to 2.52) |
| 2016 | America | 2.41 (2.22 to 2.51) |
| 2017 | America | 2.36 (2.16 to 2.45) |
| 2018 | America | 2.33 (2.13 to 2.42) |
| 2019 | America | 2.33 (2.13 to 2.42) |
| 2020 | America | 2.32 (2.12 to 2.42) |
| 2021 | America | 2.33 (2.12 to 2.45) |
| 1990 | Asia | 1.01 (0.87 to 1.18) |
| 1991 | Asia | 1.03 (0.89 to 1.21) |
| 1992 | Asia | 1.05 (0.90 to 1.23) |
| 1993 | Asia | 1.07 (0.93 to 1.26) |
| 1994 | Asia | 1.09 (0.95 to 1.27) |
| 1995 | Asia | 1.12 (0.98 to 1.31) |
| 1996 | Asia | 1.14 (0.99 to 1.32) |
| 1997 | Asia | 1.14 (1.00 to 1.32) |
| 1998 | Asia | 1.16 (1.01 to 1.34) |
| 1999 | Asia | 1.17 (1.03 to 1.35) |
| 2000 | Asia | 1.18 (1.03 to 1.36) |
| 2001 | Asia | 1.21 (1.06 to 1.39) |
| 2002 | Asia | 1.22 (1.06 to 1.40) |
| 2003 | Asia | 1.24 (1.08 to 1.42) |
| 2004 | Asia | 1.27 (1.10 to 1.45) |
| 2005 | Asia | 1.28 (1.12 to 1.46) |
| 2006 | Asia | 1.29 (1.12 to 1.47) |
| 2007 | Asia | 1.32 (1.15 to 1.50) |
| 2008 | Asia | 1.34 (1.16 to 1.53) |
| 2009 | Asia | 1.36 (1.19 to 1.55) |
| 2010 | Asia | 1.40 (1.21 to 1.58) |
| 2011 | Asia | 1.41 (1.23 to 1.59) |
| 2012 | Asia | 1.39 (1.20 to 1.58) |
| 2013 | Asia | 1.39 (1.20 to 1.59) |
| 2014 | Asia | 1.40 (1.21 to 1.61) |
| 2015 | Asia | 1.41 (1.22 to 1.61) |
| 2016 | Asia | 1.44 (1.22 to 1.62) |
| 2017 | Asia | 1.45 (1.23 to 1.64) |
| 2018 | Asia | 1.44 (1.23 to 1.63) |
| 2019 | Asia | 1.43 (1.23 to 1.63) |
| 2020 | Asia | 1.40 (1.19 to 1.58) |
| 2021 | Asia | 1.40 (1.20 to 1.59) |
| 1990 | Europe | 3.95 (3.74 to 4.06) |
| 1991 | Europe | 4.04 (3.82 to 4.15) |
| 1992 | Europe | 4.12 (3.90 to 4.23) |
| 1993 | Europe | 4.28 (4.04 to 4.38) |
| 1994 | Europe | 4.36 (4.13 to 4.47) |
| 1995 | Europe | 4.37 (4.13 to 4.48) |
| 1996 | Europe | 4.32 (4.08 to 4.43) |
| 1997 | Europe | 4.22 (3.98 to 4.34) |
| 1998 | Europe | 4.18 (3.95 to 4.30) |
| 1999 | Europe | 4.31 (4.07 to 4.43) |
| 2000 | Europe | 4.23 (3.99 to 4.35) |
| 2001 | Europe | 4.01 (3.76 to 4.13) |
| 2002 | Europe | 3.97 (3.73 to 4.09) |
| 2003 | Europe | 3.93 (3.68 to 4.05) |
| 2004 | Europe | 3.84 (3.61 to 3.97) |
| 2005 | Europe | 3.79 (3.55 to 3.92) |
| 2006 | Europe | 3.68 (3.44 to 3.81) |
| 2007 | Europe | 3.62 (3.39 to 3.75) |
| 2008 | Europe | 3.57 (3.34 to 3.69) |
| 2009 | Europe | 3.46 (3.24 to 3.58) |
| 2010 | Europe | 3.38 (3.17 to 3.50) |
| 2011 | Europe | 3.27 (3.06 to 3.39) |
| 2012 | Europe | 3.23 (3.01 to 3.34) |
| 2013 | Europe | 3.18 (2.96 to 3.29) |
| 2014 | Europe | 3.16 (2.95 to 3.27) |
| 2015 | Europe | 3.15 (2.92 to 3.25) |
| 2016 | Europe | 3.14 (2.91 to 3.25) |
| 2017 | Europe | 3.07 (2.86 to 3.18) |
| 2018 | Europe | 3.07 (2.86 to 3.19) |
| 2019 | Europe | 3.04 (2.81 to 3.16) |
| 2020 | Europe | 3.00 (2.78 to 3.15) |
| 2021 | Europe | 2.97 (2.76 to 3.15) |

#### Fig 2B. ASDR across Health System Groupings from 1990 to 2021

| **Year** | **Location** | **ASDR per 100,000** |
| --- | --- | --- |
| 1990 | Advanced Health System | 4.15 (3.90 to 4.28) |
| 1991 | Advanced Health System | 4.21 (3.94 to 4.34) |
| 1992 | Advanced Health System | 4.23 (3.97 to 4.35) |
| 1993 | Advanced Health System | 4.32 (4.05 to 4.44) |
| 1994 | Advanced Health System | 4.36 (4.09 to 4.48) |
| 1995 | Advanced Health System | 4.36 (4.09 to 4.49) |
| 1996 | Advanced Health System | 4.31 (4.03 to 4.44) |
| 1997 | Advanced Health System | 4.23 (3.95 to 4.36) |
| 1998 | Advanced Health System | 4.19 (3.91 to 4.32) |
| 1999 | Advanced Health System | 4.25 (3.96 to 4.38) |
| 2000 | Advanced Health System | 4.17 (3.88 to 4.31) |
| 2001 | Advanced Health System | 4.01 (3.72 to 4.15) |
| 2002 | Advanced Health System | 3.96 (3.67 to 4.10) |
| 2003 | Advanced Health System | 3.90 (3.61 to 4.04) |
| 2004 | Advanced Health System | 3.81 (3.52 to 3.96) |
| 2005 | Advanced Health System | 3.76 (3.47 to 3.91) |
| 2006 | Advanced Health System | 3.65 (3.36 to 3.80) |
| 2007 | Advanced Health System | 3.58 (3.29 to 3.73) |
| 2008 | Advanced Health System | 3.49 (3.20 to 3.64) |
| 2009 | Advanced Health System | 3.40 (3.11 to 3.55) |
| 2010 | Advanced Health System | 3.34 (3.05 to 3.49) |
| 2011 | Advanced Health System | 3.26 (2.97 to 3.40) |
| 2012 | Advanced Health System | 3.20 (2.91 to 3.34) |
| 2013 | Advanced Health System | 3.15 (2.86 to 3.29) |
| 2014 | Advanced Health System | 3.13 (2.84 to 3.28) |
| 2015 | Advanced Health System | 3.11 (2.81 to 3.26) |
| 2016 | Advanced Health System | 3.12 (2.82 to 3.27) |
| 2017 | Advanced Health System | 3.09 (2.78 to 3.24) |
| 2018 | Advanced Health System | 3.07 (2.76 to 3.22) |
| 2019 | Advanced Health System | 3.04 (2.73 to 3.19) |
| 2020 | Advanced Health System | 2.98 (2.68 to 3.16) |
| 2021 | Advanced Health System | 2.99 (2.69 to 3.17) |
| 1990 | Basic Health System | 1.02 (0.94 to 1.11) |
| 1991 | Basic Health System | 1.03 (0.95 to 1.11) |
| 1992 | Basic Health System | 1.06 (0.98 to 1.14) |
| 1993 | Basic Health System | 1.08 (1.00 to 1.16) |
| 1994 | Basic Health System | 1.09 (1.01 to 1.17) |
| 1995 | Basic Health System | 1.10 (1.02 to 1.18) |
| 1996 | Basic Health System | 1.11 (1.04 to 1.18) |
| 1997 | Basic Health System | 1.11 (1.04 to 1.18) |
| 1998 | Basic Health System | 1.11 (1.04 to 1.18) |
| 1999 | Basic Health System | 1.12 (1.04 to 1.18) |
| 2000 | Basic Health System | 1.13 (1.05 to 1.19) |
| 2001 | Basic Health System | 1.12 (1.05 to 1.19) |
| 2002 | Basic Health System | 1.13 (1.05 to 1.19) |
| 2003 | Basic Health System | 1.14 (1.06 to 1.20) |
| 2004 | Basic Health System | 1.13 (1.05 to 1.19) |
| 2005 | Basic Health System | 1.12 (1.05 to 1.18) |
| 2006 | Basic Health System | 1.12 (1.04 to 1.17) |
| 2007 | Basic Health System | 1.12 (1.05 to 1.17) |
| 2008 | Basic Health System | 1.12 (1.04 to 1.17) |
| 2009 | Basic Health System | 1.11 (1.04 to 1.16) |
| 2010 | Basic Health System | 1.11 (1.04 to 1.16) |
| 2011 | Basic Health System | 1.10 (1.03 to 1.14) |
| 2012 | Basic Health System | 1.08 (1.00 to 1.13) |
| 2013 | Basic Health System | 1.06 (0.99 to 1.12) |
| 2014 | Basic Health System | 1.06 (0.99 to 1.12) |
| 2015 | Basic Health System | 1.06 (0.99 to 1.12) |
| 2016 | Basic Health System | 1.07 (0.98 to 1.13) |
| 2017 | Basic Health System | 1.05 (0.97 to 1.12) |
| 2018 | Basic Health System | 1.05 (0.97 to 1.13) |
| 2019 | Basic Health System | 1.06 (0.97 to 1.14) |
| 2020 | Basic Health System | 1.05 (0.96 to 1.15) |
| 2021 | Basic Health System | 1.05 (0.95 to 1.15) |
| 1990 | Limited Health System | 0.99 (0.73 to 1.45) |
| 1991 | Limited Health System | 0.99 (0.74 to 1.46) |
| 1992 | Limited Health System | 1.01 (0.75 to 1.48) |
| 1993 | Limited Health System | 1.02 (0.77 to 1.49) |
| 1994 | Limited Health System | 1.04 (0.79 to 1.52) |
| 1995 | Limited Health System | 1.07 (0.81 to 1.58) |
| 1996 | Limited Health System | 1.08 (0.81 to 1.56) |
| 1997 | Limited Health System | 1.07 (0.81 to 1.55) |
| 1998 | Limited Health System | 1.06 (0.81 to 1.52) |
| 1999 | Limited Health System | 1.06 (0.81 to 1.53) |
| 2000 | Limited Health System | 1.05 (0.81 to 1.50) |
| 2001 | Limited Health System | 1.05 (0.81 to 1.51) |
| 2002 | Limited Health System | 1.05 (0.81 to 1.50) |
| 2003 | Limited Health System | 1.06 (0.81 to 1.52) |
| 2004 | Limited Health System | 1.08 (0.82 to 1.55) |
| 2005 | Limited Health System | 1.07 (0.82 to 1.51) |
| 2006 | Limited Health System | 1.07 (0.83 to 1.52) |
| 2007 | Limited Health System | 1.09 (0.85 to 1.55) |
| 2008 | Limited Health System | 1.09 (0.87 to 1.54) |
| 2009 | Limited Health System | 1.10 (0.87 to 1.55) |
| 2010 | Limited Health System | 1.12 (0.89 to 1.57) |
| 2011 | Limited Health System | 1.13 (0.89 to 1.61) |
| 2012 | Limited Health System | 1.15 (0.90 to 1.64) |
| 2013 | Limited Health System | 1.19 (0.94 to 1.68) |
| 2014 | Limited Health System | 1.23 (0.98 to 1.76) |
| 2015 | Limited Health System | 1.26 (0.99 to 1.79) |
| 2016 | Limited Health System | 1.27 (1.00 to 1.81) |
| 2017 | Limited Health System | 1.30 (1.03 to 1.83) |
| 2018 | Limited Health System | 1.31 (1.05 to 1.84) |
| 2019 | Limited Health System | 1.33 (1.07 to 1.88) |
| 2020 | Limited Health System | 1.33 (1.07 to 1.89) |
| 2021 | Limited Health System | 1.32 (1.06 to 1.83) |
| 1990 | Minimal Health System | 1.71 (0.93 to 3.14) |
| 1991 | Minimal Health System | 1.71 (0.93 to 3.15) |
| 1992 | Minimal Health System | 1.71 (0.93 to 3.13) |
| 1993 | Minimal Health System | 1.71 (0.93 to 3.12) |
| 1994 | Minimal Health System | 1.71 (0.93 to 3.13) |
| 1995 | Minimal Health System | 1.71 (0.93 to 3.13) |
| 1996 | Minimal Health System | 1.71 (0.93 to 3.14) |
| 1997 | Minimal Health System | 1.69 (0.91 to 3.11) |
| 1998 | Minimal Health System | 1.68 (0.90 to 3.09) |
| 1999 | Minimal Health System | 1.67 (0.88 to 3.08) |
| 2000 | Minimal Health System | 1.66 (0.87 to 3.08) |
| 2001 | Minimal Health System | 1.64 (0.86 to 3.08) |
| 2002 | Minimal Health System | 1.60 (0.83 to 3.04) |
| 2003 | Minimal Health System | 1.57 (0.80 to 2.97) |
| 2004 | Minimal Health System | 1.54 (0.79 to 2.92) |
| 2005 | Minimal Health System | 1.52 (0.78 to 2.89) |
| 2006 | Minimal Health System | 1.52 (0.78 to 2.92) |
| 2007 | Minimal Health System | 1.51 (0.78 to 2.92) |
| 2008 | Minimal Health System | 1.52 (0.79 to 2.94) |
| 2009 | Minimal Health System | 1.53 (0.79 to 2.96) |
| 2010 | Minimal Health System | 1.55 (0.79 to 2.97) |
| 2011 | Minimal Health System | 1.57 (0.80 to 2.99) |
| 2012 | Minimal Health System | 1.58 (0.80 to 2.99) |
| 2013 | Minimal Health System | 1.60 (0.81 to 3.01) |
| 2014 | Minimal Health System | 1.61 (0.81 to 3.01) |
| 2015 | Minimal Health System | 1.62 (0.82 to 3.02) |
| 2016 | Minimal Health System | 1.63 (0.82 to 3.05) |
| 2017 | Minimal Health System | 1.64 (0.82 to 3.06) |
| 2018 | Minimal Health System | 1.65 (0.82 to 3.07) |
| 2019 | Minimal Health System | 1.67 (0.82 to 3.08) |
| 2020 | Minimal Health System | 1.68 (0.83 to 3.09) |
| 2021 | Minimal Health System | 1.70 (0.84 to 3.09) |

#### Fig 2C. ASDR across different Commonwealth Income regions from 1990 to 2021

| **Year** | **Location** | **ASDR per 100,000** |
| --- | --- | --- |
| 1990 | Commonwealth High Income | 8.66 (8.18 to 8.94) |
| 1991 | Commonwealth High Income | 8.67 (8.18 to 8.95) |
| 1992 | Commonwealth High Income | 8.78 (8.26 to 9.06) |
| 1993 | Commonwealth High Income | 8.79 (8.28 to 9.06) |
| 1994 | Commonwealth High Income | 8.71 (8.19 to 8.99) |
| 1995 | Commonwealth High Income | 8.74 (8.23 to 9.02) |
| 1996 | Commonwealth High Income | 8.69 (8.19 to 8.96) |
| 1997 | Commonwealth High Income | 8.49 (7.98 to 8.77) |
| 1998 | Commonwealth High Income | 8.38 (7.86 to 8.66) |
| 1999 | Commonwealth High Income | 8.21 (7.71 to 8.49) |
| 2000 | Commonwealth High Income | 7.96 (7.46 to 8.25) |
| 2001 | Commonwealth High Income | 7.65 (7.13 to 7.94) |
| 2002 | Commonwealth High Income | 7.40 (6.89 to 7.67) |
| 2003 | Commonwealth High Income | 7.10 (6.61 to 7.37) |
| 2004 | Commonwealth High Income | 6.79 (6.29 to 7.06) |
| 2005 | Commonwealth High Income | 6.44 (5.93 to 6.69) |
| 2006 | Commonwealth High Income | 6.14 (5.63 to 6.40) |
| 2007 | Commonwealth High Income | 5.85 (5.35 to 6.10) |
| 2008 | Commonwealth High Income | 5.51 (5.02 to 5.74) |
| 2009 | Commonwealth High Income | 5.15 (4.68 to 5.38) |
| 2010 | Commonwealth High Income | 4.87 (4.43 to 5.09) |
| 2011 | Commonwealth High Income | 4.55 (4.13 to 4.75) |
| 2012 | Commonwealth High Income | 4.36 (3.94 to 4.55) |
| 2013 | Commonwealth High Income | 4.15 (3.76 to 4.34) |
| 2014 | Commonwealth High Income | 4.00 (3.62 to 4.18) |
| 2015 | Commonwealth High Income | 3.84 (3.47 to 4.02) |
| 2016 | Commonwealth High Income | 3.71 (3.35 to 3.88) |
| 2017 | Commonwealth High Income | 3.57 (3.22 to 3.73) |
| 2018 | Commonwealth High Income | 3.48 (3.13 to 3.64) |
| 2019 | Commonwealth High Income | 3.37 (3.03 to 3.53) |
| 2020 | Commonwealth High Income | 3.22 (2.87 to 3.38) |
| 2021 | Commonwealth High Income | 3.22 (2.86 to 3.40) |
| 1990 | Commonwealth Low Income | 1.27 (0.80 to 2.11) |
| 1991 | Commonwealth Low Income | 1.28 (0.80 to 2.14) |
| 1992 | Commonwealth Low Income | 1.31 (0.81 to 2.18) |
| 1993 | Commonwealth Low Income | 1.33 (0.82 to 2.21) |
| 1994 | Commonwealth Low Income | 1.34 (0.82 to 2.24) |
| 1995 | Commonwealth Low Income | 1.36 (0.83 to 2.28) |
| 1996 | Commonwealth Low Income | 1.36 (0.83 to 2.29) |
| 1997 | Commonwealth Low Income | 1.37 (0.84 to 2.30) |
| 1998 | Commonwealth Low Income | 1.38 (0.84 to 2.27) |
| 1999 | Commonwealth Low Income | 1.40 (0.86 to 2.31) |
| 2000 | Commonwealth Low Income | 1.41 (0.86 to 2.31) |
| 2001 | Commonwealth Low Income | 1.40 (0.86 to 2.28) |
| 2002 | Commonwealth Low Income | 1.40 (0.87 to 2.27) |
| 2003 | Commonwealth Low Income | 1.39 (0.87 to 2.26) |
| 2004 | Commonwealth Low Income | 1.38 (0.86 to 2.27) |
| 2005 | Commonwealth Low Income | 1.37 (0.85 to 2.25) |
| 2006 | Commonwealth Low Income | 1.38 (0.86 to 2.27) |
| 2007 | Commonwealth Low Income | 1.40 (0.89 to 2.26) |
| 2008 | Commonwealth Low Income | 1.41 (0.89 to 2.26) |
| 2009 | Commonwealth Low Income | 1.41 (0.90 to 2.27) |
| 2010 | Commonwealth Low Income | 1.42 (0.91 to 2.26) |
| 2011 | Commonwealth Low Income | 1.39 (0.87 to 2.25) |
| 2012 | Commonwealth Low Income | 1.35 (0.84 to 2.20) |
| 2013 | Commonwealth Low Income | 1.35 (0.84 to 2.19) |
| 2014 | Commonwealth Low Income | 1.38 (0.86 to 2.23) |
| 2015 | Commonwealth Low Income | 1.39 (0.88 to 2.25) |
| 2016 | Commonwealth Low Income | 1.41 (0.88 to 2.31) |
| 2017 | Commonwealth Low Income | 1.47 (0.92 to 2.33) |
| 2018 | Commonwealth Low Income | 1.49 (0.94 to 2.34) |
| 2019 | Commonwealth Low Income | 1.51 (0.97 to 2.36) |
| 2020 | Commonwealth Low Income | 1.52 (0.98 to 2.38) |
| 2021 | Commonwealth Low Income | 1.53 (0.98 to 2.38) |
| 1990 | Commonwealth Middle Income | 1.06 (0.83 to 1.45) |
| 1991 | Commonwealth Middle Income | 1.07 (0.84 to 1.47) |
| 1992 | Commonwealth Middle Income | 1.09 (0.87 to 1.48) |
| 1993 | Commonwealth Middle Income | 1.11 (0.88 to 1.51) |
| 1994 | Commonwealth Middle Income | 1.13 (0.90 to 1.54) |
| 1995 | Commonwealth Middle Income | 1.18 (0.93 to 1.60) |
| 1996 | Commonwealth Middle Income | 1.19 (0.94 to 1.59) |
| 1997 | Commonwealth Middle Income | 1.19 (0.95 to 1.60) |
| 1998 | Commonwealth Middle Income | 1.18 (0.95 to 1.58) |
| 1999 | Commonwealth Middle Income | 1.17 (0.94 to 1.56) |
| 2000 | Commonwealth Middle Income | 1.16 (0.94 to 1.55) |
| 2001 | Commonwealth Middle Income | 1.15 (0.94 to 1.54) |
| 2002 | Commonwealth Middle Income | 1.14 (0.93 to 1.54) |
| 2003 | Commonwealth Middle Income | 1.16 (0.94 to 1.56) |
| 2004 | Commonwealth Middle Income | 1.18 (0.95 to 1.59) |
| 2005 | Commonwealth Middle Income | 1.16 (0.94 to 1.58) |
| 2006 | Commonwealth Middle Income | 1.15 (0.94 to 1.57) |
| 2007 | Commonwealth Middle Income | 1.17 (0.96 to 1.59) |
| 2008 | Commonwealth Middle Income | 1.17 (0.96 to 1.58) |
| 2009 | Commonwealth Middle Income | 1.18 (0.96 to 1.61) |
| 2010 | Commonwealth Middle Income | 1.19 (0.97 to 1.62) |
| 2011 | Commonwealth Middle Income | 1.21 (0.98 to 1.65) |
| 2012 | Commonwealth Middle Income | 1.23 (0.99 to 1.67) |
| 2013 | Commonwealth Middle Income | 1.26 (1.03 to 1.73) |
| 2014 | Commonwealth Middle Income | 1.31 (1.07 to 1.81) |
| 2015 | Commonwealth Middle Income | 1.35 (1.08 to 1.85) |
| 2016 | Commonwealth Middle Income | 1.36 (1.10 to 1.85) |
| 2017 | Commonwealth Middle Income | 1.38 (1.13 to 1.85) |
| 2018 | Commonwealth Middle Income | 1.39 (1.14 to 1.87) |
| 2019 | Commonwealth Middle Income | 1.41 (1.15 to 1.90) |
| 2020 | Commonwealth Middle Income | 1.39 (1.13 to 1.89) |
| 2021 | Commonwealth Middle Income | 1.39 (1.13 to 1.85) |

#### Fig 2D. ASDR across SDI regions from 1990 to 2021

| **Year** | **Location** | **ASDR per 100,000** |
| --- | --- | --- |
| 1990 | High SDI | 4.76 (4.46 to 4.91) |
| 1991 | High SDI | 4.81 (4.49 to 4.96) |
| 1992 | High SDI | 4.77 (4.45 to 4.92) |
| 1993 | High SDI | 4.76 (4.45 to 4.91) |
| 1994 | High SDI | 4.73 (4.42 to 4.89) |
| 1995 | High SDI | 4.74 (4.43 to 4.90) |
| 1996 | High SDI | 4.71 (4.39 to 4.87) |
| 1997 | High SDI | 4.64 (4.32 to 4.79) |
| 1998 | High SDI | 4.59 (4.26 to 4.74) |
| 1999 | High SDI | 4.53 (4.19 to 4.69) |
| 2000 | High SDI | 4.45 (4.11 to 4.62) |
| 2001 | High SDI | 4.36 (4.01 to 4.53) |
| 2002 | High SDI | 4.27 (3.93 to 4.44) |
| 2003 | High SDI | 4.17 (3.82 to 4.34) |
| 2004 | High SDI | 4.05 (3.70 to 4.22) |
| 2005 | High SDI | 3.95 (3.60 to 4.12) |
| 2006 | High SDI | 3.83 (3.49 to 4.00) |
| 2007 | High SDI | 3.72 (3.38 to 3.89) |
| 2008 | High SDI | 3.58 (3.24 to 3.75) |
| 2009 | High SDI | 3.47 (3.13 to 3.64) |
| 2010 | High SDI | 3.39 (3.04 to 3.56) |
| 2011 | High SDI | 3.30 (2.96 to 3.47) |
| 2012 | High SDI | 3.21 (2.87 to 3.38) |
| 2013 | High SDI | 3.14 (2.80 to 3.30) |
| 2014 | High SDI | 3.08 (2.74 to 3.25) |
| 2015 | High SDI | 3.04 (2.70 to 3.20) |
| 2016 | High SDI | 3.04 (2.70 to 3.22) |
| 2017 | High SDI | 3.00 (2.66 to 3.18) |
| 2018 | High SDI | 2.96 (2.61 to 3.14) |
| 2019 | High SDI | 2.92 (2.57 to 3.10) |
| 2020 | High SDI | 2.85 (2.50 to 3.05) |
| 2021 | High SDI | 2.87 (2.51 to 3.06) |
| 1990 | High-middle SDI | 1.99 (1.88 to 2.08) |
| 1991 | High-middle SDI | 2.03 (1.93 to 2.12) |
| 1992 | High-middle SDI | 2.10 (1.98 to 2.19) |
| 1993 | High-middle SDI | 2.23 (2.12 to 2.32) |
| 1994 | High-middle SDI | 2.32 (2.21 to 2.40) |
| 1995 | High-middle SDI | 2.30 (2.19 to 2.38) |
| 1996 | High-middle SDI | 2.23 (2.12 to 2.32) |
| 1997 | High-middle SDI | 2.17 (2.06 to 2.25) |
| 1998 | High-middle SDI | 2.15 (2.04 to 2.23) |
| 1999 | High-middle SDI | 2.32 (2.21 to 2.39) |
| 2000 | High-middle SDI | 2.25 (2.13 to 2.32) |
| 2001 | High-middle SDI | 2.06 (1.95 to 2.13) |
| 2002 | High-middle SDI | 2.06 (1.95 to 2.12) |
| 2003 | High-middle SDI | 2.07 (1.97 to 2.14) |
| 2004 | High-middle SDI | 2.03 (1.92 to 2.10) |
| 2005 | High-middle SDI | 2.05 (1.94 to 2.12) |
| 2006 | High-middle SDI | 2.00 (1.89 to 2.07) |
| 2007 | High-middle SDI | 2.00 (1.89 to 2.07) |
| 2008 | High-middle SDI | 1.99 (1.89 to 2.06) |
| 2009 | High-middle SDI | 1.96 (1.86 to 2.03) |
| 2010 | High-middle SDI | 1.95 (1.84 to 2.01) |
| 2011 | High-middle SDI | 1.91 (1.81 to 1.97) |
| 2012 | High-middle SDI | 1.89 (1.79 to 1.96) |
| 2013 | High-middle SDI | 1.87 (1.77 to 1.94) |
| 2014 | High-middle SDI | 1.89 (1.79 to 1.97) |
| 2015 | High-middle SDI | 1.90 (1.79 to 1.98) |
| 2016 | High-middle SDI | 1.90 (1.78 to 1.99) |
| 2017 | High-middle SDI | 1.87 (1.75 to 1.96) |
| 2018 | High-middle SDI | 1.87 (1.75 to 1.96) |
| 2019 | High-middle SDI | 1.85 (1.73 to 1.95) |
| 2020 | High-middle SDI | 1.82 (1.70 to 1.93) |
| 2021 | High-middle SDI | 1.79 (1.66 to 1.92) |
| 1990 | Low SDI | 1.37 (0.83 to 2.37) |
| 1991 | Low SDI | 1.37 (0.83 to 2.36) |
| 1992 | Low SDI | 1.38 (0.84 to 2.37) |
| 1993 | Low SDI | 1.39 (0.84 to 2.40) |
| 1994 | Low SDI | 1.40 (0.85 to 2.43) |
| 1995 | Low SDI | 1.41 (0.85 to 2.45) |
| 1996 | Low SDI | 1.40 (0.84 to 2.41) |
| 1997 | Low SDI | 1.38 (0.83 to 2.41) |
| 1998 | Low SDI | 1.37 (0.82 to 2.40) |
| 1999 | Low SDI | 1.35 (0.81 to 2.38) |
| 2000 | Low SDI | 1.34 (0.80 to 2.33) |
| 2001 | Low SDI | 1.32 (0.80 to 2.30) |
| 2002 | Low SDI | 1.30 (0.79 to 2.27) |
| 2003 | Low SDI | 1.29 (0.78 to 2.24) |
| 2004 | Low SDI | 1.27 (0.76 to 2.21) |
| 2005 | Low SDI | 1.25 (0.75 to 2.18) |
| 2006 | Low SDI | 1.25 (0.74 to 2.17) |
| 2007 | Low SDI | 1.25 (0.74 to 2.17) |
| 2008 | Low SDI | 1.25 (0.75 to 2.16) |
| 2009 | Low SDI | 1.26 (0.75 to 2.20) |
| 2010 | Low SDI | 1.28 (0.76 to 2.21) |
| 2011 | Low SDI | 1.30 (0.78 to 2.24) |
| 2012 | Low SDI | 1.33 (0.80 to 2.28) |
| 2013 | Low SDI | 1.39 (0.84 to 2.34) |
| 2014 | Low SDI | 1.42 (0.86 to 2.41) |
| 2015 | Low SDI | 1.44 (0.87 to 2.47) |
| 2016 | Low SDI | 1.46 (0.88 to 2.46) |
| 2017 | Low SDI | 1.49 (0.90 to 2.49) |
| 2018 | Low SDI | 1.48 (0.89 to 2.46) |
| 2019 | Low SDI | 1.49 (0.91 to 2.46) |
| 2020 | Low SDI | 1.49 (0.92 to 2.47) |
| 2021 | Low SDI | 1.48 (0.91 to 2.44) |
| 1990 | Low-middle SDI | 0.89 (0.71 to 1.20) |
| 1991 | Low-middle SDI | 0.90 (0.71 to 1.21) |
| 1992 | Low-middle SDI | 0.92 (0.73 to 1.23) |
| 1993 | Low-middle SDI | 0.93 (0.74 to 1.25) |
| 1994 | Low-middle SDI | 0.95 (0.77 to 1.27) |
| 1995 | Low-middle SDI | 0.98 (0.79 to 1.31) |
| 1996 | Low-middle SDI | 1.00 (0.80 to 1.33) |
| 1997 | Low-middle SDI | 1.00 (0.80 to 1.34) |
| 1998 | Low-middle SDI | 1.01 (0.82 to 1.34) |
| 1999 | Low-middle SDI | 1.02 (0.83 to 1.34) |
| 2000 | Low-middle SDI | 1.03 (0.84 to 1.36) |
| 2001 | Low-middle SDI | 1.03 (0.85 to 1.37) |
| 2002 | Low-middle SDI | 1.04 (0.86 to 1.38) |
| 2003 | Low-middle SDI | 1.06 (0.86 to 1.40) |
| 2004 | Low-middle SDI | 1.07 (0.88 to 1.42) |
| 2005 | Low-middle SDI | 1.07 (0.88 to 1.42) |
| 2006 | Low-middle SDI | 1.07 (0.88 to 1.42) |
| 2007 | Low-middle SDI | 1.10 (0.91 to 1.47) |
| 2008 | Low-middle SDI | 1.12 (0.92 to 1.48) |
| 2009 | Low-middle SDI | 1.13 (0.93 to 1.50) |
| 2010 | Low-middle SDI | 1.14 (0.95 to 1.52) |
| 2011 | Low-middle SDI | 1.15 (0.96 to 1.54) |
| 2012 | Low-middle SDI | 1.16 (0.97 to 1.55) |
| 2013 | Low-middle SDI | 1.19 (0.99 to 1.58) |
| 2014 | Low-middle SDI | 1.22 (1.01 to 1.65) |
| 2015 | Low-middle SDI | 1.24 (1.03 to 1.66) |
| 2016 | Low-middle SDI | 1.26 (1.06 to 1.68) |
| 2017 | Low-middle SDI | 1.29 (1.08 to 1.72) |
| 2018 | Low-middle SDI | 1.31 (1.09 to 1.73) |
| 2019 | Low-middle SDI | 1.33 (1.11 to 1.79) |
| 2020 | Low-middle SDI | 1.33 (1.10 to 1.79) |
| 2021 | Low-middle SDI | 1.31 (1.09 to 1.76) |
| 1990 | Middle SDI | 1.03 (0.94 to 1.14) |
| 1991 | Middle SDI | 1.04 (0.96 to 1.16) |
| 1992 | Middle SDI | 1.07 (0.98 to 1.19) |
| 1993 | Middle SDI | 1.10 (1.01 to 1.21) |
| 1994 | Middle SDI | 1.12 (1.04 to 1.24) |
| 1995 | Middle SDI | 1.14 (1.05 to 1.25) |
| 1996 | Middle SDI | 1.16 (1.07 to 1.27) |
| 1997 | Middle SDI | 1.15 (1.07 to 1.26) |
| 1998 | Middle SDI | 1.16 (1.07 to 1.27) |
| 1999 | Middle SDI | 1.16 (1.08 to 1.27) |
| 2000 | Middle SDI | 1.17 (1.09 to 1.28) |
| 2001 | Middle SDI | 1.18 (1.09 to 1.29) |
| 2002 | Middle SDI | 1.17 (1.08 to 1.28) |
| 2003 | Middle SDI | 1.18 (1.09 to 1.28) |
| 2004 | Middle SDI | 1.20 (1.11 to 1.30) |
| 2005 | Middle SDI | 1.18 (1.10 to 1.29) |
| 2006 | Middle SDI | 1.18 (1.09 to 1.27) |
| 2007 | Middle SDI | 1.17 (1.09 to 1.27) |
| 2008 | Middle SDI | 1.17 (1.09 to 1.26) |
| 2009 | Middle SDI | 1.17 (1.07 to 1.25) |
| 2010 | Middle SDI | 1.17 (1.07 to 1.25) |
| 2011 | Middle SDI | 1.15 (1.06 to 1.24) |
| 2012 | Middle SDI | 1.14 (1.05 to 1.22) |
| 2013 | Middle SDI | 1.13 (1.04 to 1.22) |
| 2014 | Middle SDI | 1.14 (1.04 to 1.23) |
| 2015 | Middle SDI | 1.15 (1.06 to 1.23) |
| 2016 | Middle SDI | 1.15 (1.06 to 1.24) |
| 2017 | Middle SDI | 1.14 (1.04 to 1.22) |
| 2018 | Middle SDI | 1.14 (1.04 to 1.22) |
| 2019 | Middle SDI | 1.16 (1.05 to 1.24) |
| 2020 | Middle SDI | 1.16 (1.05 to 1.26) |
| 2021 | Middle SDI | 1.15 (1.04 to 1.25) |

#### Fig 2E. ASDR in males and females across the Four World Regions in 2021

| **Sex** | **Location** | **ASDR per 100,000** |
| --- | --- | --- |
| Male | Africa | 2.16 (1.21 to 3.36) |
| Female | Africa | 1.20 (0.76 to 1.75) |
| Male | America | 3.17 (2.97 to 3.32) |
| Female | America | 1.63 (1.42 to 1.76) |
| Male | Asia | 1.81 (1.60 to 2.11) |
| Female | Asia | 1.05 (0.85 to 1.21) |
| Male | Europe | 4.69 (4.38 to 4.92) |
| Female | Europe | 1.70 (1.50 to 1.92) |

#### Fig 2F. ASDR in males and females across Health System Groupings in 2021

| **Sex** | **Location** | **ASDR per 100,000** |
| --- | --- | --- |
| Male | Basic Health System | 1.49 (1.33 to 1.65) |
| Female | Basic Health System | 0.68 (0.59 to 0.75) |
| Male | Limited Health System | 1.72 (1.35 to 2.67) |
| Female | Limited Health System | 0.96 (0.70 to 1.25) |
| Male | Minimal Health System | 2.06 (0.68 to 4.01) |
| Female | Minimal Health System | 1.38 (0.71 to 2.50) |
| Male | Advanced Health System | 4.31 (3.99 to 4.49) |
| Female | Advanced Health System | 1.94 (1.63 to 2.18) |

#### Fig 2G. ASDR in males and females across different Commonwealth Income regions in 2021

| **Sex** | **Location** | **ASDR per 100,000** |
| --- | --- | --- |
| Male | Commonwealth High Income | 4.42 (4.07 to 4.62) |
| Female | Commonwealth High Income | 2.25 (1.90 to 2.44) |
| Male | Commonwealth Middle Income | 1.85 (1.46 to 2.74) |
| Female | Commonwealth Middle Income | 0.99 (0.73 to 1.27) |
| Male | Commonwealth Low Income | 1.96 (1.17 to 3.42) |
| Female | Commonwealth Low Income | 1.12 (0.74 to 1.70) |

#### Fig 2H. ASDR in males and females across SDI regions in 2021

| **Sex** | **Location** | **ASDR per 100,000** |
| --- | --- | --- |
| Female | High SDI | 2.00 (1.63 to 2.24) |
| Male | High SDI | 3.92 (3.60 to 4.09) |
| Female | High-middle SDI | 1.01 (0.90 to 1.12) |
| Male | High-middle SDI | 2.79 (2.59 to 3.00) |
| Female | Low SDI | 1.17 (0.74 to 1.81) |
| Male | Low SDI | 1.81 (0.88 to 3.18) |
| Female | Low-middle SDI | 0.96 (0.74 to 1.21) |
| Male | Low-middle SDI | 1.71 (1.38 to 2.54) |
| Female | Middle SDI | 0.78 (0.69 to 0.87) |
| Male | Middle SDI | 1.60 (1.44 to 1.80) |

### Data for Figure 3

#### Fig 3A. ASDR in Europe from 1990 to 2021

| **Year** | **Location** | **ASDR per 100,000** |
| --- | --- | --- |
| 1990 | Central Europe | 3.07 (2.95 to 3.18) |
| 1991 | Central Europe | 3.30 (3.16 to 3.41) |
| 1992 | Central Europe | 3.38 (3.23 to 3.49) |
| 1993 | Central Europe | 3.39 (3.23 to 3.50) |
| 1994 | Central Europe | 3.43 (3.27 to 3.54) |
| 1995 | Central Europe | 3.44 (3.29 to 3.54) |
| 1996 | Central Europe | 3.42 (3.28 to 3.52) |
| 1997 | Central Europe | 3.39 (3.25 to 3.49) |
| 1998 | Central Europe | 3.24 (3.11 to 3.34) |
| 1999 | Central Europe | 3.27 (3.14 to 3.37) |
| 2000 | Central Europe | 3.34 (3.19 to 3.43) |
| 2001 | Central Europe | 3.40 (3.25 to 3.50) |
| 2002 | Central Europe | 3.45 (3.30 to 3.55) |
| 2003 | Central Europe | 3.51 (3.37 to 3.62) |
| 2004 | Central Europe | 3.52 (3.39 to 3.62) |
| 2005 | Central Europe | 3.49 (3.35 to 3.60) |
| 2006 | Central Europe | 3.46 (3.31 to 3.56) |
| 2007 | Central Europe | 3.45 (3.30 to 3.55) |
| 2008 | Central Europe | 3.44 (3.30 to 3.54) |
| 2009 | Central Europe | 3.39 (3.24 to 3.49) |
| 2010 | Central Europe | 3.29 (3.15 to 3.40) |
| 2011 | Central Europe | 3.20 (3.05 to 3.31) |
| 2012 | Central Europe | 3.23 (3.07 to 3.35) |
| 2013 | Central Europe | 3.17 (3.01 to 3.28) |
| 2014 | Central Europe | 3.09 (2.93 to 3.20) |
| 2015 | Central Europe | 3.06 (2.90 to 3.17) |
| 2016 | Central Europe | 3.00 (2.85 to 3.11) |
| 2017 | Central Europe | 2.97 (2.81 to 3.08) |
| 2018 | Central Europe | 2.97 (2.81 to 3.09) |
| 2019 | Central Europe | 2.91 (2.75 to 3.03) |
| 2020 | Central Europe | 2.93 (2.75 to 3.14) |
| 2021 | Central Europe | 2.93 (2.69 to 3.21) |
| 1990 | Eastern Europe | 2.52 (2.43 to 2.62) |
| 1991 | Eastern Europe | 2.60 (2.51 to 2.70) |
| 1992 | Eastern Europe | 2.78 (2.69 to 2.88) |
| 1993 | Eastern Europe | 3.24 (3.14 to 3.34) |
| 1994 | Eastern Europe | 3.52 (3.41 to 3.63) |
| 1995 | Eastern Europe | 3.33 (3.23 to 3.45) |
| 1996 | Eastern Europe | 3.10 (3.01 to 3.20) |
| 1997 | Eastern Europe | 2.93 (2.84 to 3.02) |
| 1998 | Eastern Europe | 2.91 (2.82 to 3.00) |
| 1999 | Eastern Europe | 3.68 (3.56 to 3.81) |
| 2000 | Eastern Europe | 3.51 (3.40 to 3.64) |
| 2001 | Eastern Europe | 2.84 (2.76 to 2.94) |
| 2002 | Eastern Europe | 2.83 (2.74 to 2.93) |
| 2003 | Eastern Europe | 2.85 (2.75 to 2.94) |
| 2004 | Eastern Europe | 2.86 (2.77 to 2.95) |
| 2005 | Eastern Europe | 3.09 (2.99 to 3.20) |
| 2006 | Eastern Europe | 3.01 (2.91 to 3.10) |
| 2007 | Eastern Europe | 3.08 (2.98 to 3.18) |
| 2008 | Eastern Europe | 3.24 (3.14 to 3.34) |
| 2009 | Eastern Europe | 3.24 (3.13 to 3.33) |
| 2010 | Eastern Europe | 3.34 (3.23 to 3.42) |
| 2011 | Eastern Europe | 3.32 (3.21 to 3.40) |
| 2012 | Eastern Europe | 3.36 (3.24 to 3.44) |
| 2013 | Eastern Europe | 3.49 (3.36 to 3.56) |
| 2014 | Eastern Europe | 3.71 (3.57 to 3.80) |
| 2015 | Eastern Europe | 3.78 (3.62 to 3.86) |
| 2016 | Eastern Europe | 3.87 (3.71 to 3.96) |
| 2017 | Eastern Europe | 3.80 (3.63 to 3.90) |
| 2018 | Eastern Europe | 3.87 (3.69 to 4.00) |
| 2019 | Eastern Europe | 3.88 (3.70 to 4.04) |
| 2020 | Eastern Europe | 3.84 (3.64 to 4.04) |
| 2021 | Eastern Europe | 3.82 (3.52 to 4.12) |
| 1990 | Western Europe | 4.78 (4.50 to 4.92) |
| 1991 | Western Europe | 4.84 (4.54 to 4.98) |
| 1992 | Western Europe | 4.87 (4.58 to 5.02) |
| 1993 | Western Europe | 4.92 (4.61 to 5.07) |
| 1994 | Western Europe | 4.93 (4.62 to 5.08) |
| 1995 | Western Europe | 5.01 (4.70 to 5.16) |
| 1996 | Western Europe | 5.02 (4.72 to 5.18) |
| 1997 | Western Europe | 4.94 (4.64 to 5.09) |
| 1998 | Western Europe | 4.92 (4.62 to 5.08) |
| 1999 | Western Europe | 4.84 (4.54 to 4.99) |
| 2000 | Western Europe | 4.75 (4.44 to 4.91) |
| 2001 | Western Europe | 4.61 (4.29 to 4.77) |
| 2002 | Western Europe | 4.53 (4.21 to 4.69) |
| 2003 | Western Europe | 4.43 (4.11 to 4.59) |
| 2004 | Western Europe | 4.27 (3.95 to 4.44) |
| 2005 | Western Europe | 4.10 (3.78 to 4.25) |
| 2006 | Western Europe | 3.95 (3.64 to 4.10) |
| 2007 | Western Europe | 3.82 (3.52 to 3.97) |
| 2008 | Western Europe | 3.67 (3.36 to 3.81) |
| 2009 | Western Europe | 3.51 (3.21 to 3.65) |
| 2010 | Western Europe | 3.36 (3.07 to 3.50) |
| 2011 | Western Europe | 3.20 (2.92 to 3.34) |
| 2012 | Western Europe | 3.11 (2.84 to 3.25) |
| 2013 | Western Europe | 2.99 (2.72 to 3.12) |
| 2014 | Western Europe | 2.89 (2.63 to 3.03) |
| 2015 | Western Europe | 2.85 (2.58 to 2.98) |
| 2016 | Western Europe | 2.81 (2.54 to 2.94) |
| 2017 | Western Europe | 2.74 (2.48 to 2.87) |
| 2018 | Western Europe | 2.72 (2.44 to 2.84) |
| 2019 | Western Europe | 2.66 (2.40 to 2.79) |
| 2020 | Western Europe | 2.60 (2.33 to 2.74) |
| 2021 | Western Europe | 2.57 (2.30 to 2.71) |

#### Fig 3B. ASDR in Western Europe from 1990 to 2021

| **Year** | **Location** | **ASDR per 100,000** |
| --- | --- | --- |
| 1990 | Andorra | 6.87 (4.36 to 10.33) |
| 1991 | Andorra | 6.89 (4.36 to 10.07) |
| 1992 | Andorra | 6.90 (4.39 to 10.01) |
| 1993 | Andorra | 6.99 (4.39 to 10.20) |
| 1994 | Andorra | 6.95 (4.41 to 10.04) |
| 1995 | Andorra | 6.88 (4.42 to 9.87) |
| 1996 | Andorra | 6.82 (4.43 to 9.73) |
| 1997 | Andorra | 6.71 (4.42 to 9.50) |
| 1998 | Andorra | 6.62 (4.44 to 9.35) |
| 1999 | Andorra | 6.54 (4.42 to 9.26) |
| 2000 | Andorra | 6.47 (4.44 to 9.09) |
| 2001 | Andorra | 5.96 (3.98 to 8.45) |
| 2002 | Andorra | 5.57 (3.62 to 8.07) |
| 2003 | Andorra | 5.43 (3.56 to 7.75) |
| 2004 | Andorra | 5.28 (3.50 to 7.49) |
| 2005 | Andorra | 5.04 (3.36 to 7.21) |
| 2006 | Andorra | 4.93 (3.28 to 7.26) |
| 2007 | Andorra | 4.79 (3.21 to 7.17) |
| 2008 | Andorra | 5.00 (3.40 to 7.26) |
| 2009 | Andorra | 5.17 (3.61 to 7.42) |
| 2010 | Andorra | 5.29 (3.75 to 7.45) |
| 2011 | Andorra | 5.08 (3.69 to 7.01) |
| 2012 | Andorra | 4.90 (3.58 to 6.66) |
| 2013 | Andorra | 4.88 (3.54 to 6.60) |
| 2014 | Andorra | 4.88 (3.50 to 6.52) |
| 2015 | Andorra | 4.83 (3.45 to 6.45) |
| 2016 | Andorra | 4.82 (3.39 to 6.40) |
| 2017 | Andorra | 4.79 (3.34 to 6.50) |
| 2018 | Andorra | 4.76 (3.32 to 6.54) |
| 2019 | Andorra | 4.70 (3.22 to 6.62) |
| 2020 | Andorra | 3.96 (2.52 to 5.92) |
| 2021 | Andorra | 3.96 (2.50 to 5.97) |
| 1990 | Austria | 3.20 (3.03 to 3.37) |
| 1991 | Austria | 3.23 (3.03 to 3.41) |
| 1992 | Austria | 3.27 (3.06 to 3.46) |
| 1993 | Austria | 3.37 (3.14 to 3.56) |
| 1994 | Austria | 3.42 (3.18 to 3.63) |
| 1995 | Austria | 3.42 (3.17 to 3.62) |
| 1996 | Austria | 3.37 (3.11 to 3.56) |
| 1997 | Austria | 3.29 (3.02 to 3.48) |
| 1998 | Austria | 3.13 (2.86 to 3.32) |
| 1999 | Austria | 2.95 (2.69 to 3.14) |
| 2000 | Austria | 2.92 (2.67 to 3.11) |
| 2001 | Austria | 2.93 (2.67 to 3.11) |
| 2002 | Austria | 2.88 (2.63 to 3.05) |
| 2003 | Austria | 2.60 (2.37 to 2.75) |
| 2004 | Austria | 2.46 (2.24 to 2.60) |
| 2005 | Austria | 2.41 (2.20 to 2.56) |
| 2006 | Austria | 2.43 (2.20 to 2.58) |
| 2007 | Austria | 2.36 (2.14 to 2.51) |
| 2008 | Austria | 2.27 (2.07 to 2.42) |
| 2009 | Austria | 2.30 (2.09 to 2.46) |
| 2010 | Austria | 2.24 (2.04 to 2.39) |
| 2011 | Austria | 2.17 (1.97 to 2.31) |
| 2012 | Austria | 2.18 (1.98 to 2.32) |
| 2013 | Austria | 2.18 (2.00 to 2.32) |
| 2014 | Austria | 2.14 (1.95 to 2.28) |
| 2015 | Austria | 2.12 (1.94 to 2.26) |
| 2016 | Austria | 2.08 (1.89 to 2.22) |
| 2017 | Austria | 1.98 (1.80 to 2.12) |
| 2018 | Austria | 2.03 (1.82 to 2.16) |
| 2019 | Austria | 1.94 (1.74 to 2.07) |
| 2020 | Austria | 1.92 (1.72 to 2.06) |
| 2021 | Austria | 1.80 (1.61 to 1.94) |
| 1990 | Belgium | 5.11 (4.73 to 5.45) |
| 1991 | Belgium | 5.13 (4.74 to 5.48) |
| 1992 | Belgium | 5.24 (4.84 to 5.59) |
| 1993 | Belgium | 5.01 (4.62 to 5.34) |
| 1994 | Belgium | 4.98 (4.58 to 5.29) |
| 1995 | Belgium | 4.85 (4.47 to 5.15) |
| 1996 | Belgium | 4.97 (4.57 to 5.29) |
| 1997 | Belgium | 4.77 (4.37 to 5.06) |
| 1998 | Belgium | 4.59 (4.22 to 4.87) |
| 1999 | Belgium | 4.58 (4.22 to 4.86) |
| 2000 | Belgium | 4.57 (4.19 to 4.87) |
| 2001 | Belgium | 4.37 (4.00 to 4.66) |
| 2002 | Belgium | 4.28 (3.94 to 4.57) |
| 2003 | Belgium | 4.31 (3.93 to 4.59) |
| 2004 | Belgium | 4.19 (3.82 to 4.48) |
| 2005 | Belgium | 3.95 (3.59 to 4.24) |
| 2006 | Belgium | 3.69 (3.34 to 3.96) |
| 2007 | Belgium | 3.57 (3.22 to 3.83) |
| 2008 | Belgium | 3.56 (3.22 to 3.83) |
| 2009 | Belgium | 3.40 (3.07 to 3.65) |
| 2010 | Belgium | 3.23 (2.93 to 3.48) |
| 2011 | Belgium | 3.11 (2.81 to 3.34) |
| 2012 | Belgium | 2.97 (2.68 to 3.18) |
| 2013 | Belgium | 2.88 (2.59 to 3.08) |
| 2014 | Belgium | 2.67 (2.39 to 2.87) |
| 2015 | Belgium | 2.68 (2.41 to 2.89) |
| 2016 | Belgium | 2.59 (2.32 to 2.78) |
| 2017 | Belgium | 2.56 (2.28 to 2.75) |
| 2018 | Belgium | 2.43 (2.16 to 2.62) |
| 2019 | Belgium | 2.37 (2.10 to 2.56) |
| 2020 | Belgium | 2.26 (1.98 to 2.43) |
| 2021 | Belgium | 2.25 (1.96 to 2.43) |
| 1990 | Cyprus | 7.82 (6.01 to 10.06) |
| 1991 | Cyprus | 8.07 (6.33 to 10.25) |
| 1992 | Cyprus | 8.26 (6.56 to 10.34) |
| 1993 | Cyprus | 8.15 (6.61 to 10.19) |
| 1994 | Cyprus | 8.11 (6.51 to 10.29) |
| 1995 | Cyprus | 7.83 (6.40 to 9.65) |
| 1996 | Cyprus | 7.50 (6.02 to 9.13) |
| 1997 | Cyprus | 7.44 (5.92 to 9.06) |
| 1998 | Cyprus | 7.52 (6.10 to 9.09) |
| 1999 | Cyprus | 7.43 (6.17 to 8.88) |
| 2000 | Cyprus | 7.50 (6.31 to 8.87) |
| 2001 | Cyprus | 7.54 (6.25 to 8.92) |
| 2002 | Cyprus | 7.45 (6.19 to 8.88) |
| 2003 | Cyprus | 7.13 (6.00 to 8.37) |
| 2004 | Cyprus | 6.43 (5.48 to 7.43) |
| 2005 | Cyprus | 6.05 (5.23 to 6.94) |
| 2006 | Cyprus | 5.86 (5.05 to 6.69) |
| 2007 | Cyprus | 5.61 (4.83 to 6.42) |
| 2008 | Cyprus | 5.39 (4.68 to 6.12) |
| 2009 | Cyprus | 5.23 (4.56 to 5.93) |
| 2010 | Cyprus | 5.13 (4.50 to 5.83) |
| 2011 | Cyprus | 4.92 (4.29 to 5.56) |
| 2012 | Cyprus | 4.75 (4.11 to 5.39) |
| 2013 | Cyprus | 4.61 (3.96 to 5.24) |
| 2014 | Cyprus | 4.49 (3.83 to 5.12) |
| 2015 | Cyprus | 4.43 (3.78 to 5.09) |
| 2016 | Cyprus | 4.30 (3.64 to 4.96) |
| 2017 | Cyprus | 4.29 (3.62 to 4.96) |
| 2018 | Cyprus | 4.24 (3.52 to 4.97) |
| 2019 | Cyprus | 4.26 (3.46 to 5.10) |
| 2020 | Cyprus | 4.00 (3.11 to 4.95) |
| 2021 | Cyprus | 3.96 (3.04 to 4.88) |
| 1990 | Denmark | 6.71 (6.21 to 7.17) |
| 1991 | Denmark | 6.71 (6.22 to 7.14) |
| 1992 | Denmark | 6.87 (6.40 to 7.28) |
| 1993 | Denmark | 7.13 (6.65 to 7.55) |
| 1994 | Denmark | 7.62 (7.11 to 8.07) |
| 1995 | Denmark | 7.61 (7.12 to 8.06) |
| 1996 | Denmark | 7.65 (7.12 to 8.07) |
| 1997 | Denmark | 7.73 (7.21 to 8.17) |
| 1998 | Denmark | 7.77 (7.23 to 8.19) |
| 1999 | Denmark | 7.57 (7.07 to 8.01) |
| 2000 | Denmark | 7.48 (6.96 to 7.93) |
| 2001 | Denmark | 7.53 (6.97 to 7.99) |
| 2002 | Denmark | 7.43 (6.91 to 7.89) |
| 2003 | Denmark | 7.27 (6.69 to 7.75) |
| 2004 | Denmark | 7.36 (6.78 to 7.83) |
| 2005 | Denmark | 7.19 (6.60 to 7.69) |
| 2006 | Denmark | 7.02 (6.44 to 7.47) |
| 2007 | Denmark | 6.78 (6.20 to 7.20) |
| 2008 | Denmark | 6.38 (5.83 to 6.79) |
| 2009 | Denmark | 5.98 (5.45 to 6.38) |
| 2010 | Denmark | 5.70 (5.19 to 6.08) |
| 2011 | Denmark | 5.63 (5.12 to 6.02) |
| 2012 | Denmark | 5.55 (5.02 to 5.93) |
| 2013 | Denmark | 5.50 (4.96 to 5.88) |
| 2014 | Denmark | 5.12 (4.62 to 5.49) |
| 2015 | Denmark | 4.98 (4.46 to 5.34) |
| 2016 | Denmark | 4.90 (4.40 to 5.27) |
| 2017 | Denmark | 4.77 (4.28 to 5.15) |
| 2018 | Denmark | 4.84 (4.33 to 5.25) |
| 2019 | Denmark | 4.72 (4.21 to 5.14) |
| 2020 | Denmark | 4.63 (4.14 to 5.03) |
| 2021 | Denmark | 4.61 (4.10 to 5.02) |
| 1990 | Finland | 7.88 (7.35 to 8.36) |
| 1991 | Finland | 7.63 (7.08 to 8.08) |
| 1992 | Finland | 7.48 (6.98 to 7.93) |
| 1993 | Finland | 7.32 (6.76 to 7.77) |
| 1994 | Finland | 7.04 (6.48 to 7.42) |
| 1995 | Finland | 7.02 (6.45 to 7.39) |
| 1996 | Finland | 6.98 (6.38 to 7.41) |
| 1997 | Finland | 6.99 (6.40 to 7.44) |
| 1998 | Finland | 6.95 (6.38 to 7.41) |
| 1999 | Finland | 6.74 (6.17 to 7.17) |
| 2000 | Finland | 6.67 (6.09 to 7.07) |
| 2001 | Finland | 6.57 (6.02 to 6.97) |
| 2002 | Finland | 6.73 (6.17 to 7.15) |
| 2003 | Finland | 6.57 (6.01 to 6.99) |
| 2004 | Finland | 6.40 (5.84 to 6.80) |
| 2005 | Finland | 6.11 (5.59 to 6.50) |
| 2006 | Finland | 6.01 (5.45 to 6.38) |
| 2007 | Finland | 6.08 (5.49 to 6.45) |
| 2008 | Finland | 5.87 (5.25 to 6.23) |
| 2009 | Finland | 5.67 (5.07 to 6.03) |
| 2010 | Finland | 5.42 (4.85 to 5.79) |
| 2011 | Finland | 5.14 (4.58 to 5.51) |
| 2012 | Finland | 5.03 (4.45 to 5.39) |
| 2013 | Finland | 4.86 (4.29 to 5.24) |
| 2014 | Finland | 4.66 (4.10 to 5.03) |
| 2015 | Finland | 4.35 (3.84 to 4.68) |
| 2016 | Finland | 4.18 (3.71 to 4.51) |
| 2017 | Finland | 4.03 (3.59 to 4.35) |
| 2018 | Finland | 3.78 (3.37 to 4.07) |
| 2019 | Finland | 3.72 (3.31 to 4.01) |
| 2020 | Finland | 3.66 (3.26 to 3.95) |
| 2021 | Finland | 3.63 (3.20 to 3.93) |
| 1990 | France | 3.65 (3.41 to 3.85) |
| 1991 | France | 3.70 (3.44 to 3.91) |
| 1992 | France | 3.75 (3.49 to 3.97) |
| 1993 | France | 3.78 (3.51 to 4.00) |
| 1994 | France | 3.73 (3.44 to 3.96) |
| 1995 | France | 3.84 (3.53 to 4.07) |
| 1996 | France | 3.93 (3.61 to 4.18) |
| 1997 | France | 3.82 (3.50 to 4.06) |
| 1998 | France | 3.73 (3.43 to 3.95) |
| 1999 | France | 3.71 (3.42 to 3.93) |
| 2000 | France | 3.70 (3.42 to 3.93) |
| 2001 | France | 3.59 (3.30 to 3.81) |
| 2002 | France | 3.51 (3.23 to 3.74) |
| 2003 | France | 3.39 (3.11 to 3.60) |
| 2004 | France | 3.23 (2.96 to 3.42) |
| 2005 | France | 3.17 (2.90 to 3.35) |
| 2006 | France | 3.07 (2.79 to 3.25) |
| 2007 | France | 2.95 (2.68 to 3.12) |
| 2008 | France | 2.85 (2.60 to 3.03) |
| 2009 | France | 2.73 (2.49 to 2.90) |
| 2010 | France | 2.58 (2.35 to 2.75) |
| 2011 | France | 2.38 (2.17 to 2.55) |
| 2012 | France | 2.28 (2.08 to 2.44) |
| 2013 | France | 2.18 (1.98 to 2.33) |
| 2014 | France | 2.10 (1.89 to 2.24) |
| 2015 | France | 2.08 (1.88 to 2.22) |
| 2016 | France | 2.09 (1.88 to 2.23) |
| 2017 | France | 2.07 (1.87 to 2.21) |
| 2018 | France | 2.01 (1.81 to 2.16) |
| 2019 | France | 1.98 (1.76 to 2.13) |
| 2020 | France | 1.96 (1.73 to 2.11) |
| 2021 | France | 1.89 (1.67 to 2.04) |
| 1990 | Germany | 3.32 (3.07 to 3.56) |
| 1991 | Germany | 3.32 (3.06 to 3.55) |
| 1992 | Germany | 3.25 (3.00 to 3.47) |
| 1993 | Germany | 3.23 (2.98 to 3.44) |
| 1994 | Germany | 3.16 (2.93 to 3.37) |
| 1995 | Germany | 3.12 (2.89 to 3.32) |
| 1996 | Germany | 3.12 (2.88 to 3.32) |
| 1997 | Germany | 3.11 (2.87 to 3.32) |
| 1998 | Germany | 3.17 (2.91 to 3.39) |
| 1999 | Germany | 3.07 (2.82 to 3.29) |
| 2000 | Germany | 2.94 (2.70 to 3.15) |
| 2001 | Germany | 2.81 (2.57 to 3.00) |
| 2002 | Germany | 2.78 (2.55 to 2.96) |
| 2003 | Germany | 2.74 (2.50 to 2.92) |
| 2004 | Germany | 2.66 (2.41 to 2.84) |
| 2005 | Germany | 2.58 (2.33 to 2.77) |
| 2006 | Germany | 2.57 (2.32 to 2.75) |
| 2007 | Germany | 2.56 (2.31 to 2.74) |
| 2008 | Germany | 2.55 (2.32 to 2.73) |
| 2009 | Germany | 2.46 (2.23 to 2.62) |
| 2010 | Germany | 2.38 (2.15 to 2.54) |
| 2011 | Germany | 2.35 (2.13 to 2.50) |
| 2012 | Germany | 2.31 (2.10 to 2.46) |
| 2013 | Germany | 2.30 (2.09 to 2.46) |
| 2014 | Germany | 2.28 (2.07 to 2.44) |
| 2015 | Germany | 2.24 (2.04 to 2.38) |
| 2016 | Germany | 2.25 (2.04 to 2.40) |
| 2017 | Germany | 2.24 (2.03 to 2.39) |
| 2018 | Germany | 2.32 (2.09 to 2.47) |
| 2019 | Germany | 2.33 (2.10 to 2.49) |
| 2020 | Germany | 2.30 (2.06 to 2.45) |
| 2021 | Germany | 2.25 (2.02 to 2.41) |
| 1990 | Greece | 3.81 (3.55 to 4.03) |
| 1991 | Greece | 3.92 (3.66 to 4.14) |
| 1992 | Greece | 4.00 (3.75 to 4.23) |
| 1993 | Greece | 4.02 (3.78 to 4.25) |
| 1994 | Greece | 4.24 (3.96 to 4.47) |
| 1995 | Greece | 4.47 (4.20 to 4.72) |
| 1996 | Greece | 4.53 (4.25 to 4.76) |
| 1997 | Greece | 4.53 (4.27 to 4.76) |
| 1998 | Greece | 4.73 (4.46 to 4.97) |
| 1999 | Greece | 4.69 (4.37 to 4.92) |
| 2000 | Greece | 4.67 (4.35 to 4.91) |
| 2001 | Greece | 4.69 (4.36 to 4.94) |
| 2002 | Greece | 4.65 (4.34 to 4.88) |
| 2003 | Greece | 4.64 (4.34 to 4.88) |
| 2004 | Greece | 4.68 (4.35 to 4.92) |
| 2005 | Greece | 4.58 (4.26 to 4.82) |
| 2006 | Greece | 4.51 (4.19 to 4.75) |
| 2007 | Greece | 4.46 (4.13 to 4.71) |
| 2008 | Greece | 4.15 (3.85 to 4.40) |
| 2009 | Greece | 4.06 (3.76 to 4.31) |
| 2010 | Greece | 3.94 (3.64 to 4.19) |
| 2011 | Greece | 3.79 (3.50 to 4.04) |
| 2012 | Greece | 3.70 (3.41 to 3.96) |
| 2013 | Greece | 3.60 (3.32 to 3.86) |
| 2014 | Greece | 3.60 (3.32 to 3.83) |
| 2015 | Greece | 3.86 (3.56 to 4.11) |
| 2016 | Greece | 4.04 (3.72 to 4.31) |
| 2017 | Greece | 3.97 (3.65 to 4.21) |
| 2018 | Greece | 3.91 (3.59 to 4.15) |
| 2019 | Greece | 3.93 (3.62 to 4.17) |
| 2020 | Greece | 3.99 (3.67 to 4.25) |
| 2021 | Greece | 3.99 (3.65 to 4.26) |
| 1990 | Iceland | 4.34 (3.91 to 4.65) |
| 1991 | Iceland | 4.47 (4.01 to 4.80) |
| 1992 | Iceland | 4.60 (4.14 to 4.95) |
| 1993 | Iceland | 4.54 (4.08 to 4.88) |
| 1994 | Iceland | 4.91 (4.44 to 5.28) |
| 1995 | Iceland | 5.05 (4.57 to 5.45) |
| 1996 | Iceland | 5.12 (4.64 to 5.52) |
| 1997 | Iceland | 5.09 (4.60 to 5.48) |
| 1998 | Iceland | 4.98 (4.54 to 5.34) |
| 1999 | Iceland | 4.76 (4.34 to 5.12) |
| 2000 | Iceland | 4.61 (4.21 to 4.97) |
| 2001 | Iceland | 4.77 (4.32 to 5.12) |
| 2002 | Iceland | 4.69 (4.25 to 5.04) |
| 2003 | Iceland | 4.46 (4.05 to 4.81) |
| 2004 | Iceland | 4.43 (4.01 to 4.78) |
| 2005 | Iceland | 4.29 (3.89 to 4.63) |
| 2006 | Iceland | 4.21 (3.82 to 4.54) |
| 2007 | Iceland | 4.03 (3.66 to 4.34) |
| 2008 | Iceland | 3.90 (3.53 to 4.20) |
| 2009 | Iceland | 3.81 (3.43 to 4.11) |
| 2010 | Iceland | 3.76 (3.38 to 4.08) |
| 2011 | Iceland | 3.71 (3.31 to 4.03) |
| 2012 | Iceland | 3.51 (3.12 to 3.83) |
| 2013 | Iceland | 3.41 (3.03 to 3.73) |
| 2014 | Iceland | 3.33 (2.97 to 3.62) |
| 2015 | Iceland | 3.18 (2.83 to 3.45) |
| 2016 | Iceland | 3.11 (2.76 to 3.39) |
| 2017 | Iceland | 2.95 (2.60 to 3.23) |
| 2018 | Iceland | 2.86 (2.49 to 3.16) |
| 2019 | Iceland | 2.77 (2.37 to 3.08) |
| 2020 | Iceland | 2.61 (2.24 to 2.90) |
| 2021 | Iceland | 2.61 (2.23 to 2.92) |
| 1990 | Ireland | 6.36 (5.94 to 6.78) |
| 1991 | Ireland | 6.40 (5.96 to 6.79) |
| 1992 | Ireland | 6.86 (6.40 to 7.29) |
| 1993 | Ireland | 6.97 (6.50 to 7.39) |
| 1994 | Ireland | 6.74 (6.32 to 7.16) |
| 1995 | Ireland | 6.95 (6.47 to 7.37) |
| 1996 | Ireland | 6.90 (6.46 to 7.30) |
| 1997 | Ireland | 6.99 (6.54 to 7.40) |
| 1998 | Ireland | 6.93 (6.49 to 7.33) |
| 1999 | Ireland | 6.95 (6.46 to 7.35) |
| 2000 | Ireland | 6.90 (6.41 to 7.30) |
| 2001 | Ireland | 6.71 (6.20 to 7.12) |
| 2002 | Ireland | 6.57 (6.04 to 6.98) |
| 2003 | Ireland | 6.15 (5.64 to 6.52) |
| 2004 | Ireland | 5.81 (5.33 to 6.17) |
| 2005 | Ireland | 5.57 (5.11 to 5.91) |
| 2006 | Ireland | 5.44 (4.97 to 5.81) |
| 2007 | Ireland | 5.38 (4.93 to 5.74) |
| 2008 | Ireland | 5.00 (4.55 to 5.33) |
| 2009 | Ireland | 4.66 (4.23 to 4.98) |
| 2010 | Ireland | 4.42 (3.99 to 4.73) |
| 2011 | Ireland | 4.42 (3.99 to 4.74) |
| 2012 | Ireland | 4.27 (3.86 to 4.58) |
| 2013 | Ireland | 4.13 (3.73 to 4.44) |
| 2014 | Ireland | 3.85 (3.44 to 4.13) |
| 2015 | Ireland | 3.67 (3.30 to 3.97) |
| 2016 | Ireland | 3.69 (3.29 to 3.99) |
| 2017 | Ireland | 3.54 (3.14 to 3.84) |
| 2018 | Ireland | 3.50 (3.07 to 3.81) |
| 2019 | Ireland | 3.38 (2.93 to 3.69) |
| 2020 | Ireland | 3.16 (2.72 to 3.49) |
| 2021 | Ireland | 2.93 (2.47 to 3.25) |
| 1990 | Israel | 2.34 (2.15 to 2.50) |
| 1991 | Israel | 2.39 (2.20 to 2.55) |
| 1992 | Israel | 2.44 (2.26 to 2.60) |
| 1993 | Israel | 2.40 (2.22 to 2.54) |
| 1994 | Israel | 2.41 (2.23 to 2.55) |
| 1995 | Israel | 2.40 (2.22 to 2.54) |
| 1996 | Israel | 2.34 (2.15 to 2.48) |
| 1997 | Israel | 2.42 (2.22 to 2.56) |
| 1998 | Israel | 2.42 (2.22 to 2.56) |
| 1999 | Israel | 2.43 (2.23 to 2.58) |
| 2000 | Israel | 2.24 (2.05 to 2.39) |
| 2001 | Israel | 2.13 (1.95 to 2.26) |
| 2002 | Israel | 2.22 (2.02 to 2.36) |
| 2003 | Israel | 2.10 (1.91 to 2.23) |
| 2004 | Israel | 2.03 (1.85 to 2.15) |
| 2005 | Israel | 1.93 (1.76 to 2.05) |
| 2006 | Israel | 1.84 (1.67 to 1.96) |
| 2007 | Israel | 1.81 (1.64 to 1.93) |
| 2008 | Israel | 1.71 (1.54 to 1.83) |
| 2009 | Israel | 1.70 (1.53 to 1.83) |
| 2010 | Israel | 1.65 (1.48 to 1.78) |
| 2011 | Israel | 1.53 (1.38 to 1.66) |
| 2012 | Israel | 1.51 (1.35 to 1.62) |
| 2013 | Israel | 1.53 (1.37 to 1.64) |
| 2014 | Israel | 1.47 (1.31 to 1.58) |
| 2015 | Israel | 1.50 (1.34 to 1.61) |
| 2016 | Israel | 1.49 (1.34 to 1.60) |
| 2017 | Israel | 1.44 (1.28 to 1.55) |
| 2018 | Israel | 1.37 (1.22 to 1.47) |
| 2019 | Israel | 1.36 (1.21 to 1.47) |
| 2020 | Israel | 1.31 (1.17 to 1.42) |
| 2021 | Israel | 1.28 (1.13 to 1.40) |
| 1990 | Italy | 3.10 (2.91 to 3.21) |
| 1991 | Italy | 3.18 (2.98 to 3.30) |
| 1992 | Italy | 3.27 (3.06 to 3.39) |
| 1993 | Italy | 3.38 (3.15 to 3.52) |
| 1994 | Italy | 3.56 (3.32 to 3.70) |
| 1995 | Italy | 3.75 (3.49 to 3.90) |
| 1996 | Italy | 3.74 (3.48 to 3.90) |
| 1997 | Italy | 3.68 (3.43 to 3.84) |
| 1998 | Italy | 3.68 (3.43 to 3.83) |
| 1999 | Italy | 3.59 (3.34 to 3.74) |
| 2000 | Italy | 3.47 (3.21 to 3.62) |
| 2001 | Italy | 3.41 (3.14 to 3.56) |
| 2002 | Italy | 3.40 (3.13 to 3.55) |
| 2003 | Italy | 3.52 (3.23 to 3.68) |
| 2004 | Italy | 3.41 (3.11 to 3.59) |
| 2005 | Italy | 3.32 (3.02 to 3.49) |
| 2006 | Italy | 3.15 (2.84 to 3.31) |
| 2007 | Italy | 3.12 (2.81 to 3.27) |
| 2008 | Italy | 2.94 (2.66 to 3.09) |
| 2009 | Italy | 2.82 (2.55 to 2.96) |
| 2010 | Italy | 2.72 (2.46 to 2.86) |
| 2011 | Italy | 2.64 (2.38 to 2.77) |
| 2012 | Italy | 2.58 (2.33 to 2.71) |
| 2013 | Italy | 2.44 (2.20 to 2.56) |
| 2014 | Italy | 2.41 (2.16 to 2.54) |
| 2015 | Italy | 2.40 (2.16 to 2.53) |
| 2016 | Italy | 2.34 (2.11 to 2.48) |
| 2017 | Italy | 2.28 (2.06 to 2.41) |
| 2018 | Italy | 2.26 (2.02 to 2.39) |
| 2019 | Italy | 2.20 (1.96 to 2.34) |
| 2020 | Italy | 2.21 (1.98 to 2.36) |
| 2021 | Italy | 2.18 (1.95 to 2.34) |
| 1990 | Luxembourg | 4.33 (4.06 to 4.64) |
| 1991 | Luxembourg | 4.66 (4.37 to 4.98) |
| 1992 | Luxembourg | 4.74 (4.42 to 5.06) |
| 1993 | Luxembourg | 4.68 (4.39 to 5.01) |
| 1994 | Luxembourg | 4.77 (4.46 to 5.08) |
| 1995 | Luxembourg | 4.76 (4.44 to 5.07) |
| 1996 | Luxembourg | 4.77 (4.46 to 5.07) |
| 1997 | Luxembourg | 4.67 (4.37 to 4.98) |
| 1998 | Luxembourg | 4.57 (4.27 to 4.87) |
| 1999 | Luxembourg | 4.39 (4.07 to 4.70) |
| 2000 | Luxembourg | 4.24 (3.92 to 4.54) |
| 2001 | Luxembourg | 4.27 (3.96 to 4.56) |
| 2002 | Luxembourg | 4.15 (3.86 to 4.43) |
| 2003 | Luxembourg | 4.14 (3.84 to 4.41) |
| 2004 | Luxembourg | 3.91 (3.59 to 4.18) |
| 2005 | Luxembourg | 3.74 (3.43 to 4.01) |
| 2006 | Luxembourg | 3.72 (3.44 to 3.97) |
| 2007 | Luxembourg | 3.55 (3.26 to 3.77) |
| 2008 | Luxembourg | 3.35 (3.08 to 3.57) |
| 2009 | Luxembourg | 3.18 (2.92 to 3.40) |
| 2010 | Luxembourg | 2.96 (2.72 to 3.18) |
| 2011 | Luxembourg | 2.99 (2.74 to 3.22) |
| 2012 | Luxembourg | 2.90 (2.66 to 3.14) |
| 2013 | Luxembourg | 2.65 (2.42 to 2.86) |
| 2014 | Luxembourg | 2.65 (2.42 to 2.87) |
| 2015 | Luxembourg | 2.74 (2.50 to 2.97) |
| 2016 | Luxembourg | 2.60 (2.37 to 2.79) |
| 2017 | Luxembourg | 2.49 (2.26 to 2.68) |
| 2018 | Luxembourg | 2.49 (2.25 to 2.68) |
| 2019 | Luxembourg | 2.40 (2.16 to 2.61) |
| 2020 | Luxembourg | 2.27 (2.02 to 2.48) |
| 2021 | Luxembourg | 2.23 (1.97 to 2.46) |
| 1990 | Malta | 2.60 (2.41 to 2.80) |
| 1991 | Malta | 2.69 (2.50 to 2.89) |
| 1992 | Malta | 2.74 (2.55 to 2.94) |
| 1993 | Malta | 2.73 (2.52 to 2.92) |
| 1994 | Malta | 2.73 (2.52 to 2.94) |
| 1995 | Malta | 2.74 (2.53 to 2.95) |
| 1996 | Malta | 2.71 (2.51 to 2.92) |
| 1997 | Malta | 2.88 (2.66 to 3.10) |
| 1998 | Malta | 2.78 (2.58 to 3.00) |
| 1999 | Malta | 2.75 (2.55 to 2.98) |
| 2000 | Malta | 2.81 (2.60 to 3.05) |
| 2001 | Malta | 2.71 (2.50 to 2.94) |
| 2002 | Malta | 2.52 (2.31 to 2.72) |
| 2003 | Malta | 2.46 (2.26 to 2.67) |
| 2004 | Malta | 2.47 (2.27 to 2.67) |
| 2005 | Malta | 2.47 (2.26 to 2.68) |
| 2006 | Malta | 2.40 (2.19 to 2.59) |
| 2007 | Malta | 2.30 (2.10 to 2.48) |
| 2008 | Malta | 2.16 (1.98 to 2.34) |
| 2009 | Malta | 1.95 (1.77 to 2.11) |
| 2010 | Malta | 1.86 (1.69 to 2.01) |
| 2011 | Malta | 1.80 (1.64 to 1.95) |
| 2012 | Malta | 1.71 (1.57 to 1.86) |
| 2013 | Malta | 1.65 (1.51 to 1.79) |
| 2014 | Malta | 1.60 (1.45 to 1.73) |
| 2015 | Malta | 1.51 (1.37 to 1.63) |
| 2016 | Malta | 1.46 (1.32 to 1.59) |
| 2017 | Malta | 1.39 (1.25 to 1.53) |
| 2018 | Malta | 1.40 (1.26 to 1.54) |
| 2019 | Malta | 1.39 (1.23 to 1.54) |
| 2020 | Malta | 1.26 (1.11 to 1.40) |
| 2021 | Malta | 1.32 (1.17 to 1.47) |
| 1990 | Monaco | 5.66 (4.29 to 7.02) |
| 1991 | Monaco | 5.64 (4.26 to 7.03) |
| 1992 | Monaco | 5.62 (4.23 to 6.98) |
| 1993 | Monaco | 5.63 (4.20 to 7.03) |
| 1994 | Monaco | 5.67 (4.18 to 7.01) |
| 1995 | Monaco | 5.71 (4.22 to 7.06) |
| 1996 | Monaco | 5.76 (4.34 to 7.13) |
| 1997 | Monaco | 5.84 (4.40 to 7.15) |
| 1998 | Monaco | 5.92 (4.43 to 7.25) |
| 1999 | Monaco | 6.00 (4.51 to 7.32) |
| 2000 | Monaco | 6.10 (4.66 to 7.35) |
| 2001 | Monaco | 6.16 (4.79 to 7.47) |
| 2002 | Monaco | 6.15 (4.83 to 7.37) |
| 2003 | Monaco | 6.14 (4.90 to 7.35) |
| 2004 | Monaco | 6.10 (4.91 to 7.27) |
| 2005 | Monaco | 6.08 (4.93 to 7.19) |
| 2006 | Monaco | 6.04 (4.98 to 7.14) |
| 2007 | Monaco | 5.95 (4.94 to 7.02) |
| 2008 | Monaco | 5.87 (4.84 to 6.94) |
| 2009 | Monaco | 5.85 (4.89 to 6.96) |
| 2010 | Monaco | 5.85 (4.95 to 6.95) |
| 2011 | Monaco | 5.78 (4.84 to 6.86) |
| 2012 | Monaco | 5.74 (4.81 to 6.79) |
| 2013 | Monaco | 5.69 (4.73 to 6.73) |
| 2014 | Monaco | 5.62 (4.64 to 6.71) |
| 2015 | Monaco | 5.54 (4.53 to 6.73) |
| 2016 | Monaco | 5.46 (4.38 to 6.79) |
| 2017 | Monaco | 5.38 (4.31 to 6.80) |
| 2018 | Monaco | 5.31 (4.18 to 6.82) |
| 2019 | Monaco | 5.23 (4.04 to 6.84) |
| 2020 | Monaco | 5.17 (3.90 to 6.90) |
| 2021 | Monaco | 5.15 (3.88 to 6.95) |
| 1990 | Netherlands | 7.44 (6.89 to 7.96) |
| 1991 | Netherlands | 7.60 (7.04 to 8.10) |
| 1992 | Netherlands | 7.58 (7.00 to 8.07) |
| 1993 | Netherlands | 7.69 (7.08 to 8.19) |
| 1994 | Netherlands | 7.73 (7.09 to 8.20) |
| 1995 | Netherlands | 7.98 (7.33 to 8.45) |
| 1996 | Netherlands | 7.87 (7.26 to 8.36) |
| 1997 | Netherlands | 7.82 (7.20 to 8.32) |
| 1998 | Netherlands | 7.74 (7.12 to 8.22) |
| 1999 | Netherlands | 7.65 (7.02 to 8.13) |
| 2000 | Netherlands | 7.68 (7.05 to 8.19) |
| 2001 | Netherlands | 7.68 (7.04 to 8.18) |
| 2002 | Netherlands | 7.69 (7.02 to 8.18) |
| 2003 | Netherlands | 7.32 (6.65 to 7.80) |
| 2004 | Netherlands | 7.07 (6.42 to 7.54) |
| 2005 | Netherlands | 6.83 (6.19 to 7.30) |
| 2006 | Netherlands | 6.49 (5.90 to 6.95) |
| 2007 | Netherlands | 6.10 (5.54 to 6.53) |
| 2008 | Netherlands | 5.82 (5.27 to 6.24) |
| 2009 | Netherlands | 5.55 (5.00 to 5.96) |
| 2010 | Netherlands | 5.42 (4.86 to 5.80) |
| 2011 | Netherlands | 5.20 (4.66 to 5.57) |
| 2012 | Netherlands | 4.90 (4.38 to 5.27) |
| 2013 | Netherlands | 4.43 (3.94 to 4.79) |
| 2014 | Netherlands | 4.20 (3.71 to 4.54) |
| 2015 | Netherlands | 4.12 (3.66 to 4.47) |
| 2016 | Netherlands | 4.07 (3.60 to 4.41) |
| 2017 | Netherlands | 3.88 (3.42 to 4.21) |
| 2018 | Netherlands | 3.84 (3.37 to 4.17) |
| 2019 | Netherlands | 3.77 (3.30 to 4.10) |
| 2020 | Netherlands | 3.75 (3.26 to 4.08) |
| 2021 | Netherlands | 3.74 (3.24 to 4.08) |
| 1990 | Norway | 7.58 (7.16 to 7.90) |
| 1991 | Norway | 7.44 (6.99 to 7.72) |
| 1992 | Norway | 7.68 (7.20 to 7.99) |
| 1993 | Norway | 7.99 (7.49 to 8.31) |
| 1994 | Norway | 7.92 (7.40 to 8.25) |
| 1995 | Norway | 8.05 (7.49 to 8.39) |
| 1996 | Norway | 8.20 (7.59 to 8.55) |
| 1997 | Norway | 8.07 (7.46 to 8.44) |
| 1998 | Norway | 8.27 (7.71 to 8.65) |
| 1999 | Norway | 8.25 (7.65 to 8.65) |
| 2000 | Norway | 8.09 (7.47 to 8.51) |
| 2001 | Norway | 7.95 (7.31 to 8.36) |
| 2002 | Norway | 7.79 (7.19 to 8.19) |
| 2003 | Norway | 7.68 (7.05 to 8.06) |
| 2004 | Norway | 7.62 (6.97 to 8.03) |
| 2005 | Norway | 7.24 (6.62 to 7.61) |
| 2006 | Norway | 6.89 (6.27 to 7.24) |
| 2007 | Norway | 6.82 (6.18 to 7.15) |
| 2008 | Norway | 6.25 (5.66 to 6.58) |
| 2009 | Norway | 6.24 (5.63 to 6.55) |
| 2010 | Norway | 6.09 (5.51 to 6.43) |
| 2011 | Norway | 5.87 (5.33 to 6.20) |
| 2012 | Norway | 5.66 (5.13 to 5.97) |
| 2013 | Norway | 5.43 (4.89 to 5.75) |
| 2014 | Norway | 4.96 (4.43 to 5.25) |
| 2015 | Norway | 4.98 (4.45 to 5.27) |
| 2016 | Norway | 4.71 (4.22 to 4.98) |
| 2017 | Norway | 4.70 (4.19 to 4.98) |
| 2018 | Norway | 4.74 (4.24 to 5.03) |
| 2019 | Norway | 4.84 (4.29 to 5.13) |
| 2020 | Norway | 4.69 (4.07 to 5.00) |
| 2021 | Norway | 4.75 (4.15 to 5.08) |
| 1990 | Portugal | 1.75 (1.64 to 1.86) |
| 1991 | Portugal | 1.83 (1.72 to 1.94) |
| 1992 | Portugal | 1.80 (1.69 to 1.91) |
| 1993 | Portugal | 1.86 (1.75 to 1.96) |
| 1994 | Portugal | 1.81 (1.70 to 1.92) |
| 1995 | Portugal | 1.95 (1.82 to 2.05) |
| 1996 | Portugal | 1.95 (1.83 to 2.06) |
| 1997 | Portugal | 1.95 (1.83 to 2.06) |
| 1998 | Portugal | 1.94 (1.82 to 2.05) |
| 1999 | Portugal | 1.85 (1.74 to 1.96) |
| 2000 | Portugal | 1.89 (1.76 to 2.00) |
| 2001 | Portugal | 1.89 (1.75 to 2.00) |
| 2002 | Portugal | 1.82 (1.68 to 1.93) |
| 2003 | Portugal | 1.81 (1.68 to 1.92) |
| 2004 | Portugal | 1.71 (1.58 to 1.82) |
| 2005 | Portugal | 1.50 (1.39 to 1.61) |
| 2006 | Portugal | 1.52 (1.42 to 1.62) |
| 2007 | Portugal | 1.61 (1.50 to 1.72) |
| 2008 | Portugal | 1.66 (1.54 to 1.77) |
| 2009 | Portugal | 1.66 (1.54 to 1.77) |
| 2010 | Portugal | 1.62 (1.49 to 1.73) |
| 2011 | Portugal | 1.57 (1.45 to 1.67) |
| 2012 | Portugal | 1.58 (1.46 to 1.68) |
| 2013 | Portugal | 1.50 (1.37 to 1.59) |
| 2014 | Portugal | 1.49 (1.37 to 1.59) |
| 2015 | Portugal | 1.53 (1.41 to 1.63) |
| 2016 | Portugal | 1.52 (1.40 to 1.62) |
| 2017 | Portugal | 1.54 (1.41 to 1.64) |
| 2018 | Portugal | 1.58 (1.43 to 1.68) |
| 2019 | Portugal | 1.56 (1.42 to 1.67) |
| 2020 | Portugal | 1.50 (1.37 to 1.62) |
| 2021 | Portugal | 1.51 (1.37 to 1.62) |
| 1990 | San Marino | 3.09 (2.49 to 3.76) |
| 1991 | San Marino | 3.09 (2.50 to 3.71) |
| 1992 | San Marino | 3.09 (2.53 to 3.72) |
| 1993 | San Marino | 3.09 (2.55 to 3.74) |
| 1994 | San Marino | 3.10 (2.55 to 3.73) |
| 1995 | San Marino | 3.07 (2.52 to 3.62) |
| 1996 | San Marino | 3.05 (2.50 to 3.56) |
| 1997 | San Marino | 3.00 (2.47 to 3.53) |
| 1998 | San Marino | 2.98 (2.49 to 3.49) |
| 1999 | San Marino | 2.94 (2.45 to 3.45) |
| 2000 | San Marino | 2.91 (2.41 to 3.51) |
| 2001 | San Marino | 2.88 (2.37 to 3.50) |
| 2002 | San Marino | 2.84 (2.33 to 3.47) |
| 2003 | San Marino | 2.80 (2.26 to 3.47) |
| 2004 | San Marino | 2.73 (2.16 to 3.43) |
| 2005 | San Marino | 2.72 (2.12 to 3.40) |
| 2006 | San Marino | 2.73 (2.14 to 3.43) |
| 2007 | San Marino | 2.72 (2.08 to 3.48) |
| 2008 | San Marino | 2.73 (2.07 to 3.52) |
| 2009 | San Marino | 2.73 (2.03 to 3.60) |
| 2010 | San Marino | 2.75 (2.05 to 3.62) |
| 2011 | San Marino | 2.77 (2.07 to 3.68) |
| 2012 | San Marino | 2.78 (2.05 to 3.78) |
| 2013 | San Marino | 2.80 (2.09 to 3.77) |
| 2014 | San Marino | 2.81 (2.06 to 3.82) |
| 2015 | San Marino | 2.83 (2.07 to 3.80) |
| 2016 | San Marino | 2.84 (2.06 to 3.82) |
| 2017 | San Marino | 2.86 (2.06 to 3.77) |
| 2018 | San Marino | 2.85 (2.08 to 3.81) |
| 2019 | San Marino | 2.84 (2.06 to 3.85) |
| 2020 | San Marino | 1.77 (1.19 to 2.59) |
| 2021 | San Marino | 1.67 (1.06 to 2.59) |
| 1990 | Spain | 2.54 (2.37 to 2.68) |
| 1991 | Spain | 2.76 (2.57 to 2.92) |
| 1992 | Spain | 2.78 (2.59 to 2.95) |
| 1993 | Spain | 2.86 (2.67 to 3.02) |
| 1994 | Spain | 2.97 (2.76 to 3.14) |
| 1995 | Spain | 3.08 (2.88 to 3.26) |
| 1996 | Spain | 3.10 (2.91 to 3.26) |
| 1997 | Spain | 3.08 (2.88 to 3.23) |
| 1998 | Spain | 3.11 (2.91 to 3.27) |
| 1999 | Spain | 3.15 (2.95 to 3.32) |
| 2000 | Spain | 3.14 (2.95 to 3.32) |
| 2001 | Spain | 3.07 (2.87 to 3.24) |
| 2002 | Spain | 3.08 (2.88 to 3.26) |
| 2003 | Spain | 3.04 (2.85 to 3.21) |
| 2004 | Spain | 2.89 (2.69 to 3.05) |
| 2005 | Spain | 2.78 (2.59 to 2.95) |
| 2006 | Spain | 2.73 (2.54 to 2.89) |
| 2007 | Spain | 2.65 (2.46 to 2.81) |
| 2008 | Spain | 2.59 (2.39 to 2.75) |
| 2009 | Spain | 2.51 (2.33 to 2.67) |
| 2010 | Spain | 2.40 (2.20 to 2.55) |
| 2011 | Spain | 2.30 (2.09 to 2.45) |
| 2012 | Spain | 2.26 (2.04 to 2.40) |
| 2013 | Spain | 2.18 (1.98 to 2.32) |
| 2014 | Spain | 2.17 (1.96 to 2.31) |
| 2015 | Spain | 2.19 (1.99 to 2.33) |
| 2016 | Spain | 2.19 (2.00 to 2.34) |
| 2017 | Spain | 2.22 (2.04 to 2.37) |
| 2018 | Spain | 2.22 (2.02 to 2.37) |
| 2019 | Spain | 2.17 (1.97 to 2.32) |
| 2020 | Spain | 2.09 (1.89 to 2.25) |
| 2021 | Spain | 2.08 (1.87 to 2.23) |
| 1990 | Sweden | 7.41 (6.91 to 7.79) |
| 1991 | Sweden | 7.56 (7.07 to 7.94) |
| 1992 | Sweden | 7.58 (7.07 to 7.97) |
| 1993 | Sweden | 7.77 (7.27 to 8.17) |
| 1994 | Sweden | 7.71 (7.19 to 8.13) |
| 1995 | Sweden | 7.66 (7.15 to 8.08) |
| 1996 | Sweden | 7.85 (7.34 to 8.26) |
| 1997 | Sweden | 7.90 (7.39 to 8.33) |
| 1998 | Sweden | 7.97 (7.44 to 8.41) |
| 1999 | Sweden | 7.87 (7.31 to 8.31) |
| 2000 | Sweden | 7.71 (7.17 to 8.14) |
| 2001 | Sweden | 7.64 (7.05 to 8.06) |
| 2002 | Sweden | 7.38 (6.82 to 7.82) |
| 2003 | Sweden | 7.25 (6.68 to 7.66) |
| 2004 | Sweden | 7.24 (6.68 to 7.65) |
| 2005 | Sweden | 6.77 (6.22 to 7.15) |
| 2006 | Sweden | 6.55 (5.98 to 6.92) |
| 2007 | Sweden | 5.90 (5.38 to 6.24) |
| 2008 | Sweden | 5.63 (5.12 to 5.96) |
| 2009 | Sweden | 5.54 (5.03 to 5.87) |
| 2010 | Sweden | 5.30 (4.85 to 5.64) |
| 2011 | Sweden | 5.10 (4.68 to 5.42) |
| 2012 | Sweden | 4.87 (4.46 to 5.18) |
| 2013 | Sweden | 4.77 (4.36 to 5.08) |
| 2014 | Sweden | 4.48 (4.05 to 4.78) |
| 2015 | Sweden | 4.38 (3.94 to 4.69) |
| 2016 | Sweden | 4.40 (3.96 to 4.71) |
| 2017 | Sweden | 4.42 (3.99 to 4.72) |
| 2018 | Sweden | 4.32 (3.88 to 4.63) |
| 2019 | Sweden | 4.16 (3.72 to 4.47) |
| 2020 | Sweden | 4.02 (3.58 to 4.34) |
| 2021 | Sweden | 3.88 (3.38 to 4.33) |
| 1990 | Switzerland | 4.87 (4.43 to 5.26) |
| 1991 | Switzerland | 4.82 (4.40 to 5.17) |
| 1992 | Switzerland | 4.69 (4.26 to 5.04) |
| 1993 | Switzerland | 4.54 (4.14 to 4.87) |
| 1994 | Switzerland | 4.50 (4.12 to 4.81) |
| 1995 | Switzerland | 4.67 (4.25 to 5.00) |
| 1996 | Switzerland | 4.64 (4.22 to 4.96) |
| 1997 | Switzerland | 4.70 (4.30 to 5.00) |
| 1998 | Switzerland | 4.55 (4.17 to 4.84) |
| 1999 | Switzerland | 4.30 (3.92 to 4.57) |
| 2000 | Switzerland | 4.37 (3.98 to 4.66) |
| 2001 | Switzerland | 4.29 (3.90 to 4.56) |
| 2002 | Switzerland | 4.19 (3.80 to 4.47) |
| 2003 | Switzerland | 3.94 (3.58 to 4.22) |
| 2004 | Switzerland | 3.71 (3.35 to 3.99) |
| 2005 | Switzerland | 3.78 (3.40 to 4.09) |
| 2006 | Switzerland | 3.69 (3.29 to 3.99) |
| 2007 | Switzerland | 3.60 (3.22 to 3.87) |
| 2008 | Switzerland | 3.48 (3.12 to 3.75) |
| 2009 | Switzerland | 3.37 (3.00 to 3.63) |
| 2010 | Switzerland | 3.26 (2.90 to 3.50) |
| 2011 | Switzerland | 3.08 (2.73 to 3.32) |
| 2012 | Switzerland | 3.04 (2.69 to 3.28) |
| 2013 | Switzerland | 3.01 (2.66 to 3.25) |
| 2014 | Switzerland | 2.92 (2.58 to 3.16) |
| 2015 | Switzerland | 2.75 (2.43 to 2.97) |
| 2016 | Switzerland | 2.72 (2.39 to 2.94) |
| 2017 | Switzerland | 2.57 (2.26 to 2.77) |
| 2018 | Switzerland | 2.53 (2.21 to 2.74) |
| 2019 | Switzerland | 2.40 (2.09 to 2.61) |
| 2020 | Switzerland | 2.36 (2.04 to 2.59) |
| 2021 | Switzerland | 2.28 (1.96 to 2.51) |
| 1990 | United Kingdom of Great Britain and Northern Ireland | 9.72 (9.16 to 10.00) |
| 1991 | United Kingdom of Great Britain and Northern Ireland | 9.84 (9.31 to 10.12) |
| 1992 | United Kingdom of Great Britain and Northern Ireland | 10.04 (9.47 to 10.34) |
| 1993 | United Kingdom of Great Britain and Northern Ireland | 10.19 (9.65 to 10.47) |
| 1994 | United Kingdom of Great Britain and Northern Ireland | 10.17 (9.57 to 10.48) |
| 1995 | United Kingdom of Great Britain and Northern Ireland | 10.36 (9.78 to 10.66) |
| 1996 | United Kingdom of Great Britain and Northern Ireland | 10.42 (9.86 to 10.73) |
| 1997 | United Kingdom of Great Britain and Northern Ireland | 10.18 (9.61 to 10.49) |
| 1998 | United Kingdom of Great Britain and Northern Ireland | 10.17 (9.57 to 10.49) |
| 1999 | United Kingdom of Great Britain and Northern Ireland | 10.04 (9.45 to 10.35) |
| 2000 | United Kingdom of Great Britain and Northern Ireland | 9.89 (9.27 to 10.21) |
| 2001 | United Kingdom of Great Britain and Northern Ireland | 9.56 (8.90 to 9.89) |
| 2002 | United Kingdom of Great Britain and Northern Ireland | 9.33 (8.71 to 9.65) |
| 2003 | United Kingdom of Great Britain and Northern Ireland | 9.06 (8.42 to 9.37) |
| 2004 | United Kingdom of Great Britain and Northern Ireland | 8.67 (8.02 to 9.01) |
| 2005 | United Kingdom of Great Britain and Northern Ireland | 8.19 (7.56 to 8.49) |
| 2006 | United Kingdom of Great Britain and Northern Ireland | 7.83 (7.21 to 8.14) |
| 2007 | United Kingdom of Great Britain and Northern Ireland | 7.48 (6.88 to 7.78) |
| 2008 | United Kingdom of Great Britain and Northern Ireland | 7.06 (6.48 to 7.34) |
| 2009 | United Kingdom of Great Britain and Northern Ireland | 6.62 (6.03 to 6.91) |
| 2010 | United Kingdom of Great Britain and Northern Ireland | 6.26 (5.72 to 6.53) |
| 2011 | United Kingdom of Great Britain and Northern Ireland | 5.83 (5.29 to 6.10) |
| 2012 | United Kingdom of Great Britain and Northern Ireland | 5.64 (5.11 to 5.90) |
| 2013 | United Kingdom of Great Britain and Northern Ireland | 5.38 (4.88 to 5.62) |
| 2014 | United Kingdom of Great Britain and Northern Ireland | 5.15 (4.65 to 5.39) |
| 2015 | United Kingdom of Great Britain and Northern Ireland | 4.94 (4.45 to 5.17) |
| 2016 | United Kingdom of Great Britain and Northern Ireland | 4.76 (4.29 to 4.98) |
| 2017 | United Kingdom of Great Britain and Northern Ireland | 4.55 (4.11 to 4.76) |
| 2018 | United Kingdom of Great Britain and Northern Ireland | 4.38 (3.94 to 4.58) |
| 2019 | United Kingdom of Great Britain and Northern Ireland | 4.19 (3.76 to 4.38) |
| 2020 | United Kingdom of Great Britain and Northern Ireland | 4.02 (3.60 to 4.23) |
| 2021 | United Kingdom of Great Britain and Northern Ireland | 4.03 (3.61 to 4.25) |

#### Fig 3C. ASDR in Central Europe from 1990 to 2021

| **Year** | **Location** | **ASDR per 100,000** |
| --- | --- | --- |
| 1990 | Albania | 0.91 (0.76 to 1.08) |
| 1991 | Albania | 0.94 (0.78 to 1.11) |
| 1992 | Albania | 0.91 (0.75 to 1.08) |
| 1993 | Albania | 0.88 (0.73 to 1.06) |
| 1994 | Albania | 0.84 (0.68 to 1.01) |
| 1995 | Albania | 0.87 (0.71 to 1.03) |
| 1996 | Albania | 0.90 (0.76 to 1.05) |
| 1997 | Albania | 0.92 (0.78 to 1.06) |
| 1998 | Albania | 0.92 (0.78 to 1.08) |
| 1999 | Albania | 0.94 (0.79 to 1.10) |
| 2000 | Albania | 0.96 (0.82 to 1.13) |
| 2001 | Albania | 0.97 (0.80 to 1.16) |
| 2002 | Albania | 1.00 (0.83 to 1.20) |
| 2003 | Albania | 1.03 (0.87 to 1.24) |
| 2004 | Albania | 1.05 (0.87 to 1.25) |
| 2005 | Albania | 1.03 (0.86 to 1.23) |
| 2006 | Albania | 1.00 (0.82 to 1.22) |
| 2007 | Albania | 0.98 (0.77 to 1.22) |
| 2008 | Albania | 0.97 (0.76 to 1.21) |
| 2009 | Albania | 0.96 (0.75 to 1.21) |
| 2010 | Albania | 0.98 (0.75 to 1.28) |
| 2011 | Albania | 1.00 (0.75 to 1.31) |
| 2012 | Albania | 1.02 (0.77 to 1.33) |
| 2013 | Albania | 1.04 (0.78 to 1.36) |
| 2014 | Albania | 1.05 (0.79 to 1.42) |
| 2015 | Albania | 1.06 (0.78 to 1.42) |
| 2016 | Albania | 1.07 (0.79 to 1.45) |
| 2017 | Albania | 1.09 (0.79 to 1.46) |
| 2018 | Albania | 1.10 (0.80 to 1.49) |
| 2019 | Albania | 1.14 (0.80 to 1.58) |
| 2020 | Albania | 1.06 (0.76 to 1.43) |
| 2021 | Albania | 1.09 (0.76 to 1.50) |
| 1990 | Bosnia and Herzegovina | 2.12 (1.55 to 2.89) |
| 1991 | Bosnia and Herzegovina | 2.22 (1.59 to 3.00) |
| 1992 | Bosnia and Herzegovina | 2.21 (1.58 to 3.11) |
| 1993 | Bosnia and Herzegovina | 2.24 (1.59 to 3.15) |
| 1994 | Bosnia and Herzegovina | 2.24 (1.54 to 3.16) |
| 1995 | Bosnia and Herzegovina | 2.23 (1.49 to 3.21) |
| 1996 | Bosnia and Herzegovina | 2.24 (1.56 to 3.08) |
| 1997 | Bosnia and Herzegovina | 2.29 (1.63 to 3.14) |
| 1998 | Bosnia and Herzegovina | 2.28 (1.70 to 3.09) |
| 1999 | Bosnia and Herzegovina | 2.29 (1.73 to 3.08) |
| 2000 | Bosnia and Herzegovina | 2.34 (1.74 to 3.15) |
| 2001 | Bosnia and Herzegovina | 2.37 (1.74 to 3.16) |
| 2002 | Bosnia and Herzegovina | 2.44 (1.80 to 3.23) |
| 2003 | Bosnia and Herzegovina | 2.51 (1.93 to 3.23) |
| 2004 | Bosnia and Herzegovina | 2.55 (1.98 to 3.23) |
| 2005 | Bosnia and Herzegovina | 2.68 (2.08 to 3.47) |
| 2006 | Bosnia and Herzegovina | 2.73 (2.14 to 3.50) |
| 2007 | Bosnia and Herzegovina | 2.91 (2.29 to 3.69) |
| 2008 | Bosnia and Herzegovina | 2.86 (2.26 to 3.60) |
| 2009 | Bosnia and Herzegovina | 2.87 (2.29 to 3.58) |
| 2010 | Bosnia and Herzegovina | 2.84 (2.27 to 3.52) |
| 2011 | Bosnia and Herzegovina | 2.87 (2.34 to 3.49) |
| 2012 | Bosnia and Herzegovina | 2.83 (2.32 to 3.45) |
| 2013 | Bosnia and Herzegovina | 2.88 (2.37 to 3.53) |
| 2014 | Bosnia and Herzegovina | 2.90 (2.37 to 3.56) |
| 2015 | Bosnia and Herzegovina | 2.94 (2.38 to 3.65) |
| 2016 | Bosnia and Herzegovina | 2.99 (2.39 to 3.76) |
| 2017 | Bosnia and Herzegovina | 2.95 (2.34 to 3.71) |
| 2018 | Bosnia and Herzegovina | 2.98 (2.33 to 3.76) |
| 2019 | Bosnia and Herzegovina | 3.06 (2.38 to 3.94) |
| 2020 | Bosnia and Herzegovina | 3.16 (2.42 to 4.09) |
| 2021 | Bosnia and Herzegovina | 3.00 (2.15 to 3.99) |
| 1990 | Bulgaria | 2.03 (1.82 to 2.23) |
| 1991 | Bulgaria | 1.94 (1.76 to 2.13) |
| 1992 | Bulgaria | 2.00 (1.80 to 2.19) |
| 1993 | Bulgaria | 2.18 (1.95 to 2.40) |
| 1994 | Bulgaria | 2.31 (2.08 to 2.57) |
| 1995 | Bulgaria | 2.37 (2.14 to 2.62) |
| 1996 | Bulgaria | 2.39 (2.14 to 2.65) |
| 1997 | Bulgaria | 2.34 (2.08 to 2.63) |
| 1998 | Bulgaria | 2.22 (1.99 to 2.50) |
| 1999 | Bulgaria | 2.24 (1.99 to 2.52) |
| 2000 | Bulgaria | 2.48 (2.22 to 2.78) |
| 2001 | Bulgaria | 2.58 (2.29 to 2.89) |
| 2002 | Bulgaria | 2.54 (2.27 to 2.85) |
| 2003 | Bulgaria | 2.67 (2.39 to 3.02) |
| 2004 | Bulgaria | 2.66 (2.38 to 2.98) |
| 2005 | Bulgaria | 2.88 (2.56 to 3.22) |
| 2006 | Bulgaria | 2.88 (2.59 to 3.24) |
| 2007 | Bulgaria | 2.82 (2.53 to 3.17) |
| 2008 | Bulgaria | 2.76 (2.46 to 3.07) |
| 2009 | Bulgaria | 2.69 (2.41 to 2.98) |
| 2010 | Bulgaria | 2.57 (2.31 to 2.86) |
| 2011 | Bulgaria | 2.60 (2.33 to 2.88) |
| 2012 | Bulgaria | 2.61 (2.32 to 2.92) |
| 2013 | Bulgaria | 2.58 (2.30 to 2.89) |
| 2014 | Bulgaria | 2.56 (2.30 to 2.90) |
| 2015 | Bulgaria | 2.57 (2.27 to 2.94) |
| 2016 | Bulgaria | 2.58 (2.26 to 2.97) |
| 2017 | Bulgaria | 2.61 (2.27 to 3.01) |
| 2018 | Bulgaria | 2.57 (2.23 to 2.99) |
| 2019 | Bulgaria | 2.58 (2.21 to 3.05) |
| 2020 | Bulgaria | 2.59 (2.15 to 3.13) |
| 2021 | Bulgaria | 2.62 (2.12 to 3.23) |
| 1990 | Croatia | 2.79 (2.45 to 3.19) |
| 1991 | Croatia | 3.02 (2.67 to 3.48) |
| 1992 | Croatia | 3.06 (2.71 to 3.51) |
| 1993 | Croatia | 3.11 (2.74 to 3.54) |
| 1994 | Croatia | 3.15 (2.74 to 3.59) |
| 1995 | Croatia | 3.24 (2.80 to 3.73) |
| 1996 | Croatia | 3.24 (2.74 to 3.74) |
| 1997 | Croatia | 3.21 (2.73 to 3.73) |
| 1998 | Croatia | 3.21 (2.73 to 3.72) |
| 1999 | Croatia | 3.34 (2.86 to 3.87) |
| 2000 | Croatia | 3.47 (2.99 to 3.97) |
| 2001 | Croatia | 3.61 (3.14 to 4.12) |
| 2002 | Croatia | 3.67 (3.23 to 4.22) |
| 2003 | Croatia | 3.68 (3.24 to 4.20) |
| 2004 | Croatia | 3.50 (3.06 to 3.98) |
| 2005 | Croatia | 3.57 (3.11 to 4.09) |
| 2006 | Croatia | 3.42 (2.95 to 3.92) |
| 2007 | Croatia | 3.54 (3.03 to 4.08) |
| 2008 | Croatia | 3.79 (3.26 to 4.38) |
| 2009 | Croatia | 3.82 (3.29 to 4.42) |
| 2010 | Croatia | 3.92 (3.37 to 4.56) |
| 2011 | Croatia | 3.62 (3.13 to 4.19) |
| 2012 | Croatia | 3.78 (3.26 to 4.34) |
| 2013 | Croatia | 3.76 (3.26 to 4.33) |
| 2014 | Croatia | 3.61 (3.12 to 4.14) |
| 2015 | Croatia | 3.67 (3.19 to 4.19) |
| 2016 | Croatia | 3.44 (2.99 to 3.92) |
| 2017 | Croatia | 3.42 (2.99 to 3.85) |
| 2018 | Croatia | 3.42 (2.97 to 3.86) |
| 2019 | Croatia | 3.34 (2.91 to 3.78) |
| 2020 | Croatia | 3.31 (2.87 to 3.76) |
| 2021 | Croatia | 3.34 (2.87 to 3.86) |
| 1990 | Czech Republic | 2.96 (2.76 to 3.18) |
| 1991 | Czech Republic | 3.06 (2.86 to 3.28) |
| 1992 | Czech Republic | 3.08 (2.87 to 3.30) |
| 1993 | Czech Republic | 3.14 (2.90 to 3.38) |
| 1994 | Czech Republic | 3.24 (2.97 to 3.47) |
| 1995 | Czech Republic | 3.19 (2.91 to 3.44) |
| 1996 | Czech Republic | 3.18 (2.89 to 3.43) |
| 1997 | Czech Republic | 3.22 (2.92 to 3.48) |
| 1998 | Czech Republic | 3.16 (2.86 to 3.44) |
| 1999 | Czech Republic | 3.34 (3.04 to 3.62) |
| 2000 | Czech Republic | 3.78 (3.46 to 4.11) |
| 2001 | Czech Republic | 4.03 (3.69 to 4.35) |
| 2002 | Czech Republic | 4.00 (3.68 to 4.30) |
| 2003 | Czech Republic | 4.08 (3.74 to 4.38) |
| 2004 | Czech Republic | 4.03 (3.70 to 4.30) |
| 2005 | Czech Republic | 3.85 (3.55 to 4.12) |
| 2006 | Czech Republic | 3.73 (3.44 to 3.99) |
| 2007 | Czech Republic | 3.61 (3.34 to 3.87) |
| 2008 | Czech Republic | 3.55 (3.28 to 3.82) |
| 2009 | Czech Republic | 3.57 (3.29 to 3.84) |
| 2010 | Czech Republic | 3.67 (3.38 to 3.96) |
| 2011 | Czech Republic | 3.64 (3.35 to 3.93) |
| 2012 | Czech Republic | 3.47 (3.19 to 3.74) |
| 2013 | Czech Republic | 3.31 (3.04 to 3.59) |
| 2014 | Czech Republic | 3.24 (2.99 to 3.49) |
| 2015 | Czech Republic | 3.16 (2.91 to 3.42) |
| 2016 | Czech Republic | 3.00 (2.77 to 3.26) |
| 2017 | Czech Republic | 2.86 (2.62 to 3.09) |
| 2018 | Czech Republic | 2.90 (2.64 to 3.12) |
| 2019 | Czech Republic | 2.87 (2.62 to 3.08) |
| 2020 | Czech Republic | 2.86 (2.57 to 3.11) |
| 2021 | Czech Republic | 2.83 (2.46 to 3.19) |
| 1990 | Hungary | 3.23 (3.00 to 3.48) |
| 1991 | Hungary | 3.42 (3.16 to 3.67) |
| 1992 | Hungary | 3.68 (3.40 to 3.94) |
| 1993 | Hungary | 3.70 (3.42 to 3.97) |
| 1994 | Hungary | 3.76 (3.47 to 4.04) |
| 1995 | Hungary | 3.73 (3.45 to 3.99) |
| 1996 | Hungary | 3.64 (3.35 to 3.91) |
| 1997 | Hungary | 3.49 (3.21 to 3.78) |
| 1998 | Hungary | 3.48 (3.17 to 3.77) |
| 1999 | Hungary | 3.55 (3.22 to 3.86) |
| 2000 | Hungary | 3.38 (3.06 to 3.69) |
| 2001 | Hungary | 3.34 (3.00 to 3.65) |
| 2002 | Hungary | 3.38 (3.04 to 3.70) |
| 2003 | Hungary | 3.43 (3.11 to 3.74) |
| 2004 | Hungary | 3.44 (3.13 to 3.72) |
| 2005 | Hungary | 3.55 (3.26 to 3.82) |
| 2006 | Hungary | 3.41 (3.15 to 3.65) |
| 2007 | Hungary | 3.52 (3.26 to 3.75) |
| 2008 | Hungary | 3.31 (3.07 to 3.54) |
| 2009 | Hungary | 3.31 (3.08 to 3.54) |
| 2010 | Hungary | 3.29 (3.05 to 3.52) |
| 2011 | Hungary | 3.19 (2.97 to 3.42) |
| 2012 | Hungary | 3.20 (2.99 to 3.43) |
| 2013 | Hungary | 3.13 (2.91 to 3.36) |
| 2014 | Hungary | 3.05 (2.85 to 3.27) |
| 2015 | Hungary | 3.03 (2.82 to 3.25) |
| 2016 | Hungary | 2.90 (2.69 to 3.09) |
| 2017 | Hungary | 2.97 (2.75 to 3.19) |
| 2018 | Hungary | 3.11 (2.88 to 3.31) |
| 2019 | Hungary | 2.96 (2.74 to 3.17) |
| 2020 | Hungary | 2.94 (2.67 to 3.21) |
| 2021 | Hungary | 2.87 (2.54 to 3.23) |
| 1990 | Montenegro | 6.70 (5.35 to 8.64) |
| 1991 | Montenegro | 6.77 (5.29 to 8.87) |
| 1992 | Montenegro | 6.72 (5.26 to 8.72) |
| 1993 | Montenegro | 6.63 (5.26 to 8.46) |
| 1994 | Montenegro | 6.61 (5.27 to 8.35) |
| 1995 | Montenegro | 6.50 (5.27 to 8.08) |
| 1996 | Montenegro | 6.41 (5.25 to 7.86) |
| 1997 | Montenegro | 6.50 (5.37 to 7.82) |
| 1998 | Montenegro | 6.66 (5.59 to 7.97) |
| 1999 | Montenegro | 6.72 (5.75 to 8.06) |
| 2000 | Montenegro | 6.66 (5.71 to 8.01) |
| 2001 | Montenegro | 6.77 (5.73 to 8.13) |
| 2002 | Montenegro | 6.87 (5.84 to 8.20) |
| 2003 | Montenegro | 7.04 (6.02 to 8.35) |
| 2004 | Montenegro | 6.99 (5.96 to 8.31) |
| 2005 | Montenegro | 6.91 (5.92 to 8.17) |
| 2006 | Montenegro | 6.99 (6.05 to 8.20) |
| 2007 | Montenegro | 6.80 (5.83 to 8.01) |
| 2008 | Montenegro | 6.84 (5.75 to 8.10) |
| 2009 | Montenegro | 7.27 (5.98 to 8.64) |
| 2010 | Montenegro | 7.62 (6.17 to 9.18) |
| 2011 | Montenegro | 7.71 (6.30 to 9.40) |
| 2012 | Montenegro | 7.79 (6.49 to 9.48) |
| 2013 | Montenegro | 7.97 (6.59 to 9.80) |
| 2014 | Montenegro | 8.09 (6.64 to 9.90) |
| 2015 | Montenegro | 8.35 (6.81 to 10.30) |
| 2016 | Montenegro | 8.61 (7.03 to 10.78) |
| 2017 | Montenegro | 8.77 (7.08 to 11.05) |
| 2018 | Montenegro | 8.76 (6.95 to 10.97) |
| 2019 | Montenegro | 8.85 (6.95 to 11.21) |
| 2020 | Montenegro | 9.31 (7.28 to 11.88) |
| 2021 | Montenegro | 8.65 (6.59 to 11.28) |
| 1990 | North Macedonia | 1.94 (1.51 to 2.42) |
| 1991 | North Macedonia | 1.96 (1.51 to 2.43) |
| 1992 | North Macedonia | 2.06 (1.63 to 2.50) |
| 1993 | North Macedonia | 2.12 (1.68 to 2.55) |
| 1994 | North Macedonia | 2.16 (1.73 to 2.57) |
| 1995 | North Macedonia | 2.23 (1.84 to 2.64) |
| 1996 | North Macedonia | 2.31 (1.94 to 2.74) |
| 1997 | North Macedonia | 2.37 (1.98 to 2.81) |
| 1998 | North Macedonia | 2.49 (2.11 to 2.95) |
| 1999 | North Macedonia | 2.59 (2.20 to 3.04) |
| 2000 | North Macedonia | 2.56 (2.16 to 3.05) |
| 2001 | North Macedonia | 2.62 (2.21 to 3.12) |
| 2002 | North Macedonia | 2.64 (2.21 to 3.16) |
| 2003 | North Macedonia | 2.58 (2.18 to 3.10) |
| 2004 | North Macedonia | 2.51 (2.11 to 2.96) |
| 2005 | North Macedonia | 2.55 (2.14 to 3.02) |
| 2006 | North Macedonia | 2.69 (2.26 to 3.20) |
| 2007 | North Macedonia | 2.73 (2.34 to 3.22) |
| 2008 | North Macedonia | 2.67 (2.22 to 3.13) |
| 2009 | North Macedonia | 2.68 (2.22 to 3.18) |
| 2010 | North Macedonia | 2.64 (2.17 to 3.12) |
| 2011 | North Macedonia | 2.55 (2.11 to 3.00) |
| 2012 | North Macedonia | 2.56 (2.12 to 3.04) |
| 2013 | North Macedonia | 2.53 (2.08 to 2.99) |
| 2014 | North Macedonia | 2.54 (2.10 to 3.05) |
| 2015 | North Macedonia | 2.49 (2.03 to 2.97) |
| 2016 | North Macedonia | 2.55 (2.07 to 3.07) |
| 2017 | North Macedonia | 2.46 (2.02 to 2.98) |
| 2018 | North Macedonia | 2.54 (2.06 to 3.13) |
| 2019 | North Macedonia | 2.64 (2.12 to 3.27) |
| 2020 | North Macedonia | 2.68 (1.86 to 3.67) |
| 2021 | North Macedonia | 2.63 (1.80 to 3.81) |
| 1990 | Poland | 4.54 (4.36 to 4.66) |
| 1991 | Poland | 5.14 (4.95 to 5.29) |
| 1992 | Poland | 5.25 (5.05 to 5.40) |
| 1993 | Poland | 5.19 (4.98 to 5.34) |
| 1994 | Poland | 5.21 (5.00 to 5.35) |
| 1995 | Poland | 5.21 (5.02 to 5.36) |
| 1996 | Poland | 5.16 (4.95 to 5.31) |
| 1997 | Poland | 5.11 (4.92 to 5.26) |
| 1998 | Poland | 4.74 (4.54 to 4.88) |
| 1999 | Poland | 4.71 (4.52 to 4.86) |
| 2000 | Poland | 4.71 (4.49 to 4.84) |
| 2001 | Poland | 4.73 (4.50 to 4.87) |
| 2002 | Poland | 4.81 (4.58 to 4.96) |
| 2003 | Poland | 4.85 (4.61 to 4.99) |
| 2004 | Poland | 4.89 (4.66 to 5.03) |
| 2005 | Poland | 4.75 (4.52 to 4.89) |
| 2006 | Poland | 4.76 (4.55 to 4.90) |
| 2007 | Poland | 4.72 (4.50 to 4.86) |
| 2008 | Poland | 4.74 (4.52 to 4.88) |
| 2009 | Poland | 4.55 (4.33 to 4.68) |
| 2010 | Poland | 4.26 (4.05 to 4.40) |
| 2011 | Poland | 4.11 (3.89 to 4.25) |
| 2012 | Poland | 4.17 (3.95 to 4.32) |
| 2013 | Poland | 4.11 (3.88 to 4.25) |
| 2014 | Poland | 3.90 (3.68 to 4.05) |
| 2015 | Poland | 3.83 (3.62 to 3.97) |
| 2016 | Poland | 3.72 (3.52 to 3.86) |
| 2017 | Poland | 3.62 (3.42 to 3.75) |
| 2018 | Poland | 3.55 (3.36 to 3.69) |
| 2019 | Poland | 3.41 (3.22 to 3.53) |
| 2020 | Poland | 3.42 (3.17 to 3.66) |
| 2021 | Poland | 3.44 (3.09 to 3.79) |
| 1990 | Romania | 1.40 (1.30 to 1.52) |
| 1991 | Romania | 1.42 (1.32 to 1.53) |
| 1992 | Romania | 1.49 (1.39 to 1.60) |
| 1993 | Romania | 1.51 (1.41 to 1.62) |
| 1994 | Romania | 1.54 (1.44 to 1.64) |
| 1995 | Romania | 1.59 (1.49 to 1.69) |
| 1996 | Romania | 1.64 (1.55 to 1.75) |
| 1997 | Romania | 1.62 (1.53 to 1.72) |
| 1998 | Romania | 1.56 (1.48 to 1.65) |
| 1999 | Romania | 1.55 (1.46 to 1.63) |
| 2000 | Romania | 1.55 (1.47 to 1.64) |
| 2001 | Romania | 1.61 (1.52 to 1.70) |
| 2002 | Romania | 1.68 (1.59 to 1.77) |
| 2003 | Romania | 1.75 (1.66 to 1.85) |
| 2004 | Romania | 1.78 (1.67 to 1.88) |
| 2005 | Romania | 1.77 (1.67 to 1.86) |
| 2006 | Romania | 1.70 (1.60 to 1.80) |
| 2007 | Romania | 1.66 (1.56 to 1.76) |
| 2008 | Romania | 1.71 (1.61 to 1.82) |
| 2009 | Romania | 1.78 (1.67 to 1.89) |
| 2010 | Romania | 1.77 (1.67 to 1.89) |
| 2011 | Romania | 1.64 (1.54 to 1.74) |
| 2012 | Romania | 1.72 (1.62 to 1.83) |
| 2013 | Romania | 1.63 (1.53 to 1.73) |
| 2014 | Romania | 1.69 (1.59 to 1.80) |
| 2015 | Romania | 1.63 (1.52 to 1.74) |
| 2016 | Romania | 1.73 (1.61 to 1.84) |
| 2017 | Romania | 1.78 (1.65 to 1.91) |
| 2018 | Romania | 1.87 (1.72 to 2.01) |
| 2019 | Romania | 1.86 (1.69 to 2.02) |
| 2020 | Romania | 1.85 (1.66 to 2.06) |
| 2021 | Romania | 1.90 (1.65 to 2.18) |
| 1990 | Serbia | 4.22 (3.50 to 5.14) |
| 1991 | Serbia | 4.21 (3.41 to 5.21) |
| 1992 | Serbia | 4.15 (3.34 to 5.24) |
| 1993 | Serbia | 4.15 (3.35 to 5.12) |
| 1994 | Serbia | 4.18 (3.51 to 4.97) |
| 1995 | Serbia | 4.23 (3.69 to 4.88) |
| 1996 | Serbia | 4.22 (3.70 to 4.81) |
| 1997 | Serbia | 4.17 (3.60 to 4.83) |
| 1998 | Serbia | 3.94 (3.48 to 4.45) |
| 1999 | Serbia | 3.96 (3.51 to 4.44) |
| 2000 | Serbia | 4.07 (3.64 to 4.58) |
| 2001 | Serbia | 4.03 (3.60 to 4.51) |
| 2002 | Serbia | 4.03 (3.59 to 4.46) |
| 2003 | Serbia | 4.11 (3.67 to 4.62) |
| 2004 | Serbia | 4.17 (3.69 to 4.70) |
| 2005 | Serbia | 4.19 (3.71 to 4.77) |
| 2006 | Serbia | 4.20 (3.66 to 4.75) |
| 2007 | Serbia | 4.25 (3.68 to 4.80) |
| 2008 | Serbia | 4.26 (3.71 to 4.82) |
| 2009 | Serbia | 4.20 (3.67 to 4.81) |
| 2010 | Serbia | 4.14 (3.62 to 4.77) |
| 2011 | Serbia | 4.14 (3.57 to 4.74) |
| 2012 | Serbia | 4.14 (3.58 to 4.67) |
| 2013 | Serbia | 4.07 (3.54 to 4.59) |
| 2014 | Serbia | 4.07 (3.55 to 4.60) |
| 2015 | Serbia | 4.12 (3.62 to 4.71) |
| 2016 | Serbia | 4.06 (3.56 to 4.67) |
| 2017 | Serbia | 4.08 (3.53 to 4.72) |
| 2018 | Serbia | 3.98 (3.36 to 4.64) |
| 2019 | Serbia | 4.01 (3.37 to 4.68) |
| 2020 | Serbia | 4.18 (3.39 to 5.09) |
| 2021 | Serbia | 4.05 (3.21 to 5.10) |
| 1990 | Slovakia | 2.29 (1.97 to 2.68) |
| 1991 | Slovakia | 2.24 (1.94 to 2.60) |
| 1992 | Slovakia | 2.23 (1.97 to 2.56) |
| 1993 | Slovakia | 2.21 (1.98 to 2.52) |
| 1994 | Slovakia | 2.21 (1.99 to 2.51) |
| 1995 | Slovakia | 2.22 (2.00 to 2.51) |
| 1996 | Slovakia | 2.21 (2.00 to 2.46) |
| 1997 | Slovakia | 2.28 (2.06 to 2.53) |
| 1998 | Slovakia | 2.32 (2.12 to 2.58) |
| 1999 | Slovakia | 2.34 (2.14 to 2.58) |
| 2000 | Slovakia | 2.34 (2.14 to 2.58) |
| 2001 | Slovakia | 2.37 (2.13 to 2.61) |
| 2002 | Slovakia | 2.38 (2.14 to 2.62) |
| 2003 | Slovakia | 2.38 (2.13 to 2.62) |
| 2004 | Slovakia | 2.36 (2.10 to 2.61) |
| 2005 | Slovakia | 2.42 (2.17 to 2.69) |
| 2006 | Slovakia | 2.41 (2.16 to 2.69) |
| 2007 | Slovakia | 2.43 (2.18 to 2.71) |
| 2008 | Slovakia | 2.39 (2.12 to 2.65) |
| 2009 | Slovakia | 2.35 (2.11 to 2.61) |
| 2010 | Slovakia | 2.30 (2.06 to 2.54) |
| 2011 | Slovakia | 2.31 (2.08 to 2.55) |
| 2012 | Slovakia | 2.30 (2.04 to 2.53) |
| 2013 | Slovakia | 2.26 (2.03 to 2.52) |
| 2014 | Slovakia | 2.26 (2.01 to 2.54) |
| 2015 | Slovakia | 2.28 (2.04 to 2.57) |
| 2016 | Slovakia | 2.22 (1.95 to 2.54) |
| 2017 | Slovakia | 2.24 (1.94 to 2.55) |
| 2018 | Slovakia | 2.26 (1.96 to 2.60) |
| 2019 | Slovakia | 2.21 (1.87 to 2.61) |
| 2020 | Slovakia | 2.27 (1.84 to 2.78) |
| 2021 | Slovakia | 2.29 (1.79 to 2.90) |
| 1990 | Slovenia | 2.66 (2.47 to 2.84) |
| 1991 | Slovenia | 2.94 (2.72 to 3.13) |
| 1992 | Slovenia | 3.02 (2.81 to 3.22) |
| 1993 | Slovenia | 3.15 (2.91 to 3.36) |
| 1994 | Slovenia | 3.07 (2.86 to 3.28) |
| 1995 | Slovenia | 2.96 (2.76 to 3.17) |
| 1996 | Slovenia | 2.99 (2.77 to 3.21) |
| 1997 | Slovenia | 3.05 (2.81 to 3.29) |
| 1998 | Slovenia | 3.13 (2.87 to 3.39) |
| 1999 | Slovenia | 3.21 (2.92 to 3.49) |
| 2000 | Slovenia | 3.23 (2.95 to 3.51) |
| 2001 | Slovenia | 3.38 (3.10 to 3.67) |
| 2002 | Slovenia | 3.40 (3.12 to 3.68) |
| 2003 | Slovenia | 3.46 (3.17 to 3.76) |
| 2004 | Slovenia | 3.27 (2.98 to 3.54) |
| 2005 | Slovenia | 3.14 (2.85 to 3.40) |
| 2006 | Slovenia | 3.11 (2.86 to 3.38) |
| 2007 | Slovenia | 2.97 (2.71 to 3.23) |
| 2008 | Slovenia | 2.93 (2.68 to 3.15) |
| 2009 | Slovenia | 2.82 (2.59 to 3.04) |
| 2010 | Slovenia | 2.84 (2.60 to 3.07) |
| 2011 | Slovenia | 2.73 (2.48 to 2.95) |
| 2012 | Slovenia | 2.84 (2.58 to 3.06) |
| 2013 | Slovenia | 2.71 (2.44 to 2.92) |
| 2014 | Slovenia | 2.56 (2.29 to 2.78) |
| 2015 | Slovenia | 2.64 (2.40 to 2.86) |
| 2016 | Slovenia | 2.58 (2.33 to 2.82) |
| 2017 | Slovenia | 2.50 (2.25 to 2.71) |
| 2018 | Slovenia | 2.43 (2.17 to 2.66) |
| 2019 | Slovenia | 2.25 (2.01 to 2.47) |
| 2020 | Slovenia | 2.22 (1.94 to 2.58) |
| 2021 | Slovenia | 2.16 (1.82 to 2.58) |

#### Fig 3D. ASDR in Eastern Europe from 1990 to 2021

| **Year** | **Location** | **ASDR per 100,000** |
| --- | --- | --- |
| 1990 | Belarus | 2.67 (2.31 to 3.23) |
| 1991 | Belarus | 2.75 (2.40 to 3.33) |
| 1992 | Belarus | 2.88 (2.52 to 3.43) |
| 1993 | Belarus | 3.19 (2.80 to 3.78) |
| 1994 | Belarus | 3.25 (2.85 to 3.84) |
| 1995 | Belarus | 3.42 (3.00 to 4.04) |
| 1996 | Belarus | 3.40 (2.97 to 3.99) |
| 1997 | Belarus | 3.43 (3.01 to 4.00) |
| 1998 | Belarus | 3.49 (3.04 to 4.09) |
| 1999 | Belarus | 3.58 (3.12 to 4.19) |
| 2000 | Belarus | 3.42 (3.00 to 3.96) |
| 2001 | Belarus | 3.64 (3.22 to 4.24) |
| 2002 | Belarus | 4.04 (3.55 to 4.69) |
| 2003 | Belarus | 4.00 (3.53 to 4.65) |
| 2004 | Belarus | 3.99 (3.50 to 4.59) |
| 2005 | Belarus | 4.17 (3.67 to 4.80) |
| 2006 | Belarus | 4.08 (3.63 to 4.68) |
| 2007 | Belarus | 3.91 (3.49 to 4.44) |
| 2008 | Belarus | 3.98 (3.55 to 4.48) |
| 2009 | Belarus | 4.04 (3.61 to 4.53) |
| 2010 | Belarus | 4.12 (3.71 to 4.64) |
| 2011 | Belarus | 4.23 (3.82 to 4.69) |
| 2012 | Belarus | 3.73 (3.38 to 4.11) |
| 2013 | Belarus | 3.70 (3.39 to 4.02) |
| 2014 | Belarus | 3.65 (3.36 to 3.96) |
| 2015 | Belarus | 3.54 (3.27 to 3.83) |
| 2016 | Belarus | 3.55 (3.28 to 3.83) |
| 2017 | Belarus | 3.54 (3.28 to 3.82) |
| 2018 | Belarus | 3.66 (3.38 to 3.98) |
| 2019 | Belarus | 3.74 (3.45 to 4.07) |
| 2020 | Belarus | 3.76 (3.27 to 4.32) |
| 2021 | Belarus | 3.75 (3.05 to 4.49) |
| 1990 | Estonia | 2.86 (2.60 to 3.13) |
| 1991 | Estonia | 3.01 (2.78 to 3.29) |
| 1992 | Estonia | 3.08 (2.84 to 3.31) |
| 1993 | Estonia | 3.39 (3.13 to 3.66) |
| 1994 | Estonia | 3.81 (3.54 to 4.09) |
| 1995 | Estonia | 3.65 (3.40 to 3.93) |
| 1996 | Estonia | 3.28 (3.06 to 3.53) |
| 1997 | Estonia | 3.43 (3.19 to 3.68) |
| 1998 | Estonia | 3.53 (3.28 to 3.78) |
| 1999 | Estonia | 4.04 (3.73 to 4.32) |
| 2000 | Estonia | 3.73 (3.47 to 3.99) |
| 2001 | Estonia | 3.22 (2.99 to 3.43) |
| 2002 | Estonia | 3.06 (2.86 to 3.27) |
| 2003 | Estonia | 2.87 (2.66 to 3.07) |
| 2004 | Estonia | 2.84 (2.63 to 3.06) |
| 2005 | Estonia | 2.79 (2.58 to 3.00) |
| 2006 | Estonia | 2.90 (2.68 to 3.10) |
| 2007 | Estonia | 3.17 (2.92 to 3.43) |
| 2008 | Estonia | 2.93 (2.72 to 3.17) |
| 2009 | Estonia | 3.05 (2.79 to 3.31) |
| 2010 | Estonia | 3.06 (2.79 to 3.33) |
| 2011 | Estonia | 3.13 (2.89 to 3.41) |
| 2012 | Estonia | 3.22 (2.93 to 3.48) |
| 2013 | Estonia | 3.23 (2.95 to 3.51) |
| 2014 | Estonia | 3.47 (3.18 to 3.77) |
| 2015 | Estonia | 3.31 (3.02 to 3.61) |
| 2016 | Estonia | 3.43 (3.13 to 3.72) |
| 2017 | Estonia | 3.37 (3.07 to 3.67) |
| 2018 | Estonia | 3.51 (3.19 to 3.86) |
| 2019 | Estonia | 3.40 (3.07 to 3.71) |
| 2020 | Estonia | 3.36 (3.03 to 3.71) |
| 2021 | Estonia | 3.34 (2.90 to 3.78) |
| 1990 | Latvia | 2.21 (2.03 to 2.40) |
| 1991 | Latvia | 2.27 (2.10 to 2.47) |
| 1992 | Latvia | 2.38 (2.20 to 2.56) |
| 1993 | Latvia | 2.76 (2.57 to 2.96) |
| 1994 | Latvia | 3.08 (2.86 to 3.31) |
| 1995 | Latvia | 2.93 (2.74 to 3.12) |
| 1996 | Latvia | 2.61 (2.44 to 2.78) |
| 1997 | Latvia | 2.57 (2.42 to 2.74) |
| 1998 | Latvia | 2.72 (2.55 to 2.90) |
| 1999 | Latvia | 3.01 (2.81 to 3.19) |
| 2000 | Latvia | 2.75 (2.57 to 2.91) |
| 2001 | Latvia | 2.44 (2.30 to 2.59) |
| 2002 | Latvia | 2.39 (2.23 to 2.54) |
| 2003 | Latvia | 2.39 (2.23 to 2.55) |
| 2004 | Latvia | 2.39 (2.24 to 2.55) |
| 2005 | Latvia | 2.55 (2.40 to 2.72) |
| 2006 | Latvia | 2.65 (2.50 to 2.81) |
| 2007 | Latvia | 2.73 (2.57 to 2.89) |
| 2008 | Latvia | 2.45 (2.31 to 2.59) |
| 2009 | Latvia | 2.47 (2.32 to 2.61) |
| 2010 | Latvia | 2.51 (2.35 to 2.66) |
| 2011 | Latvia | 2.50 (2.35 to 2.66) |
| 2012 | Latvia | 2.60 (2.43 to 2.75) |
| 2013 | Latvia | 2.73 (2.55 to 2.90) |
| 2014 | Latvia | 2.74 (2.56 to 2.92) |
| 2015 | Latvia | 2.77 (2.59 to 2.97) |
| 2016 | Latvia | 2.91 (2.70 to 3.10) |
| 2017 | Latvia | 3.15 (2.91 to 3.40) |
| 2018 | Latvia | 3.17 (2.92 to 3.42) |
| 2019 | Latvia | 3.02 (2.75 to 3.28) |
| 2020 | Latvia | 3.07 (2.76 to 3.44) |
| 2021 | Latvia | 3.05 (2.63 to 3.47) |
| 1990 | Lithuania | 2.16 (1.97 to 2.36) |
| 1991 | Lithuania | 2.27 (2.10 to 2.47) |
| 1992 | Lithuania | 2.30 (2.13 to 2.48) |
| 1993 | Lithuania | 2.63 (2.45 to 2.82) |
| 1994 | Lithuania | 2.78 (2.59 to 2.97) |
| 1995 | Lithuania | 2.63 (2.45 to 2.80) |
| 1996 | Lithuania | 2.53 (2.36 to 2.71) |
| 1997 | Lithuania | 2.42 (2.27 to 2.58) |
| 1998 | Lithuania | 2.51 (2.35 to 2.67) |
| 1999 | Lithuania | 3.01 (2.83 to 3.18) |
| 2000 | Lithuania | 2.73 (2.57 to 2.88) |
| 2001 | Lithuania | 2.56 (2.40 to 2.69) |
| 2002 | Lithuania | 2.49 (2.35 to 2.62) |
| 2003 | Lithuania | 2.43 (2.30 to 2.56) |
| 2004 | Lithuania | 2.43 (2.30 to 2.57) |
| 2005 | Lithuania | 2.72 (2.56 to 2.87) |
| 2006 | Lithuania | 2.71 (2.55 to 2.88) |
| 2007 | Lithuania | 2.92 (2.73 to 3.11) |
| 2008 | Lithuania | 2.89 (2.71 to 3.08) |
| 2009 | Lithuania | 2.74 (2.56 to 2.93) |
| 2010 | Lithuania | 2.83 (2.64 to 3.04) |
| 2011 | Lithuania | 2.95 (2.76 to 3.18) |
| 2012 | Lithuania | 2.86 (2.67 to 3.08) |
| 2013 | Lithuania | 3.00 (2.79 to 3.23) |
| 2014 | Lithuania | 3.06 (2.83 to 3.30) |
| 2015 | Lithuania | 3.24 (3.01 to 3.47) |
| 2016 | Lithuania | 3.30 (3.07 to 3.54) |
| 2017 | Lithuania | 3.25 (3.02 to 3.50) |
| 2018 | Lithuania | 3.12 (2.88 to 3.36) |
| 2019 | Lithuania | 3.14 (2.90 to 3.37) |
| 2020 | Lithuania | 3.18 (2.84 to 3.50) |
| 2021 | Lithuania | 3.15 (2.73 to 3.56) |
| 1990 | Republic of Moldova | 0.96 (0.89 to 1.06) |
| 1991 | Republic of Moldova | 1.05 (0.96 to 1.14) |
| 1992 | Republic of Moldova | 1.02 (0.94 to 1.10) |
| 1993 | Republic of Moldova | 1.05 (0.98 to 1.13) |
| 1994 | Republic of Moldova | 1.18 (1.10 to 1.27) |
| 1995 | Republic of Moldova | 1.21 (1.13 to 1.30) |
| 1996 | Republic of Moldova | 1.15 (1.07 to 1.23) |
| 1997 | Republic of Moldova | 1.08 (1.00 to 1.15) |
| 1998 | Republic of Moldova | 1.07 (0.99 to 1.14) |
| 1999 | Republic of Moldova | 1.26 (1.17 to 1.34) |
| 2000 | Republic of Moldova | 1.14 (1.07 to 1.21) |
| 2001 | Republic of Moldova | 0.92 (0.85 to 0.97) |
| 2002 | Republic of Moldova | 0.93 (0.87 to 0.99) |
| 2003 | Republic of Moldova | 0.96 (0.89 to 1.02) |
| 2004 | Republic of Moldova | 0.93 (0.87 to 1.00) |
| 2005 | Republic of Moldova | 1.04 (0.97 to 1.11) |
| 2006 | Republic of Moldova | 1.11 (1.04 to 1.19) |
| 2007 | Republic of Moldova | 1.11 (1.03 to 1.19) |
| 2008 | Republic of Moldova | 1.15 (1.06 to 1.24) |
| 2009 | Republic of Moldova | 1.19 (1.10 to 1.29) |
| 2010 | Republic of Moldova | 1.25 (1.15 to 1.36) |
| 2011 | Republic of Moldova | 1.18 (1.08 to 1.28) |
| 2012 | Republic of Moldova | 1.24 (1.15 to 1.35) |
| 2013 | Republic of Moldova | 1.32 (1.23 to 1.43) |
| 2014 | Republic of Moldova | 1.41 (1.31 to 1.52) |
| 2015 | Republic of Moldova | 1.49 (1.39 to 1.61) |
| 2016 | Republic of Moldova | 1.46 (1.36 to 1.60) |
| 2017 | Republic of Moldova | 1.39 (1.29 to 1.51) |
| 2018 | Republic of Moldova | 1.46 (1.35 to 1.60) |
| 2019 | Republic of Moldova | 1.47 (1.32 to 1.62) |
| 2020 | Republic of Moldova | 1.47 (1.34 to 1.61) |
| 2021 | Republic of Moldova | 1.49 (1.32 to 1.67) |
| 1990 | Russian Federation | 2.72 (2.62 to 2.78) |
| 1991 | Russian Federation | 2.78 (2.69 to 2.84) |
| 1992 | Russian Federation | 2.99 (2.90 to 3.06) |
| 1993 | Russian Federation | 3.60 (3.50 to 3.68) |
| 1994 | Russian Federation | 3.95 (3.85 to 4.04) |
| 1995 | Russian Federation | 3.59 (3.49 to 3.66) |
| 1996 | Russian Federation | 3.27 (3.19 to 3.34) |
| 1997 | Russian Federation | 3.05 (2.97 to 3.11) |
| 1998 | Russian Federation | 3.07 (2.98 to 3.14) |
| 1999 | Russian Federation | 4.18 (4.03 to 4.33) |
| 2000 | Russian Federation | 3.91 (3.77 to 4.06) |
| 2001 | Russian Federation | 2.91 (2.83 to 3.00) |
| 2002 | Russian Federation | 2.85 (2.78 to 2.90) |
| 2003 | Russian Federation | 2.87 (2.80 to 2.93) |
| 2004 | Russian Federation | 2.85 (2.78 to 2.91) |
| 2005 | Russian Federation | 3.10 (3.02 to 3.16) |
| 2006 | Russian Federation | 3.02 (2.94 to 3.08) |
| 2007 | Russian Federation | 3.09 (3.00 to 3.16) |
| 2008 | Russian Federation | 3.31 (3.22 to 3.39) |
| 2009 | Russian Federation | 3.42 (3.32 to 3.50) |
| 2010 | Russian Federation | 3.60 (3.49 to 3.67) |
| 2011 | Russian Federation | 3.60 (3.48 to 3.68) |
| 2012 | Russian Federation | 3.68 (3.56 to 3.77) |
| 2013 | Russian Federation | 3.85 (3.71 to 3.94) |
| 2014 | Russian Federation | 4.21 (4.05 to 4.31) |
| 2015 | Russian Federation | 4.33 (4.15 to 4.43) |
| 2016 | Russian Federation | 4.46 (4.27 to 4.57) |
| 2017 | Russian Federation | 4.33 (4.13 to 4.44) |
| 2018 | Russian Federation | 4.40 (4.21 to 4.51) |
| 2019 | Russian Federation | 4.42 (4.21 to 4.52) |
| 2020 | Russian Federation | 4.39 (4.19 to 4.61) |
| 2021 | Russian Federation | 4.38 (4.01 to 4.74) |
| 1990 | Ukraine | 2.13 (1.92 to 2.37) |
| 1991 | Ukraine | 2.27 (2.04 to 2.51) |
| 1992 | Ukraine | 2.42 (2.19 to 2.66) |
| 1993 | Ukraine | 2.54 (2.31 to 2.76) |
| 1994 | Ukraine | 2.66 (2.42 to 2.89) |
| 1995 | Ukraine | 2.87 (2.60 to 3.13) |
| 1996 | Ukraine | 2.80 (2.53 to 3.06) |
| 1997 | Ukraine | 2.69 (2.44 to 2.95) |
| 1998 | Ukraine | 2.53 (2.30 to 2.76) |
| 1999 | Ukraine | 2.61 (2.38 to 2.86) |
| 2000 | Ukraine | 2.71 (2.47 to 2.98) |
| 2001 | Ukraine | 2.66 (2.43 to 2.93) |
| 2002 | Ukraine | 2.72 (2.49 to 3.00) |
| 2003 | Ukraine | 2.74 (2.49 to 3.01) |
| 2004 | Ukraine | 2.86 (2.62 to 3.14) |
| 2005 | Ukraine | 3.06 (2.82 to 3.35) |
| 2006 | Ukraine | 2.95 (2.71 to 3.22) |
| 2007 | Ukraine | 3.08 (2.83 to 3.35) |
| 2008 | Ukraine | 3.12 (2.89 to 3.38) |
| 2009 | Ukraine | 2.78 (2.57 to 2.99) |
| 2010 | Ukraine | 2.67 (2.49 to 2.86) |
| 2011 | Ukraine | 2.58 (2.42 to 2.74) |
| 2012 | Ukraine | 2.59 (2.44 to 2.76) |
| 2013 | Ukraine | 2.61 (2.45 to 2.76) |
| 2014 | Ukraine | 2.55 (2.39 to 2.70) |
| 2015 | Ukraine | 2.49 (2.34 to 2.65) |
| 2016 | Ukraine | 2.50 (2.34 to 2.66) |
| 2017 | Ukraine | 2.53 (2.26 to 2.81) |
| 2018 | Ukraine | 2.59 (2.21 to 3.03) |
| 2019 | Ukraine | 2.62 (2.16 to 3.14) |
| 2020 | Ukraine | 2.47 (2.07 to 2.97) |
| 2021 | Ukraine | 2.47 (1.82 to 3.25) |

#### Fig 3E. ASDR in males and females from Europe in 2021

| **Sex** | **Location** | **ASDR per 100,000** |
| --- | --- | --- |
| Male | Central Europe | 4.74 (4.38 to 5.15) |
| Female | Central Europe | 1.64 (1.42 to 1.95) |
| Male | Eastern Europe | 6.52 (5.85 to 7.16) |
| Female | Eastern Europe | 2.17 (1.92 to 2.45) |
| Male | Western Europe | 3.95 (3.63 to 4.13) |
| Female | Western Europe | 1.50 (1.27 to 1.68) |

#### Fig 3F. ASDR in males and females from Western Europe in 2021

| **Sex** | **Location** | **ASDR per 100,000** |
| --- | --- | --- |
| Male | Andorra | 5.80 (3.35 to 9.23) |
| Female | Andorra | 2.24 (1.45 to 3.29) |
| Male | Austria | 2.52 (2.25 to 2.72) |
| Female | Austria | 1.24 (1.04 to 1.38) |
| Male | Belgium | 3.70 (3.28 to 4.03) |
| Female | Belgium | 1.16 (0.96 to 1.32) |
| Male | Cyprus | 6.54 (4.90 to 8.30) |
| Female | Cyprus | 1.87 (1.42 to 2.38) |
| Male | Denmark | 6.77 (6.01 to 7.44) |
| Female | Denmark | 2.96 (2.55 to 3.34) |
| Male | Finland | 5.58 (4.96 to 6.06) |
| Female | Finland | 2.10 (1.76 to 2.41) |
| Male | France | 3.11 (2.74 to 3.35) |
| Female | France | 1.00 (0.82 to 1.13) |
| Male | Germany | 3.30 (2.96 to 3.57) |
| Female | Germany | 1.39 (1.19 to 1.57) |
| Male | Greece | 6.71 (6.13 to 7.18) |
| Female | Greece | 1.71 (1.46 to 1.89) |
| Male | Iceland | 3.28 (2.86 to 3.67) |
| Female | Iceland | 2.01 (1.65 to 2.31) |
| Male | Ireland | 3.74 (3.21 to 4.19) |
| Female | Ireland | 2.24 (1.81 to 2.65) |
| Male | Israel | 1.88 (1.66 to 2.07) |
| Female | Israel | 0.79 (0.65 to 0.90) |
| Male | Italy | 3.59 (3.28 to 3.79) |
| Female | Italy | 1.12 (0.90 to 1.36) |
| Male | Luxembourg | 3.69 (3.29 to 4.12) |
| Female | Luxembourg | 1.10 (0.95 to 1.27) |
| Male | Malta | 2.30 (2.05 to 2.60) |
| Female | Malta | 0.52 (0.43 to 0.59) |
| Male | Monaco | 6.35 (4.63 to 8.60) |
| Female | Monaco | 4.11 (2.83 to 5.96) |
| Male | Netherlands | 5.64 (4.95 to 6.22) |
| Female | Netherlands | 2.37 (1.98 to 2.70) |
| Male | Norway | 6.32 (5.66 to 6.71) |
| Female | Norway | 3.46 (2.92 to 3.79) |
| Male | Portugal | 2.61 (2.39 to 2.83) |
| Female | Portugal | 0.67 (0.57 to 0.77) |
| Male | San Marino | 3.13 (1.98 to 4.92) |
| Female | San Marino | 0.37 (0.22 to 0.57) |
| Male | Spain | 3.81 (3.43 to 4.08) |
| Female | Spain | 0.76 (0.62 to 0.89) |
| Male | Sweden | 4.83 (4.11 to 5.51) |
| Female | Sweden | 3.09 (2.55 to 3.55) |
| Male | Switzerland | 3.39 (2.99 to 3.70) |
| Female | Switzerland | 1.42 (1.11 to 1.67) |
| Male | United Kingdom of Great Britain and Northern Ireland | 5.51 (5.06 to 5.74) |
| Female | United Kingdom of Great Britain and Northern Ireland | 2.84 (2.41 to 3.13) |

#### Fig 3G. ASDR in males and females from Central Europe

| **Sex** | **Location** | **ASDR per 100,000** |
| --- | --- | --- |
| Male | Albania | 1.58 (0.95 to 2.31) |
| Female | Albania | 0.68 (0.46 to 0.96) |
| Male | Bosnia and Herzegovina | 4.94 (3.26 to 6.82) |
| Female | Bosnia and Herzegovina | 1.54 (1.05 to 2.23) |
| Male | Bulgaria | 4.49 (3.68 to 5.52) |
| Female | Bulgaria | 1.23 (0.94 to 1.61) |
| Male | Croatia | 5.47 (4.69 to 6.32) |
| Female | Croatia | 1.84 (1.49 to 2.34) |
| Male | Czech Republic | 4.15 (3.60 to 4.68) |
| Female | Czech Republic | 1.83 (1.51 to 2.26) |
| Male | Hungary | 4.15 (3.64 to 4.69) |
| Female | Hungary | 1.98 (1.73 to 2.35) |
| Male | Montenegro | 14.76 (10.31 to 21.14) |
| Female | Montenegro | 4.29 (3.31 to 5.25) |
| Male | North Macedonia | 4.06 (2.69 to 5.99) |
| Female | North Macedonia | 1.45 (0.77 to 2.28) |
| Male | Poland | 5.96 (5.38 to 6.55) |
| Female | Poland | 1.79 (1.50 to 2.20) |
| Male | Romania | 2.78 (2.38 to 3.25) |
| Female | Romania | 1.22 (1.00 to 1.55) |
| Male | Serbia | 6.70 (5.06 to 8.79) |
| Female | Serbia | 2.01 (1.54 to 2.61) |
| Male | Slovakia | 3.72 (2.81 to 4.86) |
| Female | Slovakia | 1.28 (0.93 to 1.69) |
| Male | Slovenia | 3.49 (2.91 to 4.11) |
| Female | Slovenia | 1.19 (0.91 to 1.72) |

#### Fig 3H. ASDR in males and females from Eastern Europe in 2021

| **Sex** | **Location** | **ASDR per 100,000** |
| --- | --- | --- |
| Male | Belarus | 7.18 (5.87 to 8.77) |
| Female | Belarus | 1.73 (1.39 to 2.11) |
| Male | Estonia | 6.16 (5.28 to 7.17) |
| Female | Estonia | 1.69 (1.39 to 1.97) |
| Male | Latvia | 5.81 (5.00 to 6.76) |
| Female | Latvia | 1.46 (1.18 to 1.87) |
| Male | Lithuania | 5.67 (4.97 to 6.42) |
| Female | Lithuania | 1.62 (1.32 to 2.05) |
| Male | Republic of Moldova | 2.45 (2.11 to 2.81) |
| Female | Republic of Moldova | 0.84 (0.72 to 0.98) |
| Male | Russian Federation | 7.34 (6.55 to 8.04) |
| Female | Russian Federation | 2.60 (2.30 to 2.98) |
| Male | Ukraine | 4.53 (3.00 to 6.31) |
| Female | Ukraine | 1.16 (0.79 to 1.67) |

### Data for Figure 4

#### Fig 4A. ASDR in Asia from 1990 to 2021

| **Year** | **Location** | **ASDR per 100,000** |
| --- | --- | --- |
| 1990 | Central Asia | 0.94 (0.81 to 1.13) |
| 1991 | Central Asia | 1.00 (0.87 to 1.18) |
| 1992 | Central Asia | 1.07 (0.94 to 1.24) |
| 1993 | Central Asia | 1.14 (1.01 to 1.31) |
| 1994 | Central Asia | 1.16 (1.02 to 1.32) |
| 1995 | Central Asia | 1.20 (1.06 to 1.37) |
| 1996 | Central Asia | 1.21 (1.07 to 1.37) |
| 1997 | Central Asia | 1.18 (1.03 to 1.34) |
| 1998 | Central Asia | 1.16 (1.02 to 1.34) |
| 1999 | Central Asia | 1.11 (0.98 to 1.27) |
| 2000 | Central Asia | 1.15 (1.02 to 1.30) |
| 2001 | Central Asia | 1.15 (1.02 to 1.29) |
| 2002 | Central Asia | 1.19 (1.06 to 1.34) |
| 2003 | Central Asia | 1.27 (1.14 to 1.41) |
| 2004 | Central Asia | 1.27 (1.15 to 1.40) |
| 2005 | Central Asia | 1.32 (1.20 to 1.45) |
| 2006 | Central Asia | 1.36 (1.24 to 1.48) |
| 2007 | Central Asia | 1.43 (1.31 to 1.55) |
| 2008 | Central Asia | 1.45 (1.33 to 1.56) |
| 2009 | Central Asia | 1.45 (1.33 to 1.56) |
| 2010 | Central Asia | 1.54 (1.41 to 1.66) |
| 2011 | Central Asia | 1.60 (1.47 to 1.72) |
| 2012 | Central Asia | 1.69 (1.56 to 1.81) |
| 2013 | Central Asia | 1.74 (1.60 to 1.86) |
| 2014 | Central Asia | 1.81 (1.68 to 1.95) |
| 2015 | Central Asia | 1.95 (1.80 to 2.10) |
| 2016 | Central Asia | 2.00 (1.84 to 2.16) |
| 2017 | Central Asia | 1.97 (1.81 to 2.13) |
| 2018 | Central Asia | 2.05 (1.88 to 2.23) |
| 2019 | Central Asia | 2.01 (1.85 to 2.20) |
| 2020 | Central Asia | 2.00 (1.83 to 2.20) |
| 2021 | Central Asia | 1.98 (1.77 to 2.21) |
| 1990 | East Asia | 0.36 (0.29 to 0.44) |
| 1991 | East Asia | 0.37 (0.30 to 0.45) |
| 1992 | East Asia | 0.37 (0.31 to 0.46) |
| 1993 | East Asia | 0.38 (0.32 to 0.46) |
| 1994 | East Asia | 0.39 (0.34 to 0.46) |
| 1995 | East Asia | 0.40 (0.35 to 0.47) |
| 1996 | East Asia | 0.40 (0.36 to 0.47) |
| 1997 | East Asia | 0.41 (0.36 to 0.46) |
| 1998 | East Asia | 0.41 (0.37 to 0.46) |
| 1999 | East Asia | 0.41 (0.37 to 0.46) |
| 2000 | East Asia | 0.42 (0.38 to 0.46) |
| 2001 | East Asia | 0.42 (0.38 to 0.46) |
| 2002 | East Asia | 0.43 (0.39 to 0.47) |
| 2003 | East Asia | 0.43 (0.40 to 0.47) |
| 2004 | East Asia | 0.42 (0.39 to 0.46) |
| 2005 | East Asia | 0.43 (0.39 to 0.46) |
| 2006 | East Asia | 0.44 (0.40 to 0.47) |
| 2007 | East Asia | 0.45 (0.42 to 0.49) |
| 2008 | East Asia | 0.47 (0.43 to 0.51) |
| 2009 | East Asia | 0.49 (0.44 to 0.53) |
| 2010 | East Asia | 0.50 (0.45 to 0.55) |
| 2011 | East Asia | 0.50 (0.45 to 0.55) |
| 2012 | East Asia | 0.50 (0.44 to 0.55) |
| 2013 | East Asia | 0.49 (0.43 to 0.55) |
| 2014 | East Asia | 0.49 (0.43 to 0.55) |
| 2015 | East Asia | 0.49 (0.43 to 0.56) |
| 2016 | East Asia | 0.50 (0.43 to 0.58) |
| 2017 | East Asia | 0.50 (0.42 to 0.60) |
| 2018 | East Asia | 0.50 (0.42 to 0.59) |
| 2019 | East Asia | 0.50 (0.41 to 0.60) |
| 2020 | East Asia | 0.50 (0.41 to 0.62) |
| 2021 | East Asia | 0.50 (0.41 to 0.63) |
| 1990 | Japan | 2.92 (2.68 to 3.04) |
| 1991 | Japan | 3.05 (2.81 to 3.19) |
| 1992 | Japan | 3.10 (2.84 to 3.23) |
| 1993 | Japan | 3.21 (2.93 to 3.35) |
| 1994 | Japan | 3.27 (2.98 to 3.42) |
| 1995 | Japan | 3.39 (3.09 to 3.56) |
| 1996 | Japan | 3.44 (3.12 to 3.61) |
| 1997 | Japan | 3.44 (3.12 to 3.62) |
| 1998 | Japan | 3.61 (3.26 to 3.80) |
| 1999 | Japan | 3.69 (3.31 to 3.87) |
| 2000 | Japan | 3.77 (3.38 to 3.98) |
| 2001 | Japan | 3.91 (3.49 to 4.14) |
| 2002 | Japan | 3.96 (3.52 to 4.20) |
| 2003 | Japan | 4.01 (3.56 to 4.26) |
| 2004 | Japan | 4.16 (3.68 to 4.43) |
| 2005 | Japan | 4.25 (3.75 to 4.52) |
| 2006 | Japan | 4.26 (3.75 to 4.54) |
| 2007 | Japan | 4.36 (3.82 to 4.65) |
| 2008 | Japan | 4.50 (3.93 to 4.80) |
| 2009 | Japan | 4.63 (4.04 to 4.94) |
| 2010 | Japan | 4.81 (4.19 to 5.15) |
| 2011 | Japan | 4.90 (4.26 to 5.24) |
| 2012 | Japan | 4.86 (4.22 to 5.21) |
| 2013 | Japan | 4.86 (4.20 to 5.23) |
| 2014 | Japan | 4.88 (4.21 to 5.25) |
| 2015 | Japan | 4.93 (4.25 to 5.32) |
| 2016 | Japan | 5.12 (4.42 to 5.53) |
| 2017 | Japan | 5.19 (4.46 to 5.60) |
| 2018 | Japan | 5.16 (4.41 to 5.55) |
| 2019 | Japan | 5.14 (4.39 to 5.53) |
| 2020 | Japan | 5.01 (4.28 to 5.43) |
| 2021 | Japan | 5.07 (4.33 to 5.47) |
| 1990 | Republic of Korea | 1.84 (1.25 to 2.57) |
| 1991 | Republic of Korea | 1.85 (1.30 to 2.51) |
| 1992 | Republic of Korea | 1.84 (1.37 to 2.43) |
| 1993 | Republic of Korea | 1.85 (1.43 to 2.37) |
| 1994 | Republic of Korea | 1.87 (1.52 to 2.33) |
| 1995 | Republic of Korea | 1.87 (1.58 to 2.28) |
| 1996 | Republic of Korea | 1.89 (1.63 to 2.27) |
| 1997 | Republic of Korea | 1.91 (1.66 to 2.25) |
| 1998 | Republic of Korea | 1.98 (1.74 to 2.31) |
| 1999 | Republic of Korea | 2.03 (1.79 to 2.38) |
| 2000 | Republic of Korea | 2.10 (1.86 to 2.43) |
| 2001 | Republic of Korea | 2.12 (1.89 to 2.42) |
| 2002 | Republic of Korea | 2.13 (1.89 to 2.44) |
| 2003 | Republic of Korea | 2.16 (1.91 to 2.46) |
| 2004 | Republic of Korea | 2.16 (1.91 to 2.47) |
| 2005 | Republic of Korea | 2.17 (1.92 to 2.47) |
| 2006 | Republic of Korea | 2.15 (1.89 to 2.42) |
| 2007 | Republic of Korea | 2.13 (1.86 to 2.39) |
| 2008 | Republic of Korea | 2.09 (1.81 to 2.35) |
| 2009 | Republic of Korea | 2.08 (1.78 to 2.35) |
| 2010 | Republic of Korea | 2.05 (1.77 to 2.32) |
| 2011 | Republic of Korea | 2.03 (1.74 to 2.30) |
| 2012 | Republic of Korea | 1.99 (1.69 to 2.24) |
| 2013 | Republic of Korea | 1.96 (1.67 to 2.20) |
| 2014 | Republic of Korea | 1.94 (1.64 to 2.18) |
| 2015 | Republic of Korea | 1.92 (1.62 to 2.18) |
| 2016 | Republic of Korea | 1.93 (1.61 to 2.19) |
| 2017 | Republic of Korea | 1.90 (1.58 to 2.18) |
| 2018 | Republic of Korea | 1.91 (1.58 to 2.20) |
| 2019 | Republic of Korea | 1.91 (1.54 to 2.22) |
| 2020 | Republic of Korea | 1.87 (1.51 to 2.20) |
| 2021 | Republic of Korea | 1.92 (1.56 to 2.27) |
| 1990 | Singapore | 2.65 (2.49 to 2.79) |
| 1991 | Singapore | 2.76 (2.60 to 2.91) |
| 1992 | Singapore | 2.76 (2.59 to 2.92) |
| 1993 | Singapore | 2.71 (2.54 to 2.86) |
| 1994 | Singapore | 2.63 (2.47 to 2.78) |
| 1995 | Singapore | 2.67 (2.51 to 2.82) |
| 1996 | Singapore | 2.61 (2.45 to 2.76) |
| 1997 | Singapore | 2.54 (2.38 to 2.69) |
| 1998 | Singapore | 2.80 (2.62 to 2.96) |
| 1999 | Singapore | 2.75 (2.56 to 2.92) |
| 2000 | Singapore | 2.84 (2.65 to 3.02) |
| 2001 | Singapore | 2.83 (2.63 to 3.02) |
| 2002 | Singapore | 2.91 (2.69 to 3.09) |
| 2003 | Singapore | 2.72 (2.51 to 2.89) |
| 2004 | Singapore | 2.79 (2.57 to 2.96) |
| 2005 | Singapore | 2.71 (2.50 to 2.88) |
| 2006 | Singapore | 2.81 (2.59 to 2.99) |
| 2007 | Singapore | 2.79 (2.57 to 2.96) |
| 2008 | Singapore | 2.76 (2.54 to 2.93) |
| 2009 | Singapore | 2.70 (2.47 to 2.88) |
| 2010 | Singapore | 2.71 (2.48 to 2.89) |
| 2011 | Singapore | 2.58 (2.36 to 2.74) |
| 2012 | Singapore | 2.58 (2.35 to 2.74) |
| 2013 | Singapore | 2.56 (2.32 to 2.74) |
| 2014 | Singapore | 2.54 (2.30 to 2.72) |
| 2015 | Singapore | 2.40 (2.16 to 2.57) |
| 2016 | Singapore | 2.40 (2.15 to 2.56) |
| 2017 | Singapore | 2.44 (2.20 to 2.60) |
| 2018 | Singapore | 2.41 (2.16 to 2.57) |
| 2019 | Singapore | 2.30 (2.04 to 2.46) |
| 2020 | Singapore | 2.25 (2.02 to 2.42) |
| 2021 | Singapore | 2.15 (1.91 to 2.32) |
| 1990 | South Asia | 0.71 (0.45 to 1.13) |
| 1991 | South Asia | 0.72 (0.45 to 1.14) |
| 1992 | South Asia | 0.74 (0.47 to 1.16) |
| 1993 | South Asia | 0.75 (0.48 to 1.18) |
| 1994 | South Asia | 0.78 (0.51 to 1.21) |
| 1995 | South Asia | 0.82 (0.54 to 1.27) |
| 1996 | South Asia | 0.82 (0.54 to 1.27) |
| 1997 | South Asia | 0.82 (0.54 to 1.28) |
| 1998 | South Asia | 0.81 (0.52 to 1.28) |
| 1999 | South Asia | 0.81 (0.54 to 1.25) |
| 2000 | South Asia | 0.80 (0.53 to 1.26) |
| 2001 | South Asia | 0.81 (0.54 to 1.27) |
| 2002 | South Asia | 0.83 (0.55 to 1.27) |
| 2003 | South Asia | 0.85 (0.57 to 1.31) |
| 2004 | South Asia | 0.90 (0.61 to 1.36) |
| 2005 | South Asia | 0.89 (0.61 to 1.34) |
| 2006 | South Asia | 0.90 (0.61 to 1.37) |
| 2007 | South Asia | 0.94 (0.64 to 1.41) |
| 2008 | South Asia | 0.95 (0.64 to 1.44) |
| 2009 | South Asia | 0.96 (0.66 to 1.47) |
| 2010 | South Asia | 0.99 (0.68 to 1.48) |
| 2011 | South Asia | 1.01 (0.69 to 1.52) |
| 2012 | South Asia | 1.03 (0.72 to 1.52) |
| 2013 | South Asia | 1.07 (0.73 to 1.61) |
| 2014 | South Asia | 1.12 (0.76 to 1.67) |
| 2015 | South Asia | 1.16 (0.82 to 1.71) |
| 2016 | South Asia | 1.17 (0.84 to 1.72) |
| 2017 | South Asia | 1.21 (0.87 to 1.76) |
| 2018 | South Asia | 1.22 (0.87 to 1.76) |
| 2019 | South Asia | 1.24 (0.87 to 1.79) |
| 2020 | South Asia | 1.24 (0.88 to 1.78) |
| 2021 | South Asia | 1.22 (0.88 to 1.78) |
| 1990 | Southeast Asia | 1.03 (0.83 to 1.29) |
| 1991 | Southeast Asia | 1.04 (0.85 to 1.30) |
| 1992 | Southeast Asia | 1.06 (0.86 to 1.31) |
| 1993 | Southeast Asia | 1.09 (0.89 to 1.33) |
| 1994 | Southeast Asia | 1.10 (0.91 to 1.34) |
| 1995 | Southeast Asia | 1.12 (0.93 to 1.35) |
| 1996 | Southeast Asia | 1.14 (0.96 to 1.37) |
| 1997 | Southeast Asia | 1.14 (0.96 to 1.35) |
| 1998 | Southeast Asia | 1.17 (0.98 to 1.38) |
| 1999 | Southeast Asia | 1.18 (0.99 to 1.39) |
| 2000 | Southeast Asia | 1.19 (1.01 to 1.40) |
| 2001 | Southeast Asia | 1.20 (1.02 to 1.41) |
| 2002 | Southeast Asia | 1.20 (1.03 to 1.40) |
| 2003 | Southeast Asia | 1.22 (1.05 to 1.40) |
| 2004 | Southeast Asia | 1.23 (1.05 to 1.41) |
| 2005 | Southeast Asia | 1.23 (1.07 to 1.42) |
| 2006 | Southeast Asia | 1.24 (1.07 to 1.42) |
| 2007 | Southeast Asia | 1.24 (1.07 to 1.43) |
| 2008 | Southeast Asia | 1.25 (1.09 to 1.43) |
| 2009 | Southeast Asia | 1.26 (1.10 to 1.42) |
| 2010 | Southeast Asia | 1.27 (1.11 to 1.44) |
| 2011 | Southeast Asia | 1.27 (1.12 to 1.45) |
| 2012 | Southeast Asia | 1.26 (1.11 to 1.43) |
| 2013 | Southeast Asia | 1.26 (1.10 to 1.43) |
| 2014 | Southeast Asia | 1.27 (1.11 to 1.43) |
| 2015 | Southeast Asia | 1.28 (1.13 to 1.45) |
| 2016 | Southeast Asia | 1.29 (1.14 to 1.46) |
| 2017 | Southeast Asia | 1.31 (1.15 to 1.48) |
| 2018 | Southeast Asia | 1.34 (1.18 to 1.52) |
| 2019 | Southeast Asia | 1.36 (1.20 to 1.54) |
| 2020 | Southeast Asia | 1.35 (1.16 to 1.53) |
| 2021 | Southeast Asia | 1.39 (1.21 to 1.60) |

#### Fig 4B. ASDR in South Asia from 1990 to 2021

| **Year** | **Location** | **ASDR** |
| --- | --- | --- |
| 1990 | Nepal | 0.64 (0.37 to 1.13) |
| 1991 | Nepal | 0.65 (0.38 to 1.14) |
| 1992 | Nepal | 0.65 (0.38 to 1.11) |
| 1993 | Nepal | 0.66 (0.39 to 1.12) |
| 1994 | Nepal | 0.67 (0.40 to 1.13) |
| 1995 | Nepal | 0.68 (0.40 to 1.14) |
| 1996 | Nepal | 0.68 (0.42 to 1.13) |
| 1997 | Nepal | 0.68 (0.43 to 1.13) |
| 1998 | Nepal | 0.69 (0.43 to 1.14) |
| 1999 | Nepal | 0.70 (0.44 to 1.15) |
| 2000 | Nepal | 0.70 (0.45 to 1.15) |
| 2001 | Nepal | 0.71 (0.46 to 1.14) |
| 2002 | Nepal | 0.72 (0.48 to 1.16) |
| 2003 | Nepal | 0.73 (0.48 to 1.17) |
| 2004 | Nepal | 0.74 (0.50 to 1.18) |
| 2005 | Nepal | 0.76 (0.51 to 1.22) |
| 2006 | Nepal | 0.78 (0.53 to 1.26) |
| 2007 | Nepal | 0.81 (0.54 to 1.28) |
| 2008 | Nepal | 0.83 (0.55 to 1.32) |
| 2009 | Nepal | 0.86 (0.58 to 1.37) |
| 2010 | Nepal | 0.89 (0.60 to 1.41) |
| 2011 | Nepal | 0.93 (0.63 to 1.49) |
| 2012 | Nepal | 0.95 (0.64 to 1.54) |
| 2013 | Nepal | 0.99 (0.67 to 1.57) |
| 2014 | Nepal | 1.01 (0.68 to 1.62) |
| 2015 | Nepal | 1.06 (0.72 to 1.67) |
| 2016 | Nepal | 1.08 (0.73 to 1.70) |
| 2017 | Nepal | 1.12 (0.76 to 1.72) |
| 2018 | Nepal | 1.14 (0.77 to 1.79) |
| 2019 | Nepal | 1.16 (0.79 to 1.82) |
| 2020 | Nepal | 1.17 (0.81 to 1.84) |
| 2021 | Nepal | 1.19 (0.82 to 1.85) |
| 1990 | Pakistan | 0.84 (0.57 to 1.29) |
| 1991 | Pakistan | 0.87 (0.59 to 1.33) |
| 1992 | Pakistan | 0.91 (0.61 to 1.37) |
| 1993 | Pakistan | 0.95 (0.64 to 1.42) |
| 1994 | Pakistan | 1.00 (0.68 to 1.50) |
| 1995 | Pakistan | 1.05 (0.70 to 1.59) |
| 1996 | Pakistan | 1.09 (0.75 to 1.61) |
| 1997 | Pakistan | 1.13 (0.77 to 1.67) |
| 1998 | Pakistan | 1.16 (0.80 to 1.71) |
| 1999 | Pakistan | 1.19 (0.82 to 1.74) |
| 2000 | Pakistan | 1.22 (0.86 to 1.78) |
| 2001 | Pakistan | 1.26 (0.87 to 1.84) |
| 2002 | Pakistan | 1.29 (0.90 to 1.87) |
| 2003 | Pakistan | 1.32 (0.94 to 1.91) |
| 2004 | Pakistan | 1.35 (0.97 to 1.95) |
| 2005 | Pakistan | 1.37 (0.97 to 1.95) |
| 2006 | Pakistan | 1.38 (0.98 to 1.93) |
| 2007 | Pakistan | 1.39 (0.99 to 1.97) |
| 2008 | Pakistan | 1.40 (1.01 to 1.98) |
| 2009 | Pakistan | 1.41 (1.02 to 1.98) |
| 2010 | Pakistan | 1.42 (1.03 to 1.98) |
| 2011 | Pakistan | 1.41 (1.02 to 1.95) |
| 2012 | Pakistan | 1.40 (1.02 to 2.00) |
| 2013 | Pakistan | 1.40 (1.05 to 1.94) |
| 2014 | Pakistan | 1.40 (1.01 to 1.97) |
| 2015 | Pakistan | 1.41 (1.03 to 1.99) |
| 2016 | Pakistan | 1.42 (1.02 to 1.93) |
| 2017 | Pakistan | 1.44 (1.04 to 1.97) |
| 2018 | Pakistan | 1.44 (1.06 to 1.96) |
| 2019 | Pakistan | 1.45 (1.08 to 2.01) |
| 2020 | Pakistan | 1.47 (1.06 to 2.01) |
| 2021 | Pakistan | 1.48 (1.06 to 2.04) |
| 1990 | Bhutan | 0.78 (0.51 to 1.16) |
| 1991 | Bhutan | 0.79 (0.53 to 1.15) |
| 1992 | Bhutan | 0.80 (0.53 to 1.17) |
| 1993 | Bhutan | 0.82 (0.54 to 1.19) |
| 1994 | Bhutan | 0.84 (0.56 to 1.21) |
| 1995 | Bhutan | 0.86 (0.58 to 1.24) |
| 1996 | Bhutan | 0.86 (0.58 to 1.26) |
| 1997 | Bhutan | 0.89 (0.61 to 1.28) |
| 1998 | Bhutan | 0.90 (0.62 to 1.27) |
| 1999 | Bhutan | 0.90 (0.63 to 1.27) |
| 2000 | Bhutan | 0.93 (0.66 to 1.31) |
| 2001 | Bhutan | 0.95 (0.67 to 1.31) |
| 2002 | Bhutan | 0.97 (0.70 to 1.32) |
| 2003 | Bhutan | 1.00 (0.73 to 1.37) |
| 2004 | Bhutan | 1.03 (0.75 to 1.42) |
| 2005 | Bhutan | 1.06 (0.76 to 1.45) |
| 2006 | Bhutan | 1.08 (0.76 to 1.50) |
| 2007 | Bhutan | 1.11 (0.78 to 1.53) |
| 2008 | Bhutan | 1.15 (0.79 to 1.59) |
| 2009 | Bhutan | 1.19 (0.82 to 1.66) |
| 2010 | Bhutan | 1.22 (0.82 to 1.70) |
| 2011 | Bhutan | 1.26 (0.83 to 1.74) |
| 2012 | Bhutan | 1.28 (0.84 to 1.77) |
| 2013 | Bhutan | 1.31 (0.86 to 1.82) |
| 2014 | Bhutan | 1.33 (0.86 to 1.85) |
| 2015 | Bhutan | 1.36 (0.87 to 1.93) |
| 2016 | Bhutan | 1.39 (0.89 to 1.97) |
| 2017 | Bhutan | 1.42 (0.90 to 2.01) |
| 2018 | Bhutan | 1.44 (0.90 to 2.05) |
| 2019 | Bhutan | 1.46 (0.92 to 2.07) |
| 2020 | Bhutan | 1.48 (0.94 to 2.12) |
| 2021 | Bhutan | 1.49 (0.93 to 2.16) |
| 1990 | Bangladesh | 0.82 (0.49 to 1.42) |
| 1991 | Bangladesh | 0.80 (0.49 to 1.41) |
| 1992 | Bangladesh | 0.81 (0.50 to 1.42) |
| 1993 | Bangladesh | 0.82 (0.51 to 1.44) |
| 1994 | Bangladesh | 0.83 (0.52 to 1.46) |
| 1995 | Bangladesh | 0.85 (0.53 to 1.49) |
| 1996 | Bangladesh | 0.86 (0.53 to 1.50) |
| 1997 | Bangladesh | 0.86 (0.54 to 1.53) |
| 1998 | Bangladesh | 0.87 (0.55 to 1.54) |
| 1999 | Bangladesh | 0.92 (0.57 to 1.60) |
| 2000 | Bangladesh | 0.94 (0.58 to 1.66) |
| 2001 | Bangladesh | 0.95 (0.58 to 1.70) |
| 2002 | Bangladesh | 0.98 (0.60 to 1.75) |
| 2003 | Bangladesh | 1.00 (0.61 to 1.78) |
| 2004 | Bangladesh | 0.99 (0.61 to 1.78) |
| 2005 | Bangladesh | 1.00 (0.62 to 1.77) |
| 2006 | Bangladesh | 1.03 (0.64 to 1.84) |
| 2007 | Bangladesh | 1.07 (0.67 to 1.89) |
| 2008 | Bangladesh | 1.09 (0.71 to 1.90) |
| 2009 | Bangladesh | 1.10 (0.74 to 1.86) |
| 2010 | Bangladesh | 1.12 (0.74 to 1.88) |
| 2011 | Bangladesh | 1.05 (0.72 to 1.74) |
| 2012 | Bangladesh | 0.99 (0.68 to 1.66) |
| 2013 | Bangladesh | 1.00 (0.68 to 1.67) |
| 2014 | Bangladesh | 1.04 (0.70 to 1.79) |
| 2015 | Bangladesh | 1.05 (0.71 to 1.82) |
| 2016 | Bangladesh | 1.08 (0.72 to 1.89) |
| 2017 | Bangladesh | 1.15 (0.77 to 2.03) |
| 2018 | Bangladesh | 1.17 (0.78 to 2.05) |
| 2019 | Bangladesh | 1.19 (0.79 to 2.04) |
| 2020 | Bangladesh | 1.20 (0.80 to 2.06) |
| 2021 | Bangladesh | 1.21 (0.80 to 1.99) |
| 1990 | India | 0.68 (0.40 to 1.08) |
| 1991 | India | 0.68 (0.41 to 1.09) |
| 1992 | India | 0.70 (0.41 to 1.12) |
| 1993 | India | 0.71 (0.43 to 1.13) |
| 1994 | India | 0.74 (0.46 to 1.17) |
| 1995 | India | 0.78 (0.49 to 1.23) |
| 1996 | India | 0.78 (0.49 to 1.23) |
| 1997 | India | 0.77 (0.48 to 1.23) |
| 1998 | India | 0.76 (0.46 to 1.21) |
| 1999 | India | 0.75 (0.48 to 1.17) |
| 2000 | India | 0.73 (0.46 to 1.17) |
| 2001 | India | 0.75 (0.47 to 1.18) |
| 2002 | India | 0.76 (0.48 to 1.20) |
| 2003 | India | 0.79 (0.51 to 1.23) |
| 2004 | India | 0.84 (0.56 to 1.29) |
| 2005 | India | 0.84 (0.56 to 1.27) |
| 2006 | India | 0.84 (0.55 to 1.29) |
| 2007 | India | 0.89 (0.58 to 1.34) |
| 2008 | India | 0.90 (0.58 to 1.38) |
| 2009 | India | 0.91 (0.60 to 1.39) |
| 2010 | India | 0.94 (0.63 to 1.42) |
| 2011 | India | 0.97 (0.65 to 1.48) |
| 2012 | India | 1.00 (0.68 to 1.48) |
| 2013 | India | 1.05 (0.69 to 1.59) |
| 2014 | India | 1.10 (0.72 to 1.67) |
| 2015 | India | 1.15 (0.78 to 1.69) |
| 2016 | India | 1.16 (0.81 to 1.71) |
| 2017 | India | 1.20 (0.84 to 1.74) |
| 2018 | India | 1.21 (0.84 to 1.75) |
| 2019 | India | 1.23 (0.86 to 1.77) |
| 2020 | India | 1.22 (0.83 to 1.76) |
| 2021 | India | 1.20 (0.86 to 1.74) |

#### Fig 4C. ASDR in Central Asia from 1990 to 2021

| **Year** | **Location** | **ASDR per 100,000** |
| --- | --- | --- |
| 1990 | Armenia | 4.55 (3.71 to 5.57) |
| 1991 | Armenia | 4.75 (3.85 to 5.85) |
| 1992 | Armenia | 5.18 (4.25 to 6.32) |
| 1993 | Armenia | 5.42 (4.47 to 6.61) |
| 1994 | Armenia | 5.24 (4.30 to 6.40) |
| 1995 | Armenia | 5.11 (4.22 to 6.25) |
| 1996 | Armenia | 5.13 (4.24 to 6.39) |
| 1997 | Armenia | 4.89 (4.03 to 6.09) |
| 1998 | Armenia | 4.80 (3.94 to 5.84) |
| 1999 | Armenia | 4.86 (4.06 to 5.87) |
| 2000 | Armenia | 4.80 (4.03 to 5.74) |
| 2001 | Armenia | 4.88 (4.10 to 5.85) |
| 2002 | Armenia | 4.97 (4.18 to 5.98) |
| 2003 | Armenia | 5.14 (4.27 to 6.14) |
| 2004 | Armenia | 5.31 (4.43 to 6.26) |
| 2005 | Armenia | 5.70 (4.76 to 6.68) |
| 2006 | Armenia | 5.93 (4.91 to 6.95) |
| 2007 | Armenia | 6.08 (5.08 to 7.07) |
| 2008 | Armenia | 6.25 (5.26 to 7.20) |
| 2009 | Armenia | 6.67 (5.60 to 7.70) |
| 2010 | Armenia | 7.58 (6.39 to 8.67) |
| 2011 | Armenia | 7.79 (6.56 to 8.92) |
| 2012 | Armenia | 7.88 (6.62 to 9.00) |
| 2013 | Armenia | 7.95 (6.73 to 9.04) |
| 2014 | Armenia | 8.35 (7.11 to 9.48) |
| 2015 | Armenia | 9.02 (7.74 to 10.25) |
| 2016 | Armenia | 9.02 (7.81 to 10.26) |
| 2017 | Armenia | 8.55 (7.36 to 9.69) |
| 2018 | Armenia | 9.26 (8.08 to 10.51) |
| 2019 | Armenia | 9.06 (7.89 to 10.25) |
| 2020 | Armenia | 9.10 (7.74 to 10.51) |
| 2021 | Armenia | 9.16 (7.61 to 10.81) |
| 1990 | Azerbaijan | 0.73 (0.55 to 0.96) |
| 1991 | Azerbaijan | 0.74 (0.57 to 0.96) |
| 1992 | Azerbaijan | 0.77 (0.61 to 0.96) |
| 1993 | Azerbaijan | 0.77 (0.62 to 0.95) |
| 1994 | Azerbaijan | 0.80 (0.64 to 1.00) |
| 1995 | Azerbaijan | 0.77 (0.62 to 0.99) |
| 1996 | Azerbaijan | 0.77 (0.60 to 0.99) |
| 1997 | Azerbaijan | 0.75 (0.57 to 0.99) |
| 1998 | Azerbaijan | 0.75 (0.55 to 1.00) |
| 1999 | Azerbaijan | 0.75 (0.54 to 1.01) |
| 2000 | Azerbaijan | 0.75 (0.55 to 1.02) |
| 2001 | Azerbaijan | 0.77 (0.56 to 1.05) |
| 2002 | Azerbaijan | 0.83 (0.61 to 1.09) |
| 2003 | Azerbaijan | 0.90 (0.69 to 1.18) |
| 2004 | Azerbaijan | 0.97 (0.75 to 1.22) |
| 2005 | Azerbaijan | 1.04 (0.83 to 1.27) |
| 2006 | Azerbaijan | 1.08 (0.88 to 1.34) |
| 2007 | Azerbaijan | 1.14 (0.92 to 1.40) |
| 2008 | Azerbaijan | 1.19 (0.95 to 1.46) |
| 2009 | Azerbaijan | 1.22 (0.98 to 1.55) |
| 2010 | Azerbaijan | 1.26 (0.99 to 1.61) |
| 2011 | Azerbaijan | 1.27 (0.99 to 1.64) |
| 2012 | Azerbaijan | 1.29 (1.00 to 1.68) |
| 2013 | Azerbaijan | 1.31 (1.02 to 1.71) |
| 2014 | Azerbaijan | 1.30 (0.99 to 1.70) |
| 2015 | Azerbaijan | 1.33 (1.00 to 1.75) |
| 2016 | Azerbaijan | 1.32 (0.95 to 1.75) |
| 2017 | Azerbaijan | 1.35 (0.98 to 1.80) |
| 2018 | Azerbaijan | 1.31 (0.95 to 1.77) |
| 2019 | Azerbaijan | 1.37 (0.98 to 1.97) |
| 2020 | Azerbaijan | 1.34 (0.96 to 1.98) |
| 2021 | Azerbaijan | 1.37 (0.93 to 2.08) |
| 1990 | Georgia | 0.61 (0.51 to 0.72) |
| 1991 | Georgia | 0.61 (0.52 to 0.73) |
| 1992 | Georgia | 0.61 (0.52 to 0.73) |
| 1993 | Georgia | 0.58 (0.50 to 0.69) |
| 1994 | Georgia | 0.56 (0.49 to 0.66) |
| 1995 | Georgia | 0.56 (0.49 to 0.66) |
| 1996 | Georgia | 0.59 (0.51 to 0.68) |
| 1997 | Georgia | 0.59 (0.50 to 0.70) |
| 1998 | Georgia | 0.61 (0.52 to 0.71) |
| 1999 | Georgia | 0.37 (0.32 to 0.43) |
| 2000 | Georgia | 0.44 (0.40 to 0.49) |
| 2001 | Georgia | 0.40 (0.36 to 0.44) |
| 2002 | Georgia | 0.45 (0.41 to 0.48) |
| 2003 | Georgia | 0.62 (0.57 to 0.68) |
| 2004 | Georgia | 0.71 (0.64 to 0.78) |
| 2005 | Georgia | 0.72 (0.63 to 0.79) |
| 2006 | Georgia | 0.81 (0.71 to 0.91) |
| 2007 | Georgia | 1.09 (0.96 to 1.23) |
| 2008 | Georgia | 1.36 (1.18 to 1.54) |
| 2009 | Georgia | 1.41 (1.22 to 1.60) |
| 2010 | Georgia | 1.59 (1.36 to 1.78) |
| 2011 | Georgia | 2.05 (1.78 to 2.31) |
| 2012 | Georgia | 2.26 (1.96 to 2.54) |
| 2013 | Georgia | 2.43 (2.11 to 2.73) |
| 2014 | Georgia | 2.52 (2.19 to 2.85) |
| 2015 | Georgia | 2.75 (2.40 to 3.13) |
| 2016 | Georgia | 2.93 (2.58 to 3.31) |
| 2017 | Georgia | 2.73 (2.40 to 3.10) |
| 2018 | Georgia | 2.96 (2.58 to 3.35) |
| 2019 | Georgia | 2.89 (2.50 to 3.27) |
| 2020 | Georgia | 2.99 (2.58 to 3.40) |
| 2021 | Georgia | 2.88 (2.43 to 3.35) |
| 1990 | Kazakhstan | 1.46 (1.19 to 1.87) |
| 1991 | Kazakhstan | 1.61 (1.32 to 2.04) |
| 1992 | Kazakhstan | 1.76 (1.44 to 2.18) |
| 1993 | Kazakhstan | 1.97 (1.62 to 2.43) |
| 1994 | Kazakhstan | 2.10 (1.71 to 2.58) |
| 1995 | Kazakhstan | 2.31 (1.88 to 2.82) |
| 1996 | Kazakhstan | 2.33 (1.91 to 2.84) |
| 1997 | Kazakhstan | 2.29 (1.85 to 2.81) |
| 1998 | Kazakhstan | 2.24 (1.81 to 2.74) |
| 1999 | Kazakhstan | 2.15 (1.73 to 2.62) |
| 2000 | Kazakhstan | 2.25 (1.82 to 2.76) |
| 2001 | Kazakhstan | 2.26 (1.82 to 2.77) |
| 2002 | Kazakhstan | 2.28 (1.84 to 2.80) |
| 2003 | Kazakhstan | 2.36 (1.93 to 2.86) |
| 2004 | Kazakhstan | 2.28 (1.89 to 2.73) |
| 2005 | Kazakhstan | 2.28 (1.89 to 2.72) |
| 2006 | Kazakhstan | 2.29 (1.91 to 2.70) |
| 2007 | Kazakhstan | 2.30 (1.94 to 2.69) |
| 2008 | Kazakhstan | 2.16 (1.82 to 2.50) |
| 2009 | Kazakhstan | 2.06 (1.75 to 2.38) |
| 2010 | Kazakhstan | 2.12 (1.83 to 2.41) |
| 2011 | Kazakhstan | 2.10 (1.81 to 2.37) |
| 2012 | Kazakhstan | 2.12 (1.85 to 2.40) |
| 2013 | Kazakhstan | 2.09 (1.82 to 2.41) |
| 2014 | Kazakhstan | 2.11 (1.82 to 2.42) |
| 2015 | Kazakhstan | 2.18 (1.88 to 2.51) |
| 2016 | Kazakhstan | 2.25 (1.92 to 2.60) |
| 2017 | Kazakhstan | 2.29 (1.94 to 2.65) |
| 2018 | Kazakhstan | 2.31 (1.94 to 2.73) |
| 2019 | Kazakhstan | 2.32 (1.94 to 2.76) |
| 2020 | Kazakhstan | 2.28 (1.87 to 2.72) |
| 2021 | Kazakhstan | 2.23 (1.80 to 2.71) |
| 1990 | Kyrgyzstan | 0.28 (0.24 to 0.33) |
| 1991 | Kyrgyzstan | 0.29 (0.25 to 0.34) |
| 1992 | Kyrgyzstan | 0.30 (0.26 to 0.34) |
| 1993 | Kyrgyzstan | 0.32 (0.28 to 0.36) |
| 1994 | Kyrgyzstan | 0.33 (0.30 to 0.37) |
| 1995 | Kyrgyzstan | 0.33 (0.30 to 0.37) |
| 1996 | Kyrgyzstan | 0.32 (0.29 to 0.36) |
| 1997 | Kyrgyzstan | 0.32 (0.28 to 0.36) |
| 1998 | Kyrgyzstan | 0.31 (0.28 to 0.33) |
| 1999 | Kyrgyzstan | 0.30 (0.28 to 0.32) |
| 2000 | Kyrgyzstan | 0.31 (0.29 to 0.33) |
| 2001 | Kyrgyzstan | 0.28 (0.26 to 0.31) |
| 2002 | Kyrgyzstan | 0.47 (0.44 to 0.51) |
| 2003 | Kyrgyzstan | 0.61 (0.56 to 0.66) |
| 2004 | Kyrgyzstan | 0.42 (0.38 to 0.46) |
| 2005 | Kyrgyzstan | 0.41 (0.38 to 0.45) |
| 2006 | Kyrgyzstan | 0.46 (0.42 to 0.51) |
| 2007 | Kyrgyzstan | 0.57 (0.51 to 0.63) |
| 2008 | Kyrgyzstan | 0.63 (0.56 to 0.70) |
| 2009 | Kyrgyzstan | 0.50 (0.44 to 0.56) |
| 2010 | Kyrgyzstan | 0.43 (0.37 to 0.48) |
| 2011 | Kyrgyzstan | 0.44 (0.39 to 0.49) |
| 2012 | Kyrgyzstan | 0.62 (0.55 to 0.69) |
| 2013 | Kyrgyzstan | 0.69 (0.61 to 0.77) |
| 2014 | Kyrgyzstan | 0.73 (0.65 to 0.82) |
| 2015 | Kyrgyzstan | 0.79 (0.70 to 0.89) |
| 2016 | Kyrgyzstan | 0.81 (0.72 to 0.92) |
| 2017 | Kyrgyzstan | 0.81 (0.71 to 0.91) |
| 2018 | Kyrgyzstan | 0.84 (0.73 to 0.96) |
| 2019 | Kyrgyzstan | 0.81 (0.70 to 0.94) |
| 2020 | Kyrgyzstan | 0.81 (0.67 to 0.95) |
| 2021 | Kyrgyzstan | 0.79 (0.63 to 0.96) |
| 1990 | Mongolia | 0.31 (0.22 to 0.42) |
| 1991 | Mongolia | 0.32 (0.23 to 0.43) |
| 1992 | Mongolia | 0.33 (0.23 to 0.44) |
| 1993 | Mongolia | 0.33 (0.23 to 0.46) |
| 1994 | Mongolia | 0.33 (0.23 to 0.45) |
| 1995 | Mongolia | 0.34 (0.24 to 0.47) |
| 1996 | Mongolia | 0.35 (0.25 to 0.47) |
| 1997 | Mongolia | 0.35 (0.25 to 0.46) |
| 1998 | Mongolia | 0.35 (0.25 to 0.46) |
| 1999 | Mongolia | 0.34 (0.26 to 0.46) |
| 2000 | Mongolia | 0.34 (0.26 to 0.46) |
| 2001 | Mongolia | 0.34 (0.25 to 0.45) |
| 2002 | Mongolia | 0.36 (0.26 to 0.48) |
| 2003 | Mongolia | 0.37 (0.28 to 0.50) |
| 2004 | Mongolia | 0.39 (0.29 to 0.53) |
| 2005 | Mongolia | 0.40 (0.30 to 0.55) |
| 2006 | Mongolia | 0.41 (0.30 to 0.55) |
| 2007 | Mongolia | 0.41 (0.30 to 0.56) |
| 2008 | Mongolia | 0.41 (0.30 to 0.55) |
| 2009 | Mongolia | 0.42 (0.31 to 0.57) |
| 2010 | Mongolia | 0.41 (0.30 to 0.56) |
| 2011 | Mongolia | 0.46 (0.33 to 0.62) |
| 2012 | Mongolia | 0.50 (0.37 to 0.65) |
| 2013 | Mongolia | 0.54 (0.40 to 0.70) |
| 2014 | Mongolia | 0.58 (0.44 to 0.78) |
| 2015 | Mongolia | 0.61 (0.47 to 0.81) |
| 2016 | Mongolia | 0.64 (0.49 to 0.83) |
| 2017 | Mongolia | 0.67 (0.52 to 0.86) |
| 2018 | Mongolia | 0.70 (0.54 to 0.90) |
| 2019 | Mongolia | 0.72 (0.55 to 0.92) |
| 2020 | Mongolia | 0.70 (0.54 to 0.90) |
| 2021 | Mongolia | 0.70 (0.53 to 0.89) |
| 1990 | Tajikistan | 0.28 (0.19 to 0.36) |
| 1991 | Tajikistan | 0.29 (0.20 to 0.38) |
| 1992 | Tajikistan | 0.30 (0.22 to 0.39) |
| 1993 | Tajikistan | 0.29 (0.22 to 0.36) |
| 1994 | Tajikistan | 0.29 (0.23 to 0.36) |
| 1995 | Tajikistan | 0.30 (0.23 to 0.37) |
| 1996 | Tajikistan | 0.29 (0.22 to 0.36) |
| 1997 | Tajikistan | 0.27 (0.21 to 0.33) |
| 1998 | Tajikistan | 0.27 (0.20 to 0.34) |
| 1999 | Tajikistan | 0.27 (0.20 to 0.35) |
| 2000 | Tajikistan | 0.27 (0.19 to 0.35) |
| 2001 | Tajikistan | 0.26 (0.20 to 0.35) |
| 2002 | Tajikistan | 0.27 (0.20 to 0.36) |
| 2003 | Tajikistan | 0.28 (0.21 to 0.37) |
| 2004 | Tajikistan | 0.29 (0.22 to 0.37) |
| 2005 | Tajikistan | 0.30 (0.22 to 0.38) |
| 2006 | Tajikistan | 0.30 (0.23 to 0.38) |
| 2007 | Tajikistan | 0.31 (0.25 to 0.39) |
| 2008 | Tajikistan | 0.31 (0.25 to 0.40) |
| 2009 | Tajikistan | 0.32 (0.25 to 0.41) |
| 2010 | Tajikistan | 0.30 (0.23 to 0.38) |
| 2011 | Tajikistan | 0.29 (0.23 to 0.37) |
| 2012 | Tajikistan | 0.28 (0.22 to 0.35) |
| 2013 | Tajikistan | 0.26 (0.20 to 0.34) |
| 2014 | Tajikistan | 0.26 (0.20 to 0.33) |
| 2015 | Tajikistan | 0.25 (0.20 to 0.32) |
| 2016 | Tajikistan | 0.25 (0.19 to 0.31) |
| 2017 | Tajikistan | 0.25 (0.19 to 0.31) |
| 2018 | Tajikistan | 0.24 (0.19 to 0.32) |
| 2019 | Tajikistan | 0.24 (0.19 to 0.31) |
| 2020 | Tajikistan | 0.24 (0.18 to 0.32) |
| 2021 | Tajikistan | 0.25 (0.18 to 0.34) |
| 1990 | Turkmenistan | 1.03 (0.84 to 1.26) |
| 1991 | Turkmenistan | 1.08 (0.89 to 1.33) |
| 1992 | Turkmenistan | 1.08 (0.90 to 1.32) |
| 1993 | Turkmenistan | 1.21 (1.02 to 1.49) |
| 1994 | Turkmenistan | 1.22 (1.03 to 1.48) |
| 1995 | Turkmenistan | 1.19 (1.00 to 1.43) |
| 1996 | Turkmenistan | 1.22 (1.04 to 1.47) |
| 1997 | Turkmenistan | 1.21 (1.04 to 1.47) |
| 1998 | Turkmenistan | 1.22 (1.03 to 1.47) |
| 1999 | Turkmenistan | 1.26 (1.07 to 1.50) |
| 2000 | Turkmenistan | 1.29 (1.08 to 1.52) |
| 2001 | Turkmenistan | 1.27 (1.07 to 1.51) |
| 2002 | Turkmenistan | 1.33 (1.12 to 1.57) |
| 2003 | Turkmenistan | 1.37 (1.16 to 1.60) |
| 2004 | Turkmenistan | 1.46 (1.25 to 1.69) |
| 2005 | Turkmenistan | 1.58 (1.34 to 1.83) |
| 2006 | Turkmenistan | 1.60 (1.37 to 1.83) |
| 2007 | Turkmenistan | 1.61 (1.37 to 1.85) |
| 2008 | Turkmenistan | 1.57 (1.34 to 1.84) |
| 2009 | Turkmenistan | 1.48 (1.25 to 1.74) |
| 2010 | Turkmenistan | 1.47 (1.24 to 1.74) |
| 2011 | Turkmenistan | 1.50 (1.25 to 1.77) |
| 2012 | Turkmenistan | 1.52 (1.26 to 1.81) |
| 2013 | Turkmenistan | 1.62 (1.35 to 1.94) |
| 2014 | Turkmenistan | 1.65 (1.37 to 1.99) |
| 2015 | Turkmenistan | 1.69 (1.39 to 2.08) |
| 2016 | Turkmenistan | 1.72 (1.39 to 2.17) |
| 2017 | Turkmenistan | 1.78 (1.42 to 2.31) |
| 2018 | Turkmenistan | 1.82 (1.40 to 2.44) |
| 2019 | Turkmenistan | 1.87 (1.41 to 2.58) |
| 2020 | Turkmenistan | 1.88 (1.43 to 2.63) |
| 2021 | Turkmenistan | 1.88 (1.42 to 2.67) |
| 1990 | Uzbekistan | 0.21 (0.16 to 0.30) |
| 1991 | Uzbekistan | 0.22 (0.17 to 0.31) |
| 1992 | Uzbekistan | 0.23 (0.17 to 0.31) |
| 1993 | Uzbekistan | 0.24 (0.18 to 0.32) |
| 1994 | Uzbekistan | 0.25 (0.19 to 0.34) |
| 1995 | Uzbekistan | 0.26 (0.19 to 0.34) |
| 1996 | Uzbekistan | 0.26 (0.20 to 0.35) |
| 1997 | Uzbekistan | 0.26 (0.20 to 0.34) |
| 1998 | Uzbekistan | 0.26 (0.20 to 0.34) |
| 1999 | Uzbekistan | 0.26 (0.21 to 0.34) |
| 2000 | Uzbekistan | 0.27 (0.22 to 0.34) |
| 2001 | Uzbekistan | 0.27 (0.22 to 0.34) |
| 2002 | Uzbekistan | 0.28 (0.23 to 0.35) |
| 2003 | Uzbekistan | 0.29 (0.24 to 0.36) |
| 2004 | Uzbekistan | 0.30 (0.25 to 0.37) |
| 2005 | Uzbekistan | 0.33 (0.27 to 0.40) |
| 2006 | Uzbekistan | 0.36 (0.29 to 0.43) |
| 2007 | Uzbekistan | 0.37 (0.30 to 0.45) |
| 2008 | Uzbekistan | 0.38 (0.31 to 0.45) |
| 2009 | Uzbekistan | 0.37 (0.31 to 0.45) |
| 2010 | Uzbekistan | 0.38 (0.31 to 0.45) |
| 2011 | Uzbekistan | 0.38 (0.32 to 0.45) |
| 2012 | Uzbekistan | 0.52 (0.44 to 0.61) |
| 2013 | Uzbekistan | 0.62 (0.53 to 0.72) |
| 2014 | Uzbekistan | 0.75 (0.65 to 0.88) |
| 2015 | Uzbekistan | 0.90 (0.77 to 1.05) |
| 2016 | Uzbekistan | 0.99 (0.84 to 1.15) |
| 2017 | Uzbekistan | 1.03 (0.87 to 1.19) |
| 2018 | Uzbekistan | 1.08 (0.91 to 1.25) |
| 2019 | Uzbekistan | 1.04 (0.89 to 1.22) |
| 2020 | Uzbekistan | 1.04 (0.86 to 1.25) |
| 2021 | Uzbekistan | 1.04 (0.85 to 1.28) |

#### Fig 4D. ASDR in Southeast Asia from 1990 to 2021

| **Year** | **Location** | **ASDR per 100,000** |
| --- | --- | --- |
| 1990 | Cambodia | 0.55 (0.32 to 0.87) |
| 1991 | Cambodia | 0.55 (0.33 to 0.85) |
| 1992 | Cambodia | 0.56 (0.33 to 0.85) |
| 1993 | Cambodia | 0.56 (0.34 to 0.86) |
| 1994 | Cambodia | 0.57 (0.34 to 0.86) |
| 1995 | Cambodia | 0.57 (0.35 to 0.86) |
| 1996 | Cambodia | 0.57 (0.35 to 0.85) |
| 1997 | Cambodia | 0.57 (0.36 to 0.85) |
| 1998 | Cambodia | 0.57 (0.36 to 0.84) |
| 1999 | Cambodia | 0.57 (0.37 to 0.84) |
| 2000 | Cambodia | 0.57 (0.37 to 0.84) |
| 2001 | Cambodia | 0.57 (0.38 to 0.84) |
| 2002 | Cambodia | 0.57 (0.38 to 0.84) |
| 2003 | Cambodia | 0.57 (0.39 to 0.83) |
| 2004 | Cambodia | 0.58 (0.39 to 0.82) |
| 2005 | Cambodia | 0.58 (0.40 to 0.82) |
| 2006 | Cambodia | 0.59 (0.40 to 0.83) |
| 2007 | Cambodia | 0.60 (0.41 to 0.85) |
| 2008 | Cambodia | 0.61 (0.42 to 0.86) |
| 2009 | Cambodia | 0.63 (0.43 to 0.86) |
| 2010 | Cambodia | 0.64 (0.43 to 0.88) |
| 2011 | Cambodia | 0.65 (0.44 to 0.91) |
| 2012 | Cambodia | 0.66 (0.44 to 0.92) |
| 2013 | Cambodia | 0.67 (0.45 to 0.93) |
| 2014 | Cambodia | 0.68 (0.45 to 0.96) |
| 2015 | Cambodia | 0.70 (0.46 to 0.99) |
| 2016 | Cambodia | 0.71 (0.47 to 1.04) |
| 2017 | Cambodia | 0.72 (0.47 to 1.07) |
| 2018 | Cambodia | 0.73 (0.48 to 1.11) |
| 2019 | Cambodia | 0.74 (0.48 to 1.15) |
| 2020 | Cambodia | 0.74 (0.48 to 1.17) |
| 2021 | Cambodia | 0.75 (0.47 to 1.21) |
| 1990 | Indonesia | 0.63 (0.41 to 0.87) |
| 1991 | Indonesia | 0.65 (0.42 to 0.89) |
| 1992 | Indonesia | 0.67 (0.44 to 0.90) |
| 1993 | Indonesia | 0.69 (0.46 to 0.92) |
| 1994 | Indonesia | 0.71 (0.49 to 0.95) |
| 1995 | Indonesia | 0.72 (0.49 to 0.95) |
| 1996 | Indonesia | 0.74 (0.51 to 0.97) |
| 1997 | Indonesia | 0.75 (0.52 to 0.99) |
| 1998 | Indonesia | 0.77 (0.53 to 1.00) |
| 1999 | Indonesia | 0.78 (0.54 to 1.04) |
| 2000 | Indonesia | 0.80 (0.56 to 1.04) |
| 2001 | Indonesia | 0.81 (0.56 to 1.07) |
| 2002 | Indonesia | 0.82 (0.57 to 1.07) |
| 2003 | Indonesia | 0.83 (0.58 to 1.09) |
| 2004 | Indonesia | 0.85 (0.59 to 1.12) |
| 2005 | Indonesia | 0.86 (0.60 to 1.12) |
| 2006 | Indonesia | 0.87 (0.61 to 1.14) |
| 2007 | Indonesia | 0.88 (0.61 to 1.17) |
| 2008 | Indonesia | 0.89 (0.61 to 1.18) |
| 2009 | Indonesia | 0.89 (0.62 to 1.19) |
| 2010 | Indonesia | 0.90 (0.62 to 1.20) |
| 2011 | Indonesia | 0.92 (0.62 to 1.23) |
| 2012 | Indonesia | 0.92 (0.64 to 1.24) |
| 2013 | Indonesia | 0.93 (0.64 to 1.24) |
| 2014 | Indonesia | 0.94 (0.65 to 1.27) |
| 2015 | Indonesia | 0.95 (0.65 to 1.28) |
| 2016 | Indonesia | 0.97 (0.65 to 1.28) |
| 2017 | Indonesia | 0.98 (0.67 to 1.32) |
| 2018 | Indonesia | 0.99 (0.68 to 1.34) |
| 2019 | Indonesia | 1.01 (0.68 to 1.34) |
| 2020 | Indonesia | 1.02 (0.68 to 1.38) |
| 2021 | Indonesia | 1.03 (0.71 to 1.36) |
| 1990 | Lao People's Democratic Republic | 0.78 (0.51 to 1.22) |
| 1991 | Lao People's Democratic Republic | 0.79 (0.51 to 1.21) |
| 1992 | Lao People's Democratic Republic | 0.79 (0.52 to 1.21) |
| 1993 | Lao People's Democratic Republic | 0.80 (0.52 to 1.20) |
| 1994 | Lao People's Democratic Republic | 0.80 (0.53 to 1.19) |
| 1995 | Lao People's Democratic Republic | 0.80 (0.54 to 1.18) |
| 1996 | Lao People's Democratic Republic | 0.81 (0.55 to 1.17) |
| 1997 | Lao People's Democratic Republic | 0.81 (0.56 to 1.17) |
| 1998 | Lao People's Democratic Republic | 0.82 (0.57 to 1.16) |
| 1999 | Lao People's Democratic Republic | 0.82 (0.58 to 1.16) |
| 2000 | Lao People's Democratic Republic | 0.82 (0.58 to 1.13) |
| 2001 | Lao People's Democratic Republic | 0.83 (0.60 to 1.13) |
| 2002 | Lao People's Democratic Republic | 0.82 (0.60 to 1.11) |
| 2003 | Lao People's Democratic Republic | 0.83 (0.60 to 1.09) |
| 2004 | Lao People's Democratic Republic | 0.83 (0.61 to 1.09) |
| 2005 | Lao People's Democratic Republic | 0.83 (0.61 to 1.09) |
| 2006 | Lao People's Democratic Republic | 0.84 (0.61 to 1.11) |
| 2007 | Lao People's Democratic Republic | 0.84 (0.62 to 1.11) |
| 2008 | Lao People's Democratic Republic | 0.85 (0.64 to 1.13) |
| 2009 | Lao People's Democratic Republic | 0.86 (0.65 to 1.12) |
| 2010 | Lao People's Democratic Republic | 0.86 (0.66 to 1.12) |
| 2011 | Lao People's Democratic Republic | 0.87 (0.67 to 1.11) |
| 2012 | Lao People's Democratic Republic | 0.87 (0.66 to 1.14) |
| 2013 | Lao People's Democratic Republic | 0.88 (0.67 to 1.16) |
| 2014 | Lao People's Democratic Republic | 0.89 (0.67 to 1.20) |
| 2015 | Lao People's Democratic Republic | 0.90 (0.68 to 1.21) |
| 2016 | Lao People's Democratic Republic | 0.90 (0.68 to 1.23) |
| 2017 | Lao People's Democratic Republic | 0.91 (0.69 to 1.26) |
| 2018 | Lao People's Democratic Republic | 0.91 (0.69 to 1.25) |
| 2019 | Lao People's Democratic Republic | 0.92 (0.70 to 1.24) |
| 2020 | Lao People's Democratic Republic | 0.92 (0.69 to 1.27) |
| 2021 | Lao People's Democratic Republic | 0.93 (0.68 to 1.28) |
| 1990 | Malaysia | 3.30 (2.66 to 4.00) |
| 1991 | Malaysia | 3.26 (2.66 to 3.96) |
| 1992 | Malaysia | 3.35 (2.74 to 4.01) |
| 1993 | Malaysia | 3.51 (2.87 to 4.14) |
| 1994 | Malaysia | 3.49 (2.92 to 4.21) |
| 1995 | Malaysia | 3.67 (3.09 to 4.41) |
| 1996 | Malaysia | 3.81 (3.24 to 4.45) |
| 1997 | Malaysia | 3.94 (3.35 to 4.51) |
| 1998 | Malaysia | 4.15 (3.54 to 4.74) |
| 1999 | Malaysia | 4.45 (3.90 to 5.01) |
| 2000 | Malaysia | 4.35 (3.79 to 4.87) |
| 2001 | Malaysia | 4.21 (3.75 to 4.71) |
| 2002 | Malaysia | 4.08 (3.66 to 4.48) |
| 2003 | Malaysia | 4.38 (3.94 to 4.83) |
| 2004 | Malaysia | 4.40 (3.96 to 4.85) |
| 2005 | Malaysia | 4.32 (3.88 to 4.74) |
| 2006 | Malaysia | 4.18 (3.73 to 4.60) |
| 2007 | Malaysia | 4.11 (3.67 to 4.54) |
| 2008 | Malaysia | 4.17 (3.73 to 4.59) |
| 2009 | Malaysia | 4.26 (3.81 to 4.72) |
| 2010 | Malaysia | 4.27 (3.69 to 4.83) |
| 2011 | Malaysia | 4.13 (3.62 to 4.71) |
| 2012 | Malaysia | 3.99 (3.56 to 4.48) |
| 2013 | Malaysia | 3.92 (3.41 to 4.50) |
| 2014 | Malaysia | 3.95 (3.46 to 4.49) |
| 2015 | Malaysia | 4.02 (3.52 to 4.57) |
| 2016 | Malaysia | 3.97 (3.43 to 4.59) |
| 2017 | Malaysia | 3.97 (3.42 to 4.59) |
| 2018 | Malaysia | 3.97 (3.41 to 4.65) |
| 2019 | Malaysia | 4.01 (3.40 to 4.74) |
| 2020 | Malaysia | 3.43 (2.87 to 4.08) |
| 2021 | Malaysia | 4.04 (3.38 to 4.88) |
| 1990 | Maldives | 0.66 (0.47 to 0.91) |
| 1991 | Maldives | 0.68 (0.49 to 0.92) |
| 1992 | Maldives | 0.70 (0.51 to 0.94) |
| 1993 | Maldives | 0.72 (0.51 to 0.96) |
| 1994 | Maldives | 0.73 (0.51 to 0.98) |
| 1995 | Maldives | 0.74 (0.53 to 1.00) |
| 1996 | Maldives | 0.74 (0.56 to 0.98) |
| 1997 | Maldives | 0.76 (0.57 to 0.99) |
| 1998 | Maldives | 0.76 (0.56 to 0.99) |
| 1999 | Maldives | 0.73 (0.56 to 0.93) |
| 2000 | Maldives | 0.74 (0.57 to 0.94) |
| 2001 | Maldives | 0.73 (0.55 to 0.93) |
| 2002 | Maldives | 0.71 (0.53 to 0.93) |
| 2003 | Maldives | 0.70 (0.51 to 0.92) |
| 2004 | Maldives | 0.69 (0.50 to 0.92) |
| 2005 | Maldives | 0.68 (0.48 to 0.90) |
| 2006 | Maldives | 0.67 (0.46 to 0.89) |
| 2007 | Maldives | 0.66 (0.45 to 0.87) |
| 2008 | Maldives | 0.65 (0.44 to 0.85) |
| 2009 | Maldives | 0.64 (0.42 to 0.85) |
| 2010 | Maldives | 0.63 (0.41 to 0.84) |
| 2011 | Maldives | 0.62 (0.40 to 0.83) |
| 2012 | Maldives | 0.62 (0.39 to 0.83) |
| 2013 | Maldives | 0.60 (0.38 to 0.81) |
| 2014 | Maldives | 0.60 (0.37 to 0.82) |
| 2015 | Maldives | 0.61 (0.36 to 0.85) |
| 2016 | Maldives | 0.60 (0.35 to 0.85) |
| 2017 | Maldives | 0.57 (0.34 to 0.79) |
| 2018 | Maldives | 0.60 (0.33 to 0.88) |
| 2019 | Maldives | 0.61 (0.32 to 0.95) |
| 2020 | Maldives | 0.59 (0.30 to 0.94) |
| 2021 | Maldives | 0.60 (0.31 to 0.97) |
| 1990 | Mauritius | 1.50 (1.40 to 1.60) |
| 1991 | Mauritius | 1.42 (1.31 to 1.52) |
| 1992 | Mauritius | 1.25 (1.17 to 1.34) |
| 1993 | Mauritius | 1.03 (0.97 to 1.11) |
| 1994 | Mauritius | 1.11 (1.03 to 1.18) |
| 1995 | Mauritius | 1.07 (0.99 to 1.15) |
| 1996 | Mauritius | 0.85 (0.79 to 0.91) |
| 1997 | Mauritius | 0.99 (0.92 to 1.05) |
| 1998 | Mauritius | 0.91 (0.85 to 0.97) |
| 1999 | Mauritius | 0.86 (0.79 to 0.91) |
| 2000 | Mauritius | 1.02 (0.95 to 1.09) |
| 2001 | Mauritius | 1.11 (1.03 to 1.17) |
| 2002 | Mauritius | 1.05 (0.98 to 1.11) |
| 2003 | Mauritius | 1.06 (0.99 to 1.12) |
| 2004 | Mauritius | 1.05 (0.99 to 1.11) |
| 2005 | Mauritius | 0.95 (0.88 to 1.00) |
| 2006 | Mauritius | 0.87 (0.81 to 0.92) |
| 2007 | Mauritius | 0.80 (0.75 to 0.85) |
| 2008 | Mauritius | 0.81 (0.76 to 0.86) |
| 2009 | Mauritius | 0.84 (0.78 to 0.88) |
| 2010 | Mauritius | 0.84 (0.78 to 0.89) |
| 2011 | Mauritius | 0.80 (0.74 to 0.85) |
| 2012 | Mauritius | 0.76 (0.70 to 0.80) |
| 2013 | Mauritius | 0.76 (0.70 to 0.81) |
| 2014 | Mauritius | 0.78 (0.72 to 0.83) |
| 2015 | Mauritius | 0.75 (0.70 to 0.80) |
| 2016 | Mauritius | 0.82 (0.76 to 0.88) |
| 2017 | Mauritius | 0.83 (0.77 to 0.89) |
| 2018 | Mauritius | 0.86 (0.80 to 0.92) |
| 2019 | Mauritius | 0.86 (0.79 to 0.92) |
| 2020 | Mauritius | 0.83 (0.76 to 0.89) |
| 2021 | Mauritius | 0.88 (0.81 to 0.94) |
| 1990 | Myanmar | 0.80 (0.47 to 1.16) |
| 1991 | Myanmar | 0.80 (0.47 to 1.19) |
| 1992 | Myanmar | 0.80 (0.48 to 1.20) |
| 1993 | Myanmar | 0.81 (0.49 to 1.21) |
| 1994 | Myanmar | 0.81 (0.50 to 1.21) |
| 1995 | Myanmar | 0.82 (0.52 to 1.21) |
| 1996 | Myanmar | 0.83 (0.52 to 1.20) |
| 1997 | Myanmar | 0.83 (0.52 to 1.21) |
| 1998 | Myanmar | 0.84 (0.53 to 1.21) |
| 1999 | Myanmar | 0.84 (0.55 to 1.21) |
| 2000 | Myanmar | 0.85 (0.56 to 1.21) |
| 2001 | Myanmar | 0.86 (0.57 to 1.23) |
| 2002 | Myanmar | 0.86 (0.57 to 1.21) |
| 2003 | Myanmar | 0.86 (0.57 to 1.20) |
| 2004 | Myanmar | 0.87 (0.58 to 1.21) |
| 2005 | Myanmar | 0.87 (0.60 to 1.19) |
| 2006 | Myanmar | 0.87 (0.61 to 1.19) |
| 2007 | Myanmar | 0.87 (0.63 to 1.19) |
| 2008 | Myanmar | 0.87 (0.63 to 1.19) |
| 2009 | Myanmar | 0.87 (0.64 to 1.19) |
| 2010 | Myanmar | 0.87 (0.65 to 1.19) |
| 2011 | Myanmar | 0.89 (0.66 to 1.20) |
| 2012 | Myanmar | 0.88 (0.66 to 1.18) |
| 2013 | Myanmar | 0.89 (0.67 to 1.18) |
| 2014 | Myanmar | 0.90 (0.69 to 1.18) |
| 2015 | Myanmar | 0.91 (0.70 to 1.17) |
| 2016 | Myanmar | 0.92 (0.71 to 1.17) |
| 2017 | Myanmar | 0.93 (0.72 to 1.20) |
| 2018 | Myanmar | 0.94 (0.71 to 1.23) |
| 2019 | Myanmar | 0.95 (0.72 to 1.23) |
| 2020 | Myanmar | 0.95 (0.72 to 1.25) |
| 2021 | Myanmar | 0.96 (0.73 to 1.26) |
| 1990 | Philippines | 1.22 (1.07 to 1.39) |
| 1991 | Philippines | 1.18 (1.02 to 1.35) |
| 1992 | Philippines | 1.16 (1.02 to 1.31) |
| 1993 | Philippines | 1.16 (1.03 to 1.31) |
| 1994 | Philippines | 1.18 (1.05 to 1.32) |
| 1995 | Philippines | 1.19 (1.06 to 1.33) |
| 1996 | Philippines | 1.22 (1.09 to 1.35) |
| 1997 | Philippines | 1.22 (1.08 to 1.36) |
| 1998 | Philippines | 1.24 (1.10 to 1.36) |
| 1999 | Philippines | 1.24 (1.11 to 1.38) |
| 2000 | Philippines | 1.26 (1.14 to 1.38) |
| 2001 | Philippines | 1.28 (1.15 to 1.40) |
| 2002 | Philippines | 1.27 (1.17 to 1.37) |
| 2003 | Philippines | 1.26 (1.15 to 1.36) |
| 2004 | Philippines | 1.25 (1.13 to 1.37) |
| 2005 | Philippines | 1.25 (1.14 to 1.35) |
| 2006 | Philippines | 1.25 (1.14 to 1.34) |
| 2007 | Philippines | 1.25 (1.14 to 1.36) |
| 2008 | Philippines | 1.26 (1.15 to 1.36) |
| 2009 | Philippines | 1.27 (1.16 to 1.37) |
| 2010 | Philippines | 1.27 (1.16 to 1.38) |
| 2011 | Philippines | 1.28 (1.18 to 1.39) |
| 2012 | Philippines | 1.28 (1.16 to 1.38) |
| 2013 | Philippines | 1.28 (1.16 to 1.40) |
| 2014 | Philippines | 1.26 (1.17 to 1.37) |
| 2015 | Philippines | 1.32 (1.21 to 1.45) |
| 2016 | Philippines | 1.32 (1.20 to 1.42) |
| 2017 | Philippines | 1.32 (1.21 to 1.42) |
| 2018 | Philippines | 1.33 (1.20 to 1.46) |
| 2019 | Philippines | 1.34 (1.21 to 1.49) |
| 2020 | Philippines | 1.34 (1.20 to 1.47) |
| 2021 | Philippines | 1.35 (1.12 to 1.58) |
| 1990 | Seychelles | 1.08 (0.88 to 1.29) |
| 1991 | Seychelles | 1.09 (0.90 to 1.30) |
| 1992 | Seychelles | 1.10 (0.92 to 1.31) |
| 1993 | Seychelles | 1.12 (0.93 to 1.33) |
| 1994 | Seychelles | 1.14 (0.95 to 1.35) |
| 1995 | Seychelles | 1.15 (0.96 to 1.37) |
| 1996 | Seychelles | 1.16 (0.96 to 1.39) |
| 1997 | Seychelles | 1.16 (0.96 to 1.43) |
| 1998 | Seychelles | 1.16 (0.94 to 1.45) |
| 1999 | Seychelles | 1.16 (0.92 to 1.46) |
| 2000 | Seychelles | 1.14 (0.89 to 1.46) |
| 2001 | Seychelles | 1.13 (0.88 to 1.45) |
| 2002 | Seychelles | 1.17 (0.90 to 1.49) |
| 2003 | Seychelles | 1.21 (0.94 to 1.54) |
| 2004 | Seychelles | 1.22 (0.95 to 1.57) |
| 2005 | Seychelles | 1.25 (0.97 to 1.58) |
| 2006 | Seychelles | 1.24 (0.95 to 1.57) |
| 2007 | Seychelles | 1.23 (0.93 to 1.56) |
| 2008 | Seychelles | 1.21 (0.92 to 1.54) |
| 2009 | Seychelles | 1.22 (0.93 to 1.55) |
| 2010 | Seychelles | 1.18 (0.89 to 1.54) |
| 2011 | Seychelles | 1.16 (0.85 to 1.55) |
| 2012 | Seychelles | 1.16 (0.87 to 1.51) |
| 2013 | Seychelles | 1.16 (0.86 to 1.54) |
| 2014 | Seychelles | 1.15 (0.85 to 1.54) |
| 2015 | Seychelles | 1.16 (0.84 to 1.57) |
| 2016 | Seychelles | 1.18 (0.84 to 1.59) |
| 2017 | Seychelles | 1.20 (0.85 to 1.62) |
| 2018 | Seychelles | 1.21 (0.85 to 1.65) |
| 2019 | Seychelles | 1.21 (0.85 to 1.68) |
| 2020 | Seychelles | 1.01 (0.70 to 1.42) |
| 2021 | Seychelles | 1.07 (0.74 to 1.51) |
| 1990 | Sri Lanka | 0.35 (0.29 to 0.42) |
| 1991 | Sri Lanka | 0.33 (0.28 to 0.38) |
| 1992 | Sri Lanka | 0.33 (0.28 to 0.38) |
| 1993 | Sri Lanka | 0.32 (0.27 to 0.38) |
| 1994 | Sri Lanka | 0.33 (0.28 to 0.38) |
| 1995 | Sri Lanka | 0.34 (0.29 to 0.39) |
| 1996 | Sri Lanka | 0.33 (0.28 to 0.39) |
| 1997 | Sri Lanka | 0.35 (0.30 to 0.40) |
| 1998 | Sri Lanka | 0.34 (0.29 to 0.39) |
| 1999 | Sri Lanka | 0.34 (0.30 to 0.40) |
| 2000 | Sri Lanka | 0.35 (0.30 to 0.40) |
| 2001 | Sri Lanka | 0.34 (0.30 to 0.39) |
| 2002 | Sri Lanka | 0.34 (0.30 to 0.39) |
| 2003 | Sri Lanka | 0.36 (0.32 to 0.41) |
| 2004 | Sri Lanka | 0.37 (0.32 to 0.42) |
| 2005 | Sri Lanka | 0.39 (0.34 to 0.44) |
| 2006 | Sri Lanka | 0.40 (0.35 to 0.45) |
| 2007 | Sri Lanka | 0.40 (0.36 to 0.46) |
| 2008 | Sri Lanka | 0.41 (0.36 to 0.45) |
| 2009 | Sri Lanka | 0.40 (0.35 to 0.45) |
| 2010 | Sri Lanka | 0.39 (0.35 to 0.44) |
| 2011 | Sri Lanka | 0.40 (0.36 to 0.45) |
| 2012 | Sri Lanka | 0.40 (0.35 to 0.44) |
| 2013 | Sri Lanka | 0.40 (0.36 to 0.45) |
| 2014 | Sri Lanka | 0.40 (0.36 to 0.45) |
| 2015 | Sri Lanka | 0.41 (0.34 to 0.47) |
| 2016 | Sri Lanka | 0.39 (0.31 to 0.48) |
| 2017 | Sri Lanka | 0.39 (0.30 to 0.49) |
| 2018 | Sri Lanka | 0.39 (0.29 to 0.51) |
| 2019 | Sri Lanka | 0.38 (0.27 to 0.50) |
| 2020 | Sri Lanka | 0.34 (0.20 to 0.49) |
| 2021 | Sri Lanka | 0.37 (0.25 to 0.50) |
| 1990 | Thailand | 2.02 (1.53 to 2.71) |
| 1991 | Thailand | 2.07 (1.58 to 2.72) |
| 1992 | Thailand | 2.11 (1.58 to 2.75) |
| 1993 | Thailand | 2.16 (1.64 to 2.84) |
| 1994 | Thailand | 2.18 (1.66 to 2.83) |
| 1995 | Thailand | 2.19 (1.67 to 2.82) |
| 1996 | Thailand | 2.23 (1.71 to 2.82) |
| 1997 | Thailand | 2.14 (1.65 to 2.74) |
| 1998 | Thailand | 2.19 (1.65 to 2.82) |
| 1999 | Thailand | 2.15 (1.62 to 2.73) |
| 2000 | Thailand | 2.15 (1.68 to 2.69) |
| 2001 | Thailand | 2.14 (1.71 to 2.67) |
| 2002 | Thailand | 2.16 (1.74 to 2.66) |
| 2003 | Thailand | 2.15 (1.72 to 2.65) |
| 2004 | Thailand | 2.16 (1.74 to 2.58) |
| 2005 | Thailand | 2.13 (1.74 to 2.59) |
| 2006 | Thailand | 2.11 (1.71 to 2.61) |
| 2007 | Thailand | 2.08 (1.68 to 2.52) |
| 2008 | Thailand | 2.06 (1.71 to 2.45) |
| 2009 | Thailand | 2.02 (1.69 to 2.40) |
| 2010 | Thailand | 2.01 (1.69 to 2.39) |
| 2011 | Thailand | 1.98 (1.66 to 2.35) |
| 2012 | Thailand | 1.92 (1.60 to 2.31) |
| 2013 | Thailand | 1.86 (1.55 to 2.22) |
| 2014 | Thailand | 1.86 (1.53 to 2.22) |
| 2015 | Thailand | 1.83 (1.54 to 2.15) |
| 2016 | Thailand | 1.85 (1.56 to 2.15) |
| 2017 | Thailand | 1.86 (1.55 to 2.18) |
| 2018 | Thailand | 1.93 (1.60 to 2.29) |
| 2019 | Thailand | 1.98 (1.64 to 2.37) |
| 2020 | Thailand | 1.99 (1.61 to 2.46) |
| 2021 | Thailand | 2.01 (1.55 to 2.58) |
| 1990 | Timor-Leste | 0.54 (0.38 to 0.82) |
| 1991 | Timor-Leste | 0.55 (0.39 to 0.82) |
| 1992 | Timor-Leste | 0.56 (0.40 to 0.83) |
| 1993 | Timor-Leste | 0.56 (0.41 to 0.82) |
| 1994 | Timor-Leste | 0.57 (0.42 to 0.84) |
| 1995 | Timor-Leste | 0.57 (0.43 to 0.83) |
| 1996 | Timor-Leste | 0.57 (0.43 to 0.82) |
| 1997 | Timor-Leste | 0.57 (0.43 to 0.80) |
| 1998 | Timor-Leste | 0.58 (0.44 to 0.80) |
| 1999 | Timor-Leste | 0.58 (0.43 to 0.80) |
| 2000 | Timor-Leste | 0.58 (0.43 to 0.81) |
| 2001 | Timor-Leste | 0.58 (0.43 to 0.81) |
| 2002 | Timor-Leste | 0.58 (0.44 to 0.82) |
| 2003 | Timor-Leste | 0.59 (0.44 to 0.82) |
| 2004 | Timor-Leste | 0.58 (0.44 to 0.82) |
| 2005 | Timor-Leste | 0.59 (0.44 to 0.83) |
| 2006 | Timor-Leste | 0.60 (0.45 to 0.85) |
| 2007 | Timor-Leste | 0.60 (0.45 to 0.86) |
| 2008 | Timor-Leste | 0.61 (0.46 to 0.88) |
| 2009 | Timor-Leste | 0.63 (0.47 to 0.91) |
| 2010 | Timor-Leste | 0.65 (0.47 to 0.94) |
| 2011 | Timor-Leste | 0.67 (0.48 to 0.99) |
| 2012 | Timor-Leste | 0.69 (0.49 to 1.04) |
| 2013 | Timor-Leste | 0.71 (0.49 to 1.07) |
| 2014 | Timor-Leste | 0.72 (0.50 to 1.08) |
| 2015 | Timor-Leste | 0.72 (0.49 to 1.09) |
| 2016 | Timor-Leste | 0.72 (0.49 to 1.09) |
| 2017 | Timor-Leste | 0.73 (0.49 to 1.12) |
| 2018 | Timor-Leste | 0.73 (0.49 to 1.13) |
| 2019 | Timor-Leste | 0.74 (0.49 to 1.14) |
| 2020 | Timor-Leste | 0.75 (0.49 to 1.15) |
| 2021 | Timor-Leste | 0.74 (0.49 to 1.15) |
| 1990 | Viet Nam | 0.67 (0.48 to 0.90) |
| 1991 | Viet Nam | 0.68 (0.49 to 0.92) |
| 1992 | Viet Nam | 0.69 (0.50 to 0.94) |
| 1993 | Viet Nam | 0.70 (0.51 to 0.94) |
| 1994 | Viet Nam | 0.72 (0.52 to 0.96) |
| 1995 | Viet Nam | 0.73 (0.54 to 0.97) |
| 1996 | Viet Nam | 0.75 (0.55 to 0.99) |
| 1997 | Viet Nam | 0.76 (0.56 to 1.00) |
| 1998 | Viet Nam | 0.77 (0.58 to 1.02) |
| 1999 | Viet Nam | 0.79 (0.59 to 1.03) |
| 2000 | Viet Nam | 0.80 (0.59 to 1.04) |
| 2001 | Viet Nam | 0.81 (0.60 to 1.07) |
| 2002 | Viet Nam | 0.83 (0.61 to 1.09) |
| 2003 | Viet Nam | 0.85 (0.62 to 1.10) |
| 2004 | Viet Nam | 0.87 (0.64 to 1.12) |
| 2005 | Viet Nam | 0.89 (0.66 to 1.13) |
| 2006 | Viet Nam | 0.90 (0.67 to 1.15) |
| 2007 | Viet Nam | 0.92 (0.69 to 1.15) |
| 2008 | Viet Nam | 0.93 (0.70 to 1.16) |
| 2009 | Viet Nam | 0.94 (0.71 to 1.20) |
| 2010 | Viet Nam | 0.96 (0.71 to 1.22) |
| 2011 | Viet Nam | 0.97 (0.72 to 1.24) |
| 2012 | Viet Nam | 0.99 (0.74 to 1.27) |
| 2013 | Viet Nam | 1.00 (0.74 to 1.30) |
| 2014 | Viet Nam | 1.01 (0.75 to 1.32) |
| 2015 | Viet Nam | 1.03 (0.76 to 1.34) |
| 2016 | Viet Nam | 1.03 (0.76 to 1.36) |
| 2017 | Viet Nam | 1.04 (0.76 to 1.39) |
| 2018 | Viet Nam | 1.05 (0.77 to 1.40) |
| 2019 | Viet Nam | 1.06 (0.79 to 1.41) |
| 2020 | Viet Nam | 1.07 (0.79 to 1.42) |
| 2021 | Viet Nam | 1.07 (0.78 to 1.42) |

#### Fig 4E. ASDR in East Asia from 1990 to 2021

| **Year** | **Location** | **ASDR per 100,000** |
| --- | --- | --- |
| 1990 | China | 0.33 (0.27 to 0.42) |
| 1991 | China | 0.34 (0.27 to 0.42) |
| 1992 | China | 0.34 (0.27 to 0.43) |
| 1993 | China | 0.35 (0.28 to 0.43) |
| 1994 | China | 0.35 (0.29 to 0.42) |
| 1995 | China | 0.35 (0.30 to 0.42) |
| 1996 | China | 0.35 (0.30 to 0.41) |
| 1997 | China | 0.35 (0.31 to 0.41) |
| 1998 | China | 0.35 (0.31 to 0.40) |
| 1999 | China | 0.35 (0.31 to 0.40) |
| 2000 | China | 0.35 (0.31 to 0.40) |
| 2001 | China | 0.35 (0.32 to 0.39) |
| 2002 | China | 0.36 (0.32 to 0.40) |
| 2003 | China | 0.36 (0.33 to 0.40) |
| 2004 | China | 0.36 (0.33 to 0.40) |
| 2005 | China | 0.37 (0.33 to 0.40) |
| 2006 | China | 0.38 (0.35 to 0.41) |
| 2007 | China | 0.39 (0.36 to 0.43) |
| 2008 | China | 0.41 (0.37 to 0.45) |
| 2009 | China | 0.43 (0.39 to 0.48) |
| 2010 | China | 0.45 (0.40 to 0.50) |
| 2011 | China | 0.45 (0.40 to 0.50) |
| 2012 | China | 0.44 (0.39 to 0.50) |
| 2013 | China | 0.44 (0.38 to 0.50) |
| 2014 | China | 0.44 (0.38 to 0.51) |
| 2015 | China | 0.44 (0.38 to 0.52) |
| 2016 | China | 0.45 (0.38 to 0.53) |
| 2017 | China | 0.45 (0.37 to 0.55) |
| 2018 | China | 0.46 (0.37 to 0.55) |
| 2019 | China | 0.46 (0.36 to 0.56) |
| 2020 | China | 0.46 (0.37 to 0.58) |
| 2021 | China | 0.46 (0.36 to 0.59) |
| 1990 | Democratic People's Republic of Korea | 0.55 (0.41 to 0.73) |
| 1991 | Democratic People's Republic of Korea | 0.55 (0.41 to 0.72) |
| 1992 | Democratic People's Republic of Korea | 0.55 (0.41 to 0.72) |
| 1993 | Democratic People's Republic of Korea | 0.55 (0.41 to 0.71) |
| 1994 | Democratic People's Republic of Korea | 0.54 (0.41 to 0.70) |
| 1995 | Democratic People's Republic of Korea | 0.54 (0.41 to 0.68) |
| 1996 | Democratic People's Republic of Korea | 0.54 (0.40 to 0.68) |
| 1997 | Democratic People's Republic of Korea | 0.53 (0.40 to 0.68) |
| 1998 | Democratic People's Republic of Korea | 0.53 (0.40 to 0.68) |
| 1999 | Democratic People's Republic of Korea | 0.52 (0.40 to 0.67) |
| 2000 | Democratic People's Republic of Korea | 0.52 (0.39 to 0.67) |
| 2001 | Democratic People's Republic of Korea | 0.52 (0.39 to 0.66) |
| 2002 | Democratic People's Republic of Korea | 0.52 (0.40 to 0.66) |
| 2003 | Democratic People's Republic of Korea | 0.52 (0.39 to 0.67) |
| 2004 | Democratic People's Republic of Korea | 0.53 (0.40 to 0.67) |
| 2005 | Democratic People's Republic of Korea | 0.53 (0.40 to 0.68) |
| 2006 | Democratic People's Republic of Korea | 0.54 (0.41 to 0.70) |
| 2007 | Democratic People's Republic of Korea | 0.55 (0.42 to 0.71) |
| 2008 | Democratic People's Republic of Korea | 0.55 (0.42 to 0.72) |
| 2009 | Democratic People's Republic of Korea | 0.56 (0.43 to 0.73) |
| 2010 | Democratic People's Republic of Korea | 0.57 (0.43 to 0.74) |
| 2011 | Democratic People's Republic of Korea | 0.56 (0.43 to 0.74) |
| 2012 | Democratic People's Republic of Korea | 0.56 (0.42 to 0.74) |
| 2013 | Democratic People's Republic of Korea | 0.56 (0.42 to 0.73) |
| 2014 | Democratic People's Republic of Korea | 0.56 (0.42 to 0.73) |
| 2015 | Democratic People's Republic of Korea | 0.56 (0.42 to 0.73) |
| 2016 | Democratic People's Republic of Korea | 0.56 (0.43 to 0.74) |
| 2017 | Democratic People's Republic of Korea | 0.55 (0.42 to 0.74) |
| 2018 | Democratic People's Republic of Korea | 0.55 (0.43 to 0.73) |
| 2019 | Democratic People's Republic of Korea | 0.55 (0.42 to 0.73) |
| 2020 | Democratic People's Republic of Korea | 0.55 (0.42 to 0.73) |
| 2021 | Democratic People's Republic of Korea | 0.55 (0.41 to 0.72) |

#### Fig 4F. ASDR in males and females from Asia in 2021

| **Sex** | **Location** | **ASDR per 100,000** |
| --- | --- | --- |
| Male | Central Asia | 3.08 (2.75 to 3.43) |
| Female | Central Asia | 1.22 (1.07 to 1.39) |
| Male | East Asia | 0.78 (0.58 to 1.00) |
| Female | East Asia | 0.27 (0.20 to 0.34) |
| Male | Japan | 6.61 (6.07 to 6.89) |
| Female | Japan | 3.72 (2.89 to 4.25) |
| Male | Republic of Korea | 2.48 (1.94 to 3.02) |
| Female | Republic of Korea | 1.50 (1.04 to 1.90) |
| Male | Singapore | 2.86 (2.56 to 3.13) |
| Female | Singapore | 1.55 (1.30 to 1.75) |
| Male | South Asia | 1.57 (1.01 to 2.63) |
| Female | South Asia | 0.90 (0.59 to 1.26) |
| Male | Southeast Asia | 2.00 (1.69 to 2.41) |
| Female | Southeast Asia | 0.92 (0.77 to 1.11) |

#### Fig 4G. ASDR in males and females from South Asia in 2021

| **Sex** | **Location** | **ASDR per 100,000** |
| --- | --- | --- |
| Female | Nepal | 1.04 (0.62 to 1.66) |
| Male | Nepal | 1.35 (0.75 to 2.59) |
| Female | Pakistan | 1.24 (0.77 to 1.79) |
| Male | Pakistan | 1.69 (1.06 to 2.77) |
| Female | Bhutan | 1.19 (0.77 to 1.63) |
| Male | Bhutan | 1.78 (0.81 to 3.14) |
| Female | Bangladesh | 0.92 (0.53 to 1.46) |
| Male | Bangladesh | 1.48 (0.82 to 2.94) |
| Female | India | 0.88 (0.55 to 1.23) |
| Male | India | 1.57 (0.95 to 2.61) |

#### Fig 4H. ASDR in males and females from Central Asia in 2021

| **Sex** | **Location** | **ASDR per 100,000** |
| --- | --- | --- |
| Male | Armenia | 13.82 (11.42 to 16.43) |
| Female | Armenia | 5.85 (4.76 to 7.20) |
| Male | Azerbaijan | 2.10 (1.27 to 3.34) |
| Female | Azerbaijan | 0.83 (0.53 to 1.31) |
| Male | Georgia | 5.57 (4.73 to 6.50) |
| Female | Georgia | 1.11 (0.88 to 1.49) |
| Male | Kazakhstan | 3.75 (3.02 to 4.56) |
| Female | Kazakhstan | 1.31 (1.02 to 1.63) |
| Male | Kyrgyzstan | 1.26 (1.00 to 1.54) |
| Female | Kyrgyzstan | 0.47 (0.38 to 0.57) |
| Male | Mongolia | 0.86 (0.55 to 1.28) |
| Female | Mongolia | 0.57 (0.41 to 0.77) |
| Male | Tajikistan | 0.43 (0.29 to 0.61) |
| Female | Tajikistan | 0.08 (0.06 to 0.11) |
| Male | Turkmenistan | 2.87 (2.13 to 4.30) |
| Female | Turkmenistan | 1.14 (0.81 to 1.51) |
| Male | Uzbekistan | 1.38 (1.12 to 1.68) |
| Female | Uzbekistan | 0.78 (0.61 to 0.98) |

#### Fig 4I. ASDR in males and females from Southeast Asia in 2021

| **Sex** | **Location** | **ASDR per 100,000** |
| --- | --- | --- |
| Male | Cambodia | 1.15 (0.67 to 2.07) |
| Female | Cambodia | 0.51 (0.31 to 0.80) |
| Male | Indonesia | 1.33 (0.74 to 1.96) |
| Female | Indonesia | 0.78 (0.49 to 1.17) |
| Male | Lao People's Democratic Republic | 1.25 (0.82 to 1.84) |
| Female | Lao People's Democratic Republic | 0.66 (0.47 to 0.94) |
| Male | Malaysia | 5.94 (4.63 to 7.45) |
| Female | Malaysia | 2.22 (1.74 to 2.73) |
| Male | Maldives | 0.81 (0.32 to 1.42) |
| Female | Maldives | 0.37 (0.24 to 0.54) |
| Male | Mauritius | 1.49 (1.36 to 1.59) |
| Female | Mauritius | 0.44 (0.39 to 0.49) |
| Male | Myanmar | 1.49 (1.02 to 2.10) |
| Female | Myanmar | 0.61 (0.44 to 0.84) |
| Male | Philippines | 1.88 (1.45 to 2.39) |
| Female | Philippines | 0.97 (0.76 to 1.21) |
| Male | Seychelles | 1.65 (0.91 to 2.51) |
| Female | Seychelles | 0.63 (0.42 to 0.88) |
| Male | Sri Lanka | 0.63 (0.41 to 0.90) |
| Female | Sri Lanka | 0.19 (0.12 to 0.27) |
| Male | Thailand | 2.82 (2.16 to 3.73) |
| Female | Thailand | 1.36 (0.98 to 1.82) |
| Male | Timor-Leste | 0.92 (0.55 to 1.64) |
| Female | Timor-Leste | 0.58 (0.38 to 0.91) |
| Male | Viet Nam | 1.80 (1.18 to 2.62) |
| Female | Viet Nam | 0.63 (0.46 to 0.85) |

#### Fig 4J. ASDR in males and females from GBD East Asia in 2021

| **Sex** | **Location** | **ASDR per 100,000** |
| --- | --- | --- |
| Male | China | 0.71 (0.51 to 0.94) |
| Female | China | 0.24 (0.18 to 0.32) |
| Male | Democratic People's Republic of Korea | 0.74 (0.53 to 1.03) |
| Female | Democratic People's Republic of Korea | 0.39 (0.27 to 0.53) |

### Data for Figure 5

#### Fig 5A. Global ASDR in males and females from 1990 to 2021

| **Year** | **Sex** | **ASDR per 100,000** |
| --- | --- | --- |
| 1990 | Female | 1.58 (1.41 to 1.76) |
| 1991 | Female | 1.60 (1.43 to 1.77) |
| 1992 | Female | 1.60 (1.43 to 1.77) |
| 1993 | Female | 1.62 (1.45 to 1.79) |
| 1994 | Female | 1.64 (1.47 to 1.81) |
| 1995 | Female | 1.64 (1.47 to 1.81) |
| 1996 | Female | 1.64 (1.45 to 1.80) |
| 1997 | Female | 1.61 (1.43 to 1.78) |
| 1998 | Female | 1.61 (1.43 to 1.77) |
| 1999 | Female | 1.65 (1.46 to 1.81) |
| 2000 | Female | 1.63 (1.44 to 1.78) |
| 2001 | Female | 1.57 (1.38 to 1.72) |
| 2002 | Female | 1.54 (1.36 to 1.69) |
| 2003 | Female | 1.53 (1.34 to 1.67) |
| 2004 | Female | 1.51 (1.32 to 1.65) |
| 2005 | Female | 1.49 (1.30 to 1.63) |
| 2006 | Female | 1.46 (1.27 to 1.60) |
| 2007 | Female | 1.44 (1.25 to 1.58) |
| 2008 | Female | 1.41 (1.23 to 1.55) |
| 2009 | Female | 1.39 (1.21 to 1.53) |
| 2010 | Female | 1.37 (1.19 to 1.51) |
| 2011 | Female | 1.36 (1.17 to 1.49) |
| 2012 | Female | 1.34 (1.16 to 1.47) |
| 2013 | Female | 1.33 (1.15 to 1.46) |
| 2014 | Female | 1.33 (1.15 to 1.46) |
| 2015 | Female | 1.33 (1.15 to 1.46) |
| 2016 | Female | 1.34 (1.15 to 1.46) |
| 2017 | Female | 1.33 (1.14 to 1.45) |
| 2018 | Female | 1.32 (1.13 to 1.44) |
| 2019 | Female | 1.31 (1.13 to 1.43) |
| 2020 | Female | 1.29 (1.09 to 1.41) |
| 2021 | Female | 1.28 (1.10 to 1.42) |
| 1990 | Male | 3.87 (3.61 to 4.18) |
| 1991 | Male | 3.89 (3.63 to 4.20) |
| 1992 | Male | 3.90 (3.64 to 4.21) |
| 1993 | Male | 3.95 (3.69 to 4.25) |
| 1994 | Male | 3.94 (3.68 to 4.24) |
| 1995 | Male | 3.94 (3.68 to 4.24) |
| 1996 | Male | 3.89 (3.64 to 4.19) |
| 1997 | Male | 3.81 (3.57 to 4.11) |
| 1998 | Male | 3.76 (3.52 to 4.06) |
| 1999 | Male | 3.74 (3.50 to 4.02) |
| 2000 | Male | 3.67 (3.42 to 3.95) |
| 2001 | Male | 3.56 (3.32 to 3.83) |
| 2002 | Male | 3.52 (3.28 to 3.79) |
| 2003 | Male | 3.46 (3.23 to 3.73) |
| 2004 | Male | 3.37 (3.14 to 3.64) |
| 2005 | Male | 3.32 (3.09 to 3.58) |
| 2006 | Male | 3.23 (3.00 to 3.48) |
| 2007 | Male | 3.16 (2.94 to 3.41) |
| 2008 | Male | 3.09 (2.87 to 3.34) |
| 2009 | Male | 3.00 (2.79 to 3.25) |
| 2010 | Male | 2.95 (2.73 to 3.19) |
| 2011 | Male | 2.87 (2.66 to 3.10) |
| 2012 | Male | 2.80 (2.59 to 3.03) |
| 2013 | Male | 2.75 (2.53 to 3.00) |
| 2014 | Male | 2.72 (2.52 to 2.97) |
| 2015 | Male | 2.71 (2.49 to 2.93) |
| 2016 | Male | 2.70 (2.48 to 2.92) |
| 2017 | Male | 2.66 (2.46 to 2.88) |
| 2018 | Male | 2.64 (2.44 to 2.85) |
| 2019 | Male | 2.62 (2.41 to 2.83) |
| 2020 | Male | 2.58 (2.37 to 2.80) |
| 2021 | Male | 2.57 (2.36 to 2.79) |

#### Fig 5B. Global death rate per 100,000 in males and females per age groups in 2021

| **Age** | **Sex** | **Death rate per 100,000** |
| --- | --- | --- |
| 15-19 years | Female | 0.02 (0.02 to 0.03) |
| 20-24 years | Female | 0.05 (0.04 to 0.06) |
| 25-29 years | Female | 0.06 (0.05 to 0.07) |
| 30-34 years | Female | 0.09 (0.08 to 0.10) |
| 35-39 years | Female | 0.15 (0.13 to 0.16) |
| 40-44 years | Female | 0.23 (0.21 to 0.26) |
| 45-49 years | Female | 0.39 (0.35 to 0.44) |
| 50-54 years | Female | 0.62 (0.56 to 0.70) |
| 55-59 years | Female | 1.14 (1.01 to 1.29) |
| 60-64 years | Female | 2.02 (1.83 to 2.30) |
| 65-69 years | Female | 3.51 (3.20 to 3.86) |
| 70-74 years | Female | 6.59 (5.92 to 7.36) |
| 75-79 years | Female | 11.39 (10.08 to 12.44) |
| 80-84 years | Female | 19.30 (16.14 to 21.35) |
| 85-89 years | Female | 34.09 (26.06 to 38.92) |
| 90-94 years | Female | 57.91 (41.84 to 66.94) |
| 95+ years | Female | 89.24 (60.11 to 105.90) |
| 15-19 years | Male | 0.04 (0.03 to 0.05) |
| 20-24 years | Male | 0.08 (0.07 to 0.09) |
| 25-29 years | Male | 0.14 (0.12 to 0.16) |
| 30-34 years | Male | 0.25 (0.22 to 0.29) |
| 35-39 years | Male | 0.45 (0.39 to 0.52) |
| 40-44 years | Male | 0.76 (0.68 to 0.87) |
| 45-49 years | Male | 1.18 (1.07 to 1.32) |
| 50-54 years | Male | 1.81 (1.66 to 2.01) |
| 55-59 years | Male | 3.08 (2.82 to 3.40) |
| 60-64 years | Male | 5.37 (5.01 to 5.90) |
| 65-69 years | Male | 8.67 (8.04 to 9.57) |
| 70-74 years | Male | 14.32 (13.43 to 15.78) |
| 75-79 years | Male | 22.24 (20.60 to 24.35) |
| 80-84 years | Male | 34.72 (30.99 to 37.91) |
| 85-89 years | Male | 57.94 (50.41 to 62.44) |
| 90-94 years | Male | 89.29 (75.27 to 97.35) |
| 95+ years | Male | 100.50 (76.52 to 112.24) |

#### Fig 5C. Percentage changes in AA-caused ASDR of men and women in 2021 compared to 1990

| **Four World Regions to % change (95%UI)** | |  |
| --- | --- | --- |
| **Location** | **Men** | **Women** |
| **Europe** | -34.5 (-40.3 to -28.2) | -17.9 (-30.9 to 0.7) |
| **America** | -54.6 (-58.8 to -50.0) | -36.2 (-47.0 to -23.1) |
| **Africa** | -12.0 (-52.6 to 140.7) | -33.3 (-65.7 to 19.0) |
| **Asia** | 65.3 (-30.5 to 203.1) | 58.9 (-22.9 to 188.5) |
| **USA and some European countries to % change (95%UI)** | |  |
| **Location** | **Men** | **Women** |
| **USA** | -68.0 (-71.1 to -64.4) | -49.5 (-58.7 to -38.0) |
| **UK** | -65.6 (-69.1 to -62.7) | -50.4 (-59.9 to -40.0) |
| **Sweden** | -59.7 (-67.7 to -51.1) | -24.2 (-41.7 to -1.8) |
| **Montenegro** | 32.0 (-34.0 to 155.7) | 23.8 (-23.2 to 97.7) |
| **Monaco** | -13.0 (-51.1 to 53.9) | -8.4 (-52.0 to 94.2) |
| **Norway** | -50.6 (-57.5 to -44.9) | -10.8 (-28.6 to 8.5) |
| **Denmark** | -40.9 (-51.4 to -29.0) | -9.0 (-28.5 to 15.0) |
| **Russia** | 44.5 (26.1 to 63.4) | 58.5 (36.0 to 90.1) |
| **Selected Asian Countries to % change (95%UI)** | | |
| **Location** | **Men** | **Women** |
| **Georgia** | 478.1 (322.5 to 672.4) | 181.5 (83.9 to 380.0) |
| **Armenia** | 128.6 (51.1 to 234.0) | 69.9 (5.7 to 169.6) |
| **India** | 103.6 (-45.9 to 511.5) | 51.9 (-53.1 to 343.0) |
| **China** | 71.6 (-10.5 to 241.5) | -8.8 (-52.6 to 67.5) |
| **Japan** | 44.7 (28.3 to 59.4) | 106.1 (49.6 to 167.9) |
| **Malaysia** | 24.8 (-26.4 to 113.0) | 11.5 (-27.8 to 68.4) |
| **South Korea** | 8.6 (-43.4 to 116.6) | -4.3 (-52.3 to 83.5) |
| **North Korea** | 2.0 (-48.6 to 106.8) | -10.8 (-53.9 to 74.6) |
| **Singapore** | -31.0 (-42.0 to -19.2) | 4.7 (-18.8 to 29.9) |
| **Top 10 countries with highest ASDR to % change (95%UI)** | | |
| **Location** | **Men** | **Women** |
| **Armenia** | 128.6 (51.1 to 234.0) | 69.9 (5.7 to 169.6) |
| **Montenegro** | 32.0 (-34.0 to 155.7) | 23.8 (-23.2 to 97.7) |
| **Nauru** | 4.5 (-50.0 to 139.3) | 52.7 (-45.3 to 270.9) |
| **Japan** | 44.7 (28.3 to 59.4) | 106.1 (49.6 to 167.9) |
| **Saint Lucia** | -45.9 (-61.2 to -29.6) | -57.2 (-68.9 to -43.6) |
| **Brunei Darussalam** | -3.1 (-47.0 to 90.7) | -16.6 (-47.8 to 46.4) |
| **Norway** | -50.6 (-57.5 to -44.9) | -10.8 (-28.6 to 8.5) |
| **Denmark** | -40.9 (-51.4 to -29.0) | -9.0 (-28.5 to 15.0) |
| **Monaco** | -13.0 (-51.1 to 53.9) | -8.4 (-52.0 to 94.2) |
| **Grenada** | -4.2 (-36.2 to 24.2) | 5.3 (-32.7 to 44.7) |

### Data for Figure 6

#### Fig 6A. Percentage contribution of risk factors to global AA-ASDR grouped by sex in 2021

| **Risk factor** | **Sex** | **Percent contribution to ASDR (%)** |
| --- | --- | --- |
| Diet high in sodium | Male | 1.01 (0.10 to 2.99) |
| Diet high in sodium | Female | 0.64 (0.03 to 2.26) |
| Diet high in sodium | Both | 0.87 (0.08 to 2.73) |
| Diet low in fruits | Male | 3.50 (2.41 to 4.65) |
| Diet low in fruits | Female | 3.76 (2.57 to 4.95) |
| Diet low in fruits | Both | 3.63 (2.48 to 4.78) |
| Diet low in vegetables | Male | 2.80 (1.88 to 3.97) |
| Diet low in vegetables | Female | 2.92 (1.94 to 4.16) |
| Diet low in vegetables | Both | 2.85 (1.90 to 4.02) |
| High body-mass index | Male | 6.99 (3.79 to 11.85) |
| High body-mass index | Female | 7.89 (4.26 to 13.30) |
| High body-mass index | Both | 7.39 (4.01 to 12.72) |
| High systolic blood pressure | Male | 16.80 (12.64 to 21.29) |
| High systolic blood pressure | Female | 17.94 (13.40 to 22.59) |
| High systolic blood pressure | Both | 17.32 (13.04 to 21.92) |
| Lead exposure | Male | 0.73 (-0.10 to 1.89) |
| Lead exposure | Female | 0.55 (-0.08 to 1.43) |
| Lead exposure | Both | 0.65 (-0.09 to 1.70) |
| Smoking | Male | 39.31 (33.55 to 45.51) |
| Smoking | Female | 14.16 (11.23 to 17.36) |
| Smoking | Both | 29.98 (25.47 to 34.85) |

#### Fig 6B. Percentage contribution of risk factors to AA-ASDR in world regions

Four world regions

| **Risk factor** | **Location** | **Percent contribution to ASDR (%)** |
| --- | --- | --- |
| Diet high in sodium | Africa | 0.45 (0.01 to 1.82) |
| Diet low in fruits | Africa | 4.45 (3.07 to 5.78) |
| Diet low in vegetables | Africa | 4.84 (3.31 to 6.65) |
| High body-mass index | Africa | 6.78 (3.81 to 11.57) |
| High systolic blood pressure | Africa | 19.23 (14.57 to 24.09) |
| Lead exposure | Africa | 0.87 (-0.12 to 2.19) |
| Smoking | Africa | 16.85 (13.72 to 20.35) |
| Diet high in sodium | America | 0.79 (0.03 to 2.70) |
| Diet low in fruits | America | 2.77 (1.90 to 3.60) |
| Diet low in vegetables | America | 3.60 (2.43 to 4.96) |
| High body-mass index | America | 9.91 (5.25 to 16.90) |
| High systolic blood pressure | America | 16.17 (11.81 to 20.63) |
| Lead exposure | America | 0.64 (-0.08 to 1.67) |
| Smoking | America | 33.05 (27.52 to 39.04) |
| Diet high in sodium | Asia | 1.12 (0.12 to 3.26) |
| Diet low in fruits | Asia | 4.03 (2.72 to 5.37) |
| Diet low in vegetables | Asia | 2.32 (1.49 to 3.31) |
| High body-mass index | Asia | 4.40 (2.40 to 7.04) |
| High systolic blood pressure | Asia | 16.54 (12.33 to 21.47) |
| Lead exposure | Asia | 0.77 (-0.11 to 2.00) |
| Smoking | Asia | 26.82 (22.73 to 31.41) |
| Diet high in sodium | Europe | 0.70 (0.06 to 2.31) |
| Diet low in fruits | Europe | 3.53 (2.46 to 4.67) |
| Diet low in vegetables | Europe | 2.65 (1.72 to 3.81) |
| High body-mass index | Europe | 9.95 (5.27 to 17.18) |
| High systolic blood pressure | Europe | 18.69 (14.18 to 23.62) |
| Lead exposure | Europe | 0.46 (-0.06 to 1.16) |
| Smoking | Europe | 37.24 (31.83 to 42.89) |

Health System Grouping

| **Risk factor** | **Location** | **Percent contribution to ASDR (%)** |
| --- | --- | --- |
| Lead exposure | Advanced Health System | 0.44 (-0.06 to 1.15) |
| Smoking | Advanced Health System | 33.93 (28.57 to 39.27) |
| Diet low in fruits | Advanced Health System | 3.53 (2.42 to 4.65) |
| Diet low in vegetables | Advanced Health System | 2.33 (1.50 to 3.38) |
| Diet high in sodium | Advanced Health System | 0.80 (0.05 to 2.63) |
| High systolic blood pressure | Advanced Health System | 17.35 (13.00 to 22.07) |
| High body-mass index | Advanced Health System | 8.56 (4.57 to 14.71) |
| Lead exposure | Basic Health System | 0.80 (-0.11 to 2.08) |
| Smoking | Basic Health System | 33.68 (28.72 to 38.74) |
| Diet low in fruits | Basic Health System | 2.98 (2.05 to 3.98) |
| Diet low in vegetables | Basic Health System | 3.14 (2.12 to 4.36) |
| Diet high in sodium | Basic Health System | 1.21 (0.17 to 3.27) |
| High systolic blood pressure | Basic Health System | 17.65 (13.06 to 22.15) |
| High body-mass index | Basic Health System | 7.52 (4.07 to 12.76) |
| Lead exposure | Limited Health System | 1.26 (-0.17 to 3.20) |
| Smoking | Limited Health System | 23.13 (18.32 to 29.19) |
| Diet low in fruits | Limited Health System | 5.17 (3.57 to 6.82) |
| Diet low in vegetables | Limited Health System | 4.41 (3.01 to 6.07) |
| Diet high in sodium | Limited Health System | 0.69 (0.02 to 2.48) |
| High systolic blood pressure | Limited Health System | 17.03 (12.80 to 21.57) |
| High body-mass index | Limited Health System | 4.02 (2.26 to 6.55) |
| Smoking | Minimal Health System | 12.47 (7.19 to 17.54) |
| Diet low in fruits | Minimal Health System | 5.03 (3.50 to 6.63) |
| Diet low in vegetables | Minimal Health System | 5.53 (3.80 to 7.58) |
| Diet high in sodium | Minimal Health System | 0.43 (0.00 to 1.87) |
| High systolic blood pressure | Minimal Health System | 17.93 (13.31 to 22.74) |
| High body-mass index | Minimal Health System | 4.25 (2.37 to 7.09) |

Income

| **Risk factor** | **Location** | **Percent contribution to ASDR (%)** |
| --- | --- | --- |
| Diet high in sodium | Commonwealth High Income | 0.52 (0.01 to 2.20) |
| Diet low in fruits | Commonwealth High Income | 3.44 (2.39 to 4.52) |
| Diet low in vegetables | Commonwealth High Income | 2.99 (1.97 to 4.22) |
| High body-mass index | Commonwealth High Income | 8.89 (4.67 to 15.19) |
| High systolic blood pressure | Commonwealth High Income | 15.02 (11.15 to 18.97) |
| Lead exposure | Commonwealth High Income | 0.50 (-0.06 to 1.28) |
| Smoking | Commonwealth High Income | 30.56 (24.71 to 37.65) |
| Diet high in sodium | Commonwealth Low Income | 0.75 (0.02 to 2.52) |
| Diet low in fruits | Commonwealth Low Income | 4.59 (3.16 to 6.06) |
| Diet low in vegetables | Commonwealth Low Income | 5.24 (3.58 to 7.17) |
| High body-mass index | Commonwealth Low Income | 3.50 (1.90 to 5.64) |
| High systolic blood pressure | Commonwealth Low Income | 17.43 (13.13 to 22.23) |
| Lead exposure | Commonwealth Low Income | 1.25 (-0.17 to 3.23) |
| Smoking | Commonwealth Low Income | 27.63 (21.79 to 34.58) |
| Diet high in sodium | Commonwealth Middle Income | 0.66 (0.03 to 2.39) |
| Diet low in fruits | Commonwealth Middle Income | 5.28 (3.67 to 6.96) |
| Diet low in vegetables | Commonwealth Middle Income | 4.29 (2.91 to 5.94) |
| High body-mass index | Commonwealth Middle Income | 4.53 (2.54 to 7.53) |
| High systolic blood pressure | Commonwealth Middle Income | 17.57 (13.21 to 22.12) |
| Lead exposure | Commonwealth Middle Income | 1.18 (-0.16 to 3.04) |
| Smoking | Commonwealth Middle Income | 22.67 (17.78 to 28.48) |

SDI

| **Risk factor** | **Location** | **Percent contribution to ASDR (%)** |
| --- | --- | --- |
| Diet high in sodium | High SDI | 0.86 (0.06 to 2.81) |
| Diet low in fruits | High SDI | 3.56 (2.46 to 4.69) |
| Diet low in vegetables | High SDI | 2.32 (1.50 to 3.37) |
| High body-mass index | High SDI | 7.60 (4.09 to 12.96) |
| High systolic blood pressure | High SDI | 16.52 (12.20 to 21.03) |
| Lead exposure | High SDI | 0.44 (-0.06 to 1.15) |
| Smoking | High SDI | 31.74 (26.59 to 37.29) |
| Diet high in sodium | High-middle SDI | 0.92 (0.12 to 2.60) |
| Diet low in fruits | High-middle SDI | 3.32 (2.31 to 4.39) |
| Diet low in vegetables | High-middle SDI | 2.16 (1.39 to 3.14) |
| High body-mass index | High-middle SDI | 9.71 (5.17 to 16.71) |
| High systolic blood pressure | High-middle SDI | 18.75 (14.11 to 23.67) |
| Lead exposure | High-middle SDI | 0.55 (-0.07 to 1.41) |
| Smoking | High-middle SDI | 37.21 (32.14 to 41.99) |
| Diet high in sodium | Low SDI | 0.62 (0.01 to 2.32) |
| Diet low in fruits | Low SDI | 4.67 (3.24 to 6.17) |
| Diet low in vegetables | Low SDI | 4.95 (3.38 to 6.77) |
| High body-mass index | Low SDI | 3.74 (2.05 to 6.15) |
| High systolic blood pressure | Low SDI | 16.60 (12.38 to 20.83) |
| Lead exposure | Low SDI | 1.32 (-0.18 to 3.29) |
| Smoking | Low SDI | 18.02 (13.92 to 23.13) |
| Diet high in sodium | Low-middle SDI | 0.74 (0.04 to 2.53) |
| Diet low in fruits | Low-middle SDI | 4.70 (3.23 to 6.27) |
| Diet low in vegetables | Low-middle SDI | 4.41 (3.02 to 6.10) |
| High body-mass index | Low-middle SDI | 5.01 (2.74 to 8.30) |
| High systolic blood pressure | Low-middle SDI | 17.43 (13.19 to 22.10) |
| Lead exposure | Low-middle SDI | 1.28 (-0.18 to 3.27) |
| Smoking | Low-middle SDI | 26.69 (21.51 to 33.19) |
| Diet high in sodium | Middle SDI | 1.04 (0.12 to 2.97) |
| Diet low in fruits | Middle SDI | 3.38 (2.28 to 4.44) |
| Diet low in vegetables | Middle SDI | 3.61 (2.45 to 4.98) |
| High body-mass index | Middle SDI | 6.96 (3.76 to 11.86) |
| High systolic blood pressure | Middle SDI | 17.51 (13.14 to 22.19) |
| Lead exposure | Middle SDI | 0.81 (-0.11 to 2.09) |
| Smoking | Middle SDI | 29.94 (25.48 to 34.57) |

#### Fig 6C. Percentage contribution of risk factors to AA-ASDR in world regions grouped by sex in 2021

Four World Regions

| **Risk factor** | **Sex** | **Location** | **Percent contribution to ASDR (%)** |
| --- | --- | --- | --- |
| Diet high in sodium | Male | Africa | 0.44 (0.01 to 1.85) |
| Diet high in sodium | Female | Africa | 0.46 (0.01 to 1.78) |
| Diet low in fruits | Male | Africa | 4.24 (2.93 to 5.50) |
| Diet low in fruits | Female | Africa | 4.74 (3.31 to 6.24) |
| Diet low in vegetables | Male | Africa | 4.62 (3.17 to 6.38) |
| Diet low in vegetables | Female | Africa | 5.14 (3.49 to 7.06) |
| High body-mass index | Male | Africa | 6.16 (3.39 to 10.68) |
| High body-mass index | Female | Africa | 7.76 (4.24 to 12.95) |
| High systolic blood pressure | Male | Africa | 18.26 (13.64 to 22.87) |
| High systolic blood pressure | Female | Africa | 20.66 (15.65 to 25.75) |
| Lead exposure | Male | Africa | 0.99 (-0.14 to 2.52) |
| Lead exposure | Female | Africa | 0.68 (-0.10 to 1.81) |
| Smoking | Male | Africa | 24.33 (19.73 to 29.23) |
| Smoking | Female | Africa | 5.18 (3.93 to 6.62) |
| Diet high in sodium | Male | America | 0.96 (0.05 to 3.10) |
| Diet high in sodium | Female | America | 0.51 (0.00 to 2.07) |
| Diet low in fruits | Male | America | 2.68 (1.86 to 3.51) |
| Diet low in fruits | Female | America | 2.87 (1.91 to 3.80) |
| Diet low in vegetables | Male | America | 3.60 (2.43 to 5.06) |
| Diet low in vegetables | Female | America | 3.58 (2.40 to 4.98) |
| High body-mass index | Male | America | 9.55 (5.07 to 16.39) |
| High body-mass index | Female | America | 10.34 (5.44 to 17.92) |
| High systolic blood pressure | Male | America | 15.59 (11.50 to 19.62) |
| High systolic blood pressure | Female | America | 16.94 (12.39 to 21.81) |
| Lead exposure | Male | America | 0.71 (-0.09 to 1.82) |
| Lead exposure | Female | America | 0.55 (-0.07 to 1.44) |
| Smoking | Male | America | 37.80 (31.46 to 44.56) |
| Smoking | Female | America | 24.91 (20.22 to 30.08) |
| Diet high in sodium | Male | Asia | 1.29 (0.15 to 3.63) |
| Diet high in sodium | Female | Asia | 0.88 (0.06 to 2.76) |
| Diet low in fruits | Male | Asia | 3.97 (2.71 to 5.29) |
| Diet low in fruits | Female | Asia | 4.08 (2.76 to 5.46) |
| Diet low in vegetables | Male | Asia | 2.26 (1.49 to 3.34) |
| Diet low in vegetables | Female | Asia | 2.39 (1.48 to 3.47) |
| High body-mass index | Male | Asia | 4.10 (2.27 to 6.66) |
| High body-mass index | Female | Asia | 4.73 (2.57 to 7.72) |
| High systolic blood pressure | Male | Asia | 16.20 (12.18 to 20.76) |
| High systolic blood pressure | Female | Asia | 16.96 (12.35 to 22.07) |
| Lead exposure | Male | Asia | 0.87 (-0.11 to 2.21) |
| Lead exposure | Female | Asia | 0.63 (-0.09 to 1.67) |
| Smoking | Male | Asia | 39.54 (33.31 to 45.56) |
| Smoking | Female | Asia | 8.83 (6.81 to 11.46) |
| Diet high in sodium | Male | Europe | 0.83 (0.07 to 2.58) |
| Diet high in sodium | Female | Europe | 0.43 (0.02 to 1.76) |
| Diet low in fruits | Male | Europe | 3.38 (2.38 to 4.49) |
| Diet low in fruits | Female | Europe | 3.72 (2.60 to 4.88) |
| Diet low in vegetables | Male | Europe | 2.61 (1.71 to 3.77) |
| Diet low in vegetables | Female | Europe | 2.71 (1.76 to 3.88) |
| High body-mass index | Male | Europe | 9.29 (4.95 to 16.17) |
| High body-mass index | Female | Europe | 10.98 (5.77 to 18.87) |
| High systolic blood pressure | Male | Europe | 18.13 (13.72 to 23.03) |
| High systolic blood pressure | Female | Europe | 19.34 (14.58 to 24.22) |
| Lead exposure | Male | Europe | 0.51 (-0.07 to 1.30) |
| Lead exposure | Female | Europe | 0.37 (-0.05 to 0.94) |
| Smoking | Male | Europe | 45.15 (38.60 to 51.90) |
| Smoking | Female | Europe | 18.72 (15.34 to 22.36) |

Health System Grouping

| **Risk factor** | **Sex** | **Location** | **Percent contribution to ASDR (%)** |
| --- | --- | --- | --- |
| Diet high in sodium | Male | Advanced Health System | 0.93 (0.07 to 2.90) |
| Diet high in sodium | Female | Advanced Health System | 0.58 (0.02 to 2.17) |
| Diet low in fruits | Male | Advanced Health System | 3.42 (2.36 to 4.56) |
| Diet low in fruits | Female | Advanced Health System | 3.62 (2.49 to 4.73) |
| Diet low in vegetables | Male | Advanced Health System | 2.32 (1.49 to 3.39) |
| Diet low in vegetables | Female | Advanced Health System | 2.32 (1.49 to 3.38) |
| High body-mass index | Male | Advanced Health System | 8.26 (4.45 to 14.08) |
| High body-mass index | Female | Advanced Health System | 8.83 (4.72 to 15.09) |
| High systolic blood pressure | Male | Advanced Health System | 16.95 (12.71 to 21.75) |
| High systolic blood pressure | Female | Advanced Health System | 17.77 (13.08 to 22.52) |
| Lead exposure | Male | Advanced Health System | 0.49 (-0.06 to 1.28) |
| Lead exposure | Female | Advanced Health System | 0.37 (-0.05 to 0.97) |
| Smoking | Male | Advanced Health System | 42.58 (36.35 to 49.25) |
| Smoking | Female | Advanced Health System | 17.83 (14.41 to 21.44) |
| Diet high in sodium | Male | Basic Health System | 1.39 (0.22 to 3.63) |
| Diet high in sodium | Female | Basic Health System | 0.84 (0.07 to 2.56) |
| Diet low in fruits | Male | Basic Health System | 2.92 (2.01 to 3.88) |
| Diet low in fruits | Female | Basic Health System | 3.05 (2.07 to 4.09) |
| Diet low in vegetables | Male | Basic Health System | 3.02 (2.06 to 4.20) |
| Diet low in vegetables | Female | Basic Health System | 3.35 (2.24 to 4.70) |
| High body-mass index | Male | Basic Health System | 6.89 (3.71 to 11.59) |
| High body-mass index | Female | Basic Health System | 8.53 (4.57 to 14.81) |
| High systolic blood pressure | Male | Basic Health System | 17.18 (12.67 to 21.74) |
| High systolic blood pressure | Female | Basic Health System | 18.34 (13.79 to 23.24) |
| Lead exposure | Male | Basic Health System | 0.89 (-0.12 to 2.29) |
| Lead exposure | Female | Basic Health System | 0.66 (-0.09 to 1.72) |
| Smoking | Male | Basic Health System | 43.05 (37.13 to 48.85) |
| Smoking | Female | Basic Health System | 15.26 (12.27 to 18.33) |
| Diet high in sodium | Male | Limited Health System | 0.78 (0.03 to 2.65) |
| Diet high in sodium | Female | Limited Health System | 0.56 (0.01 to 2.16) |
| Diet low in fruits | Male | Limited Health System | 5.04 (3.48 to 6.69) |
| Diet low in fruits | Female | Limited Health System | 5.33 (3.69 to 7.01) |
| Diet low in vegetables | Male | Limited Health System | 4.28 (2.91 to 5.86) |
| Diet low in vegetables | Female | Limited Health System | 4.59 (3.12 to 6.32) |
| High body-mass index | Male | Limited Health System | 3.57 (2.00 to 5.89) |
| High body-mass index | Female | Limited Health System | 4.73 (2.59 to 8.06) |
| High systolic blood pressure | Male | Limited Health System | 16.31 (12.14 to 20.83) |
| High systolic blood pressure | Female | Limited Health System | 18.13 (13.62 to 22.65) |
| Lead exposure | Male | Limited Health System | 1.37 (-0.18 to 3.48) |
| Lead exposure | Female | Limited Health System | 1.08 (-0.15 to 2.74) |
| Smoking | Male | Limited Health System | 32.61 (26.92 to 39.26) |
| Smoking | Female | Limited Health System | 7.85 (6.04 to 10.19) |
| Diet high in sodium | Male | Minimal Health System | 0.45 (0.00 to 1.97) |
| Diet high in sodium | Female | Minimal Health System | 0.41 (0.00 to 1.74) |
| Diet low in fruits | Male | Minimal Health System | 4.87 (3.38 to 6.42) |
| Diet low in fruits | Female | Minimal Health System | 5.18 (3.61 to 6.85) |
| Diet low in vegetables | Male | Minimal Health System | 5.41 (3.72 to 7.36) |
| Diet low in vegetables | Female | Minimal Health System | 5.65 (3.84 to 7.73) |
| High body-mass index | Male | Minimal Health System | 3.80 (2.09 to 6.22) |
| High body-mass index | Female | Minimal Health System | 4.75 (2.63 to 8.04) |
| High systolic blood pressure | Male | Minimal Health System | 16.98 (12.40 to 21.13) |
| High systolic blood pressure | Female | Minimal Health System | 18.92 (14.09 to 24.00) |
| Lead exposure | Male | Minimal Health System | 1.37 (-0.20 to 3.37) |
| Lead exposure | Female | Minimal Health System | 0.95 (-0.13 to 2.42) |
| Smoking | Male | Minimal Health System | 20.58 (16.80 to 24.76) |
| Smoking | Female | Minimal Health System | 2.75 (2.11 to 3.50) |

Income

| **Risk factor** | **Sex** | **Location** | **Percent contribution to ASDR (%)** |
| --- | --- | --- | --- |
| Diet high in sodium | Male | Commonwealth High Income | 0.63 (0.01 to 2.45) |
| Diet high in sodium | Female | Commonwealth High Income | 0.34 (0.00 to 1.73) |
| Diet low in fruits | Male | Commonwealth High Income | 3.36 (2.33 to 4.50) |
| Diet low in fruits | Female | Commonwealth High Income | 3.53 (2.46 to 4.69) |
| Diet low in vegetables | Male | Commonwealth High Income | 2.99 (1.99 to 4.24) |
| Diet low in vegetables | Female | Commonwealth High Income | 2.97 (1.94 to 4.24) |
| High body-mass index | Male | Commonwealth High Income | 8.67 (4.51 to 15.06) |
| High body-mass index | Female | Commonwealth High Income | 9.10 (4.75 to 15.51) |
| High systolic blood pressure | Male | Commonwealth High Income | 14.42 (10.59 to 18.48) |
| High systolic blood pressure | Female | Commonwealth High Income | 15.82 (11.45 to 20.20) |
| Lead exposure | Male | Commonwealth High Income | 0.56 (-0.07 to 1.46) |
| Lead exposure | Female | Commonwealth High Income | 0.41 (-0.05 to 1.06) |
| Smoking | Male | Commonwealth High Income | 33.92 (27.47 to 41.57) |
| Smoking | Female | Commonwealth High Income | 24.36 (19.03 to 30.47) |
| Diet high in sodium | Male | Commonwealth Low Income | 0.74 (0.01 to 2.50) |
| Diet high in sodium | Female | Commonwealth Low Income | 0.77 (0.02 to 2.60) |
| Diet low in fruits | Male | Commonwealth Low Income | 4.42 (3.05 to 5.91) |
| Diet low in fruits | Female | Commonwealth Low Income | 4.84 (3.30 to 6.54) |
| Diet low in vegetables | Male | Commonwealth Low Income | 5.04 (3.42 to 6.84) |
| Diet low in vegetables | Female | Commonwealth Low Income | 5.55 (3.73 to 7.68) |
| High body-mass index | Male | Commonwealth Low Income | 2.85 (1.55 to 4.68) |
| High body-mass index | Female | Commonwealth Low Income | 4.55 (2.43 to 7.70) |
| High systolic blood pressure | Male | Commonwealth Low Income | 15.96 (11.67 to 20.38) |
| High systolic blood pressure | Female | Commonwealth Low Income | 19.74 (14.91 to 25.24) |
| Lead exposure | Male | Commonwealth Low Income | 1.48 (-0.20 to 3.86) |
| Lead exposure | Female | Commonwealth Low Income | 0.89 (-0.13 to 2.32) |
| Smoking | Male | Commonwealth Low Income | 39.50 (33.38 to 46.24) |
| Smoking | Female | Commonwealth Low Income | 8.37 (6.28 to 10.40) |
| Diet high in sodium | Male | Commonwealth Middle Income | 0.77 (0.04 to 2.63) |
| Diet high in sodium | Female | Commonwealth Middle Income | 0.48 (0.00 to 1.96) |
| Diet low in fruits | Male | Commonwealth Middle Income | 5.14 (3.58 to 6.82) |
| Diet low in fruits | Female | Commonwealth Middle Income | 5.47 (3.79 to 7.21) |
| Diet low in vegetables | Male | Commonwealth Middle Income | 4.18 (2.85 to 5.81) |
| Diet low in vegetables | Female | Commonwealth Middle Income | 4.47 (3.02 to 6.16) |
| High body-mass index | Male | Commonwealth Middle Income | 4.04 (2.22 to 6.75) |
| High body-mass index | Female | Commonwealth Middle Income | 5.34 (2.90 to 9.18) |
| High systolic blood pressure | Male | Commonwealth Middle Income | 17.01 (12.70 to 21.73) |
| High systolic blood pressure | Female | Commonwealth Middle Income | 18.48 (13.74 to 23.30) |
| Lead exposure | Male | Commonwealth Middle Income | 1.26 (-0.17 to 3.22) |
| Lead exposure | Female | Commonwealth Middle Income | 1.06 (-0.15 to 2.72) |
| Smoking | Male | Commonwealth Middle Income | 31.64 (25.94 to 38.37) |
| Smoking | Female | Commonwealth Middle Income | 7.67 (5.84 to 10.02) |

SDI

| **Risk factor** | **Sex** | **Location** | **Percent contribution to ADR (%)** |
| --- | --- | --- | --- |
| Diet high in sodium | Male | High SDI | 0.99 (0.07 to 3.11) |
| Diet high in sodium | Female | High SDI | 0.65 (0.03 to 2.35) |
| Diet low in fruits | Male | High SDI | 3.51 (2.44 to 4.63) |
| Diet low in fruits | Female | High SDI | 3.58 (2.46 to 4.70) |
| Diet low in vegetables | Male | High SDI | 2.33 (1.53 to 3.39) |
| Diet low in vegetables | Female | High SDI | 2.30 (1.46 to 3.34) |
| High body-mass index | Male | High SDI | 7.48 (4.04 to 12.70) |
| High body-mass index | Female | High SDI | 7.60 (4.07 to 12.83) |
| High systolic blood pressure | Male | High SDI | 16.10 (12.03 to 20.76) |
| High systolic blood pressure | Female | High SDI | 17.01 (12.40 to 21.91) |
| Lead exposure | Male | High SDI | 0.48 (-0.06 to 1.28) |
| Lead exposure | Female | High SDI | 0.38 (-0.05 to 0.98) |
| Smoking | Male | High SDI | 39.44 (33.05 to 46.18) |
| Smoking | Female | High SDI | 18.93 (15.01 to 23.10) |
| Diet high in sodium | Male | High-middle SDI | 1.05 (0.15 to 2.85) |
| Diet high in sodium | Female | High-middle SDI | 0.59 (0.05 to 1.94) |
| Diet low in fruits | Male | High-middle SDI | 3.12 (2.19 to 4.14) |
| Diet low in fruits | Female | High-middle SDI | 3.57 (2.45 to 4.73) |
| Diet low in vegetables | Male | High-middle SDI | 2.13 (1.37 to 3.08) |
| Diet low in vegetables | Female | High-middle SDI | 2.24 (1.42 to 3.28) |
| High body-mass index | Male | High-middle SDI | 8.81 (4.68 to 15.31) |
| High body-mass index | Female | High-middle SDI | 11.26 (5.98 to 19.23) |
| High systolic blood pressure | Male | High-middle SDI | 18.04 (13.55 to 22.66) |
| High systolic blood pressure | Female | High-middle SDI | 19.75 (14.97 to 24.57) |
| Lead exposure | Male | High-middle SDI | 0.62 (-0.08 to 1.60) |
| Lead exposure | Female | High-middle SDI | 0.43 (-0.06 to 1.10) |
| Smoking | Male | High-middle SDI | 47.19 (40.93 to 53.06) |
| Smoking | Female | High-middle SDI | 13.35 (10.85 to 15.87) |
| Diet high in sodium | Male | Low SDI | 0.65 (0.02 to 2.35) |
| Diet high in sodium | Female | Low SDI | 0.60 (0.01 to 2.29) |
| Diet low in fruits | Male | Low SDI | 4.55 (3.15 to 6.03) |
| Diet low in fruits | Female | Low SDI | 4.82 (3.36 to 6.38) |
| Diet low in vegetables | Male | Low SDI | 4.85 (3.30 to 6.69) |
| Diet low in vegetables | Female | Low SDI | 5.07 (3.48 to 6.89) |
| High body-mass index | Male | Low SDI | 3.36 (1.87 to 5.48) |
| High body-mass index | Female | Low SDI | 4.23 (2.31 to 7.15) |
| High systolic blood pressure | Male | Low SDI | 15.87 (11.79 to 20.03) |
| High systolic blood pressure | Female | Low SDI | 17.49 (13.12 to 22.07) |
| Lead exposure | Male | Low SDI | 1.45 (-0.20 to 3.65) |
| Lead exposure | Female | Low SDI | 1.14 (-0.16 to 2.91) |
| Smoking | Male | Low SDI | 25.29 (20.88 to 30.31) |
| Smoking | Female | Low SDI | 8.03 (5.97 to 10.57) |
| Diet high in sodium | Male | Low-middle SDI | 0.85 (0.05 to 2.76) |
| Diet high in sodium | Female | Low-middle SDI | 0.56 (0.01 to 2.17) |
| Diet low in fruits | Male | Low-middle SDI | 4.57 (3.14 to 6.08) |
| Diet low in fruits | Female | Low-middle SDI | 4.86 (3.34 to 6.49) |
| Diet low in vegetables | Male | Low-middle SDI | 4.27 (2.92 to 5.89) |
| Diet low in vegetables | Female | Low-middle SDI | 4.61 (3.12 to 6.38) |
| High body-mass index | Male | Low-middle SDI | 4.52 (2.50 to 7.63) |
| High body-mass index | Female | Low-middle SDI | 5.77 (3.13 to 10.05) |
| High systolic blood pressure | Male | Low-middle SDI | 16.63 (12.40 to 21.23) |
| High systolic blood pressure | Female | Low-middle SDI | 18.60 (14.12 to 23.26) |
| Lead exposure | Male | Low-middle SDI | 1.41 (-0.19 to 3.60) |
| Lead exposure | Female | Low-middle SDI | 1.08 (-0.16 to 2.74) |
| Smoking | Male | Low-middle SDI | 37.32 (31.30 to 44.46) |
| Smoking | Female | Low-middle SDI | 10.10 (7.89 to 12.78) |
| Diet high in sodium | Male | Middle SDI | 1.22 (0.16 to 3.33) |
| Diet high in sodium | Female | Middle SDI | 0.73 (0.05 to 2.39) |
| Diet low in fruits | Male | Middle SDI | 3.31 (2.26 to 4.38) |
| Diet low in fruits | Female | Middle SDI | 3.46 (2.32 to 4.67) |
| Diet low in vegetables | Male | Middle SDI | 3.51 (2.39 to 4.83) |
| Diet low in vegetables | Female | Middle SDI | 3.77 (2.56 to 5.25) |
| High body-mass index | Male | Middle SDI | 6.37 (3.44 to 10.68) |
| High body-mass index | Female | Middle SDI | 7.87 (4.29 to 13.43) |
| High systolic blood pressure | Male | Middle SDI | 17.10 (12.74 to 21.52) |
| High systolic blood pressure | Female | Middle SDI | 18.08 (13.47 to 22.96) |
| Lead exposure | Male | Middle SDI | 0.90 (-0.12 to 2.31) |
| Lead exposure | Female | Middle SDI | 0.67 (-0.09 to 1.76) |
| Smoking | Male | Middle SDI | 39.15 (33.23 to 44.76) |
| Smoking | Female | Middle SDI | 13.31 (10.65 to 16.09) |

#### Fig 6D. Percentage contribution of risk factors to AA-ASDR in high-burden countries in 2021

| **Risk factor** | **Sex** | **Location** | **Percent contribution to ASDR (%)** |
| --- | --- | --- | --- |
| Diet high in sodium | Male | Armenia | 0.98 (0.05 to 3.12) |
| Diet high in sodium | Female | Armenia | 0.58 (0.00 to 2.35) |
| Diet low in fruits | Male | Armenia | 2.29 (1.46 to 3.27) |
| Diet low in fruits | Female | Armenia | 2.50 (1.54 to 3.63) |
| Diet low in vegetables | Male | Armenia | 0.09 (0.01 to 0.28) |
| Diet low in vegetables | Female | Armenia | 0.11 (0.00 to 0.42) |
| High body-mass index | Male | Armenia | 8.01 (4.35 to 13.88) |
| High body-mass index | Female | Armenia | 12.19 (6.30 to 20.94) |
| High systolic blood pressure | Male | Armenia | 19.62 (14.30 to 25.63) |
| High systolic blood pressure | Female | Armenia | 20.10 (13.79 to 28.11) |
| Lead exposure | Male | Armenia | 0.60 (-0.08 to 1.61) |
| Lead exposure | Female | Armenia | 0.47 (-0.06 to 1.32) |
| Smoking | Male | Armenia | 60.34 (53.32 to 66.51) |
| Smoking | Female | Armenia | 4.27 (2.95 to 6.17) |
| Diet high in sodium | Male | Brunei Darussalam | 1.59 (0.16 to 4.29) |
| Diet high in sodium | Female | Brunei Darussalam | 1.21 (0.05 to 3.67) |
| Diet low in fruits | Male | Brunei Darussalam | 4.35 (3.01 to 5.85) |
| Diet low in fruits | Female | Brunei Darussalam | 4.44 (2.99 to 6.11) |
| Diet low in vegetables | Male | Brunei Darussalam | 4.13 (2.76 to 5.80) |
| Diet low in vegetables | Female | Brunei Darussalam | 4.10 (2.65 to 5.89) |
| High body-mass index | Male | Brunei Darussalam | 4.46 (2.43 to 7.53) |
| High body-mass index | Female | Brunei Darussalam | 4.86 (2.65 to 8.18) |
| High systolic blood pressure | Male | Brunei Darussalam | 15.41 (10.76 to 20.77) |
| High systolic blood pressure | Female | Brunei Darussalam | 15.91 (9.94 to 22.52) |
| Lead exposure | Male | Brunei Darussalam | 0.72 (-0.09 to 1.94) |
| Lead exposure | Female | Brunei Darussalam | 0.52 (-0.07 to 1.44) |
| Smoking | Male | Brunei Darussalam | 36.38 (28.77 to 44.41) |
| Smoking | Female | Brunei Darussalam | 13.31 (9.37 to 18.27) |
| Diet high in sodium | Male | Denmark | 0.57 (0.00 to 2.21) |
| Diet high in sodium | Female | Denmark | 0.28 (0.00 to 1.49) |
| Diet low in fruits | Male | Denmark | 2.60 (1.60 to 3.74) |
| Diet low in fruits | Female | Denmark | 2.92 (1.84 to 4.40) |
| Diet low in vegetables | Male | Denmark | 2.94 (1.84 to 4.41) |
| Diet low in vegetables | Female | Denmark | 3.05 (1.81 to 4.77) |
| High body-mass index | Male | Denmark | 7.53 (3.89 to 13.06) |
| High body-mass index | Female | Denmark | 8.50 (4.50 to 14.93) |
| High systolic blood pressure | Male | Denmark | 19.13 (13.47 to 25.36) |
| High systolic blood pressure | Female | Denmark | 21.07 (13.89 to 29.87) |
| Lead exposure | Male | Denmark | 0.41 (-0.06 to 1.12) |
| Lead exposure | Female | Denmark | 0.36 (-0.05 to 0.93) |
| Smoking | Male | Denmark | 44.89 (36.72 to 52.97) |
| Smoking | Female | Denmark | 33.35 (26.92 to 40.74) |
| Diet high in sodium | Male | Grenada | 0.71 (0.00 to 2.72) |
| Diet high in sodium | Female | Grenada | 0.40 (0.00 to 1.91) |
| Diet low in fruits | Male | Grenada | 2.17 (1.41 to 3.12) |
| Diet low in fruits | Female | Grenada | 2.30 (1.48 to 3.28) |
| Diet low in vegetables | Male | Grenada | 4.97 (3.39 to 7.00) |
| Diet low in vegetables | Female | Grenada | 5.01 (3.27 to 7.15) |
| High body-mass index | Male | Grenada | 6.10 (3.21 to 10.09) |
| High body-mass index | Female | Grenada | 8.69 (4.56 to 14.76) |
| High systolic blood pressure | Male | Grenada | 16.46 (11.63 to 22.13) |
| High systolic blood pressure | Female | Grenada | 16.92 (10.63 to 23.64) |
| Lead exposure | Male | Grenada | 1.19 (-0.16 to 3.21) |
| Lead exposure | Female | Grenada | 0.91 (-0.13 to 2.38) |
| Smoking | Male | Grenada | 22.95 (17.76 to 29.05) |
| Smoking | Female | Grenada | 8.39 (6.09 to 11.30) |
| Diet high in sodium | Male | Japan | 1.19 (0.08 to 3.32) |
| Diet high in sodium | Female | Japan | 0.86 (0.02 to 2.81) |
| Diet low in fruits | Male | Japan | 4.22 (2.92 to 5.69) |
| Diet low in fruits | Female | Japan | 3.86 (2.61 to 5.22) |
| Diet low in vegetables | Male | Japan | 1.24 (0.63 to 2.03) |
| Diet low in vegetables | Female | Japan | 1.34 (0.69 to 2.25) |
| High body-mass index | Male | Japan | 4.55 (2.46 to 7.37) |
| High body-mass index | Female | Japan | 4.45 (2.30 to 7.45) |
| High systolic blood pressure | Male | Japan | 17.86 (13.28 to 23.53) |
| High systolic blood pressure | Female | Japan | 17.19 (11.77 to 23.51) |
| Lead exposure | Male | Japan | 0.35 (-0.04 to 0.95) |
| Lead exposure | Female | Japan | 0.33 (-0.05 to 0.86) |
| Smoking | Male | Japan | 42.02 (35.73 to 48.55) |
| Smoking | Female | Japan | 11.64 (8.77 to 15.14) |
| Diet high in sodium | Male | Monaco | 0.62 (0.00 to 2.40) |
| Diet high in sodium | Female | Monaco | 0.30 (0.00 to 1.55) |
| Diet low in fruits | Male | Monaco | 1.08 (0.56 to 1.75) |
| Diet low in fruits | Female | Monaco | 1.15 (0.52 to 2.09) |
| Diet low in vegetables | Male | Monaco | 0.58 (0.14 to 1.28) |
| Diet low in vegetables | Female | Monaco | 0.53 (0.09 to 1.34) |
| High body-mass index | Male | Monaco | 9.20 (4.81 to 16.58) |
| High body-mass index | Female | Monaco | 9.78 (4.88 to 16.96) |
| High systolic blood pressure | Male | Monaco | 17.56 (11.99 to 23.00) |
| High systolic blood pressure | Female | Monaco | 18.51 (12.18 to 25.44) |
| Lead exposure | Male | Monaco | 0.43 (-0.06 to 1.07) |
| Lead exposure | Female | Monaco | 0.34 (-0.04 to 0.90) |
| Smoking | Male | Monaco | 40.67 (32.67 to 48.93) |
| Smoking | Female | Monaco | 19.31 (13.64 to 25.84) |
| Diet high in sodium | Male | Montenegro | 1.78 (0.35 to 3.90) |
| Diet high in sodium | Female | Montenegro | 1.12 (0.10 to 3.06) |
| Diet low in fruits | Male | Montenegro | 1.48 (0.93 to 2.24) |
| Diet low in fruits | Female | Montenegro | 1.70 (1.04 to 2.53) |
| Diet low in vegetables | Male | Montenegro | 0.52 (0.16 to 1.16) |
| Diet low in vegetables | Female | Montenegro | 0.47 (0.10 to 1.07) |
| High body-mass index | Male | Montenegro | 12.91 (6.71 to 21.80) |
| High body-mass index | Female | Montenegro | 12.95 (6.70 to 22.01) |
| High systolic blood pressure | Male | Montenegro | 21.03 (15.24 to 27.56) |
| High systolic blood pressure | Female | Montenegro | 21.76 (15.00 to 29.54) |
| Lead exposure | Male | Montenegro | 0.38 (-0.05 to 1.05) |
| Lead exposure | Female | Montenegro | 0.29 (-0.04 to 0.77) |
| Smoking | Male | Montenegro | 49.16 (41.22 to 56.70) |
| Smoking | Female | Montenegro | 24.78 (18.80 to 30.62) |
| Diet high in sodium | Male | Nauru | 1.25 (0.12 to 3.40) |
| Diet high in sodium | Female | Nauru | 0.97 (0.02 to 3.23) |
| Diet low in fruits | Male | Nauru | 4.10 (2.68 to 5.68) |
| Diet low in fruits | Female | Nauru | 3.86 (2.44 to 5.59) |
| Diet low in vegetables | Male | Nauru | 5.45 (3.69 to 7.55) |
| Diet low in vegetables | Female | Nauru | 5.00 (3.31 to 7.34) |
| High body-mass index | Male | Nauru | 12.19 (6.06 to 21.82) |
| High body-mass index | Female | Nauru | 11.50 (5.36 to 21.12) |
| High systolic blood pressure | Male | Nauru | 19.80 (14.01 to 26.16) |
| High systolic blood pressure | Female | Nauru | 17.22 (10.26 to 25.62) |
| Lead exposure | Male | Nauru | 0.28 (-0.04 to 0.75) |
| Lead exposure | Female | Nauru | 0.43 (-0.06 to 0.92) |
| Smoking | Male | Nauru | 38.26 (29.29 to 47.36) |
| Smoking | Female | Nauru | 15.84 (8.70 to 26.37) |
| Diet high in sodium | Male | Norway | 0.55 (0.00 to 2.08) |
| Diet high in sodium | Female | Norway | 0.18 (0.00 to 0.98) |
| Diet low in fruits | Male | Norway | 2.80 (1.90 to 3.84) |
| Diet low in fruits | Female | Norway | 3.16 (2.06 to 4.51) |
| Diet low in vegetables | Male | Norway | 4.01 (2.65 to 5.74) |
| Diet low in vegetables | Female | Norway | 4.25 (2.74 to 6.37) |
| High body-mass index | Male | Norway | 7.11 (3.72 to 12.47) |
| High body-mass index | Female | Norway | 8.26 (4.29 to 14.30) |
| High systolic blood pressure | Male | Norway | 18.32 (13.37 to 24.10) |
| High systolic blood pressure | Female | Norway | 20.60 (14.47 to 27.60) |
| Lead exposure | Male | Norway | 0.48 (-0.06 to 1.24) |
| Lead exposure | Female | Norway | 0.30 (-0.05 to 0.85) |
| Smoking | Male | Norway | 27.81 (21.97 to 35.31) |
| Smoking | Female | Norway | 16.10 (11.62 to 21.98) |
| Diet high in sodium | Male | Saint Lucia | 0.68 (0.00 to 2.58) |
| Diet high in sodium | Female | Saint Lucia | 0.37 (0.00 to 1.76) |
| Diet low in fruits | Male | Saint Lucia | 2.77 (1.83 to 3.88) |
| Diet low in fruits | Female | Saint Lucia | 2.92 (1.85 to 4.22) |
| Diet low in vegetables | Male | Saint Lucia | 4.86 (3.24 to 6.83) |
| Diet low in vegetables | Female | Saint Lucia | 4.95 (3.35 to 7.15) |
| High body-mass index | Male | Saint Lucia | 6.14 (3.27 to 10.47) |
| High body-mass index | Female | Saint Lucia | 7.95 (4.14 to 13.69) |
| High systolic blood pressure | Male | Saint Lucia | 16.20 (11.08 to 21.75) |
| High systolic blood pressure | Female | Saint Lucia | 17.07 (11.04 to 23.87) |
| Lead exposure | Male | Saint Lucia | 1.17 (-0.16 to 3.06) |
| Lead exposure | Female | Saint Lucia | 0.85 (-0.13 to 2.21) |
| Smoking | Male | Saint Lucia | 24.46 (18.95 to 31.39) |
| Smoking | Female | Saint Lucia | 8.48 (6.16 to 11.51) |

#### Fig 6E. World maps of percentage contribution of risk factors to AA-caused ASDR in 2021

**Diet high in sodium**

| **Risk factor** | **Location** | **Percent contribution to ASDR (%)** |
| --- | --- | --- |
| Diet high in sodium | People's Republic of China | 2,16 (0,42 to 4,96) |
| Diet high in sodium | Republic of Korea | 1,99 (0,25 to 5,12) |
| Diet high in sodium | Czech Republic | 1,91 (0,33 to 4,48) |
| Diet high in sodium | Democratic People's Republic of Korea | 1,81 (0,28 to 4,65) |
| Diet high in sodium | Bosnia and Herzegovina | 1,77 (0,33 to 4,13) |
| Diet high in sodium | Republic of Bulgaria | 1,77 (0,32 to 3,97) |
| Diet high in sodium | North Macedonia | 1,64 (0,28 to 3,83) |
| Diet high in sodium | Republic of Singapore | 1,63 (0,12 to 4,41) |
| Diet high in sodium | Slovak Republic | 1,61 (0,30 to 3,75) |
| Diet high in sodium | Montenegro | 1,60 (0,29 to 3,61) |
| Diet high in sodium | Republic of Slovenia | 1,60 (0,30 to 3,55) |
| Diet high in sodium | Kingdom of Cambodia | 1,59 (0,14 to 4,40) |
| Diet high in sodium | Republic of Albania | 1,59 (0,28 to 3,62) |
| Diet high in sodium | Republic of Croatia | 1,58 (0,29 to 3,66) |
| Diet high in sodium | Republic of Serbia | 1,52 (0,29 to 3,36) |
| Diet high in sodium | Kingdom of Thailand | 1,50 (0,14 to 4,14) |
| Diet high in sodium | Hungary | 1,47 (0,29 to 3,28) |
| Diet high in sodium | Republic of the Philippines | 1,45 (0,15 to 4,02) |
| Diet high in sodium | Brunei Darussalam | 1,44 (0,13 to 4,05) |
| Diet high in sodium | Lao People's Democratic Republic | 1,43 (0,13 to 4,01) |
| Diet high in sodium | Romania | 1,42 (0,26 to 3,30) |
| Diet high in sodium | Independent State of Papua New Guinea | 1,41 (0,09 to 4,18) |
| Diet high in sodium | Republic of Colombia | 1,41 (0,22 to 3,44) |
| Diet high in sodium | Republic of Poland | 1,40 (0,16 to 3,74) |
| Diet high in sodium | Solomon Islands | 1,40 (0,10 to 4,14) |
| Diet high in sodium | Republic of Mauritius | 1,39 (0,16 to 3,74) |
| Diet high in sodium | Republic of the Marshall Islands | 1,34 (0,08 to 4,13) |
| Diet high in sodium | Republic of Kiribati | 1,33 (0,11 to 3,94) |
| Diet high in sodium | Republic of the Union of Myanmar | 1,32 (0,14 to 3,50) |
| Diet high in sodium | Federated States of Micronesia | 1,31 (0,08 to 3,94) |
| Diet high in sodium | Democratic Socialist Republic of Sri Lanka | 1,31 (0,12 to 3,63) |
| Diet high in sodium | Kingdom of Tonga | 1,30 (0,10 to 3,77) |
| Diet high in sodium | Republic of Maldives | 1,28 (0,11 to 3,48) |
| Diet high in sodium | Cook Islands | 1,27 (0,08 to 3,73) |
| Diet high in sodium | Democratic Republic of Timor-Leste | 1,26 (0,12 to 3,47) |
| Diet high in sodium | Guam | 1,24 (0,10 to 3,52) |
| Diet high in sodium | Tuvalu | 1,24 (0,08 to 3,65) |
| Diet high in sodium | Tokelau | 1,24 (0,08 to 3,88) |
| Diet high in sodium | Northern Mariana Islands | 1,22 (0,08 to 3,59) |
| Diet high in sodium | Republic of Palau | 1,22 (0,07 to 3,62) |
| Diet high in sodium | Socialist Republic of Viet Nam | 1,20 (0,12 to 3,35) |
| Diet high in sodium | Republic of Seychelles | 1,18 (0,11 to 3,25) |
| Diet high in sodium | Republic of Fiji | 1,15 (0,08 to 3,39) |
| Diet high in sodium | Malaysia | 1,15 (0,12 to 3,10) |
| Diet high in sodium | Republic of Guatemala | 1,12 (0,05 to 3,50) |
| Diet high in sodium | Taiwan (Province of China) | 1,11 (0,06 to 3,39) |
| Diet high in sodium | Republic of El Salvador | 1,09 (0,06 to 3,25) |
| Diet high in sodium | Republic of Nauru | 1,09 (0,07 to 3,26) |
| Diet high in sodium | Republic of Nicaragua | 1,08 (0,06 to 3,35) |
| Diet high in sodium | Republic of Honduras | 1,05 (0,04 to 3,21) |
| Diet high in sodium | Japan | 1,05 (0,06 to 3,16) |
| Diet high in sodium | Republic of Panama | 1,03 (0,05 to 3,16) |
| Diet high in sodium | Republic of Niue | 1,03 (0,08 to 2,88) |
| Diet high in sodium | Federal Democratic Republic of Ethiopia | 1,02 (0,03 to 3,43) |
| Diet high in sodium | Republic of Ecuador | 1,00 (0,02 to 3,47) |
| Diet high in sodium | Plurinational State of Bolivia | 0,97 (0,02 to 3,36) |
| Diet high in sodium | Eastern Republic of Uruguay | 0,96 (0,02 to 3,19) |
| Diet high in sodium | Republic of Costa Rica | 0,96 (0,06 to 2,85) |
| Diet high in sodium | Argentine Republic | 0,95 (0,02 to 3,29) |
| Diet high in sodium | Republic of Zambia | 0,95 (0,02 to 3,30) |
| Diet high in sodium | State of Eritrea | 0,94 (0,02 to 3,27) |
| Diet high in sodium | Republic of Rwanda | 0,94 (0,02 to 3,26) |
| Diet high in sodium | Republic of Tajikistan | 0,93 (0,04 to 3,04) |
| Diet high in sodium | Republic of Peru | 0,92 (0,02 to 3,22) |
| Diet high in sodium | Federal Republic of Somalia | 0,92 (0,01 to 3,01) |
| Diet high in sodium | Kyrgyz Republic | 0,92 (0,03 to 3,11) |
| Diet high in sodium | Republic of Paraguay | 0,91 (0,02 to 2,99) |
| Diet high in sodium | Bolivarian Republic of Venezuela | 0,91 (0,05 to 2,61) |
| Diet high in sodium | Republic of Vanuatu | 0,89 (0,07 to 2,55) |
| Diet high in sodium | Republic of Uzbekistan | 0,89 (0,03 to 2,94) |
| Diet high in sodium | United Republic of Tanzania | 0,87 (0,05 to 2,60) |
| Diet high in sodium | Republic of South Sudan | 0,87 (0,01 to 3,05) |
| Diet high in sodium | Federal Democratic Republic of Nepal | 0,86 (0,01 to 3,20) |
| Diet high in sodium | Union of the Comoros | 0,85 (0,02 to 2,80) |
| Diet high in sodium | Republic of Austria | 0,85 (0,02 to 2,66) |
| Diet high in sodium | Republic of Burundi | 0,84 (0,02 to 2,85) |
| Diet high in sodium | Republic of Azerbaijan | 0,83 (0,03 to 2,88) |
| Diet high in sodium | Republic of Indonesia | 0,83 (0,09 to 2,15) |
| Diet high in sodium | Republic of Madagascar | 0,83 (0,01 to 2,85) |
| Diet high in sodium | Federative Republic of Brazil | 0,82 (0,02 to 2,72) |
| Diet high in sodium | Republic of Armenia | 0,82 (0,03 to 2,77) |
| Diet high in sodium | Republic of Djibouti | 0,79 (0,01 to 2,66) |
| Diet high in sodium | Republic of Uganda | 0,79 (0,01 to 2,78) |
| Diet high in sodium | People's Republic of Bangladesh | 0,79 (0,01 to 2,84) |
| Diet high in sodium | Kingdom of Bhutan | 0,79 (0,00 to 2,85) |
| Diet high in sodium | Russian Federation | 0,78 (0,04 to 2,42) |
| Diet high in sodium | Republic of Italy | 0,77 (0,03 to 2,63) |
| Diet high in sodium | Republic of Malawi | 0,76 (0,01 to 2,47) |
| Diet high in sodium | Kingdom of Belgium | 0,76 (0,01 to 2,64) |
| Diet high in sodium | Turkmenistan | 0,76 (0,03 to 2,61) |
| Diet high in sodium | Republic of Mozambique | 0,75 (0,02 to 2,50) |
| Diet high in sodium | Republic of Chile | 0,74 (0,02 to 2,41) |
| Diet high in sodium | Canada | 0,72 (0,01 to 2,74) |
| Diet high in sodium | Islamic Republic of Pakistan | 0,71 (0,01 to 2,59) |
| Diet high in sodium | Republic of Malta | 0,71 (0,02 to 2,56) |
| Diet high in sodium | Mongolia | 0,71 (0,02 to 2,33) |
| Diet high in sodium | Georgia | 0,70 (0,03 to 2,17) |
| Diet high in sodium | Republic of India | 0,70 (0,02 to 2,61) |
| Diet high in sodium | United Mexican States | 0,69 (0,01 to 2,45) |
| Diet high in sodium | United States of America | 0,69 (0,01 to 2,59) |
| Diet high in sodium | Republic of Suriname | 0,68 (0,00 to 2,82) |
| Diet high in sodium | Greenland | 0,67 (0,00 to 2,48) |
| Diet high in sodium | Republic of Cuba | 0,65 (0,00 to 2,65) |
| Diet high in sodium | American Samoa | 0,64 (0,01 to 2,47) |
| Diet high in sodium | Saint Vincent and the Grenadines | 0,64 (0,00 to 2,55) |
| Diet high in sodium | Republic of Iceland | 0,64 (0,01 to 2,46) |
| Diet high in sodium | Hellenic Republic | 0,63 (0,00 to 2,57) |
| Diet high in sodium | Bermuda | 0,62 (0,00 to 2,49) |
| Diet high in sodium | Portuguese Republic | 0,62 (0,01 to 2,46) |
| Diet high in sodium | Republic of Kazakhstan | 0,62 (0,02 to 2,10) |
| Diet high in sodium | Antigua and Barbuda | 0,61 (0,00 to 2,52) |
| Diet high in sodium | Jamaica | 0,61 (0,00 to 2,47) |
| Diet high in sodium | Dominican Republic | 0,61 (0,00 to 2,45) |
| Diet high in sodium | Republic of Ghana | 0,60 (0,00 to 2,26) |
| Diet high in sodium | United States Virgin Islands | 0,59 (0,00 to 2,28) |
| Diet high in sodium | Puerto Rico | 0,58 (0,00 to 2,47) |
| Diet high in sodium | Saint Kitts and Nevis | 0,58 (0,00 to 2,35) |
| Diet high in sodium | Federal Republic of Germany | 0,58 (0,01 to 2,19) |
| Diet high in sodium | Republic of San Marino | 0,57 (0,00 to 2,26) |
| Diet high in sodium | Swiss Confederation | 0,57 (0,00 to 2,38) |
| Diet high in sodium | Grenada | 0,57 (0,00 to 2,29) |
| Diet high in sodium | Republic of Latvia | 0,56 (0,01 to 2,02) |
| Diet high in sodium | Commonwealth of Dominica | 0,56 (0,00 to 2,35) |
| Diet high in sodium | Independent State of Samoa | 0,56 (0,00 to 2,33) |
| Diet high in sodium | Republic of Guyana | 0,56 (0,00 to 2,32) |
| Diet high in sodium | Belize | 0,56 (0,00 to 2,35) |
| Diet high in sodium | Republic of Haiti | 0,55 (0,00 to 2,29) |
| Diet high in sodium | Commonwealth of the Bahamas | 0,55 (0,00 to 2,36) |
| Diet high in sodium | Saint Lucia | 0,52 (0,00 to 2,15) |
| Diet high in sodium | Republic of Cyprus | 0,52 (0,00 to 2,14) |
| Diet high in sodium | Kingdom of Sweden | 0,52 (0,00 to 2,03) |
| Diet high in sodium | State of Kuwait | 0,51 (0,00 to 2,26) |
| Diet high in sodium | Republic of Benin | 0,51 (0,00 to 2,19) |
| Diet high in sodium | State of Israel | 0,50 (0,00 to 2,02) |
| Diet high in sodium | Grand Duchy of Luxembourg | 0,49 (0,00 to 2,05) |
| Diet high in sodium | New Zealand | 0,49 (0,00 to 2,19) |
| Diet high in sodium | Republic of Finland | 0,49 (0,00 to 1,94) |
| Diet high in sodium | Principality of Monaco | 0,48 (0,00 to 2,00) |
| Diet high in sodium | Kingdom of Denmark | 0,47 (0,00 to 1,91) |
| Diet high in sodium | French Republic | 0,47 (0,00 to 1,99) |
| Diet high in sodium | United Kingdom of Great Britain and Northern Ireland | 0,47 (0,00 to 2,06) |
| Diet high in sodium | Republic of Chad | 0,47 (0,00 to 2,09) |
| Diet high in sodium | Burkina Faso | 0,46 (0,00 to 2,09) |
| Diet high in sodium | Republic of Guinea | 0,46 (0,00 to 2,05) |
| Diet high in sodium | Republic of Trinidad and Tobago | 0,45 (0,00 to 1,86) |
| Diet high in sodium | Republic of Côte d'Ivoire | 0,44 (0,00 to 1,93) |
| Diet high in sodium | Kingdom of the Netherlands | 0,44 (0,00 to 1,88) |
| Diet high in sodium | Republic of Cameroon | 0,42 (0,00 to 1,96) |
| Diet high in sodium | Islamic Republic of Mauritania | 0,42 (0,00 to 1,90) |
| Diet high in sodium | Republic of Mali | 0,42 (0,00 to 1,84) |
| Diet high in sodium | Ukraine | 0,42 (0,00 to 1,80) |
| Diet high in sodium | Republic of Guinea-Bissau | 0,42 (0,00 to 1,90) |
| Diet high in sodium | Republic of the Niger | 0,41 (0,00 to 1,92) |
| Diet high in sodium | Republic of Lithuania | 0,41 (0,00 to 1,72) |
| Diet high in sodium | Republic of Liberia | 0,40 (0,00 to 1,78) |
| Diet high in sodium | Kingdom of Norway | 0,40 (0,00 to 1,65) |
| Diet high in sodium | Republic of Kenya | 0,39 (0,01 to 1,45) |
| Diet high in sodium | Republic of the Gambia | 0,39 (0,00 to 1,71) |
| Diet high in sodium | Gabonese Republic | 0,39 (0,00 to 1,87) |
| Diet high in sodium | Republic of Cabo Verde | 0,39 (0,00 to 1,60) |
| Diet high in sodium | Republic of Zimbabwe | 0,38 (0,00 to 1,75) |
| Diet high in sodium | Principality of Andorra | 0,38 (0,00 to 1,70) |
| Diet high in sodium | Togolese Republic | 0,37 (0,00 to 1,68) |
| Diet high in sodium | Democratic Republic of Sao Tome and Principe | 0,37 (0,00 to 1,69) |
| Diet high in sodium | Republic of Belarus | 0,37 (0,00 to 1,59) |
| Diet high in sodium | Barbados | 0,36 (0,00 to 1,78) |
| Diet high in sodium | Republic of Senegal | 0,36 (0,00 to 1,56) |
| Diet high in sodium | Republic of Angola | 0,36 (0,00 to 1,76) |
| Diet high in sodium | Central African Republic | 0,36 (0,00 to 1,75) |
| Diet high in sodium | Republic of Moldova | 0,35 (0,00 to 1,62) |
| Diet high in sodium | Republic of the Congo | 0,35 (0,00 to 1,63) |
| Diet high in sodium | Republic of Equatorial Guinea | 0,34 (0,00 to 1,62) |
| Diet high in sodium | Ireland | 0,33 (0,00 to 1,64) |
| Diet high in sodium | Kingdom of Spain | 0,32 (0,00 to 1,47) |
| Diet high in sodium | Republic of Sierra Leone | 0,30 (0,00 to 1,33) |
| Diet high in sodium | Federal Republic of Nigeria | 0,30 (0,00 to 1,36) |
| Diet high in sodium | Kingdom of Lesotho | 0,29 (0,00 to 1,64) |
| Diet high in sodium | Australia | 0,29 (0,00 to 1,59) |
| Diet high in sodium | Republic of Namibia | 0,27 (0,00 to 1,46) |
| Diet high in sodium | Kingdom of Eswatini | 0,27 (0,00 to 1,44) |
| Diet high in sodium | Republic of Estonia | 0,26 (0,00 to 1,35) |
| Diet high in sodium | Republic of Botswana | 0,25 (0,00 to 1,39) |
| Diet high in sodium | Arab Republic of Egypt | 0,24 (0,00 to 1,40) |
| Diet high in sodium | Sultanate of Oman | 0,24 (0,00 to 1,34) |
| Diet high in sodium | Palestine | 0,24 (0,00 to 1,35) |
| Diet high in sodium | Hashemite Kingdom of Jordan | 0,23 (0,00 to 1,35) |
| Diet high in sodium | Republic of Yemen | 0,23 (0,00 to 1,35) |
| Diet high in sodium | Islamic Republic of Iran | 0,23 (0,00 to 1,29) |
| Diet high in sodium | Republic of Tunisia | 0,23 (0,00 to 1,33) |
| Diet high in sodium | Islamic Republic of Afghanistan | 0,23 (0,00 to 1,35) |
| Diet high in sodium | Syrian Arab Republic | 0,23 (0,00 to 1,32) |
| Diet high in sodium | Lebanese Republic | 0,22 (0,00 to 1,27) |
| Diet high in sodium | State of Libya | 0,21 (0,00 to 1,15) |
| Diet high in sodium | Kingdom of Bahrain | 0,21 (0,00 to 1,14) |
| Diet high in sodium | State of Qatar | 0,21 (0,00 to 1,15) |
| Diet high in sodium | Republic of South Africa | 0,21 (0,00 to 1,08) |
| Diet high in sodium | People's Democratic Republic of Algeria | 0,21 (0,00 to 1,22) |
| Diet high in sodium | Republic of Sudan | 0,20 (0,00 to 1,14) |
| Diet high in sodium | Kingdom of Morocco | 0,20 (0,00 to 1,12) |
| Diet high in sodium | United Arab Emirates | 0,20 (0,00 to 1,27) |
| Diet high in sodium | Republic of Iraq | 0,19 (0,00 to 1,10) |
| Diet high in sodium | Kingdom of Saudi Arabia | 0,19 (0,00 to 1,16) |
| Diet high in sodium | Democratic Republic of the Congo | 0,19 (0,00 to 1,23) |
| Diet high in sodium | Republic of Turkey | 0,16 (0,00 to 1,06) |

**Diet low in fruits**

| **Risk factor** | **Location** | **Percent contribution to ASDR (%)** |
| --- | --- | --- |
| Diet low in fruits | Togolese Republic | 6,68 (4,67 to 8,89) |
| Diet low in fruits | Republic of Zimbabwe | 6,63 (4,57 to 8,86) |
| Diet low in fruits | Republic of the Gambia | 6,62 (4,56 to 8,85) |
| Diet low in fruits | Mongolia | 6,47 (4,42 to 8,59) |
| Diet low in fruits | Islamic Republic of Mauritania | 6,26 (4,28 to 8,42) |
| Diet low in fruits | Burkina Faso | 6,06 (4,13 to 8,19) |
| Diet low in fruits | Republic of Sierra Leone | 6,02 (4,21 to 8,03) |
| Diet low in fruits | Republic of Senegal | 5,99 (4,14 to 8,02) |
| Diet low in fruits | Republic of Djibouti | 5,98 (4,17 to 7,95) |
| Diet low in fruits | Republic of Botswana | 5,86 (4,11 to 7,99) |
| Diet low in fruits | Republic of the Niger | 5,82 (3,94 to 8,04) |
| Diet low in fruits | Republic of Mozambique | 5,77 (3,99 to 7,73) |
| Diet low in fruits | Republic of South Africa | 5,69 (3,96 to 7,55) |
| Diet low in fruits | Democratic Republic of Timor-Leste | 5,66 (3,89 to 7,51) |
| Diet low in fruits | Republic of India | 5,65 (3,94 to 7,38) |
| Diet low in fruits | Republic of Chad | 5,64 (3,95 to 7,73) |
| Diet low in fruits | Kingdom of Lesotho | 5,63 (3,77 to 7,62) |
| Diet low in fruits | Republic of Fiji | 5,51 (3,75 to 7,49) |
| Diet low in fruits | Federal Republic of Somalia | 5,49 (3,83 to 7,41) |
| Diet low in fruits | Ukraine | 5,49 (3,87 to 7,38) |
| Diet low in fruits | Republic of Nicaragua | 5,48 (3,80 to 7,35) |
| Diet low in fruits | Republic of Latvia | 5,44 (3,75 to 7,24) |
| Diet low in fruits | Republic of Namibia | 5,44 (3,78 to 7,38) |
| Diet low in fruits | Republic of Zambia | 5,42 (3,81 to 7,18) |
| Diet low in fruits | People's Republic of Bangladesh | 5,33 (3,70 to 7,16) |
| Diet low in fruits | Republic of Indonesia | 5,29 (3,62 to 7,17) |
| Diet low in fruits | Republic of Benin | 5,26 (3,66 to 6,97) |
| Diet low in fruits | Republic of Liberia | 5,24 (3,67 to 7,11) |
| Diet low in fruits | Republic of Moldova | 5,21 (3,55 to 6,97) |
| Diet low in fruits | Republic of Mauritius | 5,20 (3,60 to 6,89) |
| Diet low in fruits | Republic of Cabo Verde | 5,15 (3,60 to 6,91) |
| Diet low in fruits | Republic of Mali | 5,10 (3,50 to 6,84) |
| Diet low in fruits | Federal Democratic Republic of Ethiopia | 5,08 (3,52 to 6,87) |
| Diet low in fruits | Hungary | 5,04 (3,53 to 6,77) |
| Diet low in fruits | Georgia | 5,01 (3,40 to 6,77) |
| Diet low in fruits | Republic of Kazakhstan | 5,00 (3,50 to 6,61) |
| Diet low in fruits | Central African Republic | 4,99 (3,47 to 6,80) |
| Diet low in fruits | Republic of the Union of Myanmar | 4,97 (3,55 to 6,67) |
| Diet low in fruits | Republic of Tajikistan | 4,96 (3,49 to 6,64) |
| Diet low in fruits | Kingdom of Eswatini | 4,95 (3,41 to 6,61) |
| Diet low in fruits | Islamic Republic of Pakistan | 4,94 (3,45 to 6,52) |
| Diet low in fruits | Republic of Guinea-Bissau | 4,90 (3,39 to 6,66) |
| Diet low in fruits | Democratic Socialist Republic of Sri Lanka | 4,89 (3,37 to 6,60) |
| Diet low in fruits | Republic of Lithuania | 4,87 (3,31 to 6,40) |
| Diet low in fruits | Saint Kitts and Nevis | 4,83 (3,30 to 6,50) |
| Diet low in fruits | Republic of Trinidad and Tobago | 4,82 (3,25 to 6,42) |
| Diet low in fruits | Kyrgyz Republic | 4,80 (3,35 to 6,33) |
| Diet low in fruits | Democratic Republic of the Congo | 4,79 (3,36 to 6,40) |
| Diet low in fruits | Republic of Madagascar | 4,78 (3,33 to 6,52) |
| Diet low in fruits | Federal Republic of Nigeria | 4,76 (3,31 to 6,30) |
| Diet low in fruits | Kingdom of Cambodia | 4,76 (3,28 to 6,45) |
| Diet low in fruits | Slovak Republic | 4,74 (3,27 to 6,38) |
| Diet low in fruits | Republic of Belarus | 4,71 (3,28 to 6,19) |
| Diet low in fruits | Republic of Malawi | 4,69 (3,23 to 6,20) |
| Diet low in fruits | Republic of the Congo | 4,66 (3,22 to 6,21) |
| Diet low in fruits | State of Eritrea | 4,66 (3,15 to 6,45) |
| Diet low in fruits | Republic of Estonia | 4,66 (3,30 to 6,26) |
| Diet low in fruits | Barbados | 4,63 (3,20 to 6,22) |
| Diet low in fruits | Republic of Yemen | 4,62 (3,20 to 6,13) |
| Diet low in fruits | Republic of Bulgaria | 4,60 (3,13 to 6,04) |
| Diet low in fruits | Islamic Republic of Afghanistan | 4,57 (3,19 to 6,11) |
| Diet low in fruits | Republic of Kenya | 4,56 (3,20 to 6,10) |
| Diet low in fruits | Republic of South Sudan | 4,49 (3,07 to 6,04) |
| Diet low in fruits | Brunei Darussalam | 4,47 (3,12 to 5,93) |
| Diet low in fruits | Russian Federation | 4,47 (3,13 to 5,92) |
| Diet low in fruits | Tuvalu | 4,39 (2,99 to 5,94) |
| Diet low in fruits | Kingdom of Bhutan | 4,38 (3,04 to 5,87) |
| Diet low in fruits | Socialist Republic of Viet Nam | 4,34 (2,98 to 5,77) |
| Diet low in fruits | Republic of Angola | 4,32 (2,98 to 5,78) |
| Diet low in fruits | Republic of Vanuatu | 4,32 (2,83 to 6,00) |
| Diet low in fruits | Kingdom of Tonga | 4,29 (2,99 to 5,78) |
| Diet low in fruits | Czech Republic | 4,28 (3,01 to 5,77) |
| Diet low in fruits | Republic of Iraq | 4,22 (2,93 to 5,50) |
| Diet low in fruits | Republic of Finland | 4,21 (2,91 to 5,75) |
| Diet low in fruits | Republic of Maldives | 4,20 (2,92 to 5,69) |
| Diet low in fruits | Tokelau | 4,17 (2,76 to 5,74) |
| Diet low in fruits | Republic of Guyana | 4,15 (2,85 to 5,58) |
| Diet low in fruits | Republic of the Marshall Islands | 4,12 (2,84 to 5,62) |
| Diet low in fruits | Independent State of Samoa | 4,11 (2,85 to 5,59) |
| Diet low in fruits | Japan | 4,11 (2,78 to 5,46) |
| Diet low in fruits | Solomon Islands | 4,04 (2,83 to 5,45) |
| Diet low in fruits | Republic of Sudan | 4,04 (2,76 to 5,51) |
| Diet low in fruits | Republic of Côte d'Ivoire | 4,03 (2,78 to 5,37) |
| Diet low in fruits | Republic of Chile | 4,03 (2,73 to 5,42) |
| Diet low in fruits | Republic of Poland | 4,01 (2,80 to 5,31) |
| Diet low in fruits | French Republic | 3,98 (2,77 to 5,37) |
| Diet low in fruits | Republic of Nauru | 3,96 (2,68 to 5,48) |
| Diet low in fruits | Republic of Haiti | 3,94 (2,69 to 5,25) |
| Diet low in fruits | Federated States of Micronesia | 3,93 (2,69 to 5,32) |
| Diet low in fruits | Democratic People's Republic of Korea | 3,92 (2,71 to 5,30) |
| Diet low in fruits | Republic of Niue | 3,90 (2,70 to 5,33) |
| Diet low in fruits | Republic of Panama | 3,90 (2,71 to 5,23) |
| Diet low in fruits | Malaysia | 3,89 (2,64 to 5,28) |
| Diet low in fruits | Republic of Seychelles | 3,88 (2,68 to 5,24) |
| Diet low in fruits | Republic of El Salvador | 3,88 (2,75 to 5,12) |
| Diet low in fruits | Republic of Kiribati | 3,83 (2,65 to 5,12) |
| Diet low in fruits | American Samoa | 3,80 (2,57 to 5,18) |
| Diet low in fruits | Romania | 3,78 (2,63 to 5,07) |
| Diet low in fruits | Federal Republic of Germany | 3,78 (2,61 to 5,02) |
| Diet low in fruits | Republic of Guatemala | 3,70 (2,52 to 4,86) |
| Diet low in fruits | Federal Democratic Republic of Nepal | 3,70 (2,58 to 5,02) |
| Diet low in fruits | United Kingdom of Great Britain and Northern Ireland | 3,66 (2,55 to 4,86) |
| Diet low in fruits | United Republic of Tanzania | 3,64 (2,56 to 4,90) |
| Diet low in fruits | Republic of Palau | 3,63 (2,42 to 4,93) |
| Diet low in fruits | Republic of Iceland | 3,62 (2,54 to 4,85) |
| Diet low in fruits | Lao People's Democratic Republic | 3,59 (2,46 to 4,83) |
| Diet low in fruits | Independent State of Papua New Guinea | 3,58 (2,44 to 4,86) |
| Diet low in fruits | Hashemite Kingdom of Jordan | 3,55 (2,44 to 4,70) |
| Diet low in fruits | Republic of Honduras | 3,55 (2,41 to 4,72) |
| Diet low in fruits | Republic of Uzbekistan | 3,51 (2,47 to 4,72) |
| Diet low in fruits | Australia | 3,50 (2,42 to 4,73) |
| Diet low in fruits | Turkmenistan | 3,47 (2,40 to 4,63) |
| Diet low in fruits | Ireland | 3,44 (2,38 to 4,76) |
| Diet low in fruits | Bosnia and Herzegovina | 3,43 (2,36 to 4,68) |
| Diet low in fruits | Bolivarian Republic of Venezuela | 3,41 (2,35 to 4,65) |
| Diet low in fruits | Kingdom of Belgium | 3,39 (2,29 to 4,58) |
| Diet low in fruits | Bermuda | 3,36 (2,28 to 4,55) |
| Diet low in fruits | Grand Duchy of Luxembourg | 3,36 (2,30 to 4,57) |
| Diet low in fruits | Union of the Comoros | 3,33 (2,31 to 4,46) |
| Diet low in fruits | People's Republic of China | 3,32 (2,27 to 4,50) |
| Diet low in fruits | Republic of Cameroon | 3,31 (2,22 to 4,44) |
| Diet low in fruits | Palestine | 3,31 (2,26 to 4,48) |
| Diet low in fruits | Republic of Malta | 3,31 (2,25 to 4,43) |
| Diet low in fruits | Kingdom of Sweden | 3,30 (2,23 to 4,47) |
| Diet low in fruits | Republic of Croatia | 3,25 (2,22 to 4,43) |
| Diet low in fruits | Eastern Republic of Uruguay | 3,22 (2,16 to 4,39) |
| Diet low in fruits | Kingdom of Saudi Arabia | 3,19 (2,22 to 4,28) |
| Diet low in fruits | Republic of Peru | 3,17 (2,17 to 4,26) |
| Diet low in fruits | Republic of Suriname | 3,14 (2,19 to 4,25) |
| Diet low in fruits | Commonwealth of the Bahamas | 3,13 (2,12 to 4,21) |
| Diet low in fruits | United States of America | 3,13 (2,15 to 4,14) |
| Diet low in fruits | State of Libya | 3,11 (2,15 to 4,25) |
| Diet low in fruits | Kingdom of the Netherlands | 3,10 (2,07 to 4,24) |
| Diet low in fruits | Republic of Guinea | 3,10 (2,09 to 4,27) |
| Diet low in fruits | Cook Islands | 3,09 (2,12 to 4,23) |
| Diet low in fruits | State of Kuwait | 3,06 (2,09 to 4,08) |
| Diet low in fruits | Northern Mariana Islands | 3,04 (2,07 to 4,23) |
| Diet low in fruits | Republic of Equatorial Guinea | 3,01 (1,96 to 4,25) |
| Diet low in fruits | Republic of Azerbaijan | 2,98 (2,02 to 4,08) |
| Diet low in fruits | Republic of Korea | 2,96 (2,07 to 3,93) |
| Diet low in fruits | Kingdom of Norway | 2,96 (2,00 to 4,08) |
| Diet low in fruits | Swiss Confederation | 2,92 (2,02 to 4,01) |
| Diet low in fruits | Canada | 2,92 (1,99 to 3,95) |
| Diet low in fruits | Plurinational State of Bolivia | 2,92 (1,98 to 3,95) |
| Diet low in fruits | Republic of Colombia | 2,88 (1,98 to 3,81) |
| Diet low in fruits | Jamaica | 2,86 (1,98 to 3,78) |
| Diet low in fruits | Saint Lucia | 2,86 (1,89 to 3,88) |
| Diet low in fruits | New Zealand | 2,81 (1,95 to 3,78) |
| Diet low in fruits | Republic of Cyprus | 2,80 (1,91 to 3,75) |
| Diet low in fruits | Guam | 2,74 (1,85 to 3,78) |
| Diet low in fruits | Republic of Slovenia | 2,73 (1,91 to 3,76) |
| Diet low in fruits | Republic of San Marino | 2,72 (1,80 to 3,83) |
| Diet low in fruits | Kingdom of Denmark | 2,71 (1,83 to 3,81) |
| Diet low in fruits | North Macedonia | 2,70 (1,84 to 3,62) |
| Diet low in fruits | Puerto Rico | 2,69 (1,81 to 3,69) |
| Diet low in fruits | Republic of the Philippines | 2,69 (1,80 to 3,70) |
| Diet low in fruits | Syrian Arab Republic | 2,68 (1,80 to 3,61) |
| Diet low in fruits | People's Democratic Republic of Algeria | 2,67 (1,87 to 3,59) |
| Diet low in fruits | Republic of Paraguay | 2,60 (1,73 to 3,52) |
| Diet low in fruits | Portuguese Republic | 2,59 (1,80 to 3,50) |
| Diet low in fruits | Greenland | 2,54 (1,75 to 3,60) |
| Diet low in fruits | Republic of Costa Rica | 2,49 (1,68 to 3,36) |
| Diet low in fruits | Republic of Serbia | 2,48 (1,65 to 3,40) |
| Diet low in fruits | Principality of Andorra | 2,48 (1,54 to 3,52) |
| Diet low in fruits | United Mexican States | 2,47 (1,69 to 3,38) |
| Diet low in fruits | Antigua and Barbuda | 2,44 (1,65 to 3,36) |
| Diet low in fruits | Republic of Burundi | 2,41 (1,63 to 3,35) |
| Diet low in fruits | Democratic Republic of Sao Tome and Principe | 2,40 (1,60 to 3,39) |
| Diet low in fruits | Republic of Armenia | 2,38 (1,62 to 3,34) |
| Diet low in fruits | Federative Republic of Brazil | 2,34 (1,58 to 3,12) |
| Diet low in fruits | Saint Vincent and the Grenadines | 2,30 (1,57 to 3,19) |
| Diet low in fruits | Republic of Austria | 2,29 (1,55 to 3,20) |
| Diet low in fruits | Grenada | 2,27 (1,51 to 3,09) |
| Diet low in fruits | United Arab Emirates | 2,14 (1,43 to 2,98) |
| Diet low in fruits | Republic of Tunisia | 2,13 (1,44 to 2,91) |
| Diet low in fruits | Argentine Republic | 2,10 (1,42 to 2,92) |
| Diet low in fruits | Kingdom of Thailand | 2,07 (1,41 to 2,83) |
| Diet low in fruits | Republic of Cuba | 2,07 (1,40 to 2,77) |
| Diet low in fruits | Taiwan (Province of China) | 2,04 (1,34 to 2,78) |
| Diet low in fruits | Republic of Ghana | 1,90 (1,27 to 2,67) |
| Diet low in fruits | Republic of Singapore | 1,87 (1,21 to 2,56) |
| Diet low in fruits | Gabonese Republic | 1,85 (1,22 to 2,60) |
| Diet low in fruits | Kingdom of Morocco | 1,84 (1,23 to 2,59) |
| Diet low in fruits | Kingdom of Spain | 1,75 (1,15 to 2,47) |
| Diet low in fruits | Republic of Albania | 1,75 (1,08 to 2,47) |
| Diet low in fruits | United States Virgin Islands | 1,75 (1,14 to 2,55) |
| Diet low in fruits | Republic of Italy | 1,67 (1,12 to 2,24) |
| Diet low in fruits | Arab Republic of Egypt | 1,61 (1,05 to 2,21) |
| Diet low in fruits | Kingdom of Bahrain | 1,60 (1,00 to 2,45) |
| Diet low in fruits | Montenegro | 1,54 (1,01 to 2,18) |
| Diet low in fruits | State of Israel | 1,49 (0,99 to 2,13) |
| Diet low in fruits | Sultanate of Oman | 1,47 (0,95 to 2,09) |
| Diet low in fruits | Lebanese Republic | 1,43 (0,92 to 2,05) |
| Diet low in fruits | Republic of Uganda | 1,40 (0,92 to 2,03) |
| Diet low in fruits | Hellenic Republic | 1,22 (0,79 to 1,74) |
| Diet low in fruits | Republic of Ecuador | 1,20 (0,75 to 1,72) |
| Diet low in fruits | Islamic Republic of Iran | 1,16 (0,79 to 1,60) |
| Diet low in fruits | Principality of Monaco | 1,11 (0,63 to 1,74) |
| Diet low in fruits | Dominican Republic | 0,86 (0,51 to 1,28) |
| Diet low in fruits | Republic of Turkey | 0,86 (0,56 to 1,23) |
| Diet low in fruits | Belize | 0,85 (0,49 to 1,27) |
| Diet low in fruits | State of Qatar | 0,81 (0,44 to 1,32) |
| Diet low in fruits | Commonwealth of Dominica | 0,42 (0,24 to 0,70) |
| Diet low in fruits | Republic of Rwanda | 0,12 (0,05 to 0,24) |

**Diet low in vegetables**

| **Risk factor** | **Location** | **Percent contribution to ASDR (%)** |
| --- | --- | --- |
| Diet low in vegetables | Republic of Zimbabwe | 6,57 (4,46 to 9,16) |
| Diet low in vegetables | Togolese Republic | 6,31 (4,37 to 8,75) |
| Diet low in vegetables | Republic of Sierra Leone | 6,12 (4,09 to 8,64) |
| Diet low in vegetables | Republic of Guinea-Bissau | 6,08 (4,11 to 8,47) |
| Diet low in vegetables | Republic of the Congo | 6,08 (4,12 to 8,45) |
| Diet low in vegetables | Republic of Vanuatu | 6,06 (4,12 to 8,54) |
| Diet low in vegetables | Republic of the Gambia | 6,01 (4,04 to 8,38) |
| Diet low in vegetables | Republic of Liberia | 6,00 (4,01 to 8,52) |
| Diet low in vegetables | Democratic Republic of Sao Tome and Principe | 6,00 (4,00 to 8,32) |
| Diet low in vegetables | Republic of Mozambique | 5,98 (4,02 to 8,49) |
| Diet low in vegetables | Central African Republic | 5,96 (4,05 to 8,36) |
| Diet low in vegetables | Islamic Republic of Mauritania | 5,92 (4,00 to 8,39) |
| Diet low in vegetables | Union of the Comoros | 5,92 (4,04 to 8,15) |
| Diet low in vegetables | Kingdom of Eswatini | 5,90 (4,01 to 8,05) |
| Diet low in vegetables | Republic of Malawi | 5,89 (3,98 to 8,07) |
| Diet low in vegetables | Republic of Indonesia | 5,87 (3,94 to 8,21) |
| Diet low in vegetables | Republic of Nicaragua | 5,84 (3,96 to 8,00) |
| Diet low in vegetables | Republic of Haiti | 5,77 (3,87 to 7,89) |
| Diet low in vegetables | Republic of Equatorial Guinea | 5,72 (3,90 to 7,95) |
| Diet low in vegetables | Federal Republic of Somalia | 5,71 (3,84 to 8,10) |
| Diet low in vegetables | Republic of Madagascar | 5,66 (3,91 to 7,86) |
| Diet low in vegetables | Democratic Republic of the Congo | 5,65 (3,79 to 7,73) |
| Diet low in vegetables | Republic of Burundi | 5,64 (3,80 to 7,98) |
| Diet low in vegetables | Republic of Chad | 5,63 (3,87 to 7,94) |
| Diet low in vegetables | Burkina Faso | 5,58 (3,74 to 7,95) |
| Diet low in vegetables | Democratic Republic of Timor-Leste | 5,56 (3,71 to 7,61) |
| Diet low in vegetables | Republic of Mali | 5,55 (3,74 to 7,86) |
| Diet low in vegetables | Gabonese Republic | 5,52 (3,75 to 7,67) |
| Diet low in vegetables | Republic of Trinidad and Tobago | 5,51 (3,72 to 7,71) |
| Diet low in vegetables | State of Eritrea | 5,50 (3,68 to 7,92) |
| Diet low in vegetables | Republic of Uganda | 5,48 (3,75 to 7,50) |
| Diet low in vegetables | Independent State of Samoa | 5,47 (3,71 to 7,60) |
| Diet low in vegetables | Republic of Botswana | 5,44 (3,71 to 7,61) |
| Diet low in vegetables | Kingdom of Lesotho | 5,43 (3,56 to 7,69) |
| Diet low in vegetables | Republic of Côte d'Ivoire | 5,39 (3,66 to 7,51) |
| Diet low in vegetables | Islamic Republic of Afghanistan | 5,39 (3,71 to 7,44) |
| Diet low in vegetables | Kingdom of Tonga | 5,34 (3,59 to 7,40) |
| Diet low in vegetables | Republic of Senegal | 5,32 (3,54 to 7,57) |
| Diet low in vegetables | People's Republic of Bangladesh | 5,25 (3,59 to 7,20) |
| Diet low in vegetables | Tuvalu | 5,24 (3,58 to 7,29) |
| Diet low in vegetables | Republic of Panama | 5,23 (3,57 to 7,36) |
| Diet low in vegetables | Republic of Namibia | 5,23 (3,54 to 7,28) |
| Diet low in vegetables | Republic of Seychelles | 5,23 (3,55 to 7,27) |
| Diet low in vegetables | Islamic Republic of Pakistan | 5,22 (3,56 to 7,24) |
| Diet low in vegetables | Republic of Niue | 5,19 (3,55 to 7,14) |
| Diet low in vegetables | Republic of Nauru | 5,19 (3,47 to 7,32) |
| Diet low in vegetables | Mongolia | 5,18 (3,45 to 7,04) |
| Diet low in vegetables | Puerto Rico | 5,16 (3,48 to 7,14) |
| Diet low in vegetables | Republic of South Sudan | 5,14 (3,53 to 7,17) |
| Diet low in vegetables | Republic of Rwanda | 5,10 (3,50 to 7,06) |
| Diet low in vegetables | Republic of Costa Rica | 5,09 (3,47 to 7,07) |
| Diet low in vegetables | American Samoa | 5,09 (3,42 to 7,27) |
| Diet low in vegetables | Tokelau | 5,09 (3,35 to 7,10) |
| Diet low in vegetables | Grenada | 5,05 (3,42 to 7,05) |
| Diet low in vegetables | Republic of Fiji | 5,05 (3,36 to 7,27) |
| Diet low in vegetables | Republic of South Africa | 5,05 (3,41 to 7,01) |
| Diet low in vegetables | Republic of Zambia | 5,04 (3,46 to 7,08) |
| Diet low in vegetables | Solomon Islands | 5,03 (3,40 to 7,08) |
| Diet low in vegetables | Republic of Ghana | 5,01 (3,40 to 6,98) |
| Diet low in vegetables | Republic of Palau | 5,00 (3,31 to 6,93) |
| Diet low in vegetables | Malaysia | 4,98 (3,34 to 6,92) |
| Diet low in vegetables | United Republic of Tanzania | 4,93 (3,31 to 6,77) |
| Diet low in vegetables | Saint Lucia | 4,92 (3,35 to 6,89) |
| Diet low in vegetables | Republic of Djibouti | 4,92 (3,38 to 6,92) |
| Diet low in vegetables | Republic of Angola | 4,92 (3,25 to 6,88) |
| Diet low in vegetables | Republic of the Marshall Islands | 4,89 (3,27 to 6,87) |
| Diet low in vegetables | Republic of Guinea | 4,88 (3,26 to 6,79) |
| Diet low in vegetables | Republic of Paraguay | 4,87 (3,32 to 6,91) |
| Diet low in vegetables | Kingdom of Cambodia | 4,87 (3,28 to 6,89) |
| Diet low in vegetables | Republic of Kenya | 4,81 (3,29 to 6,71) |
| Diet low in vegetables | Republic of Yemen | 4,80 (3,27 to 6,56) |
| Diet low in vegetables | Bolivarian Republic of Venezuela | 4,76 (3,16 to 6,61) |
| Diet low in vegetables | Republic of Colombia | 4,76 (3,28 to 6,63) |
| Diet low in vegetables | Northern Mariana Islands | 4,72 (3,16 to 6,81) |
| Diet low in vegetables | Federal Democratic Republic of Ethiopia | 4,71 (3,19 to 6,53) |
| Diet low in vegetables | Democratic Socialist Republic of Sri Lanka | 4,70 (3,16 to 6,50) |
| Diet low in vegetables | Cook Islands | 4,69 (3,14 to 6,62) |
| Diet low in vegetables | Republic of Kiribati | 4,68 (3,14 to 6,57) |
| Diet low in vegetables | Federated States of Micronesia | 4,64 (3,18 to 6,56) |
| Diet low in vegetables | Belize | 4,61 (3,10 to 6,45) |
| Diet low in vegetables | Republic of Benin | 4,59 (3,10 to 6,57) |
| Diet low in vegetables | Republic of Honduras | 4,57 (3,14 to 6,44) |
| Diet low in vegetables | Saint Vincent and the Grenadines | 4,57 (3,03 to 6,43) |
| Diet low in vegetables | Federal Republic of Nigeria | 4,56 (3,08 to 6,53) |
| Diet low in vegetables | Saint Kitts and Nevis | 4,53 (2,97 to 6,29) |
| Diet low in vegetables | Republic of Ecuador | 4,51 (3,08 to 6,19) |
| Diet low in vegetables | Guam | 4,45 (2,99 to 6,16) |
| Diet low in vegetables | Republic of El Salvador | 4,44 (3,03 to 6,20) |
| Diet low in vegetables | Republic of the Niger | 4,43 (2,93 to 6,29) |
| Diet low in vegetables | Eastern Republic of Uruguay | 4,41 (3,04 to 6,24) |
| Diet low in vegetables | Federative Republic of Brazil | 4,38 (2,99 to 6,08) |
| Diet low in vegetables | Antigua and Barbuda | 4,35 (2,96 to 6,10) |
| Diet low in vegetables | Plurinational State of Bolivia | 4,33 (2,90 to 6,05) |
| Diet low in vegetables | Republic of Guyana | 4,33 (2,90 to 6,13) |
| Diet low in vegetables | Barbados | 4,32 (2,87 to 6,07) |
| Diet low in vegetables | Lao People's Democratic Republic | 4,28 (2,80 to 6,06) |
| Diet low in vegetables | Kingdom of Bhutan | 4,28 (2,82 to 6,05) |
| Diet low in vegetables | Republic of Guatemala | 4,25 (2,90 to 5,87) |
| Diet low in vegetables | Republic of Peru | 4,22 (2,87 to 5,91) |
| Diet low in vegetables | Republic of Suriname | 4,20 (2,87 to 5,88) |
| Diet low in vegetables | Brunei Darussalam | 4,18 (2,80 to 5,87) |
| Diet low in vegetables | Republic of Cameroon | 4,17 (2,73 to 5,92) |
| Diet low in vegetables | Republic of the Union of Myanmar | 4,16 (2,85 to 5,84) |
| Diet low in vegetables | Republic of Sudan | 4,12 (2,73 to 5,89) |
| Diet low in vegetables | Kingdom of Norway | 4,12 (2,76 to 5,84) |
| Diet low in vegetables | Kingdom of Thailand | 4,11 (2,75 to 5,65) |
| Diet low in vegetables | Dominican Republic | 4,04 (2,71 to 5,68) |
| Diet low in vegetables | Republic of Mauritius | 4,02 (2,67 to 5,68) |
| Diet low in vegetables | Republic of India | 4,00 (2,73 to 5,53) |
| Diet low in vegetables | Republic of the Philippines | 3,90 (2,59 to 5,54) |
| Diet low in vegetables | Republic of Moldova | 3,89 (2,54 to 5,59) |
| Diet low in vegetables | United Mexican States | 3,89 (2,62 to 5,48) |
| Diet low in vegetables | Independent State of Papua New Guinea | 3,88 (2,57 to 5,33) |
| Diet low in vegetables | Republic of Maldives | 3,84 (2,56 to 5,45) |
| Diet low in vegetables | Jamaica | 3,84 (2,56 to 5,33) |
| Diet low in vegetables | Republic of Iceland | 3,84 (2,52 to 5,40) |
| Diet low in vegetables | Czech Republic | 3,82 (2,51 to 5,59) |
| Diet low in vegetables | Republic of Finland | 3,80 (2,47 to 5,46) |
| Diet low in vegetables | United States Virgin Islands | 3,77 (2,51 to 5,28) |
| Diet low in vegetables | Kingdom of the Netherlands | 3,73 (2,56 to 5,31) |
| Diet low in vegetables | Republic of Slovenia | 3,73 (2,46 to 5,49) |
| Diet low in vegetables | Kingdom of Sweden | 3,72 (2,40 to 5,37) |
| Diet low in vegetables | French Republic | 3,68 (2,44 to 5,17) |
| Diet low in vegetables | Republic of Cyprus | 3,65 (2,36 to 5,14) |
| Diet low in vegetables | Federal Republic of Germany | 3,60 (2,40 to 5,19) |
| Diet low in vegetables | Commonwealth of Dominica | 3,60 (2,40 to 5,11) |
| Diet low in vegetables | Republic of Cabo Verde | 3,59 (2,34 to 5,33) |
| Diet low in vegetables | Federal Democratic Republic of Nepal | 3,58 (2,39 to 4,96) |
| Diet low in vegetables | Argentine Republic | 3,54 (2,36 to 5,09) |
| Diet low in vegetables | Republic of Croatia | 3,53 (2,27 to 5,13) |
| Diet low in vegetables | Georgia | 3,48 (2,20 to 5,07) |
| Diet low in vegetables | Socialist Republic of Viet Nam | 3,41 (2,21 to 4,92) |
| Diet low in vegetables | Syrian Arab Republic | 3,35 (2,21 to 4,87) |
| Diet low in vegetables | Commonwealth of the Bahamas | 3,27 (2,15 to 4,65) |
| Diet low in vegetables | Swiss Confederation | 3,23 (2,09 to 4,58) |
| Diet low in vegetables | Russian Federation | 3,20 (2,10 to 4,57) |
| Diet low in vegetables | Grand Duchy of Luxembourg | 3,17 (2,01 to 4,59) |
| Diet low in vegetables | Republic of Estonia | 3,16 (1,99 to 4,70) |
| Diet low in vegetables | Bermuda | 3,11 (2,04 to 4,48) |
| Diet low in vegetables | United Kingdom of Great Britain and Northern Ireland | 3,10 (2,03 to 4,45) |
| Diet low in vegetables | Republic of Serbia | 3,09 (1,97 to 4,51) |
| Diet low in vegetables | Slovak Republic | 3,09 (2,01 to 4,66) |
| Diet low in vegetables | Republic of Austria | 3,08 (2,00 to 4,39) |
| Diet low in vegetables | Republic of Latvia | 3,05 (1,97 to 4,50) |
| Diet low in vegetables | Australia | 3,01 (1,95 to 4,30) |
| Diet low in vegetables | Republic of Chile | 3,00 (1,90 to 4,42) |
| Diet low in vegetables | Kingdom of Denmark | 2,98 (1,85 to 4,58) |
| Diet low in vegetables | Republic of Lithuania | 2,98 (1,88 to 4,47) |
| Diet low in vegetables | Kingdom of Saudi Arabia | 2,95 (1,92 to 4,31) |
| Diet low in vegetables | New Zealand | 2,93 (1,90 to 4,18) |
| Diet low in vegetables | State of Libya | 2,85 (1,84 to 4,22) |
| Diet low in vegetables | Ireland | 2,84 (1,81 to 4,15) |
| Diet low in vegetables | People's Democratic Republic of Algeria | 2,80 (1,80 to 4,04) |
| Diet low in vegetables | Democratic People's Republic of Korea | 2,78 (1,79 to 4,00) |
| Diet low in vegetables | United States of America | 2,78 (1,83 to 3,97) |
| Diet low in vegetables | Palestine | 2,76 (1,80 to 4,01) |
| Diet low in vegetables | Republic of San Marino | 2,71 (1,64 to 4,13) |
| Diet low in vegetables | Hashemite Kingdom of Jordan | 2,69 (1,69 to 3,91) |
| Diet low in vegetables | Canada | 2,65 (1,68 to 3,82) |
| Diet low in vegetables | United Arab Emirates | 2,58 (1,59 to 3,88) |
| Diet low in vegetables | Principality of Andorra | 2,49 (1,49 to 3,77) |
| Diet low in vegetables | Hungary | 2,47 (1,46 to 3,96) |
| Diet low in vegetables | Sultanate of Oman | 2,46 (1,50 to 3,67) |
| Diet low in vegetables | Republic of Cuba | 2,43 (1,56 to 3,52) |
| Diet low in vegetables | Kingdom of Belgium | 2,24 (1,35 to 3,41) |
| Diet low in vegetables | Kingdom of Spain | 2,23 (1,34 to 3,37) |
| Diet low in vegetables | Republic of Italy | 2,15 (1,35 to 3,19) |
| Diet low in vegetables | Kingdom of Morocco | 2,04 (1,17 to 3,19) |
| Diet low in vegetables | Greenland | 1,98 (1,12 to 3,16) |
| Diet low in vegetables | Taiwan (Province of China) | 1,73 (0,99 to 2,70) |
| Diet low in vegetables | Bosnia and Herzegovina | 1,73 (0,92 to 2,81) |
| Diet low in vegetables | Republic of Iraq | 1,71 (0,91 to 2,73) |
| Diet low in vegetables | Republic of Korea | 1,71 (1,03 to 2,66) |
| Diet low in vegetables | Republic of Singapore | 1,64 (0,96 to 2,51) |
| Diet low in vegetables | Ukraine | 1,54 (0,80 to 2,61) |
| Diet low in vegetables | Republic of Malta | 1,51 (0,77 to 2,52) |
| Diet low in vegetables | Republic of Tajikistan | 1,48 (0,76 to 2,48) |
| Diet low in vegetables | Republic of Belarus | 1,46 (0,75 to 2,52) |
| Diet low in vegetables | Portuguese Republic | 1,36 (0,71 to 2,25) |
| Diet low in vegetables | Kyrgyz Republic | 1,30 (0,65 to 2,20) |
| Diet low in vegetables | Japan | 1,29 (0,72 to 2,04) |
| Diet low in vegetables | Kingdom of Bahrain | 1,26 (0,61 to 2,14) |
| Diet low in vegetables | Republic of Poland | 1,25 (0,66 to 2,10) |
| Diet low in vegetables | Republic of Tunisia | 1,15 (0,56 to 2,01) |
| Diet low in vegetables | Republic of Kazakhstan | 0,95 (0,41 to 1,79) |
| Diet low in vegetables | Islamic Republic of Iran | 0,93 (0,49 to 1,56) |
| Diet low in vegetables | Republic of Bulgaria | 0,93 (0,44 to 1,68) |
| Diet low in vegetables | State of Kuwait | 0,84 (0,33 to 1,53) |
| Diet low in vegetables | North Macedonia | 0,68 (0,25 to 1,33) |
| Diet low in vegetables | Hellenic Republic | 0,59 (0,22 to 1,20) |
| Diet low in vegetables | Principality of Monaco | 0,56 (0,16 to 1,17) |
| Diet low in vegetables | Republic of Azerbaijan | 0,53 (0,17 to 1,15) |
| Diet low in vegetables | Republic of Albania | 0,53 (0,15 to 1,11) |
| Diet low in vegetables | Montenegro | 0,49 (0,18 to 1,03) |
| Diet low in vegetables | Turkmenistan | 0,41 (0,12 to 0,84) |
| Diet low in vegetables | State of Israel | 0,40 (0,12 to 0,88) |
| Diet low in vegetables | People's Republic of China | 0,33 (0,14 to 0,63) |
| Diet low in vegetables | Lebanese Republic | 0,29 (0,07 to 0,65) |
| Diet low in vegetables | State of Qatar | 0,23 (0,04 to 0,59) |
| Diet low in vegetables | Republic of Uzbekistan | 0,15 (0,03 to 0,39) |
| Diet low in vegetables | Romania | 0,15 (0,03 to 0,38) |
| Diet low in vegetables | Arab Republic of Egypt | 0,11 (0,01 to 0,29) |
| Diet low in vegetables | Republic of Armenia | 0,10 (0,01 to 0,28) |
| Diet low in vegetables | Republic of Turkey | 0,04 (0,00 to 0,15) |

**High body-mass index**

| **Risk factor** | **Location** | **Percent contribution to ASDR (%)** |
| --- | --- | --- |
| High body-mass index | State of Qatar | 14,28 (7,71 to 24,22) |
| High body-mass index | State of Kuwait | 14,20 (7,53 to 23,42) |
| High body-mass index | Hungary | 14,15 (7,62 to 23,35) |
| High body-mass index | Kingdom of Saudi Arabia | 13,72 (7,21 to 23,44) |
| High body-mass index | Republic of Moldova | 13,72 (7,39 to 22,85) |
| High body-mass index | United Arab Emirates | 13,33 (7,08 to 22,36) |
| High body-mass index | State of Libya | 13,25 (6,98 to 22,52) |
| High body-mass index | Syrian Arab Republic | 13,15 (6,90 to 22,56) |
| High body-mass index | Montenegro | 13,07 (6,80 to 22,00) |
| High body-mass index | Hashemite Kingdom of Jordan | 12,95 (7,01 to 22,36) |
| High body-mass index | American Samoa | 12,94 (7,04 to 21,96) |
| High body-mass index | Kingdom of Tonga | 12,88 (7,00 to 21,35) |
| High body-mass index | Arab Republic of Egypt | 12,86 (6,91 to 21,67) |
| High body-mass index | Republic of Iraq | 12,73 (6,71 to 21,88) |
| High body-mass index | Republic of Lithuania | 12,45 (6,49 to 21,18) |
| High body-mass index | Sultanate of Oman | 12,43 (6,72 to 20,96) |
| High body-mass index | Republic of Chile | 12,35 (6,40 to 21,43) |
| High body-mass index | Slovak Republic | 12,25 (6,41 to 20,92) |
| High body-mass index | Republic of Fiji | 12,20 (6,59 to 20,53) |
| High body-mass index | Kingdom of Eswatini | 12,17 (6,58 to 21,19) |
| High body-mass index | Republic of Panama | 12,06 (6,33 to 20,76) |
| High body-mass index | Republic of Slovenia | 12,05 (6,21 to 20,81) |
| High body-mass index | Cook Islands | 12,00 (6,26 to 19,84) |
| High body-mass index | Republic of Latvia | 12,00 (6,31 to 20,48) |
| High body-mass index | Republic of Belarus | 11,94 (6,28 to 20,70) |
| High body-mass index | Kingdom of Bahrain | 11,85 (6,22 to 20,19) |
| High body-mass index | Republic of Croatia | 11,84 (6,23 to 20,14) |
| High body-mass index | Republic of Nauru | 11,76 (5,72 to 21,32) |
| High body-mass index | Northern Mariana Islands | 11,74 (6,39 to 19,70) |
| High body-mass index | Romania | 11,74 (6,20 to 20,07) |
| High body-mass index | Republic of Serbia | 11,63 (6,23 to 19,83) |
| High body-mass index | Republic of Estonia | 11,60 (6,04 to 19,86) |
| High body-mass index | Ukraine | 11,59 (6,23 to 19,70) |
| High body-mass index | Czech Republic | 11,54 (5,91 to 19,78) |
| High body-mass index | Lebanese Republic | 11,42 (5,93 to 19,79) |
| High body-mass index | Republic of Palau | 11,39 (6,08 to 19,59) |
| High body-mass index | Russian Federation | 11,24 (5,88 to 19,46) |
| High body-mass index | Independent State of Samoa | 11,23 (5,98 to 19,78) |
| High body-mass index | Republic of Turkey | 11,22 (6,02 to 18,97) |
| High body-mass index | Republic of Bulgaria | 11,19 (5,91 to 18,98) |
| High body-mass index | Republic of Niue | 11,15 (5,82 to 19,33) |
| High body-mass index | Bolivarian Republic of Venezuela | 11,13 (5,70 to 18,95) |
| High body-mass index | Republic of South Africa | 11,12 (5,89 to 19,05) |
| High body-mass index | Commonwealth of Dominica | 11,04 (5,78 to 18,89) |
| High body-mass index | Palestine | 10,98 (5,72 to 19,00) |
| High body-mass index | Republic of Kazakhstan | 10,95 (5,96 to 18,23) |
| High body-mass index | Puerto Rico | 10,79 (5,74 to 17,93) |
| High body-mass index | United States of America | 10,76 (5,80 to 18,29) |
| High body-mass index | Guam | 10,63 (5,71 to 17,86) |
| High body-mass index | North Macedonia | 10,62 (5,52 to 18,11) |
| High body-mass index | Republic of Albania | 10,58 (5,46 to 17,97) |
| High body-mass index | Republic of El Salvador | 10,58 (5,56 to 18,38) |
| High body-mass index | Republic of Azerbaijan | 10,43 (5,69 to 17,80) |
| High body-mass index | Bermuda | 10,41 (5,40 to 17,66) |
| High body-mass index | Republic of Nicaragua | 10,34 (5,43 to 17,78) |
| High body-mass index | Argentine Republic | 10,33 (5,36 to 17,35) |
| High body-mass index | Tokelau | 10,32 (5,09 to 18,14) |
| High body-mass index | United Mexican States | 10,18 (5,37 to 17,34) |
| High body-mass index | Republic of Sudan | 10,14 (5,55 to 16,88) |
| High body-mass index | Georgia | 10,10 (5,46 to 17,41) |
| High body-mass index | Bosnia and Herzegovina | 10,10 (5,33 to 17,16) |
| High body-mass index | Belize | 10,01 (5,20 to 17,23) |
| High body-mass index | Republic of Costa Rica | 9,99 (5,28 to 17,64) |
| High body-mass index | United States Virgin Islands | 9,96 (5,20 to 17,31) |
| High body-mass index | Tuvalu | 9,96 (5,25 to 17,15) |
| High body-mass index | Republic of Cameroon | 9,95 (5,34 to 16,87) |
| High body-mass index | Republic of Trinidad and Tobago | 9,90 (5,13 to 16,80) |
| High body-mass index | Kingdom of Spain | 9,85 (5,20 to 17,40) |
| High body-mass index | Barbados | 9,81 (5,23 to 17,26) |
| High body-mass index | Federal Republic of Germany | 9,80 (5,14 to 16,83) |
| High body-mass index | Commonwealth of the Bahamas | 9,72 (5,17 to 16,70) |
| High body-mass index | Republic of the Marshall Islands | 9,67 (5,02 to 16,48) |
| High body-mass index | Republic of Armenia | 9,67 (5,25 to 16,37) |
| High body-mass index | Republic of Colombia | 9,64 (5,16 to 16,30) |
| High body-mass index | Kyrgyz Republic | 9,64 (5,10 to 16,77) |
| High body-mass index | Eastern Republic of Uruguay | 9,59 (5,08 to 16,46) |
| High body-mass index | Republic of Paraguay | 9,55 (4,99 to 17,17) |
| High body-mass index | Republic of Poland | 9,53 (5,00 to 16,69) |
| High body-mass index | Principality of Monaco | 9,51 (4,95 to 16,55) |
| High body-mass index | Islamic Republic of Mauritania | 9,51 (4,92 to 16,44) |
| High body-mass index | Australia | 9,46 (5,00 to 16,27) |
| High body-mass index | Turkmenistan | 9,43 (4,93 to 16,15) |
| High body-mass index | Federative Republic of Brazil | 9,41 (4,95 to 16,40) |
| High body-mass index | Republic of Finland | 9,35 (4,91 to 16,57) |
| High body-mass index | Grand Duchy of Luxembourg | 9,30 (4,83 to 15,99) |
| High body-mass index | Republic of San Marino | 9,22 (4,75 to 16,41) |
| High body-mass index | New Zealand | 9,20 (4,73 to 15,82) |
| High body-mass index | Islamic Republic of Iran | 9,19 (4,95 to 15,72) |
| High body-mass index | Republic of Uzbekistan | 9,14 (4,92 to 16,03) |
| High body-mass index | Republic of Kiribati | 9,04 (4,79 to 15,54) |
| High body-mass index | Federated States of Micronesia | 9,02 (4,77 to 16,01) |
| High body-mass index | Republic of Iceland | 9,01 (4,60 to 15,72) |
| High body-mass index | United Kingdom of Great Britain and Northern Ireland | 8,97 (4,68 to 15,61) |
| High body-mass index | Ireland | 8,95 (4,58 to 16,00) |
| High body-mass index | People's Democratic Republic of Algeria | 8,89 (4,59 to 15,32) |
| High body-mass index | State of Israel | 8,89 (4,67 to 15,60) |
| High body-mass index | Saint Kitts and Nevis | 8,83 (4,53 to 15,88) |
| High body-mass index | Gabonese Republic | 8,77 (4,48 to 15,56) |
| High body-mass index | Hellenic Republic | 8,73 (4,60 to 15,34) |
| High body-mass index | Kingdom of Morocco | 8,73 (4,74 to 14,88) |
| High body-mass index | Canada | 8,71 (4,64 to 15,53) |
| High body-mass index | Republic of Austria | 8,66 (4,58 to 15,41) |
| High body-mass index | Republic of Guatemala | 8,64 (4,58 to 14,91) |
| High body-mass index | Greenland | 8,53 (4,38 to 15,00) |
| High body-mass index | Republic of Ecuador | 8,53 (4,55 to 14,81) |
| High body-mass index | Jamaica | 8,52 (4,48 to 14,61) |
| High body-mass index | Mongolia | 8,51 (4,49 to 14,40) |
| High body-mass index | Republic of Tunisia | 8,47 (4,51 to 15,31) |
| High body-mass index | Republic of Tajikistan | 8,34 (4,50 to 14,12) |
| High body-mass index | French Republic | 8,31 (4,31 to 15,08) |
| High body-mass index | Republic of Honduras | 8,26 (4,43 to 14,28) |
| High body-mass index | Kingdom of Belgium | 8,24 (4,42 to 14,35) |
| High body-mass index | Portuguese Republic | 8,19 (4,23 to 14,33) |
| High body-mass index | Republic of Cyprus | 8,16 (4,32 to 14,18) |
| High body-mass index | Federal Republic of Nigeria | 8,10 (4,46 to 13,63) |
| High body-mass index | Republic of Malta | 8,08 (4,22 to 14,67) |
| High body-mass index | Republic of Equatorial Guinea | 8,06 (4,44 to 13,67) |
| High body-mass index | Antigua and Barbuda | 8,06 (4,26 to 13,74) |
| High body-mass index | Republic of Namibia | 8,05 (4,15 to 14,13) |
| High body-mass index | Kingdom of Sweden | 8,04 (4,20 to 13,68) |
| High body-mass index | Republic of Vanuatu | 8,02 (4,22 to 14,04) |
| High body-mass index | Republic of Liberia | 7,98 (4,08 to 13,94) |
| High body-mass index | Kingdom of Lesotho | 7,98 (4,14 to 13,83) |
| High body-mass index | Republic of Botswana | 7,95 (4,08 to 14,16) |
| High body-mass index | Kingdom of Denmark | 7,95 (4,18 to 13,95) |
| High body-mass index | Democratic Republic of Sao Tome and Principe | 7,94 (4,15 to 13,82) |
| High body-mass index | Principality of Andorra | 7,91 (4,22 to 13,69) |
| High body-mass index | Republic of Seychelles | 7,86 (4,25 to 13,13) |
| High body-mass index | Kingdom of the Netherlands | 7,82 (3,95 to 13,19) |
| High body-mass index | Republic of Italy | 7,73 (4,04 to 13,63) |
| High body-mass index | Republic of Guyana | 7,63 (3,96 to 12,89) |
| High body-mass index | Kingdom of Norway | 7,61 (4,09 to 13,56) |
| High body-mass index | Republic of Peru | 7,50 (4,04 to 12,90) |
| High body-mass index | Grenada | 7,26 (3,80 to 12,34) |
| High body-mass index | Republic of Zimbabwe | 7,16 (3,79 to 12,18) |
| High body-mass index | Saint Lucia | 7,07 (3,81 to 12,09) |
| High body-mass index | Dominican Republic | 6,99 (3,83 to 11,89) |
| High body-mass index | Swiss Confederation | 6,82 (3,67 to 12,12) |
| High body-mass index | Republic of Cabo Verde | 6,79 (3,57 to 11,76) |
| High body-mass index | Plurinational State of Bolivia | 6,67 (3,52 to 11,14) |
| High body-mass index | Republic of Cuba | 6,57 (3,50 to 11,48) |
| High body-mass index | Islamic Republic of Afghanistan | 6,52 (3,52 to 11,49) |
| High body-mass index | Republic of Côte d'Ivoire | 6,42 (3,39 to 11,42) |
| High body-mass index | Republic of Yemen | 6,41 (3,52 to 10,90) |
| High body-mass index | Republic of Mauritius | 6,33 (3,40 to 11,36) |
| High body-mass index | Republic of Ghana | 6,29 (3,29 to 10,82) |
| High body-mass index | Togolese Republic | 6,17 (3,29 to 11,15) |
| High body-mass index | Republic of the Gambia | 6,10 (3,13 to 10,80) |
| High body-mass index | Taiwan (Province of China) | 6,07 (3,29 to 10,44) |
| High body-mass index | Malaysia | 5,97 (3,23 to 9,99) |
| High body-mass index | Saint Vincent and the Grenadines | 5,96 (3,20 to 10,44) |
| High body-mass index | Republic of Senegal | 5,88 (3,17 to 9,74) |
| High body-mass index | Republic of the Congo | 5,86 (3,13 to 9,83) |
| High body-mass index | Republic of Suriname | 5,80 (3,12 to 9,86) |
| High body-mass index | People's Republic of China | 5,71 (3,12 to 9,70) |
| High body-mass index | Republic of Sierra Leone | 5,57 (2,92 to 9,66) |
| High body-mass index | Solomon Islands | 5,54 (2,90 to 9,70) |
| High body-mass index | Union of the Comoros | 5,51 (3,02 to 9,56) |
| High body-mass index | Republic of Benin | 5,50 (2,86 to 9,79) |
| High body-mass index | United Republic of Tanzania | 5,28 (2,76 to 9,19) |
| High body-mass index | Republic of Kenya | 5,19 (2,82 to 8,93) |
| High body-mass index | Islamic Republic of Pakistan | 5,14 (2,81 to 8,56) |
| High body-mass index | Republic of Guinea-Bissau | 5,06 (2,73 to 8,50) |
| High body-mass index | Democratic Republic of the Congo | 5,05 (2,74 to 8,39) |
| High body-mass index | Kingdom of Bhutan | 4,99 (2,73 to 8,36) |
| High body-mass index | Brunei Darussalam | 4,85 (2,72 to 7,95) |
| High body-mass index | Republic of Malawi | 4,82 (2,59 to 8,10) |
| High body-mass index | Republic of Singapore | 4,61 (2,43 to 7,86) |
| High body-mass index | Republic of Zambia | 4,58 (2,47 to 7,66) |
| High body-mass index | Japan | 4,57 (2,49 to 7,25) |
| High body-mass index | Republic of Angola | 4,41 (2,45 to 7,45) |
| High body-mass index | Republic of Guinea | 4,23 (2,32 to 7,41) |
| High body-mass index | Kingdom of Thailand | 4,06 (2,16 to 6,60) |
| High body-mass index | Republic of Maldives | 4,06 (2,12 to 6,83) |
| High body-mass index | Republic of Mozambique | 4,05 (2,20 to 6,86) |
| High body-mass index | Democratic Socialist Republic of Sri Lanka | 4,04 (2,23 to 6,84) |
| High body-mass index | Independent State of Papua New Guinea | 4,02 (2,10 to 6,92) |
| High body-mass index | Republic of Mali | 3,95 (1,92 to 7,34) |
| High body-mass index | Republic of the Philippines | 3,89 (2,14 to 6,34) |
| High body-mass index | Democratic People's Republic of Korea | 3,86 (2,00 to 6,67) |
| High body-mass index | Republic of the Niger | 3,83 (1,79 to 7,39) |
| High body-mass index | Republic of Indonesia | 3,78 (2,07 to 5,77) |
| High body-mass index | Republic of Korea | 3,78 (2,01 to 6,14) |
| High body-mass index | Central African Republic | 3,76 (2,09 to 6,39) |
| High body-mass index | Republic of Haiti | 3,70 (2,06 to 6,23) |
| High body-mass index | Republic of Chad | 3,68 (1,90 to 6,60) |
| High body-mass index | Republic of Uganda | 3,51 (1,88 to 5,72) |
| High body-mass index | Republic of Madagascar | 3,40 (1,80 to 5,77) |
| High body-mass index | Federal Republic of Somalia | 3,31 (1,70 to 5,52) |
| High body-mass index | Republic of Rwanda | 3,27 (1,77 to 5,42) |
| High body-mass index | Republic of India | 3,14 (1,79 to 5,09) |
| High body-mass index | Lao People's Democratic Republic | 2,89 (1,55 to 4,78) |
| High body-mass index | Republic of Djibouti | 2,86 (1,48 to 4,61) |
| High body-mass index | Federal Democratic Republic of Ethiopia | 2,75 (1,49 to 4,57) |
| High body-mass index | Burkina Faso | 2,67 (1,43 to 4,40) |
| High body-mass index | Republic of the Union of Myanmar | 2,64 (1,42 to 4,18) |
| High body-mass index | Republic of Burundi | 2,51 (1,34 to 4,40) |
| High body-mass index | Republic of South Sudan | 2,36 (1,28 to 3,97) |
| High body-mass index | People's Republic of Bangladesh | 2,33 (1,26 to 3,81) |
| High body-mass index | State of Eritrea | 2,26 (1,20 to 3,89) |
| High body-mass index | Socialist Republic of Viet Nam | 2,17 (1,17 to 3,49) |
| High body-mass index | Federal Democratic Republic of Nepal | 2,15 (1,17 to 3,43) |
| High body-mass index | Kingdom of Cambodia | 1,97 (1,06 to 3,12) |
| High body-mass index | Democratic Republic of Timor-Leste | 1,76 (0,94 to 2,86) |

**High systolic blood pressure**

| **Risk factor** | **Location** | **Percent contribution to ASDR (%)** |
| --- | --- | --- |
| High systolic blood pressure | Republic of Indonesia | 26,26 (20,01 to 32,45) |
| High systolic blood pressure | Hungary | 25,90 (19,96 to 32,82) |
| High systolic blood pressure | Republic of Moldova | 24,72 (18,33 to 31,43) |
| High systolic blood pressure | Republic of Sierra Leone | 24,41 (18,01 to 31,39) |
| High systolic blood pressure | Republic of Kazakhstan | 23,87 (18,45 to 29,84) |
| High systolic blood pressure | Republic of Lithuania | 23,43 (17,71 to 29,46) |
| High systolic blood pressure | Malaysia | 23,41 (17,36 to 29,76) |
| High systolic blood pressure | Georgia | 22,78 (16,54 to 29,17) |
| High systolic blood pressure | Republic of Latvia | 22,67 (16,90 to 28,72) |
| High systolic blood pressure | Republic of Belarus | 22,62 (16,97 to 28,69) |
| High systolic blood pressure | Romania | 22,48 (16,97 to 28,18) |
| High systolic blood pressure | Republic of Vanuatu | 22,43 (16,58 to 29,13) |
| High systolic blood pressure | Republic of Serbia | 22,36 (16,79 to 28,23) |
| High systolic blood pressure | Democratic Republic of Sao Tome and Principe | 22,32 (16,91 to 28,30) |
| High systolic blood pressure | Republic of Estonia | 22,31 (16,34 to 28,21) |
| High systolic blood pressure | Federal Republic of Nigeria | 22,30 (16,50 to 28,23) |
| High systolic blood pressure | Republic of Zimbabwe | 22,19 (16,49 to 28,41) |
| High systolic blood pressure | Togolese Republic | 22,18 (16,73 to 28,22) |
| High systolic blood pressure | Republic of Cameroon | 22,07 (16,27 to 28,19) |
| High systolic blood pressure | Republic of Senegal | 21,97 (16,28 to 27,95) |
| High systolic blood pressure | Republic of Croatia | 21,70 (15,88 to 27,85) |
| High systolic blood pressure | Ukraine | 21,67 (16,09 to 27,90) |
| High systolic blood pressure | Republic of Cabo Verde | 21,62 (15,78 to 27,44) |
| High systolic blood pressure | Republic of Iraq | 21,56 (16,38 to 27,11) |
| High systolic blood pressure | Republic of Slovenia | 21,50 (15,71 to 27,83) |
| High systolic blood pressure | Republic of Albania | 21,49 (15,99 to 27,03) |
| High systolic blood pressure | Montenegro | 21,40 (15,87 to 27,79) |
| High systolic blood pressure | Slovak Republic | 21,39 (15,92 to 27,40) |
| High systolic blood pressure | Republic of Chile | 21,33 (15,99 to 26,86) |
| High systolic blood pressure | Bosnia and Herzegovina | 21,30 (15,35 to 27,55) |
| High systolic blood pressure | Republic of Trinidad and Tobago | 21,25 (15,60 to 26,90) |
| High systolic blood pressure | Socialist Republic of Viet Nam | 21,18 (15,42 to 26,97) |
| High systolic blood pressure | Republic of the Gambia | 21,10 (15,24 to 27,34) |
| High systolic blood pressure | Mongolia | 20,98 (14,87 to 26,68) |
| High systolic blood pressure | Republic of Bulgaria | 20,96 (15,92 to 27,28) |
| High systolic blood pressure | North Macedonia | 20,94 (15,26 to 27,24) |
| High systolic blood pressure | Republic of Côte d'Ivoire | 20,92 (15,49 to 27,13) |
| High systolic blood pressure | Republic of South Africa | 20,84 (15,49 to 26,64) |
| High systolic blood pressure | Turkmenistan | 20,74 (15,50 to 26,44) |
| High systolic blood pressure | Republic of Liberia | 20,71 (15,27 to 26,56) |
| High systolic blood pressure | Republic of Sudan | 20,61 (15,54 to 26,05) |
| High systolic blood pressure | Republic of Colombia | 20,60 (15,35 to 26,41) |
| High systolic blood pressure | Islamic Republic of Mauritania | 20,47 (14,14 to 26,66) |
| High systolic blood pressure | Republic of Equatorial Guinea | 20,47 (14,73 to 26,86) |
| High systolic blood pressure | Republic of Ghana | 20,27 (14,75 to 25,81) |
| High systolic blood pressure | Russian Federation | 20,24 (15,31 to 25,31) |
| High systolic blood pressure | Republic of Azerbaijan | 20,18 (14,62 to 25,94) |
| High systolic blood pressure | Republic of Malawi | 20,07 (14,47 to 26,24) |
| High systolic blood pressure | Federal Republic of Germany | 20,02 (14,35 to 25,45) |
| High systolic blood pressure | Republic of Mozambique | 20,00 (14,57 to 25,69) |
| High systolic blood pressure | Kingdom of Morocco | 19,96 (15,00 to 25,61) |
| High systolic blood pressure | Kingdom of Denmark | 19,95 (14,44 to 26,56) |
| High systolic blood pressure | Republic of Botswana | 19,92 (14,07 to 26,05) |
| High systolic blood pressure | Republic of the Congo | 19,92 (14,07 to 25,84) |
| High systolic blood pressure | Republic of Armenia | 19,85 (14,77 to 25,85) |
| High systolic blood pressure | Bolivarian Republic of Venezuela | 19,83 (14,20 to 25,93) |
| High systolic blood pressure | Czech Republic | 19,73 (14,72 to 25,99) |
| High systolic blood pressure | Grand Duchy of Luxembourg | 19,72 (13,84 to 25,13) |
| High systolic blood pressure | Republic of Costa Rica | 19,71 (14,87 to 25,12) |
| High systolic blood pressure | Republic of Finland | 19,70 (14,27 to 25,37) |
| High systolic blood pressure | Kingdom of Eswatini | 19,64 (14,12 to 25,82) |
| High systolic blood pressure | Gabonese Republic | 19,59 (14,09 to 25,41) |
| High systolic blood pressure | Republic of Guinea-Bissau | 19,56 (14,15 to 25,18) |
| High systolic blood pressure | State of Libya | 19,41 (14,32 to 24,73) |
| High systolic blood pressure | State of Israel | 19,36 (14,40 to 24,96) |
| High systolic blood pressure | Kingdom of Norway | 19,35 (14,09 to 25,25) |
| High systolic blood pressure | Republic of Benin | 19,34 (13,85 to 25,46) |
| High systolic blood pressure | Republic of Seychelles | 19,30 (13,81 to 25,75) |
| High systolic blood pressure | Republic of the Niger | 19,29 (13,60 to 26,78) |
| High systolic blood pressure | Republic of Kenya | 19,28 (14,60 to 24,54) |
| High systolic blood pressure | Kingdom of Belgium | 19,25 (14,36 to 24,31) |
| High systolic blood pressure | Republic of Fiji | 19,22 (13,87 to 25,08) |
| High systolic blood pressure | Republic of Panama | 19,20 (13,90 to 24,54) |
| High systolic blood pressure | Republic of Angola | 19,19 (14,08 to 25,33) |
| High systolic blood pressure | Republic of Djibouti | 18,98 (13,99 to 24,26) |
| High systolic blood pressure | Republic of Mali | 18,94 (13,36 to 25,17) |
| High systolic blood pressure | Barbados | 18,92 (13,99 to 24,56) |
| High systolic blood pressure | Republic of Mauritius | 18,88 (13,64 to 24,69) |
| High systolic blood pressure | State of Qatar | 18,87 (13,91 to 24,35) |
| High systolic blood pressure | Republic of Austria | 18,78 (13,57 to 24,44) |
| High systolic blood pressure | Republic of Nicaragua | 18,77 (13,55 to 24,28) |
| High systolic blood pressure | Republic of Niue | 18,77 (13,88 to 23,89) |
| High systolic blood pressure | Democratic Republic of Timor-Leste | 18,72 (13,59 to 24,15) |
| High systolic blood pressure | Republic of Tajikistan | 18,68 (13,42 to 24,43) |
| High systolic blood pressure | Sultanate of Oman | 18,68 (13,75 to 24,34) |
| High systolic blood pressure | Ireland | 18,68 (13,59 to 24,65) |
| High systolic blood pressure | Democratic Socialist Republic of Sri Lanka | 18,63 (13,51 to 24,17) |
| High systolic blood pressure | Kingdom of the Netherlands | 18,61 (13,56 to 25,11) |
| High systolic blood pressure | Republic of Cyprus | 18,59 (13,49 to 23,81) |
| High systolic blood pressure | Principality of Andorra | 18,54 (13,02 to 24,44) |
| High systolic blood pressure | American Samoa | 18,48 (12,87 to 23,90) |
| High systolic blood pressure | Republic of Maldives | 18,47 (13,34 to 23,99) |
| High systolic blood pressure | Republic of Haiti | 18,45 (13,26 to 24,04) |
| High systolic blood pressure | Kingdom of Sweden | 18,40 (13,09 to 23,89) |
| High systolic blood pressure | Union of the Comoros | 18,39 (13,58 to 23,78) |
| High systolic blood pressure | French Republic | 18,36 (13,50 to 23,63) |
| High systolic blood pressure | Republic of San Marino | 18,35 (12,94 to 24,16) |
| High systolic blood pressure | Republic of Nauru | 18,27 (12,30 to 24,81) |
| High systolic blood pressure | Central African Republic | 18,22 (12,64 to 24,38) |
| High systolic blood pressure | Islamic Republic of Pakistan | 18,22 (13,20 to 23,52) |
| High systolic blood pressure | Republic of the Union of Myanmar | 18,21 (13,23 to 24,03) |
| High systolic blood pressure | Republic of Burundi | 18,10 (12,36 to 25,21) |
| High systolic blood pressure | Burkina Faso | 18,09 (12,69 to 24,17) |
| High systolic blood pressure | Principality of Monaco | 18,07 (13,29 to 23,07) |
| High systolic blood pressure | Republic of Namibia | 18,00 (13,02 to 23,58) |
| High systolic blood pressure | Republic of Malta | 17,99 (12,82 to 24,18) |
| High systolic blood pressure | Kingdom of Tonga | 17,80 (12,46 to 23,83) |
| High systolic blood pressure | Japan | 17,64 (13,11 to 22,97) |
| High systolic blood pressure | Northern Mariana Islands | 17,63 (12,50 to 23,72) |
| High systolic blood pressure | Saint Kitts and Nevis | 17,63 (12,46 to 23,00) |
| High systolic blood pressure | Republic of El Salvador | 17,63 (12,60 to 23,05) |
| High systolic blood pressure | Kyrgyz Republic | 17,57 (12,33 to 22,95) |
| High systolic blood pressure | Kingdom of Bahrain | 17,51 (11,74 to 23,00) |
| High systolic blood pressure | Cook Islands | 17,51 (12,35 to 23,27) |
| High systolic blood pressure | Republic of Paraguay | 17,48 (12,01 to 22,82) |
| High systolic blood pressure | Puerto Rico | 17,46 (12,50 to 23,34) |
| High systolic blood pressure | Republic of Uganda | 17,44 (12,69 to 22,43) |
| High systolic blood pressure | Federative Republic of Brazil | 17,43 (13,06 to 22,13) |
| High systolic blood pressure | Republic of Poland | 17,39 (12,66 to 22,11) |
| High systolic blood pressure | Republic of Honduras | 17,37 (12,55 to 22,59) |
| High systolic blood pressure | Republic of Uzbekistan | 17,30 (12,41 to 22,22) |
| High systolic blood pressure | Republic of Iceland | 17,27 (12,28 to 22,76) |
| High systolic blood pressure | Republic of Palau | 17,25 (12,07 to 23,73) |
| High systolic blood pressure | Guam | 17,22 (12,13 to 23,27) |
| High systolic blood pressure | Argentine Republic | 17,19 (12,66 to 22,60) |
| High systolic blood pressure | Kingdom of Spain | 17,13 (12,09 to 22,29) |
| High systolic blood pressure | Republic of Madagascar | 17,12 (11,83 to 22,59) |
| High systolic blood pressure | Lebanese Republic | 17,11 (12,49 to 21,80) |
| High systolic blood pressure | Kingdom of Lesotho | 17,10 (11,38 to 23,68) |
| High systolic blood pressure | United Arab Emirates | 17,09 (12,14 to 22,16) |
| High systolic blood pressure | Lao People's Democratic Republic | 17,05 (12,17 to 22,95) |
| High systolic blood pressure | Eastern Republic of Uruguay | 17,05 (12,37 to 22,14) |
| High systolic blood pressure | Grenada | 16,86 (11,92 to 22,35) |
| High systolic blood pressure | Commonwealth of the Bahamas | 16,86 (12,08 to 22,71) |
| High systolic blood pressure | Syrian Arab Republic | 16,83 (11,97 to 22,14) |
| High systolic blood pressure | Republic of South Sudan | 16,83 (11,77 to 22,74) |
| High systolic blood pressure | Tokelau | 16,82 (11,27 to 23,08) |
| High systolic blood pressure | People's Republic of Bangladesh | 16,81 (12,07 to 22,06) |
| High systolic blood pressure | United Mexican States | 16,80 (12,15 to 21,47) |
| High systolic blood pressure | State of Eritrea | 16,74 (11,48 to 22,60) |
| High systolic blood pressure | Republic of Guinea | 16,74 (11,95 to 21,80) |
| High systolic blood pressure | Saint Lucia | 16,71 (11,67 to 22,04) |
| High systolic blood pressure | Democratic Republic of the Congo | 16,70 (11,84 to 21,61) |
| High systolic blood pressure | Antigua and Barbuda | 16,69 (11,81 to 21,55) |
| High systolic blood pressure | Arab Republic of Egypt | 16,67 (11,94 to 21,33) |
| High systolic blood pressure | Federal Republic of Somalia | 16,67 (10,87 to 23,72) |
| High systolic blood pressure | Republic of Chad | 16,64 (11,78 to 22,35) |
| High systolic blood pressure | Commonwealth of Dominica | 16,61 (11,60 to 22,07) |
| High systolic blood pressure | Republic of Guyana | 16,59 (12,06 to 21,89) |
| High systolic blood pressure | Tuvalu | 16,43 (11,40 to 22,44) |
| High systolic blood pressure | Bermuda | 16,42 (11,93 to 21,76) |
| High systolic blood pressure | State of Kuwait | 16,39 (11,85 to 21,03) |
| High systolic blood pressure | United Republic of Tanzania | 16,38 (11,53 to 21,30) |
| High systolic blood pressure | Hashemite Kingdom of Jordan | 16,35 (11,72 to 21,27) |
| High systolic blood pressure | People's Democratic Republic of Algeria | 16,31 (11,52 to 21,31) |
| High systolic blood pressure | Portuguese Republic | 16,30 (11,83 to 21,11) |
| High systolic blood pressure | Jamaica | 16,24 (11,61 to 21,27) |
| High systolic blood pressure | Kingdom of Saudi Arabia | 16,22 (12,00 to 21,32) |
| High systolic blood pressure | Republic of Rwanda | 16,15 (10,98 to 21,38) |
| High systolic blood pressure | Dominican Republic | 16,09 (11,54 to 21,42) |
| High systolic blood pressure | Republic of India | 16,08 (12,02 to 20,48) |
| High systolic blood pressure | Republic of Turkey | 16,05 (11,87 to 20,54) |
| High systolic blood pressure | Belize | 16,05 (11,35 to 20,91) |
| High systolic blood pressure | United States Virgin Islands | 16,04 (11,47 to 21,61) |
| High systolic blood pressure | Independent State of Samoa | 15,99 (11,19 to 21,21) |
| High systolic blood pressure | Brunei Darussalam | 15,94 (11,28 to 20,95) |
| High systolic blood pressure | People's Republic of China | 15,94 (11,35 to 20,62) |
| High systolic blood pressure | Republic of Guatemala | 15,76 (11,34 to 20,42) |
| High systolic blood pressure | Kingdom of Bhutan | 15,52 (10,91 to 21,01) |
| High systolic blood pressure | Saint Vincent and the Grenadines | 15,46 (11,25 to 20,57) |
| High systolic blood pressure | New Zealand | 15,41 (11,02 to 20,01) |
| High systolic blood pressure | Swiss Confederation | 15,35 (10,66 to 20,59) |
| High systolic blood pressure | United Kingdom of Great Britain and Northern Ireland | 15,27 (11,16 to 19,39) |
| High systolic blood pressure | Republic of Italy | 15,18 (11,24 to 20,00) |
| High systolic blood pressure | Islamic Republic of Iran | 15,07 (11,13 to 19,22) |
| High systolic blood pressure | Australia | 14,93 (10,87 to 19,53) |
| High systolic blood pressure | Republic of Peru | 14,86 (10,76 to 19,23) |
| High systolic blood pressure | Islamic Republic of Afghanistan | 14,80 (10,28 to 20,45) |
| High systolic blood pressure | Republic of Yemen | 14,64 (10,73 to 19,49) |
| High systolic blood pressure | Republic of the Philippines | 14,49 (10,67 to 19,23) |
| High systolic blood pressure | Hellenic Republic | 14,48 (10,49 to 18,92) |
| High systolic blood pressure | Palestine | 14,47 (10,14 to 19,00) |
| High systolic blood pressure | Republic of Zambia | 14,24 (9,88 to 18,77) |
| High systolic blood pressure | Canada | 14,19 (10,08 to 19,22) |
| High systolic blood pressure | United States of America | 14,19 (10,04 to 18,67) |
| High systolic blood pressure | Democratic People's Republic of Korea | 14,16 (9,76 to 18,76) |
| High systolic blood pressure | Kingdom of Cambodia | 14,14 (9,52 to 19,65) |
| High systolic blood pressure | Greenland | 14,00 (9,31 to 19,21) |
| High systolic blood pressure | Taiwan (Province of China) | 13,98 (9,72 to 18,84) |
| High systolic blood pressure | Republic of the Marshall Islands | 13,96 (9,61 to 18,72) |
| High systolic blood pressure | Republic of Tunisia | 13,84 (9,62 to 18,49) |
| High systolic blood pressure | Republic of Kiribati | 13,82 (9,52 to 18,70) |
| High systolic blood pressure | Republic of Cuba | 13,65 (9,33 to 18,16) |
| High systolic blood pressure | Kingdom of Thailand | 13,63 (9,78 to 17,80) |
| High systolic blood pressure | Republic of Suriname | 13,35 (9,14 to 18,23) |
| High systolic blood pressure | Solomon Islands | 12,80 (8,75 to 17,95) |
| High systolic blood pressure | Federal Democratic Republic of Ethiopia | 12,58 (8,41 to 17,44) |
| High systolic blood pressure | Plurinational State of Bolivia | 12,52 (8,60 to 17,70) |
| High systolic blood pressure | Republic of Ecuador | 12,27 (8,33 to 16,53) |
| High systolic blood pressure | Federated States of Micronesia | 12,16 (8,01 to 16,96) |
| High systolic blood pressure | Republic of Korea | 11,55 (7,86 to 15,68) |
| High systolic blood pressure | Republic of Singapore | 11,01 (7,35 to 15,23) |
| High systolic blood pressure | Federal Democratic Republic of Nepal | 10,46 (6,99 to 14,30) |
| High systolic blood pressure | Independent State of Papua New Guinea | 9,69 (6,11 to 13,73) |

**Lead exposure**

| **Risk factor** | **Location** | **Percent contribution to ASDR (%)** |
| --- | --- | --- |
| Lead exposure | Federal Democratic Republic of Nepal | 2,13 (-0,28 to 5,28) |
| Lead exposure | Republic of Yemen | 1,99 (-0,28 to 4,68) |
| Lead exposure | Islamic Republic of Afghanistan | 1,96 (-0,27 to 4,81) |
| Lead exposure | Republic of Guatemala | 1,92 (-0,28 to 4,85) |
| Lead exposure | Federal Republic of Somalia | 1,76 (-0,25 to 4,28) |
| Lead exposure | Kingdom of Bhutan | 1,75 (-0,24 to 4,33) |
| Lead exposure | Republic of Haiti | 1,71 (-0,24 to 4,33) |
| Lead exposure | Arab Republic of Egypt | 1,69 (-0,23 to 4,29) |
| Lead exposure | Republic of Honduras | 1,69 (-0,24 to 4,04) |
| Lead exposure | Federal Democratic Republic of Ethiopia | 1,67 (-0,23 to 4,24) |
| Lead exposure | People's Republic of Bangladesh | 1,66 (-0,22 to 4,09) |
| Lead exposure | Islamic Republic of Iran | 1,62 (-0,22 to 3,97) |
| Lead exposure | Republic of Chad | 1,62 (-0,22 to 3,95) |
| Lead exposure | Republic of the Niger | 1,54 (-0,24 to 3,84) |
| Lead exposure | Republic of El Salvador | 1,54 (-0,22 to 3,83) |
| Lead exposure | Burkina Faso | 1,52 (-0,21 to 3,81) |
| Lead exposure | Dominican Republic | 1,51 (-0,21 to 3,87) |
| Lead exposure | Republic of India | 1,38 (-0,19 to 3,52) |
| Lead exposure | Republic of Mali | 1,37 (-0,21 to 3,41) |
| Lead exposure | Saint Vincent and the Grenadines | 1,35 (-0,19 to 3,38) |
| Lead exposure | Republic of Guinea | 1,35 (-0,19 to 3,33) |
| Lead exposure | Islamic Republic of Pakistan | 1,34 (-0,19 to 3,35) |
| Lead exposure | Palestine | 1,33 (-0,18 to 3,34) |
| Lead exposure | Republic of Cuba | 1,33 (-0,17 to 3,50) |
| Lead exposure | Republic of Nicaragua | 1,28 (-0,19 to 3,21) |
| Lead exposure | Republic of Malta | 1,27 (-0,18 to 3,28) |
| Lead exposure | Republic of Sudan | 1,26 (-0,19 to 3,15) |
| Lead exposure | Plurinational State of Bolivia | 1,23 (-0,15 to 3,11) |
| Lead exposure | Kingdom of Lesotho | 1,23 (-0,16 to 3,19) |
| Lead exposure | Syrian Arab Republic | 1,23 (-0,16 to 3,06) |
| Lead exposure | Republic of Suriname | 1,19 (-0,15 to 3,07) |
| Lead exposure | Republic of Mozambique | 1,18 (-0,18 to 2,76) |
| Lead exposure | Republic of Burundi | 1,18 (-0,17 to 2,97) |
| Lead exposure | Republic of South Sudan | 1,14 (-0,15 to 2,85) |
| Lead exposure | Republic of Guyana | 1,13 (-0,15 to 2,84) |
| Lead exposure | Kingdom of Cambodia | 1,12 (-0,15 to 2,83) |
| Lead exposure | Republic of Tunisia | 1,12 (-0,15 to 2,94) |
| Lead exposure | Lao People's Democratic Republic | 1,12 (-0,15 to 2,86) |
| Lead exposure | Jamaica | 1,11 (-0,14 to 2,85) |
| Lead exposure | Republic of Rwanda | 1,11 (-0,15 to 2,77) |
| Lead exposure | Republic of Guinea-Bissau | 1,09 (-0,15 to 2,76) |
| Lead exposure | United Mexican States | 1,08 (-0,15 to 2,78) |
| Lead exposure | Democratic Republic of Timor-Leste | 1,07 (-0,15 to 2,64) |
| Lead exposure | Central African Republic | 1,07 (-0,14 to 2,72) |
| Lead exposure | State of Eritrea | 1,06 (-0,14 to 2,73) |
| Lead exposure | Republic of Cameroon | 1,05 (-0,15 to 2,60) |
| Lead exposure | Republic of Uganda | 1,04 (-0,14 to 2,57) |
| Lead exposure | Grenada | 1,04 (-0,14 to 2,73) |
| Lead exposure | People's Democratic Republic of Algeria | 1,03 (-0,14 to 2,71) |
| Lead exposure | Republic of Liberia | 1,03 (-0,15 to 2,73) |
| Lead exposure | Republic of Malawi | 1,02 (-0,15 to 2,56) |
| Lead exposure | Republic of Zambia | 1,02 (-0,13 to 2,58) |
| Lead exposure | Republic of the Gambia | 1,01 (-0,13 to 2,52) |
| Lead exposure | Saint Lucia | 1,00 (-0,14 to 2,59) |
| Lead exposure | Republic of Benin | 0,99 (-0,14 to 2,50) |
| Lead exposure | Kingdom of Eswatini | 0,99 (-0,14 to 2,53) |
| Lead exposure | Portuguese Republic | 0,99 (-0,13 to 2,47) |
| Lead exposure | People's Republic of China | 0,97 (-0,13 to 2,48) |
| Lead exposure | Republic of Singapore | 0,96 (-0,12 to 2,45) |
| Lead exposure | Union of the Comoros | 0,96 (-0,14 to 2,47) |
| Lead exposure | Republic of Botswana | 0,96 (-0,14 to 2,43) |
| Lead exposure | Republic of Djibouti | 0,96 (-0,13 to 2,37) |
| Lead exposure | Bolivarian Republic of Venezuela | 0,95 (-0,14 to 2,44) |
| Lead exposure | Hashemite Kingdom of Jordan | 0,94 (-0,13 to 2,50) |
| Lead exposure | Republic of Côte d'Ivoire | 0,93 (-0,13 to 2,31) |
| Lead exposure | Republic of the Union of Myanmar | 0,92 (-0,12 to 2,37) |
| Lead exposure | Democratic Republic of the Congo | 0,92 (-0,13 to 2,33) |
| Lead exposure | Republic of Madagascar | 0,92 (-0,12 to 2,35) |
| Lead exposure | Republic of Peru | 0,92 (-0,12 to 2,29) |
| Lead exposure | Republic of Zimbabwe | 0,91 (-0,13 to 2,29) |
| Lead exposure | Democratic People's Republic of Korea | 0,90 (-0,11 to 2,32) |
| Lead exposure | Republic of Panama | 0,90 (-0,12 to 2,31) |
| Lead exposure | Kingdom of Saudi Arabia | 0,90 (-0,13 to 2,24) |
| Lead exposure | Solomon Islands | 0,87 (-0,11 to 2,31) |
| Lead exposure | Republic of Namibia | 0,86 (-0,11 to 2,27) |
| Lead exposure | Kingdom of Belgium | 0,85 (-0,12 to 2,22) |
| Lead exposure | Sultanate of Oman | 0,84 (-0,11 to 2,15) |
| Lead exposure | Republic of Tajikistan | 0,84 (-0,12 to 2,18) |
| Lead exposure | State of Libya | 0,84 (-0,12 to 2,12) |
| Lead exposure | Taiwan (Province of China) | 0,83 (-0,11 to 2,15) |
| Lead exposure | Republic of Ecuador | 0,83 (-0,10 to 2,14) |
| Lead exposure | Republic of Costa Rica | 0,83 (-0,12 to 2,04) |
| Lead exposure | Republic of Angola | 0,81 (-0,12 to 2,14) |
| Lead exposure | Australia | 0,80 (-0,10 to 2,09) |
| Lead exposure | Togolese Republic | 0,80 (-0,11 to 2,00) |
| Lead exposure | Hellenic Republic | 0,79 (-0,10 to 2,05) |
| Lead exposure | Republic of Iraq | 0,79 (-0,11 to 1,95) |
| Lead exposure | Republic of Korea | 0,79 (-0,10 to 2,06) |
| Lead exposure | Federative Republic of Brazil | 0,78 (-0,10 to 2,01) |
| Lead exposure | Kingdom of Morocco | 0,78 (-0,11 to 2,02) |
| Lead exposure | Republic of Kenya | 0,77 (-0,11 to 1,99) |
| Lead exposure | Eastern Republic of Uruguay | 0,77 (-0,10 to 2,00) |
| Lead exposure | Kingdom of Spain | 0,77 (-0,10 to 1,95) |
| Lead exposure | Democratic Republic of Sao Tome and Principe | 0,77 (-0,10 to 2,06) |
| Lead exposure | Republic of Maldives | 0,76 (-0,11 to 1,99) |
| Lead exposure | Republic of Sierra Leone | 0,76 (-0,11 to 1,83) |
| Lead exposure | Republic of Colombia | 0,75 (-0,11 to 1,93) |
| Lead exposure | Antigua and Barbuda | 0,75 (-0,09 to 1,96) |
| Lead exposure | Republic of Paraguay | 0,75 (-0,10 to 1,93) |
| Lead exposure | Belize | 0,73 (-0,10 to 1,89) |
| Lead exposure | Bosnia and Herzegovina | 0,73 (-0,10 to 1,79) |
| Lead exposure | Saint Kitts and Nevis | 0,72 (-0,10 to 1,91) |
| Lead exposure | Republic of Senegal | 0,72 (-0,10 to 1,82) |
| Lead exposure | Republic of Cabo Verde | 0,72 (-0,10 to 1,83) |
| Lead exposure | Islamic Republic of Mauritania | 0,72 (-0,10 to 1,73) |
| Lead exposure | Republic of Cyprus | 0,72 (-0,09 to 1,80) |
| Lead exposure | New Zealand | 0,72 (-0,09 to 1,84) |
| Lead exposure | State of Kuwait | 0,72 (-0,10 to 1,95) |
| Lead exposure | Commonwealth of Dominica | 0,71 (-0,10 to 1,84) |
| Lead exposure | Kingdom of Bahrain | 0,71 (-0,09 to 1,88) |
| Lead exposure | Kyrgyz Republic | 0,67 (-0,09 to 1,77) |
| Lead exposure | Republic of Turkey | 0,67 (-0,09 to 1,77) |
| Lead exposure | Lebanese Republic | 0,67 (-0,09 to 1,72) |
| Lead exposure | Georgia | 0,65 (-0,09 to 1,63) |
| Lead exposure | Republic of Ghana | 0,64 (-0,09 to 1,68) |
| Lead exposure | Republic of Equatorial Guinea | 0,64 (-0,10 to 1,62) |
| Lead exposure | Socialist Republic of Viet Nam | 0,63 (-0,09 to 1,58) |
| Lead exposure | Mongolia | 0,62 (-0,09 to 1,59) |
| Lead exposure | Brunei Darussalam | 0,61 (-0,08 to 1,69) |
| Lead exposure | Republic of Italy | 0,61 (-0,08 to 1,59) |
| Lead exposure | Swiss Confederation | 0,61 (-0,08 to 1,57) |
| Lead exposure | Republic of the Philippines | 0,60 (-0,08 to 1,54) |
| Lead exposure | Tuvalu | 0,59 (-0,08 to 1,37) |
| Lead exposure | Republic of Albania | 0,59 (-0,08 to 1,55) |
| Lead exposure | Barbados | 0,59 (-0,08 to 1,55) |
| Lead exposure | Republic of Poland | 0,59 (-0,08 to 1,52) |
| Lead exposure | Argentine Republic | 0,59 (-0,08 to 1,52) |
| Lead exposure | Gabonese Republic | 0,58 (-0,07 to 1,57) |
| Lead exposure | Republic of South Africa | 0,58 (-0,08 to 1,54) |
| Lead exposure | Republic of the Congo | 0,58 (-0,08 to 1,53) |
| Lead exposure | United Arab Emirates | 0,56 (-0,07 to 1,45) |
| Lead exposure | Malaysia | 0,55 (-0,07 to 1,41) |
| Lead exposure | Independent State of Papua New Guinea | 0,55 (-0,07 to 1,42) |
| Lead exposure | Republic of Armenia | 0,55 (-0,07 to 1,45) |
| Lead exposure | Federal Republic of Nigeria | 0,55 (-0,08 to 1,39) |
| Lead exposure | United Republic of Tanzania | 0,54 (-0,08 to 1,45) |
| Lead exposure | French Republic | 0,54 (-0,08 to 1,41) |
| Lead exposure | Republic of Indonesia | 0,54 (-0,08 to 1,32) |
| Lead exposure | Republic of Uzbekistan | 0,53 (-0,07 to 1,34) |
| Lead exposure | Republic of Azerbaijan | 0,53 (-0,07 to 1,39) |
| Lead exposure | Kingdom of Thailand | 0,52 (-0,07 to 1,38) |
| Lead exposure | State of Qatar | 0,52 (-0,07 to 1,32) |
| Lead exposure | Republic of Mauritius | 0,50 (-0,06 to 1,31) |
| Lead exposure | Puerto Rico | 0,50 (-0,07 to 1,27) |
| Lead exposure | Ireland | 0,49 (-0,07 to 1,26) |
| Lead exposure | Commonwealth of the Bahamas | 0,48 (-0,06 to 1,26) |
| Lead exposure | North Macedonia | 0,48 (-0,06 to 1,27) |
| Lead exposure | Republic of Vanuatu | 0,48 (-0,06 to 1,23) |
| Lead exposure | United States Virgin Islands | 0,48 (-0,06 to 1,26) |
| Lead exposure | Republic of Iceland | 0,47 (-0,06 to 1,23) |
| Lead exposure | Republic of Kiribati | 0,46 (-0,06 to 1,22) |
| Lead exposure | Republic of Bulgaria | 0,46 (-0,06 to 1,20) |
| Lead exposure | Bermuda | 0,44 (-0,06 to 1,15) |
| Lead exposure | Republic of the Marshall Islands | 0,44 (-0,06 to 1,10) |
| Lead exposure | Kingdom of the Netherlands | 0,43 (-0,06 to 1,11) |
| Lead exposure | United States of America | 0,43 (-0,05 to 1,13) |
| Lead exposure | United Kingdom of Great Britain and Northern Ireland | 0,43 (-0,06 to 1,10) |
| Lead exposure | Democratic Socialist Republic of Sri Lanka | 0,43 (-0,06 to 1,12) |
| Lead exposure | Turkmenistan | 0,43 (-0,06 to 1,10) |
| Lead exposure | Republic of Croatia | 0,43 (-0,06 to 1,13) |
| Lead exposure | Grand Duchy of Luxembourg | 0,41 (-0,05 to 1,05) |
| Lead exposure | Republic of Serbia | 0,41 (-0,05 to 1,08) |
| Lead exposure | Canada | 0,41 (-0,05 to 1,12) |
| Lead exposure | Kingdom of Norway | 0,40 (-0,06 to 1,06) |
| Lead exposure | Republic of Moldova | 0,40 (-0,05 to 1,03) |
| Lead exposure | Romania | 0,40 (-0,06 to 1,03) |
| Lead exposure | State of Israel | 0,39 (-0,05 to 1,09) |
| Lead exposure | Kingdom of Denmark | 0,39 (-0,05 to 1,02) |
| Lead exposure | Republic of Austria | 0,39 (-0,05 to 0,97) |
| Lead exposure | Principality of Monaco | 0,39 (-0,05 to 0,98) |
| Lead exposure | Republic of Trinidad and Tobago | 0,38 (-0,05 to 1,03) |
| Lead exposure | Greenland | 0,38 (-0,05 to 0,99) |
| Lead exposure | Federated States of Micronesia | 0,38 (-0,05 to 0,99) |
| Lead exposure | Principality of Andorra | 0,38 (-0,05 to 0,98) |
| Lead exposure | Republic of Seychelles | 0,37 (-0,05 to 0,98) |
| Lead exposure | Republic of Belarus | 0,37 (-0,05 to 1,01) |
| Lead exposure | Republic of San Marino | 0,37 (-0,05 to 0,98) |
| Lead exposure | Czech Republic | 0,37 (-0,05 to 0,94) |
| Lead exposure | Republic of Nauru | 0,37 (-0,05 to 0,82) |
| Lead exposure | Kingdom of Tonga | 0,37 (-0,05 to 0,95) |
| Lead exposure | Slovak Republic | 0,36 (-0,05 to 0,92) |
| Lead exposure | Montenegro | 0,35 (-0,05 to 0,92) |
| Lead exposure | Hungary | 0,35 (-0,05 to 0,91) |
| Lead exposure | Japan | 0,34 (-0,05 to 0,91) |
| Lead exposure | Independent State of Samoa | 0,33 (-0,04 to 0,86) |
| Lead exposure | Russian Federation | 0,33 (-0,04 to 0,85) |
| Lead exposure | Republic of Slovenia | 0,32 (-0,04 to 0,86) |
| Lead exposure | Republic of Estonia | 0,31 (-0,04 to 0,83) |
| Lead exposure | Federal Republic of Germany | 0,31 (-0,04 to 0,80) |
| Lead exposure | Republic of Latvia | 0,31 (-0,04 to 0,80) |
| Lead exposure | Republic of Fiji | 0,30 (-0,04 to 0,76) |
| Lead exposure | Kingdom of Sweden | 0,29 (-0,04 to 0,76) |
| Lead exposure | Republic of Kazakhstan | 0,29 (-0,04 to 0,78) |
| Lead exposure | Tokelau | 0,29 (-0,04 to 0,76) |
| Lead exposure | Republic of Lithuania | 0,28 (-0,04 to 0,74) |
| Lead exposure | Ukraine | 0,28 (-0,04 to 0,71) |
| Lead exposure | Republic of Chile | 0,26 (-0,04 to 0,69) |
| Lead exposure | Cook Islands | 0,22 (-0,03 to 0,58) |
| Lead exposure | American Samoa | 0,21 (-0,03 to 0,53) |
| Lead exposure | Republic of Finland | 0,21 (-0,03 to 0,54) |
| Lead exposure | Republic of Palau | 0,20 (-0,02 to 0,54) |
| Lead exposure | Republic of Niue | 0,20 (-0,03 to 0,51) |
| Lead exposure | Guam | 0,20 (-0,02 to 0,52) |
| Lead exposure | Northern Mariana Islands | 0,19 (-0,02 to 0,50) |

**Smoking**

| **Risk factor** | **Location** | **Percent contribution to ASDR (%)** |
| --- | --- | --- |
| Smoking | Lebanese Republic | 48,47 (41,65 to 55,20) |
| Smoking | Georgia | 48,09 (42,09 to 53,79) |
| Smoking | Republic of Belarus | 47,75 (42,33 to 52,65) |
| Smoking | Hellenic Republic | 46,83 (40,43 to 53,29) |
| Smoking | Republic of Albania | 46,29 (38,73 to 53,73) |
| Smoking | Hashemite Kingdom of Jordan | 46,27 (39,40 to 52,30) |
| Smoking | Bosnia and Herzegovina | 45,50 (38,71 to 52,02) |
| Smoking | People's Republic of China | 44,74 (37,99 to 50,98) |
| Smoking | Republic of Kiribati | 44,63 (37,98 to 51,02) |
| Smoking | Republic of Iraq | 43,90 (37,34 to 50,41) |
| Smoking | Montenegro | 42,75 (35,74 to 49,64) |
| Smoking | Republic of Croatia | 42,66 (36,52 to 49,12) |
| Smoking | Arab Republic of Egypt | 42,37 (36,34 to 48,24) |
| Smoking | Republic of Bulgaria | 42,17 (36,80 to 47,06) |
| Smoking | Ukraine | 41,94 (34,38 to 48,99) |
| Smoking | Kyrgyz Republic | 41,82 (36,70 to 46,54) |
| Smoking | Republic of Tunisia | 41,25 (32,55 to 48,73) |
| Smoking | Kingdom of Denmark | 41,02 (33,96 to 48,51) |
| Smoking | Palestine | 40,64 (34,26 to 47,48) |
| Smoking | Republic of Paraguay | 40,31 (33,01 to 47,72) |
| Smoking | North Macedonia | 40,03 (32,74 to 47,22) |
| Smoking | Republic of Turkey | 39,92 (34,19 to 45,75) |
| Smoking | Greenland | 39,81 (33,19 to 46,92) |
| Smoking | Republic of Moldova | 39,62 (34,18 to 44,99) |
| Smoking | State of Kuwait | 39,50 (32,58 to 46,53) |
| Smoking | Republic of Serbia | 39,41 (33,67 to 45,93) |
| Smoking | Republic of Armenia | 39,26 (34,56 to 43,90) |
| Smoking | Czech Republic | 39,01 (33,12 to 45,36) |
| Smoking | Republic of Latvia | 38,67 (33,36 to 43,94) |
| Smoking | Republic of Azerbaijan | 38,66 (28,06 to 46,99) |
| Smoking | Republic of Yemen | 38,48 (31,21 to 46,37) |
| Smoking | Republic of Lithuania | 38,32 (32,83 to 43,73) |
| Smoking | Slovak Republic | 38,32 (32,29 to 44,72) |
| Smoking | Russian Federation | 38,18 (33,76 to 42,97) |
| Smoking | Kingdom of Spain | 38,06 (31,71 to 44,61) |
| Smoking | Republic of Cyprus | 37,98 (31,08 to 45,19) |
| Smoking | Syrian Arab Republic | 37,80 (31,58 to 43,77) |
| Smoking | Kingdom of Cambodia | 37,35 (28,74 to 46,20) |
| Smoking | Republic of San Marino | 37,17 (29,23 to 45,05) |
| Smoking | Taiwan (Province of China) | 36,92 (31,81 to 41,78) |
| Smoking | Federal Republic of Germany | 36,87 (31,13 to 43,36) |
| Smoking | Republic of Estonia | 36,84 (31,19 to 42,81) |
| Smoking | United States of America | 36,77 (30,52 to 43,28) |
| Smoking | Hungary | 36,74 (31,45 to 41,82) |
| Smoking | Romania | 36,29 (31,43 to 41,25) |
| Smoking | Republic of Poland | 36,21 (30,92 to 42,01) |
| Smoking | Democratic People's Republic of Korea | 36,12 (29,98 to 42,69) |
| Smoking | Kingdom of Tonga | 35,86 (29,18 to 43,27) |
| Smoking | Eastern Republic of Uruguay | 35,85 (30,09 to 41,66) |
| Smoking | Kingdom of Belgium | 35,59 (29,08 to 41,71) |
| Smoking | Republic of Maldives | 35,30 (25,44 to 44,97) |
| Smoking | Guam | 35,22 (29,63 to 41,96) |
| Smoking | People's Democratic Republic of Algeria | 35,18 (27,98 to 41,89) |
| Smoking | Republic of Austria | 35,03 (29,22 to 40,78) |
| Smoking | Kingdom of Sweden | 34,55 (27,82 to 42,26) |
| Smoking | Grand Duchy of Luxembourg | 34,54 (27,97 to 41,70) |
| Smoking | Federative Republic of Brazil | 34,53 (28,61 to 40,69) |
| Smoking | Dominican Republic | 34,45 (28,17 to 41,58) |
| Smoking | Republic of Italy | 34,38 (28,59 to 40,57) |
| Smoking | Republic of Mauritius | 34,37 (29,06 to 39,99) |
| Smoking | Kingdom of the Netherlands | 34,31 (28,10 to 41,57) |
| Smoking | Federal Democratic Republic of Nepal | 34,29 (27,45 to 41,24) |
| Smoking | State of Libya | 33,70 (26,15 to 40,76) |
| Smoking | Lao People's Democratic Republic | 33,69 (26,25 to 40,91) |
| Smoking | French Republic | 33,65 (27,88 to 39,89) |
| Smoking | Principality of Andorra | 33,48 (26,62 to 41,13) |
| Smoking | Swiss Confederation | 33,48 (27,53 to 40,17) |
| Smoking | Argentine Republic | 33,23 (28,02 to 39,05) |
| Smoking | Republic of Malta | 33,05 (27,18 to 39,23) |
| Smoking | Canada | 32,96 (26,67 to 39,38) |
| Smoking | Republic of Slovenia | 32,82 (27,28 to 39,23) |
| Smoking | Republic of Seychelles | 32,78 (24,40 to 41,37) |
| Smoking | People's Republic of Bangladesh | 32,78 (24,90 to 42,37) |
| Smoking | Mongolia | 32,63 (25,52 to 39,50) |
| Smoking | Socialist Republic of Viet Nam | 32,63 (26,72 to 39,42) |
| Smoking | Republic of Rwanda | 32,55 (25,02 to 40,41) |
| Smoking | Kingdom of Bahrain | 32,41 (26,13 to 39,02) |
| Smoking | Republic of Cuba | 32,30 (26,38 to 38,93) |
| Smoking | Republic of Kazakhstan | 31,92 (27,48 to 35,96) |
| Smoking | United Kingdom of Great Britain and Northern Ireland | 31,88 (25,41 to 39,08) |
| Smoking | Republic of Tajikistan | 31,85 (25,71 to 38,30) |
| Smoking | Republic of the Philippines | 31,34 (25,56 to 37,05) |
| Smoking | Principality of Monaco | 31,34 (25,10 to 38,70) |
| Smoking | Ireland | 31,09 (25,01 to 37,61) |
| Smoking | Republic of Iceland | 30,88 (24,63 to 37,85) |
| Smoking | New Zealand | 30,61 (24,55 to 37,20) |
| Smoking | Turkmenistan | 30,59 (25,54 to 35,93) |
| Smoking | Republic of Suriname | 30,37 (24,84 to 36,33) |
| Smoking | Islamic Republic of Iran | 30,30 (24,84 to 35,66) |
| Smoking | Republic of Costa Rica | 30,06 (24,73 to 35,99) |
| Smoking | Independent State of Samoa | 29,97 (23,49 to 36,78) |
| Smoking | Republic of Korea | 29,91 (23,92 to 36,27) |
| Smoking | Republic of Sudan | 29,75 (23,72 to 37,20) |
| Smoking | Republic of Indonesia | 29,73 (21,49 to 37,36) |
| Smoking | Japan | 29,69 (24,82 to 34,90) |
| Smoking | Portuguese Republic | 29,56 (24,70 to 34,39) |
| Smoking | State of Israel | 29,38 (23,84 to 35,21) |
| Smoking | Malaysia | 29,15 (23,49 to 34,88) |
| Smoking | Republic of the Union of Myanmar | 29,00 (23,17 to 35,46) |
| Smoking | Republic of Finland | 28,90 (23,21 to 34,99) |
| Smoking | Kingdom of Thailand | 28,58 (23,40 to 33,90) |
| Smoking | Kingdom of Lesotho | 28,33 (20,47 to 39,85) |
| Smoking | Democratic Republic of Timor-Leste | 28,32 (20,77 to 36,25) |
| Smoking | Brunei Darussalam | 28,26 (22,78 to 35,06) |
| Smoking | Solomon Islands | 27,86 (21,73 to 36,64) |
| Smoking | Republic of Zimbabwe | 27,75 (21,01 to 34,39) |
| Smoking | Federated States of Micronesia | 27,65 (21,96 to 34,11) |
| Smoking | Republic of Malawi | 27,36 (17,96 to 35,84) |
| Smoking | Republic of Botswana | 27,21 (21,71 to 32,68) |
| Smoking | Puerto Rico | 26,87 (21,03 to 33,14) |
| Smoking | Bermuda | 26,71 (20,87 to 33,37) |
| Smoking | Republic of Nicaragua | 26,63 (21,27 to 32,52) |
| Smoking | State of Qatar | 26,58 (20,65 to 34,08) |
| Smoking | Tuvalu | 26,51 (20,88 to 32,98) |
| Smoking | Cook Islands | 26,27 (19,20 to 34,32) |
| Smoking | American Samoa | 26,26 (20,94 to 31,31) |
| Smoking | Islamic Republic of Afghanistan | 26,19 (15,11 to 34,36) |
| Smoking | Sultanate of Oman | 25,99 (19,19 to 33,49) |
| Smoking | United Republic of Tanzania | 25,95 (19,63 to 32,97) |
| Smoking | Republic of Honduras | 25,92 (20,22 to 32,37) |
| Smoking | Republic of Djibouti | 25,90 (19,26 to 32,94) |
| Smoking | Republic of Chile | 25,86 (21,56 to 30,19) |
| Smoking | Islamic Republic of Pakistan | 25,56 (19,30 to 32,94) |
| Smoking | Republic of Namibia | 25,41 (20,14 to 31,11) |
| Smoking | Republic of Nauru | 25,16 (16,99 to 36,14) |
| Smoking | Jamaica | 24,85 (19,90 to 30,62) |
| Smoking | Republic of Trinidad and Tobago | 24,78 (19,81 to 30,10) |
| Smoking | Republic of Fiji | 24,52 (20,28 to 29,61) |
| Smoking | Republic of Palau | 24,25 (19,33 to 30,32) |
| Smoking | Republic of Niue | 23,98 (18,66 to 29,86) |
| Smoking | Republic of India | 23,96 (18,35 to 29,99) |
| Smoking | Republic of the Marshall Islands | 23,86 (18,72 to 31,09) |
| Smoking | Kingdom of Norway | 23,47 (18,58 to 29,50) |
| Smoking | Northern Mariana Islands | 23,38 (18,54 to 28,54) |
| Smoking | Kingdom of Morocco | 23,36 (18,05 to 28,68) |
| Smoking | United Arab Emirates | 23,30 (17,79 to 29,15) |
| Smoking | Republic of El Salvador | 23,18 (18,48 to 28,21) |
| Smoking | Independent State of Papua New Guinea | 23,08 (17,55 to 29,28) |
| Smoking | Bolivarian Republic of Venezuela | 23,05 (18,53 to 27,74) |
| Smoking | Tokelau | 22,86 (17,34 to 29,46) |
| Smoking | Republic of Peru | 22,62 (17,75 to 28,12) |
| Smoking | Republic of Uzbekistan | 22,55 (18,82 to 26,35) |
| Smoking | Republic of Panama | 22,47 (17,99 to 27,66) |
| Smoking | Antigua and Barbuda | 22,47 (17,40 to 28,94) |
| Smoking | Australia | 22,43 (17,56 to 28,05) |
| Smoking | United Mexican States | 21,85 (17,71 to 26,01) |
| Smoking | Commonwealth of the Bahamas | 21,73 (16,86 to 27,62) |
| Smoking | Republic of Guyana | 21,66 (17,00 to 26,65) |
| Smoking | Democratic Socialist Republic of Sri Lanka | 21,38 (17,09 to 26,37) |
| Smoking | United States Virgin Islands | 21,15 (15,41 to 27,57) |
| Smoking | Saint Vincent and the Grenadines | 21,14 (16,63 to 26,18) |
| Smoking | Kingdom of Saudi Arabia | 21,14 (15,14 to 28,32) |
| Smoking | Belize | 21,09 (16,62 to 26,11) |
| Smoking | Republic of South Africa | 20,87 (17,20 to 24,94) |
| Smoking | Republic of Guatemala | 20,83 (16,33 to 25,57) |
| Smoking | Republic of Angola | 20,79 (15,80 to 25,93) |
| Smoking | Republic of Colombia | 20,33 (16,40 to 24,83) |
| Smoking | Plurinational State of Bolivia | 20,17 (15,73 to 25,25) |
| Smoking | Kingdom of Bhutan | 20,13 (13,05 to 27,47) |
| Smoking | Republic of Vanuatu | 19,93 (14,03 to 26,17) |
| Smoking | Republic of the Congo | 19,76 (14,49 to 25,88) |
| Smoking | Republic of Ecuador | 19,75 (15,79 to 23,77) |
| Smoking | Togolese Republic | 19,62 (13,10 to 26,22) |
| Smoking | Republic of Singapore | 19,39 (15,48 to 23,59) |
| Smoking | Grenada | 18,94 (14,82 to 23,40) |
| Smoking | Republic of Zambia | 18,88 (14,21 to 23,60) |
| Smoking | Barbados | 18,74 (14,28 to 23,85) |
| Smoking | Union of the Comoros | 18,68 (12,05 to 25,49) |
| Smoking | Republic of Mozambique | 18,42 (10,65 to 24,98) |
| Smoking | Commonwealth of Dominica | 17,55 (13,45 to 22,49) |
| Smoking | Saint Kitts and Nevis | 17,37 (12,86 to 22,34) |
| Smoking | Republic of South Sudan | 17,20 (8,87 to 25,16) |
| Smoking | Saint Lucia | 16,92 (13,14 to 21,80) |
| Smoking | Republic of Equatorial Guinea | 16,81 (12,21 to 22,00) |
| Smoking | Republic of Côte d'Ivoire | 16,58 (11,69 to 21,91) |
| Smoking | Republic of the Gambia | 16,42 (10,52 to 22,52) |
| Smoking | Kingdom of Eswatini | 16,17 (12,29 to 20,87) |
| Smoking | Gabonese Republic | 15,91 (11,98 to 20,29) |
| Smoking | Republic of Ghana | 15,63 (11,43 to 20,59) |
| Smoking | Republic of Kenya | 15,44 (12,10 to 19,20) |
| Smoking | Republic of Guinea | 15,30 (8,95 to 21,74) |
| Smoking | Republic of Mali | 14,78 (6,03 to 23,64) |
| Smoking | Republic of Uganda | 14,50 (10,53 to 19,09) |
| Smoking | Republic of Sierra Leone | 14,38 (8,82 to 19,60) |
| Smoking | Republic of Haiti | 14,30 (10,12 to 19,43) |
| Smoking | Central African Republic | 14,23 (7,73 to 21,34) |
| Smoking | Federal Republic of Somalia | 13,56 (4,55 to 25,27) |
| Smoking | Republic of Chad | 13,36 (6,94 to 20,27) |
| Smoking | Republic of Senegal | 12,94 (8,12 to 18,06) |
| Smoking | Republic of Cameroon | 12,57 (8,92 to 16,37) |
| Smoking | Republic of Madagascar | 12,50 (8,46 to 16,79) |
| Smoking | Republic of Burundi | 12,24 (6,73 to 17,27) |
| Smoking | Republic of Cabo Verde | 12,05 (8,96 to 15,72) |
| Smoking | Republic of Guinea-Bissau | 11,29 (6,13 to 16,54) |
| Smoking | Republic of Liberia | 11,20 (6,02 to 15,98) |
| Smoking | Islamic Republic of Mauritania | 11,06 (7,40 to 15,57) |
| Smoking | Democratic Republic of Sao Tome and Principe | 10,83 (7,33 to 14,82) |
| Smoking | Republic of Benin | 10,41 (6,36 to 14,55) |
| Smoking | Democratic Republic of the Congo | 10,13 (6,06 to 14,33) |
| Smoking | State of Eritrea | 9,87 (6,00 to 15,05) |
| Smoking | Burkina Faso | 9,81 (4,67 to 15,15) |
| Smoking | Federal Republic of Nigeria | 8,70 (6,03 to 11,84) |
| Smoking | Federal Democratic Republic of Ethiopia | 8,20 (4,95 to 11,10) |
| Smoking | Republic of the Niger | 6,93 (2,19 to 12,38) |

### Data for Figure 7

#### Fig 7A. Percentage contribution of risk factors to AA- ASDR in Europe in 2021

| **Risk factor** | **Location** | **Percent contribution to ASDR (%)** |
| --- | --- | --- |
| Diet high in sodium | Central Europe | 1.53 (0.26 to 3.66) |
| Diet low in fruits | Central Europe | 3.86 (2.69 to 5.10) |
| Diet low in vegetables | Central Europe | 1.85 (1.13 to 2.81) |
| High body-mass index | Central Europe | 11.01 (5.89 to 18.69) |
| High systolic blood pressure | Central Europe | 20.29 (15.31 to 25.40) |
| Lead exposure | Central Europe | 0.48 (-0.07 to 1.24) |
| Smoking | Central Europe | 38.06 (32.84 to 43.23) |
| Diet high in sodium | Eastern Europe | 0.70 (0.03 to 2.27) |
| Diet low in fruits | Eastern Europe | 4.65 (3.25 to 6.15) |
| Diet low in vegetables | Eastern Europe | 2.89 (1.87 to 4.15) |
| High body-mass index | Eastern Europe | 11.37 (6.03 to 19.53) |
| High systolic blood pressure | Eastern Europe | 20.66 (15.72 to 25.95) |
| Lead exposure | Eastern Europe | 0.32 (-0.04 to 0.84) |
| Smoking | Eastern Europe | 39.29 (34.77 to 43.90) |
| Diet high in sodium | Western Europe | 0.53 (0.01 to 2.12) |
| Diet low in fruits | Western Europe | 3.06 (2.15 to 4.02) |
| Diet low in vegetables | Western Europe | 2.98 (1.96 to 4.25) |
| High body-mass index | Western Europe | 8.76 (4.63 to 15.32) |
| High systolic blood pressure | Western Europe | 17.40 (13.12 to 22.15) |
| Lead exposure | Western Europe | 0.50 (-0.07 to 1.28) |
| Smoking | Western Europe | 34.82 (29.10 to 41.11) |

#### Fig 7B. Percentage contribution of risk factors to AA-ASDR in Europe grouped by sex in 2021

| **Risk factor** | **Sex** | **Location** | **Percent contribution to ASDR (%)** |
| --- | --- | --- | --- |
| Diet high in sodium | Male | Central Europe | 1.76 (0.33 to 4.12) |
| Diet high in sodium | Female | Central Europe | 1.07 (0.10 to 2.93) |
| Diet low in fruits | Male | Central Europe | 3.74 (2.59 to 4.96) |
| Diet low in fruits | Female | Central Europe | 3.99 (2.77 to 5.31) |
| Diet low in vegetables | Male | Central Europe | 1.86 (1.13 to 2.81) |
| Diet low in vegetables | Female | Central Europe | 1.84 (1.10 to 2.88) |
| High body-mass index | Male | Central Europe | 10.40 (5.51 to 18.00) |
| High body-mass index | Female | Central Europe | 11.78 (6.32 to 19.60) |
| High systolic blood pressure | Male | Central Europe | 19.83 (15.00 to 24.79) |
| High systolic blood pressure | Female | Central Europe | 20.59 (15.93 to 26.04) |
| Lead exposure | Male | Central Europe | 0.55 (-0.07 to 1.42) |
| Lead exposure | Female | Central Europe | 0.36 (-0.05 to 0.93) |
| Smoking | Male | Central Europe | 44.04 (37.71 to 50.06) |
| Smoking | Female | Central Europe | 22.99 (18.93 to 27.04) |
| Diet high in sodium | Male | Eastern Europe | 0.86 (0.04 to 2.64) |
| Diet high in sodium | Female | Eastern Europe | 0.38 (0.00 to 1.58) |
| Diet low in fruits | Male | Eastern Europe | 4.53 (3.20 to 5.98) |
| Diet low in fruits | Female | Eastern Europe | 4.71 (3.29 to 6.24) |
| Diet low in vegetables | Male | Eastern Europe | 2.88 (1.84 to 4.20) |
| Diet low in vegetables | Female | Eastern Europe | 2.86 (1.82 to 4.18) |
| High body-mass index | Male | Eastern Europe | 10.15 (5.38 to 17.77) |
| High body-mass index | Female | Eastern Europe | 13.22 (7.00 to 22.24) |
| High systolic blood pressure | Male | Eastern Europe | 20.16 (15.35 to 25.50) |
| High systolic blood pressure | Female | Eastern Europe | 21.11 (15.92 to 26.32) |
| Lead exposure | Male | Eastern Europe | 0.35 (-0.05 to 0.92) |
| Lead exposure | Female | Eastern Europe | 0.28 (-0.04 to 0.71) |
| Smoking | Male | Eastern Europe | 53.67 (47.13 to 60.35) |
| Smoking | Female | Eastern Europe | 11.07 (9.14 to 13.40) |
| Diet high in sodium | Male | Western Europe | 0.64 (0.01 to 2.39) |
| Diet high in sodium | Female | Western Europe | 0.31 (0.00 to 1.57) |
| Diet low in fruits | Male | Western Europe | 2.93 (2.07 to 3.86) |
| Diet low in fruits | Female | Western Europe | 3.26 (2.24 to 4.35) |
| Diet low in vegetables | Male | Western Europe | 2.93 (1.93 to 4.17) |
| Diet low in vegetables | Female | Western Europe | 3.05 (1.99 to 4.35) |
| High body-mass index | Male | Western Europe | 8.44 (4.46 to 14.76) |
| High body-mass index | Female | Western Europe | 9.15 (4.75 to 15.83) |
| High systolic blood pressure | Male | Western Europe | 16.87 (12.58 to 21.71) |
| High systolic blood pressure | Female | Western Europe | 18.07 (13.43 to 22.74) |
| Lead exposure | Male | Western Europe | 0.56 (-0.07 to 1.43) |
| Lead exposure | Female | Western Europe | 0.40 (-0.05 to 1.03) |
| Smoking | Male | Western Europe | 39.63 (32.93 to 46.83) |
| Smoking | Female | Western Europe | 23.07 (18.65 to 27.84) |

#### Fig 7C. AA-ASDR attributable to risk factors in Europe grouped by sex from 1990 to 2021

| **Year** | **Risk factor** | **Sex** | **Location** | **ASDR per 100,000** |
| --- | --- | --- | --- | --- |
| 1990 | Diet high in sodium | Male | Central Europe | 0.09 (0.02 to 0.21) |
| 1990 | Diet high in sodium | Female | Central Europe | 0.02 (0.00 to 0.05) |
| 1990 | Diet high in sodium | Both | Central Europe | 0.05 (0.01 to 0.11) |
| 2021 | Diet high in sodium | Male | Central Europe | 0.08 (0.02 to 0.20) |
| 2021 | Diet high in sodium | Female | Central Europe | 0.02 (0.00 to 0.05) |
| 2021 | Diet high in sodium | Both | Central Europe | 0.04 (0.01 to 0.11) |
| 1990 | Diet low in fruits | Male | Central Europe | 0.24 (0.17 to 0.32) |
| 1990 | Diet low in fruits | Female | Central Europe | 0.08 (0.06 to 0.11) |
| 1990 | Diet low in fruits | Both | Central Europe | 0.15 (0.10 to 0.20) |
| 2021 | Diet low in fruits | Male | Central Europe | 0.18 (0.12 to 0.24) |
| 2021 | Diet low in fruits | Female | Central Europe | 0.07 (0.04 to 0.09) |
| 2021 | Diet low in fruits | Both | Central Europe | 0.11 (0.08 to 0.15) |
| 1990 | Diet low in vegetables | Male | Central Europe | 0.15 (0.10 to 0.22) |
| 1990 | Diet low in vegetables | Female | Central Europe | 0.05 (0.03 to 0.07) |
| 1990 | Diet low in vegetables | Both | Central Europe | 0.09 (0.06 to 0.13) |
| 2021 | Diet low in vegetables | Male | Central Europe | 0.09 (0.05 to 0.14) |
| 2021 | Diet low in vegetables | Female | Central Europe | 0.03 (0.02 to 0.05) |
| 2021 | Diet low in vegetables | Both | Central Europe | 0.05 (0.03 to 0.08) |
| 1990 | High body-mass index | Male | Central Europe | 0.47 (0.24 to 0.81) |
| 1990 | High body-mass index | Female | Central Europe | 0.17 (0.09 to 0.29) |
| 1990 | High body-mass index | Both | Central Europe | 0.30 (0.16 to 0.50) |
| 2021 | High body-mass index | Male | Central Europe | 0.49 (0.26 to 0.84) |
| 2021 | High body-mass index | Female | Central Europe | 0.19 (0.10 to 0.33) |
| 2021 | High body-mass index | Both | Central Europe | 0.32 (0.17 to 0.56) |
| 1990 | High systolic blood pressure | Male | Central Europe | 1.15 (0.89 to 1.43) |
| 1990 | High systolic blood pressure | Female | Central Europe | 0.39 (0.30 to 0.50) |
| 1990 | High systolic blood pressure | Both | Central Europe | 0.70 (0.54 to 0.87) |
| 2021 | High systolic blood pressure | Male | Central Europe | 0.94 (0.69 to 1.20) |
| 2021 | High systolic blood pressure | Female | Central Europe | 0.34 (0.25 to 0.46) |
| 2021 | High systolic blood pressure | Both | Central Europe | 0.59 (0.45 to 0.76) |
| 1990 | Lead exposure | Male | Central Europe | 0.03 (0.00 to 0.07) |
| 1990 | Lead exposure | Female | Central Europe | 0.01 (0.00 to 0.01) |
| 1990 | Lead exposure | Both | Central Europe | 0.01 (0.00 to 0.04) |
| 2021 | Lead exposure | Male | Central Europe | 0.03 (0.00 to 0.07) |
| 2021 | Lead exposure | Female | Central Europe | 0.01 (0.00 to 0.02) |
| 2021 | Lead exposure | Both | Central Europe | 0.01 (0.00 to 0.04) |
| 1990 | Smoking | Male | Central Europe | 2.75 (2.40 to 3.11) |
| 1990 | Smoking | Female | Central Europe | 0.40 (0.34 to 0.47) |
| 1990 | Smoking | Both | Central Europe | 1.38 (1.21 to 1.55) |
| 2021 | Smoking | Male | Central Europe | 2.09 (1.77 to 2.46) |
| 2021 | Smoking | Female | Central Europe | 0.38 (0.29 to 0.49) |
| 2021 | Smoking | Both | Central Europe | 1.11 (0.93 to 1.30) |
| 1990 | Diet high in sodium | Male | Eastern Europe | 0.04 (0.00 to 0.13) |
| 1990 | Diet high in sodium | Female | Eastern Europe | 0.01 (0.00 to 0.02) |
| 1990 | Diet high in sodium | Both | Eastern Europe | 0.02 (0.00 to 0.06) |
| 2021 | Diet high in sodium | Male | Eastern Europe | 0.06 (0.00 to 0.18) |
| 2021 | Diet high in sodium | Female | Eastern Europe | 0.01 (0.00 to 0.04) |
| 2021 | Diet high in sodium | Both | Eastern Europe | 0.03 (0.00 to 0.09) |
| 1990 | Diet low in fruits | Male | Eastern Europe | 0.24 (0.17 to 0.32) |
| 1990 | Diet low in fruits | Female | Eastern Europe | 0.08 (0.06 to 0.11) |
| 1990 | Diet low in fruits | Both | Eastern Europe | 0.14 (0.10 to 0.18) |
| 2021 | Diet low in fruits | Male | Eastern Europe | 0.30 (0.20 to 0.41) |
| 2021 | Diet low in fruits | Female | Eastern Europe | 0.10 (0.07 to 0.14) |
| 2021 | Diet low in fruits | Both | Eastern Europe | 0.18 (0.12 to 0.24) |
| 1990 | Diet low in vegetables | Male | Eastern Europe | 0.13 (0.08 to 0.19) |
| 1990 | Diet low in vegetables | Female | Eastern Europe | 0.04 (0.03 to 0.06) |
| 1990 | Diet low in vegetables | Both | Eastern Europe | 0.07 (0.05 to 0.10) |
| 2021 | Diet low in vegetables | Male | Eastern Europe | 0.19 (0.12 to 0.28) |
| 2021 | Diet low in vegetables | Female | Eastern Europe | 0.06 (0.04 to 0.09) |
| 2021 | Diet low in vegetables | Both | Eastern Europe | 0.11 (0.07 to 0.16) |
| 1990 | High body-mass index | Male | Eastern Europe | 0.32 (0.18 to 0.56) |
| 1990 | High body-mass index | Female | Eastern Europe | 0.16 (0.08 to 0.26) |
| 1990 | High body-mass index | Both | Eastern Europe | 0.22 (0.12 to 0.36) |
| 2021 | High body-mass index | Male | Eastern Europe | 0.66 (0.34 to 1.14) |
| 2021 | High body-mass index | Female | Eastern Europe | 0.29 (0.15 to 0.49) |
| 2021 | High body-mass index | Both | Eastern Europe | 0.44 (0.23 to 0.73) |
| 1990 | High systolic blood pressure | Male | Eastern Europe | 0.92 (0.69 to 1.17) |
| 1990 | High systolic blood pressure | Female | Eastern Europe | 0.34 (0.26 to 0.41) |
| 1990 | High systolic blood pressure | Both | Eastern Europe | 0.53 (0.41 to 0.67) |
| 2021 | High systolic blood pressure | Male | Eastern Europe | 1.32 (0.96 to 1.67) |
| 2021 | High systolic blood pressure | Female | Eastern Europe | 0.46 (0.33 to 0.59) |
| 2021 | High systolic blood pressure | Both | Eastern Europe | 0.79 (0.59 to 1.00) |
| 1990 | Lead exposure | Male | Eastern Europe | 0.01 (0.00 to 0.04) |
| 1990 | Lead exposure | Female | Eastern Europe | 0.00 (0.00 to 0.01) |
| 1990 | Lead exposure | Both | Eastern Europe | 0.01 (0.00 to 0.02) |
| 2021 | Lead exposure | Male | Eastern Europe | 0.02 (0.00 to 0.06) |
| 2021 | Lead exposure | Female | Eastern Europe | 0.01 (0.00 to 0.02) |
| 2021 | Lead exposure | Both | Eastern Europe | 0.01 (0.00 to 0.03) |
| 1990 | Smoking | Male | Eastern Europe | 2.54 (2.25 to 2.82) |
| 1990 | Smoking | Female | Eastern Europe | 0.14 (0.11 to 0.16) |
| 1990 | Smoking | Both | Eastern Europe | 0.96 (0.86 to 1.06) |
| 2021 | Smoking | Male | Eastern Europe | 3.50 (2.97 to 4.02) |
| 2021 | Smoking | Female | Eastern Europe | 0.24 (0.19 to 0.32) |
| 2021 | Smoking | Both | Eastern Europe | 1.50 (1.28 to 1.72) |
| 2021 | Diet high in sodium | Male | Western Europe | 0.03 (0.00 to 0.09) |
| 2021 | Diet high in sodium | Female | Western Europe | 0.00 (0.00 to 0.02) |
| 2021 | Diet high in sodium | Both | Western Europe | 0.01 (0.00 to 0.05) |
| 1990 | Diet high in sodium | Male | Western Europe | 0.04 (0.00 to 0.16) |
| 1990 | Diet high in sodium | Female | Western Europe | 0.01 (0.00 to 0.03) |
| 1990 | Diet high in sodium | Both | Western Europe | 0.02 (0.00 to 0.08) |
| 1990 | Diet low in fruits | Male | Western Europe | 0.34 (0.24 to 0.46) |
| 1990 | Diet low in fruits | Female | Western Europe | 0.11 (0.08 to 0.15) |
| 1990 | Diet low in fruits | Both | Western Europe | 0.20 (0.14 to 0.27) |
| 2021 | Diet low in fruits | Male | Western Europe | 0.12 (0.08 to 0.16) |
| 2021 | Diet low in fruits | Female | Western Europe | 0.05 (0.03 to 0.07) |
| 2021 | Diet low in fruits | Both | Western Europe | 0.08 (0.05 to 0.10) |
| 1990 | Diet low in vegetables | Male | Western Europe | 0.32 (0.21 to 0.46) |
| 1990 | Diet low in vegetables | Female | Western Europe | 0.10 (0.07 to 0.14) |
| 1990 | Diet low in vegetables | Both | Western Europe | 0.19 (0.12 to 0.27) |
| 2021 | Diet low in vegetables | Male | Western Europe | 0.12 (0.08 to 0.17) |
| 2021 | Diet low in vegetables | Female | Western Europe | 0.05 (0.03 to 0.07) |
| 2021 | Diet low in vegetables | Both | Western Europe | 0.08 (0.05 to 0.11) |
| 1990 | High body-mass index | Male | Western Europe | 0.69 (0.37 to 1.16) |
| 1990 | High body-mass index | Female | Western Europe | 0.21 (0.11 to 0.36) |
| 1990 | High body-mass index | Both | Western Europe | 0.40 (0.22 to 0.68) |
| 2021 | High body-mass index | Male | Western Europe | 0.33 (0.17 to 0.58) |
| 2021 | High body-mass index | Female | Western Europe | 0.14 (0.07 to 0.24) |
| 2021 | High body-mass index | Both | Western Europe | 0.23 (0.12 to 0.39) |
| 1990 | High systolic blood pressure | Male | Western Europe | 1.95 (1.49 to 2.43) |
| 1990 | High systolic blood pressure | Female | Western Europe | 0.60 (0.46 to 0.75) |
| 1990 | High systolic blood pressure | Both | Western Europe | 1.13 (0.86 to 1.41) |
| 2021 | High systolic blood pressure | Male | Western Europe | 0.67 (0.50 to 0.85) |
| 2021 | High systolic blood pressure | Female | Western Europe | 0.27 (0.20 to 0.35) |
| 2021 | High systolic blood pressure | Both | Western Europe | 0.45 (0.33 to 0.57) |
| 1990 | Lead exposure | Male | Western Europe | 0.04 (-0.01 to 0.10) |
| 1990 | Lead exposure | Female | Western Europe | 0.01 (0.00 to 0.02) |
| 1990 | Lead exposure | Both | Western Europe | 0.02 (0.00 to 0.05) |
| 2021 | Lead exposure | Male | Western Europe | 0.02 (0.00 to 0.06) |
| 2021 | Lead exposure | Female | Western Europe | 0.01 (0.00 to 0.02) |
| 2021 | Lead exposure | Both | Western Europe | 0.01 (0.00 to 0.03) |
| 1990 | Smoking | Male | Western Europe | 4.32 (3.70 to 4.96) |
| 1990 | Smoking | Female | Western Europe | 0.72 (0.59 to 0.87) |
| 1990 | Smoking | Both | Western Europe | 2.15 (1.83 to 2.48) |
| 2021 | Smoking | Male | Western Europe | 1.57 (1.30 to 1.87) |
| 2021 | Smoking | Female | Western Europe | 0.35 (0.27 to 0.43) |
| 2021 | Smoking | Both | Western Europe | 0.89 (0.74 to 1.07) |

### Data for Figure 8

#### Fig 8A. Percentage contribution of risk factors to AA-ASDR in Asia in 2021

| **Risk factor** | **Location** | **Percent contribution to ASDR (%)** |
| --- | --- | --- |
| Diet high in sodium | Central Asia | 0.77 (0.03 to 2.59) |
| Diet low in fruits | Central Asia | 3.73 (2.60 to 4.99) |
| Diet low in vegetables | Central Asia | 0.87 (0.50 to 1.41) |
| High body-mass index | Central Asia | 9.87 (5.44 to 16.87) |
| High systolic blood pressure | Central Asia | 20.82 (15.91 to 26.16) |
| Lead exposure | Central Asia | 0.50 (-0.07 to 1.29) |
| Smoking | Central Asia | 34.08 (29.64 to 38.25) |
| Diet high in sodium | East Asia | 2.05 (0.39 to 4.80) |
| Diet low in fruits | East Asia | 3.20 (2.17 to 4.30) |
| Diet low in vegetables | East Asia | 0.52 (0.26 to 0.88) |
| High body-mass index | East Asia | 5.69 (3.13 to 9.71) |
| High systolic blood pressure | East Asia | 15.69 (11.28 to 20.21) |
| Lead exposure | East Asia | 0.96 (-0.13 to 2.44) |
| Smoking | East Asia | 43.57 (37.28 to 49.59) |
| Diet high in sodium | Japan | 1.05 (0.06 to 3.16) |
| Diet low in fruits | Japan | 4.11 (2.78 to 5.46) |
| Diet low in vegetables | Japan | 1.29 (0.72 to 2.04) |
| High body-mass index | Japan | 4.57 (2.49 to 7.25) |
| High systolic blood pressure | Japan | 17.64 (13.11 to 22.97) |
| Lead exposure | Japan | 0.34 (-0.05 to 0.91) |
| Smoking | Japan | 29.69 (24.82 to 34.90) |
| Diet high in sodium | Republic of Korea | 1.99 (0.25 to 5.12) |
| Diet low in fruits | Republic of Korea | 2.96 (2.07 to 3.93) |
| Diet low in vegetables | Republic of Korea | 1.71 (1.03 to 2.66) |
| High body-mass index | Republic of Korea | 3.78 (2.01 to 6.14) |
| High systolic blood pressure | Republic of Korea | 11.55 (7.86 to 15.68) |
| Lead exposure | Republic of Korea | 0.79 (-0.10 to 2.06) |
| Smoking | Republic of Korea | 29.91 (23.92 to 36.27) |
| Diet high in sodium | Singapore | 1.63 (0.12 to 4.41) |
| Diet low in fruits | Singapore | 1.87 (1.21 to 2.56) |
| Diet low in vegetables | Singapore | 1.64 (0.96 to 2.51) |
| High body-mass index | Singapore | 4.61 (2.43 to 7.86) |
| High systolic blood pressure | Singapore | 11.01 (7.35 to 15.23) |
| Lead exposure | Singapore | 0.96 (-0.12 to 2.45) |
| Smoking | Singapore | 19.39 (15.48 to 23.59) |
| Diet high in sodium | South Asia | 0.71 (0.02 to 2.62) |
| Diet low in fruits | South Asia | 5.53 (3.84 to 7.22) |
| Diet low in vegetables | South Asia | 4.23 (2.88 to 5.86) |
| High body-mass index | South Asia | 3.24 (1.84 to 5.26) |
| High systolic blood pressure | South Asia | 16.27 (12.19 to 20.76) |
| Lead exposure | South Asia | 1.42 (-0.19 to 3.58) |
| Smoking | South Asia | 25.18 (19.66 to 31.60) |
| Diet high in sodium | Southeast Asia | 1.24 (0.13 to 3.35) |
| Diet low in fruits | Southeast Asia | 3.61 (2.48 to 4.88) |
| Diet low in vegetables | Southeast Asia | 4.51 (3.03 to 6.30) |
| High body-mass index | Southeast Asia | 3.85 (2.13 to 6.13) |
| High systolic blood pressure | Southeast Asia | 19.09 (14.31 to 24.02) |
| Lead exposure | Southeast Asia | 0.59 (-0.08 to 1.53) |
| Smoking | Southeast Asia | 29.26 (24.25 to 34.17) |

#### Fig 8B. Percentage contribution of risk factors to AA-ASDR in Asia grouped by sex in 2021

| **Risk factor** | **Sex** | **Location** | **Percent contribution to ASDR (%)** |
| --- | --- | --- | --- |
| Diet high in sodium | Male | Central Asia | 0.95 (0.06 to 2.90) |
| Diet high in sodium | Female | Central Asia | 0.52 (0.00 to 2.04) |
| Diet low in fruits | Male | Central Asia | 3.59 (2.51 to 4.79) |
| Diet low in fruits | Female | Central Asia | 3.86 (2.66 to 5.14) |
| Diet low in vegetables | Male | Central Asia | 0.93 (0.53 to 1.49) |
| Diet low in vegetables | Female | Central Asia | 0.78 (0.42 to 1.32) |
| High body-mass index | Male | Central Asia | 8.23 (4.52 to 14.15) |
| High body-mass index | Female | Central Asia | 12.18 (6.64 to 20.84) |
| High systolic blood pressure | Male | Central Asia | 20.07 (15.25 to 25.33) |
| High systolic blood pressure | Female | Central Asia | 21.65 (16.17 to 27.51) |
| Lead exposure | Male | Central Asia | 0.55 (-0.07 to 1.45) |
| Lead exposure | Female | Central Asia | 0.42 (-0.06 to 1.15) |
| Smoking | Male | Central Asia | 50.54 (44.07 to 56.73) |
| Smoking | Female | Central Asia | 4.67 (3.65 to 5.85) |
| Diet high in sodium | Male | East Asia | 2.19 (0.41 to 5.07) |
| Diet high in sodium | Female | East Asia | 1.61 (0.27 to 4.08) |
| Diet low in fruits | Male | East Asia | 3.20 (2.16 to 4.35) |
| Diet low in fruits | Female | East Asia | 3.10 (2.07 to 4.27) |
| Diet low in vegetables | Male | East Asia | 0.55 (0.27 to 0.96) |
| Diet low in vegetables | Female | East Asia | 0.53 (0.26 to 0.91) |
| High body-mass index | Male | East Asia | 5.27 (2.90 to 9.12) |
| High body-mass index | Female | East Asia | 6.52 (3.55 to 10.97) |
| High systolic blood pressure | Male | East Asia | 15.17 (10.81 to 20.05) |
| High systolic blood pressure | Female | East Asia | 16.88 (11.49 to 22.81) |
| Lead exposure | Male | East Asia | 1.05 (-0.14 to 2.66) |
| Lead exposure | Female | East Asia | 0.78 (-0.10 to 1.97) |
| Smoking | Male | East Asia | 55.28 (48.99 to 60.71) |
| Smoking | Female | East Asia | 11.04 (8.28 to 13.96) |
| Diet high in sodium | Male | Japan | 1.19 (0.08 to 3.32) |
| Diet high in sodium | Female | Japan | 0.86 (0.02 to 2.81) |
| Diet low in fruits | Male | Japan | 4.22 (2.92 to 5.69) |
| Diet low in fruits | Female | Japan | 3.86 (2.61 to 5.22) |
| Diet low in vegetables | Male | Japan | 1.24 (0.63 to 2.03) |
| Diet low in vegetables | Female | Japan | 1.34 (0.69 to 2.25) |
| High body-mass index | Male | Japan | 4.55 (2.46 to 7.37) |
| High body-mass index | Female | Japan | 4.45 (2.30 to 7.45) |
| High systolic blood pressure | Male | Japan | 17.86 (13.28 to 23.53) |
| High systolic blood pressure | Female | Japan | 17.19 (11.77 to 23.51) |
| Lead exposure | Male | Japan | 0.35 (-0.04 to 0.95) |
| Lead exposure | Female | Japan | 0.33 (-0.05 to 0.86) |
| Smoking | Male | Japan | 42.02 (35.73 to 48.55) |
| Smoking | Female | Japan | 11.64 (8.77 to 15.14) |
| Diet high in sodium | Male | Republic of Korea | 2.16 (0.28 to 5.47) |
| Diet high in sodium | Female | Republic of Korea | 1.82 (0.19 to 4.86) |
| Diet low in fruits | Male | Republic of Korea | 2.85 (1.94 to 3.84) |
| Diet low in fruits | Female | Republic of Korea | 3.02 (2.09 to 4.20) |
| Diet low in vegetables | Male | Republic of Korea | 1.62 (0.92 to 2.58) |
| Diet low in vegetables | Female | Republic of Korea | 1.82 (1.03 to 2.88) |
| High body-mass index | Male | Republic of Korea | 3.29 (1.78 to 5.55) |
| High body-mass index | Female | Republic of Korea | 4.16 (2.19 to 7.05) |
| High systolic blood pressure | Male | Republic of Korea | 10.55 (6.91 to 14.78) |
| High systolic blood pressure | Female | Republic of Korea | 12.35 (7.79 to 17.73) |
| Lead exposure | Male | Republic of Korea | 0.90 (-0.11 to 2.36) |
| Lead exposure | Female | Republic of Korea | 0.70 (-0.09 to 1.84) |
| Smoking | Male | Republic of Korea | 44.52 (36.54 to 52.34) |
| Smoking | Female | Republic of Korea | 12.61 (8.59 to 18.38) |
| Diet high in sodium | Male | Singapore | 1.81 (0.13 to 4.97) |
| Diet high in sodium | Female | Singapore | 1.34 (0.07 to 3.96) |
| Diet low in fruits | Male | Singapore | 1.83 (1.20 to 2.55) |
| Diet low in fruits | Female | Singapore | 1.89 (1.20 to 2.69) |
| Diet low in vegetables | Male | Singapore | 1.69 (0.98 to 2.57) |
| Diet low in vegetables | Female | Singapore | 1.56 (0.86 to 2.48) |
| High body-mass index | Male | Singapore | 4.69 (2.51 to 8.20) |
| High body-mass index | Female | Singapore | 4.41 (2.24 to 7.63) |
| High systolic blood pressure | Male | Singapore | 10.76 (7.11 to 15.62) |
| High systolic blood pressure | Female | Singapore | 11.19 (6.49 to 16.48) |
| Lead exposure | Male | Singapore | 1.03 (-0.13 to 2.63) |
| Lead exposure | Female | Singapore | 0.88 (-0.12 to 2.26) |
| Smoking | Male | Singapore | 25.85 (20.80 to 31.19) |
| Smoking | Female | Singapore | 7.27 (4.84 to 10.49) |
| Diet high in sodium | Male | South Asia | 0.83 (0.03 to 2.94) |
| Diet high in sodium | Female | South Asia | 0.53 (0.00 to 2.14) |
| Diet low in fruits | Male | South Asia | 5.41 (3.76 to 7.06) |
| Diet low in fruits | Female | South Asia | 5.68 (3.94 to 7.42) |
| Diet low in vegetables | Male | South Asia | 4.09 (2.79 to 5.67) |
| Diet low in vegetables | Female | South Asia | 4.43 (3.02 to 6.14) |
| High body-mass index | Male | South Asia | 2.84 (1.63 to 4.57) |
| High body-mass index | Female | South Asia | 3.85 (2.12 to 6.44) |
| High systolic blood pressure | Male | South Asia | 15.57 (11.63 to 19.92) |
| High systolic blood pressure | Female | South Asia | 17.34 (13.04 to 21.92) |
| Lead exposure | Male | South Asia | 1.54 (-0.21 to 3.87) |
| Lead exposure | Female | South Asia | 1.24 (-0.18 to 3.11) |
| Smoking | Male | South Asia | 35.88 (30.17 to 42.21) |
| Smoking | Female | South Asia | 8.47 (6.47 to 11.09) |
| Diet high in sodium | Male | Southeast Asia | 1.45 (0.17 to 3.82) |
| Diet high in sodium | Female | Southeast Asia | 0.91 (0.05 to 2.74) |
| Diet low in fruits | Male | Southeast Asia | 3.43 (2.34 to 4.61) |
| Diet low in fruits | Female | Southeast Asia | 3.83 (2.58 to 5.27) |
| Diet low in vegetables | Male | Southeast Asia | 4.39 (2.95 to 6.11) |
| Diet low in vegetables | Female | Southeast Asia | 4.67 (3.13 to 6.61) |
| High body-mass index | Male | Southeast Asia | 3.19 (1.77 to 5.18) |
| High body-mass index | Female | Southeast Asia | 4.83 (2.61 to 7.84) |
| High systolic blood pressure | Male | Southeast Asia | 18.25 (13.47 to 23.34) |
| High systolic blood pressure | Female | Southeast Asia | 20.18 (15.14 to 25.80) |
| Lead exposure | Male | Southeast Asia | 0.68 (-0.09 to 1.75) |
| Lead exposure | Female | Southeast Asia | 0.44 (-0.06 to 1.15) |
| Smoking | Male | Southeast Asia | 42.36 (36.33 to 48.85) |
| Smoking | Female | Southeast Asia | 7.42 (5.74 to 9.44) |

#### Fig 8C. AA-ASDR attributable to risk factors in Asia grouped by sex from 1990 to 2021

| **Year** | **Risk factors** | **Sex** | **Location** | **ASDR per 100,000** |
| --- | --- | --- | --- | --- |
| 2021 | Diet high in sodium | Male | Central Asia | 0.03 (0.00 to 0.09) |
| 2021 | Diet high in sodium | Female | Central Asia | 0.01 (0.00 to 0.02) |
| 2021 | Diet high in sodium | Both | Central Asia | 0.02 (0.00 to 0.05) |
| 1990 | Diet high in sodium | Male | Central Asia | 0.02 (0.00 to 0.05) |
| 1990 | Diet high in sodium | Female | Central Asia | 0.00 (0.00 to 0.02) |
| 1990 | Diet high in sodium | Both | Central Asia | 0.01 (0.00 to 0.03) |
| 1990 | Diet low in fruits | Male | Central Asia | 0.07 (0.05 to 0.10) |
| 1990 | Diet low in fruits | Female | Central Asia | 0.03 (0.02 to 0.05) |
| 1990 | Diet low in fruits | Both | Central Asia | 0.05 (0.03 to 0.07) |
| 2021 | Diet low in fruits | Male | Central Asia | 0.11 (0.08 to 0.15) |
| 2021 | Diet low in fruits | Female | Central Asia | 0.05 (0.03 to 0.07) |
| 2021 | Diet low in fruits | Both | Central Asia | 0.07 (0.05 to 0.10) |
| 1990 | Diet low in vegetables | Male | Central Asia | 0.03 (0.02 to 0.05) |
| 1990 | Diet low in vegetables | Female | Central Asia | 0.01 (0.01 to 0.02) |
| 1990 | Diet low in vegetables | Both | Central Asia | 0.02 (0.01 to 0.03) |
| 2021 | Diet low in vegetables | Male | Central Asia | 0.03 (0.02 to 0.05) |
| 2021 | Diet low in vegetables | Female | Central Asia | 0.01 (0.01 to 0.02) |
| 2021 | Diet low in vegetables | Both | Central Asia | 0.02 (0.01 to 0.03) |
| 1990 | High body-mass index | Male | Central Asia | 0.10 (0.05 to 0.17) |
| 1990 | High body-mass index | Female | Central Asia | 0.06 (0.03 to 0.11) |
| 1990 | High body-mass index | Both | Central Asia | 0.08 (0.04 to 0.14) |
| 2021 | High body-mass index | Male | Central Asia | 0.25 (0.14 to 0.43) |
| 2021 | High body-mass index | Female | Central Asia | 0.15 (0.08 to 0.25) |
| 2021 | High body-mass index | Both | Central Asia | 0.20 (0.11 to 0.34) |
| 1990 | High systolic blood pressure | Male | Central Asia | 0.28 (0.21 to 0.38) |
| 1990 | High systolic blood pressure | Female | Central Asia | 0.13 (0.09 to 0.17) |
| 1990 | High systolic blood pressure | Both | Central Asia | 0.19 (0.14 to 0.25) |
| 2021 | High systolic blood pressure | Male | Central Asia | 0.62 (0.46 to 0.80) |
| 2021 | High systolic blood pressure | Female | Central Asia | 0.26 (0.19 to 0.35) |
| 2021 | High systolic blood pressure | Both | Central Asia | 0.41 (0.31 to 0.54) |
| 1990 | Lead exposure | Male | Central Asia | 0.01 (0.00 to 0.02) |
| 1990 | Lead exposure | Female | Central Asia | 0.00 (0.00 to 0.01) |
| 1990 | Lead exposure | Both | Central Asia | 0.00 (0.00 to 0.01) |
| 2021 | Lead exposure | Male | Central Asia | 0.02 (0.00 to 0.04) |
| 2021 | Lead exposure | Female | Central Asia | 0.01 (0.00 to 0.01) |
| 2021 | Lead exposure | Both | Central Asia | 0.01 (0.00 to 0.03) |
| 1990 | Smoking | Male | Central Asia | 0.77 (0.65 to 0.95) |
| 1990 | Smoking | Female | Central Asia | 0.04 (0.03 to 0.05) |
| 1990 | Smoking | Both | Central Asia | 0.33 (0.28 to 0.40) |
| 2021 | Smoking | Male | Central Asia | 1.56 (1.30 to 1.83) |
| 2021 | Smoking | Female | Central Asia | 0.06 (0.04 to 0.08) |
| 2021 | Smoking | Both | Central Asia | 0.67 (0.56 to 0.79) |
| 1990 | Diet high in sodium | Male | East Asia | 0.01 (0.00 to 0.03) |
| 1990 | Diet high in sodium | Female | East Asia | 0.01 (0.00 to 0.01) |
| 1990 | Diet high in sodium | Both | East Asia | 0.01 (0.00 to 0.02) |
| 2021 | Diet high in sodium | Male | East Asia | 0.02 (0.00 to 0.04) |
| 2021 | Diet high in sodium | Female | East Asia | 0.00 (0.00 to 0.01) |
| 2021 | Diet high in sodium | Both | East Asia | 0.01 (0.00 to 0.03) |
| 1990 | Diet low in fruits | Male | East Asia | 0.02 (0.01 to 0.03) |
| 1990 | Diet low in fruits | Female | East Asia | 0.01 (0.01 to 0.02) |
| 1990 | Diet low in fruits | Both | East Asia | 0.01 (0.01 to 0.02) |
| 2021 | Diet low in fruits | Male | East Asia | 0.03 (0.01 to 0.04) |
| 2021 | Diet low in fruits | Female | East Asia | 0.01 (0.00 to 0.01) |
| 2021 | Diet low in fruits | Both | East Asia | 0.02 (0.01 to 0.02) |
| 1990 | Diet low in vegetables | Male | East Asia | 0.01 (0.01 to 0.02) |
| 1990 | Diet low in vegetables | Female | East Asia | 0.01 (0.00 to 0.01) |
| 1990 | Diet low in vegetables | Both | East Asia | 0.01 (0.01 to 0.01) |
| 2021 | Diet low in vegetables | Male | East Asia | 0.00 (0.00 to 0.01) |
| 2021 | Diet low in vegetables | Female | East Asia | 0.00 (0.00 to 0.00) |
| 2021 | Diet low in vegetables | Both | East Asia | 0.00 (0.00 to 0.00) |
| 1990 | High body-mass index | Male | East Asia | 0.01 (0.01 to 0.02) |
| 1990 | High body-mass index | Female | East Asia | 0.01 (0.00 to 0.02) |
| 1990 | High body-mass index | Both | East Asia | 0.01 (0.00 to 0.02) |
| 2021 | High body-mass index | Male | East Asia | 0.04 (0.02 to 0.07) |
| 2021 | High body-mass index | Female | East Asia | 0.02 (0.01 to 0.03) |
| 2021 | High body-mass index | Both | East Asia | 0.03 (0.01 to 0.05) |
| 1990 | High systolic blood pressure | Male | East Asia | 0.05 (0.03 to 0.08) |
| 1990 | High systolic blood pressure | Female | East Asia | 0.04 (0.02 to 0.06) |
| 1990 | High systolic blood pressure | Both | East Asia | 0.05 (0.03 to 0.06) |
| 2021 | High systolic blood pressure | Male | East Asia | 0.12 (0.08 to 0.18) |
| 2021 | High systolic blood pressure | Female | East Asia | 0.04 (0.03 to 0.06) |
| 2021 | High systolic blood pressure | Both | East Asia | 0.08 (0.05 to 0.11) |
| 1990 | Lead exposure | Male | East Asia | 0.01 (0.00 to 0.01) |
| 1990 | Lead exposure | Female | East Asia | 0.00 (0.00 to 0.01) |
| 1990 | Lead exposure | Both | East Asia | 0.00 (0.00 to 0.01) |
| 2021 | Lead exposure | Male | East Asia | 0.01 (0.00 to 0.02) |
| 2021 | Lead exposure | Female | East Asia | 0.00 (0.00 to 0.01) |
| 2021 | Lead exposure | Both | East Asia | 0.00 (0.00 to 0.01) |
| 1990 | Smoking | Male | East Asia | 0.26 (0.18 to 0.37) |
| 1990 | Smoking | Female | East Asia | 0.04 (0.03 to 0.06) |
| 1990 | Smoking | Both | East Asia | 0.15 (0.11 to 0.20) |
| 2021 | Smoking | Male | East Asia | 0.43 (0.31 to 0.56) |
| 2021 | Smoking | Female | East Asia | 0.03 (0.02 to 0.04) |
| 2021 | Smoking | Both | East Asia | 0.22 (0.16 to 0.28) |
| 1990 | Diet high in sodium | Male | Japan | 0.06 (0.01 to 0.15) |
| 1990 | Diet high in sodium | Female | Japan | 0.02 (0.00 to 0.05) |
| 1990 | Diet high in sodium | Both | Japan | 0.04 (0.01 to 0.09) |
| 2021 | Diet high in sodium | Male | Japan | 0.08 (0.01 to 0.22) |
| 2021 | Diet high in sodium | Female | Japan | 0.03 (0.00 to 0.11) |
| 2021 | Diet high in sodium | Both | Japan | 0.05 (0.00 to 0.16) |
| 1990 | Diet low in fruits | Male | Japan | 0.22 (0.16 to 0.29) |
| 1990 | Diet low in fruits | Female | Japan | 0.09 (0.06 to 0.12) |
| 1990 | Diet low in fruits | Both | Japan | 0.14 (0.10 to 0.19) |
| 2021 | Diet low in fruits | Male | Japan | 0.28 (0.19 to 0.38) |
| 2021 | Diet low in fruits | Female | Japan | 0.14 (0.09 to 0.20) |
| 2021 | Diet low in fruits | Both | Japan | 0.21 (0.14 to 0.28) |
| 1990 | Diet low in vegetables | Male | Japan | 0.11 (0.07 to 0.16) |
| 1990 | Diet low in vegetables | Female | Japan | 0.04 (0.03 to 0.07) |
| 1990 | Diet low in vegetables | Both | Japan | 0.07 (0.04 to 0.10) |
| 2021 | Diet low in vegetables | Male | Japan | 0.08 (0.04 to 0.13) |
| 2021 | Diet low in vegetables | Female | Japan | 0.05 (0.03 to 0.09) |
| 2021 | Diet low in vegetables | Both | Japan | 0.07 (0.04 to 0.11) |
| 1990 | High body-mass index | Male | Japan | 0.16 (0.09 to 0.27) |
| 1990 | High body-mass index | Female | Japan | 0.08 (0.04 to 0.14) |
| 1990 | High body-mass index | Both | Japan | 0.12 (0.06 to 0.19) |
| 2021 | High body-mass index | Male | Japan | 0.30 (0.16 to 0.49) |
| 2021 | High body-mass index | Female | Japan | 0.17 (0.08 to 0.28) |
| 2021 | High body-mass index | Both | Japan | 0.23 (0.12 to 0.37) |
| 1990 | High systolic blood pressure | Male | Japan | 1.01 (0.77 to 1.26) |
| 1990 | High systolic blood pressure | Female | Japan | 0.41 (0.31 to 0.51) |
| 1990 | High systolic blood pressure | Both | Japan | 0.66 (0.50 to 0.82) |
| 2021 | High systolic blood pressure | Male | Japan | 1.18 (0.89 to 1.56) |
| 2021 | High systolic blood pressure | Female | Japan | 0.64 (0.41 to 0.92) |
| 2021 | High systolic blood pressure | Both | Japan | 0.89 (0.64 to 1.18) |
| 1990 | Lead exposure | Male | Japan | 0.01 (0.00 to 0.04) |
| 1990 | Lead exposure | Female | Japan | 0.01 (0.00 to 0.01) |
| 1990 | Lead exposure | Both | Japan | 0.01 (0.00 to 0.02) |
| 2021 | Lead exposure | Male | Japan | 0.02 (0.00 to 0.06) |
| 2021 | Lead exposure | Female | Japan | 0.01 (0.00 to 0.03) |
| 2021 | Lead exposure | Both | Japan | 0.02 (0.00 to 0.05) |
| 1990 | Smoking | Male | Japan | 2.47 (2.12 to 2.81) |
| 1990 | Smoking | Female | Japan | 0.28 (0.22 to 0.36) |
| 1990 | Smoking | Both | Japan | 1.17 (1.01 to 1.34) |
| 2021 | Smoking | Male | Japan | 2.78 (2.34 to 3.26) |
| 2021 | Smoking | Female | Japan | 0.43 (0.31 to 0.59) |
| 2021 | Smoking | Both | Japan | 1.50 (1.25 to 1.78) |
| 2021 | Diet high in sodium | Male | Republic of Korea | 0.05 (0.01 to 0.15) |
| 2021 | Diet high in sodium | Female | Republic of Korea | 0.03 (0.00 to 0.07) |
| 2021 | Diet high in sodium | Both | Republic of Korea | 0.04 (0.00 to 0.10) |
| 1990 | Diet high in sodium | Male | Republic of Korea | 0.04 (0.00 to 0.12) |
| 1990 | Diet high in sodium | Female | Republic of Korea | 0.02 (0.00 to 0.07) |
| 1990 | Diet high in sodium | Both | Republic of Korea | 0.03 (0.00 to 0.08) |
| 1990 | Diet low in fruits | Male | Republic of Korea | 0.09 (0.05 to 0.16) |
| 1990 | Diet low in fruits | Female | Republic of Korea | 0.06 (0.04 to 0.11) |
| 1990 | Diet low in fruits | Both | Republic of Korea | 0.08 (0.04 to 0.12) |
| 2021 | Diet low in fruits | Male | Republic of Korea | 0.07 (0.05 to 0.10) |
| 2021 | Diet low in fruits | Female | Republic of Korea | 0.05 (0.03 to 0.07) |
| 2021 | Diet low in fruits | Both | Republic of Korea | 0.06 (0.04 to 0.08) |
| 1990 | Diet low in vegetables | Male | Republic of Korea | 0.03 (0.01 to 0.06) |
| 1990 | Diet low in vegetables | Female | Republic of Korea | 0.02 (0.01 to 0.04) |
| 1990 | Diet low in vegetables | Both | Republic of Korea | 0.02 (0.01 to 0.04) |
| 2021 | Diet low in vegetables | Male | Republic of Korea | 0.04 (0.02 to 0.06) |
| 2021 | Diet low in vegetables | Female | Republic of Korea | 0.03 (0.01 to 0.05) |
| 2021 | Diet low in vegetables | Both | Republic of Korea | 0.03 (0.02 to 0.05) |
| 1990 | High body-mass index | Male | Republic of Korea | 0.07 (0.02 to 0.14) |
| 1990 | High body-mass index | Female | Republic of Korea | 0.05 (0.02 to 0.09) |
| 1990 | High body-mass index | Both | Republic of Korea | 0.06 (0.02 to 0.11) |
| 2021 | High body-mass index | Male | Republic of Korea | 0.08 (0.04 to 0.15) |
| 2021 | High body-mass index | Female | Republic of Korea | 0.06 (0.03 to 0.11) |
| 2021 | High body-mass index | Both | Republic of Korea | 0.07 (0.03 to 0.13) |
| 1990 | High systolic blood pressure | Male | Republic of Korea | 0.35 (0.18 to 0.59) |
| 1990 | High systolic blood pressure | Female | Republic of Korea | 0.26 (0.15 to 0.41) |
| 1990 | High systolic blood pressure | Both | Republic of Korea | 0.30 (0.17 to 0.45) |
| 2021 | High systolic blood pressure | Male | Republic of Korea | 0.26 (0.15 to 0.39) |
| 2021 | High systolic blood pressure | Female | Republic of Korea | 0.18 (0.10 to 0.29) |
| 2021 | High systolic blood pressure | Both | Republic of Korea | 0.22 (0.14 to 0.31) |
| 1990 | Lead exposure | Male | Republic of Korea | 0.02 (0.00 to 0.06) |
| 1990 | Lead exposure | Female | Republic of Korea | 0.01 (0.00 to 0.03) |
| 1990 | Lead exposure | Both | Republic of Korea | 0.01 (0.00 to 0.04) |
| 2021 | Lead exposure | Male | Republic of Korea | 0.02 (0.00 to 0.06) |
| 2021 | Lead exposure | Female | Republic of Korea | 0.01 (0.00 to 0.03) |
| 2021 | Lead exposure | Both | Republic of Korea | 0.02 (0.00 to 0.04) |
| 1990 | Smoking | Male | Republic of Korea | 1.31 (0.75 to 2.00) |
| 1990 | Smoking | Female | Republic of Korea | 0.28 (0.16 to 0.45) |
| 1990 | Smoking | Both | Republic of Korea | 0.65 (0.39 to 0.97) |
| 2021 | Smoking | Male | Republic of Korea | 1.10 (0.81 to 1.43) |
| 2021 | Smoking | Female | Republic of Korea | 0.19 (0.11 to 0.30) |
| 2021 | Smoking | Both | Republic of Korea | 0.57 (0.43 to 0.72) |
| 2021 | Diet high in sodium | Male | Singapore | 0.05 (0.00 to 0.14) |
| 2021 | Diet high in sodium | Female | Singapore | 0.02 (0.00 to 0.06) |
| 2021 | Diet high in sodium | Both | Singapore | 0.04 (0.00 to 0.10) |
| 1990 | Diet high in sodium | Male | Singapore | 0.08 (0.01 to 0.20) |
| 1990 | Diet high in sodium | Female | Singapore | 0.02 (0.00 to 0.06) |
| 1990 | Diet high in sodium | Both | Singapore | 0.05 (0.01 to 0.12) |
| 1990 | Diet low in fruits | Male | Singapore | 0.14 (0.10 to 0.19) |
| 1990 | Diet low in fruits | Female | Singapore | 0.05 (0.03 to 0.08) |
| 1990 | Diet low in fruits | Both | Singapore | 0.09 (0.06 to 0.12) |
| 2021 | Diet low in fruits | Male | Singapore | 0.05 (0.03 to 0.07) |
| 2021 | Diet low in fruits | Female | Singapore | 0.03 (0.02 to 0.04) |
| 2021 | Diet low in fruits | Both | Singapore | 0.04 (0.03 to 0.06) |
| 1990 | Diet low in vegetables | Male | Singapore | 0.12 (0.08 to 0.18) |
| 1990 | Diet low in vegetables | Female | Singapore | 0.04 (0.03 to 0.07) |
| 1990 | Diet low in vegetables | Both | Singapore | 0.08 (0.05 to 0.12) |
| 2021 | Diet low in vegetables | Male | Singapore | 0.05 (0.03 to 0.07) |
| 2021 | Diet low in vegetables | Female | Singapore | 0.02 (0.01 to 0.04) |
| 2021 | Diet low in vegetables | Both | Singapore | 0.04 (0.02 to 0.05) |
| 1990 | High body-mass index | Male | Singapore | 0.14 (0.07 to 0.24) |
| 1990 | High body-mass index | Female | Singapore | 0.04 (0.02 to 0.07) |
| 1990 | High body-mass index | Both | Singapore | 0.09 (0.05 to 0.15) |
| 2021 | High body-mass index | Male | Singapore | 0.13 (0.07 to 0.23) |
| 2021 | High body-mass index | Female | Singapore | 0.07 (0.03 to 0.12) |
| 2021 | High body-mass index | Both | Singapore | 0.10 (0.05 to 0.17) |
| 1990 | High systolic blood pressure | Male | Singapore | 0.71 (0.52 to 0.92) |
| 1990 | High systolic blood pressure | Female | Singapore | 0.28 (0.18 to 0.39) |
| 1990 | High systolic blood pressure | Both | Singapore | 0.47 (0.34 to 0.61) |
| 2021 | High systolic blood pressure | Male | Singapore | 0.31 (0.19 to 0.45) |
| 2021 | High systolic blood pressure | Female | Singapore | 0.17 (0.10 to 0.26) |
| 2021 | High systolic blood pressure | Both | Singapore | 0.24 (0.15 to 0.33) |
| 1990 | Lead exposure | Male | Singapore | 0.04 (-0.01 to 0.11) |
| 1990 | Lead exposure | Female | Singapore | 0.01 (0.00 to 0.03) |
| 1990 | Lead exposure | Both | Singapore | 0.03 (0.00 to 0.07) |
| 2021 | Lead exposure | Male | Singapore | 0.03 (0.00 to 0.08) |
| 2021 | Lead exposure | Female | Singapore | 0.01 (0.00 to 0.04) |
| 2021 | Lead exposure | Both | Singapore | 0.02 (0.00 to 0.05) |
| 1990 | Smoking | Male | Singapore | 1.58 (1.30 to 1.88) |
| 1990 | Smoking | Female | Singapore | 0.17 (0.11 to 0.25) |
| 1990 | Smoking | Both | Singapore | 0.81 (0.67 to 0.97) |
| 2021 | Smoking | Male | Singapore | 0.74 (0.59 to 0.92) |
| 2021 | Smoking | Female | Singapore | 0.11 (0.07 to 0.16) |
| 2021 | Smoking | Both | Singapore | 0.42 (0.33 to 0.52) |
| 2021 | Diet high in sodium | Male | South Asia | 0.01 (0.00 to 0.05) |
| 2021 | Diet high in sodium | Female | South Asia | 0.00 (0.00 to 0.02) |
| 2021 | Diet high in sodium | Both | South Asia | 0.01 (0.00 to 0.03) |
| 1990 | Diet high in sodium | Male | South Asia | 0.01 (0.00 to 0.02) |
| 1990 | Diet high in sodium | Female | South Asia | 0.00 (0.00 to 0.02) |
| 1990 | Diet high in sodium | Both | South Asia | 0.00 (0.00 to 0.02) |
| 1990 | Diet low in fruits | Male | South Asia | 0.04 (0.02 to 0.10) |
| 1990 | Diet low in fruits | Female | South Asia | 0.04 (0.02 to 0.08) |
| 1990 | Diet low in fruits | Both | South Asia | 0.04 (0.02 to 0.07) |
| 2021 | Diet low in fruits | Male | South Asia | 0.08 (0.05 to 0.16) |
| 2021 | Diet low in fruits | Female | South Asia | 0.05 (0.03 to 0.08) |
| 2021 | Diet low in fruits | Both | South Asia | 0.07 (0.04 to 0.11) |
| 1990 | Diet low in vegetables | Male | South Asia | 0.04 (0.02 to 0.09) |
| 1990 | Diet low in vegetables | Female | South Asia | 0.03 (0.01 to 0.07) |
| 1990 | Diet low in vegetables | Both | South Asia | 0.03 (0.02 to 0.06) |
| 2021 | Diet low in vegetables | Male | South Asia | 0.06 (0.04 to 0.12) |
| 2021 | Diet low in vegetables | Female | South Asia | 0.04 (0.02 to 0.07) |
| 2021 | Diet low in vegetables | Both | South Asia | 0.05 (0.03 to 0.08) |
| 1990 | High body-mass index | Male | South Asia | 0.01 (0.00 to 0.03) |
| 1990 | High body-mass index | Female | South Asia | 0.01 (0.00 to 0.03) |
| 1990 | High body-mass index | Both | South Asia | 0.01 (0.01 to 0.02) |
| 2021 | High body-mass index | Male | South Asia | 0.04 (0.02 to 0.09) |
| 2021 | High body-mass index | Female | South Asia | 0.03 (0.02 to 0.07) |
| 2021 | High body-mass index | Both | South Asia | 0.04 (0.02 to 0.07) |
| 1990 | High systolic blood pressure | Male | South Asia | 0.11 (0.05 to 0.25) |
| 1990 | High systolic blood pressure | Female | South Asia | 0.11 (0.05 to 0.25) |
| 1990 | High systolic blood pressure | Both | South Asia | 0.11 (0.06 to 0.18) |
| 2021 | High systolic blood pressure | Male | South Asia | 0.24 (0.14 to 0.41) |
| 2021 | High systolic blood pressure | Female | South Asia | 0.16 (0.09 to 0.24) |
| 2021 | High systolic blood pressure | Both | South Asia | 0.20 (0.13 to 0.30) |
| 1990 | Lead exposure | Male | South Asia | 0.01 (0.00 to 0.04) |
| 1990 | Lead exposure | Female | South Asia | 0.01 (0.00 to 0.03) |
| 1990 | Lead exposure | Both | South Asia | 0.01 (0.00 to 0.03) |
| 2021 | Lead exposure | Male | South Asia | 0.02 (0.00 to 0.07) |
| 2021 | Lead exposure | Female | South Asia | 0.01 (0.00 to 0.03) |
| 2021 | Lead exposure | Both | South Asia | 0.02 (0.00 to 0.05) |
| 1990 | Smoking | Male | South Asia | 0.36 (0.17 to 0.81) |
| 1990 | Smoking | Female | South Asia | 0.07 (0.03 to 0.14) |
| 1990 | Smoking | Both | South Asia | 0.22 (0.13 to 0.44) |
| 2021 | Smoking | Male | South Asia | 0.57 (0.35 to 1.00) |
| 2021 | Smoking | Female | South Asia | 0.08 (0.04 to 0.12) |
| 2021 | Smoking | Both | South Asia | 0.31 (0.20 to 0.52) |
| 2021 | Diet high in sodium | Male | Southeast Asia | 0.03 (0.00 to 0.08) |
| 2021 | Diet high in sodium | Female | Southeast Asia | 0.01 (0.00 to 0.03) |
| 2021 | Diet high in sodium | Both | Southeast Asia | 0.02 (0.00 to 0.05) |
| 1990 | Diet high in sodium | Male | Southeast Asia | 0.03 (0.00 to 0.06) |
| 1990 | Diet high in sodium | Female | Southeast Asia | 0.01 (0.00 to 0.03) |
| 1990 | Diet high in sodium | Both | Southeast Asia | 0.02 (0.00 to 0.04) |
| 1990 | Diet low in fruits | Male | Southeast Asia | 0.06 (0.04 to 0.09) |
| 1990 | Diet low in fruits | Female | Southeast Asia | 0.04 (0.02 to 0.06) |
| 1990 | Diet low in fruits | Both | Southeast Asia | 0.05 (0.03 to 0.07) |
| 2021 | Diet low in fruits | Male | Southeast Asia | 0.07 (0.04 to 0.10) |
| 2021 | Diet low in fruits | Female | Southeast Asia | 0.04 (0.02 to 0.05) |
| 2021 | Diet low in fruits | Both | Southeast Asia | 0.05 (0.03 to 0.07) |
| 1990 | Diet low in vegetables | Male | Southeast Asia | 0.07 (0.04 to 0.10) |
| 1990 | Diet low in vegetables | Female | Southeast Asia | 0.04 (0.02 to 0.07) |
| 1990 | Diet low in vegetables | Both | Southeast Asia | 0.05 (0.03 to 0.08) |
| 2021 | Diet low in vegetables | Male | Southeast Asia | 0.09 (0.06 to 0.13) |
| 2021 | Diet low in vegetables | Female | Southeast Asia | 0.04 (0.03 to 0.06) |
| 2021 | Diet low in vegetables | Both | Southeast Asia | 0.06 (0.04 to 0.09) |
| 1990 | High body-mass index | Male | Southeast Asia | 0.02 (0.01 to 0.04) |
| 1990 | High body-mass index | Female | Southeast Asia | 0.02 (0.01 to 0.03) |
| 1990 | High body-mass index | Both | Southeast Asia | 0.02 (0.01 to 0.04) |
| 2021 | High body-mass index | Male | Southeast Asia | 0.06 (0.03 to 0.11) |
| 2021 | High body-mass index | Female | Southeast Asia | 0.04 (0.02 to 0.07) |
| 2021 | High body-mass index | Both | Southeast Asia | 0.05 (0.03 to 0.09) |
| 1990 | High systolic blood pressure | Male | Southeast Asia | 0.21 (0.14 to 0.32) |
| 1990 | High systolic blood pressure | Female | Southeast Asia | 0.14 (0.09 to 0.21) |
| 1990 | High systolic blood pressure | Both | Southeast Asia | 0.17 (0.12 to 0.24) |
| 2021 | High systolic blood pressure | Male | Southeast Asia | 0.37 (0.26 to 0.51) |
| 2021 | High systolic blood pressure | Female | Southeast Asia | 0.19 (0.13 to 0.25) |
| 2021 | High systolic blood pressure | Both | Southeast Asia | 0.26 (0.19 to 0.35) |
| 1990 | Lead exposure | Male | Southeast Asia | 0.01 (0.00 to 0.02) |
| 1990 | Lead exposure | Female | Southeast Asia | 0.00 (0.00 to 0.01) |
| 1990 | Lead exposure | Both | Southeast Asia | 0.01 (0.00 to 0.02) |
| 2021 | Lead exposure | Male | Southeast Asia | 0.01 (0.00 to 0.04) |
| 2021 | Lead exposure | Female | Southeast Asia | 0.00 (0.00 to 0.01) |
| 2021 | Lead exposure | Both | Southeast Asia | 0.01 (0.00 to 0.02) |
| 1990 | Smoking | Male | Southeast Asia | 0.67 (0.50 to 0.99) |
| 1990 | Smoking | Female | Southeast Asia | 0.09 (0.07 to 0.13) |
| 1990 | Smoking | Both | Southeast Asia | 0.35 (0.26 to 0.49) |
| 2021 | Smoking | Male | Southeast Asia | 0.85 (0.66 to 1.08) |
| 2021 | Smoking | Female | Southeast Asia | 0.07 (0.05 to 0.09) |
| 2021 | Smoking | Both | Southeast Asia | 0.41 (0.32 to 0.51) |
